# Ethnic Variation and its Association with Malaria Awareness: A Cross-sectional Study in the East Nusa Tenggara Province (Indonesia)

**DOI:** 10.1101/2021.11.04.21265794

**Authors:** Robertus Dole Guntur, Jonathan Kingsley, Fakir M. Amirul Islam

## Abstract

**Objectives:** This study aims to investigate ethnic variation and its association with malaria awareness in the East Nusa Tenggara Province (ENTP), Indonesia.

**Methods:** A community-based cross-sectional study was conducted upon 1495 adults recruited by multi-stage cluster random sampling technique. A malaria awareness related questionnaire was used to collect data alongside a malaria awareness index (MAI). A logistic regression method was applied to quantify the strength of associations of factors associated with the awareness index.

**Results:** Of total participants, 33% were from Manggarai, 32.3% were from Atoni, 30.2% from Sumba ethnicity. The level of MAI was significantly different between these groups with the highest in Manggarai ethnicity (65.1%, 95% confidence interval (CI): 59.9 – 70.3) and the lowest in Sumba ethnicity (35%, 95% CI: 27.6 – 42.4). The most prominent factors influencing the MAI in Sumba and Manggarai ethnicity were education level, whilst it was socio-economic status (SES) in Atoni ethnicity. The level of MAI was significantly higher for adults with diploma or above education level (adjusted odds ratio (AOR): 21.4, 95% CI: 3.59 – 127.7-for Manggarai ; AOR: 6.94, 95% CI: 1.81 – 26.6 for Sumba). The level of MAI was significantly higher for adults living in high SES in Atoni (AOR 24.48, 95% CI: 8.79 – 68.21).

**Conclusions:** Poorer education levels and low SES were more prominent factors contributing to lower levels of MAI in rural ENTP. Interventions should focus on improving malaria awareness to these groups to support the national commitment of the Indonesian government to achieve malaria elimination zone by 2030.

## INTRODUCTION

Malaria is a major health challenge globally across 87 countries and a leading cause of illness and death in many poor, tropical and sub-tropical countries [1,2]. The World Health Organization (WHO) estimated that in 2019 the number of malaria cases and death was 229 million and 409,000 respectively [2]. Almost 2.8% of cases are from South East Asia (SEA) countries of which 11% were from Indonesia [2].

In Indonesia, the Ministry of Health through the National Malaria Control Program (NMCP) has implemented various interventions to reduce malaria cases with the national commitment to achieve a malaria-free zone in the country by 2030 [3]. The funding for a malaria program was also increased by about 2.4% from US$21,170,034.7 in 2016 to US$21,683,909.81 in 2018 [3]. However, positive cases of malaria are still high vary across different provinces of Indonesia, such as 216,380 cases in Papua, 861 cases in South Sulawesi and 12 cases in Yogyakarta Special Region province [4]. In total, there were 250,644 cases in 2019 with almost 97% were in the eastern part of the country including in the East Nusa Tenggara Province (ENTP) [4].

The ENTP is a province known as “Flobamorata”, comprising of five major islands: Flores, Sumba, Timor, Alor, and Lembata [5]. The total number of malaria cases reported in 2018 was 17,192 cases from these islands with 70% of the cases contributed to Sumba and 10% Timor [6]. Although a number of studies were conducted on malaria in the ENTP [7-9], most of the investigations on the human aspect had not documented the malaria awareness which is potential to play a significant role in achieving and maintaining malaria elimination [10]. Having high malaria awareness of a community allows them to increase the use of insecticide treated nets to protect from mosquito bites [11], reduce malaria prevalence, and speed up towards malaria elimination [12].

Several studies have been undertaken to investigate malaria awareness globally [13-15]. Most of these studies focus on female participants [13,14]. One population-based study representing rural communities conducted in China in 2016 revealed malaria awareness was higher in males compared to females [15]. In SEA, some population-based studies were conducted to investigate malaria awareness [16-18] in which male and female distribution was not equivalent. In India [16] and Bangladesh [17] the majority of respondents were male whilst, in Myanmar [18] most of the respondents were female. However, the potential factors associated with malaria awareness in rural communities representing a balanced voice of male and female is less documented in this region. In fact, the burden of malaria was higher in rural areas compared with urban areas in SEA [19].

In an Indonesian context, a country with 145 ethnic groups [20], some studies were conducted at a village level to evaluate the association between malaria awareness and socio-demographic factors [21,22]. One population-based study in a rural community in Indonesia reported a low level of malaria awareness [23], however the diversity of ethnicity in rural community and their association with malaria awareness were not investigated yet. To date, there have been no studies investigating factors associated with malaria awareness in different ethnic groups of rural population in Indonesia. The investigation of predictors of malaria awareness in rural communities in Indonesia is critical given the fact that the burden of malaria in rural area is higher compared with urban areas in the country [3]. Understanding factors contributing to the malaria awareness of these communities will help the local authorities to allocate resources to strengthen the most vulnerable groups in the community and to reduce malaria awareness disparity amongst different ethnic groups in the community. Therefore, this research was undertaken to fill this gap with a dual aim for investigating ethnic variations, and their association with malaria awareness of rural adults in three different ethnic groups to support national commitment of the Indonesia government to eliminate malaria by 2030 [3].

## METHODS

### Study population and sampling

This study was based on data obtained from a cross-sectional study in rural areas of ENTP in Indonesia from October to December (2019). Data collection was conducted in 3 out of 22 districts in the province [5]. Firstly, East Sumba district was categorised as high malaria endemic settings (MES), where the majority of the population was of Sumba ethnicity [5,20,24]. Secondly, Belu district was classified as moderate MES with most of the population of Atoni ethnicity [5,20,24]. Finally, East Manggarai district was categorised as low MES with much of the population of Manggarai ethnicity [5,20,24]. These three ethnic groups are the main ethnic groups that dominated population in the province [20]. Participants were recruited using a multi-stage cluster random sampling procedure with the systematic random sampling procedure at a cluster level 4 with 49 villages from three districts of ENTP. In each village, 20 to 40 participants were selected proportionate to the population size of the village and the selection of participants was done by applying the systematic random sampling methods.

### Sample size calculation

The sample consists of 1495 participants aged 18 to 90 years from the general population in ENTP. The sample size calculation was based on the prevalence of malaria in ENTP, 1.99% in 2018 estimated by the Indonesia Health Ministry [25]. The comprehensive calculation for this sample size has been previously presented in a prior publication [26]. Overall, the sample size was sufficiently large enough to detect a minimum 5% difference in the proportion of malaria awareness amongst high, moderate, and low malaria endemic settings with the statistical power of at least 90% with a type I error of 5%.

### Outcome variable

The outcome variable of the study was the malaria awareness of participants. The awareness was evaluated by ten questions including basic understanding of malaria, main symptoms, transmission mode, prevention measures, and seeking malaria treatment within 24 hours when they suffer from malaria. For each question, the correct answer was assigned at score of one, and the incorrect answer was marked as zero. Therefore the total score ranged from zero to ten. Scores of malaria knowledge were evaluated as follows: participants having score ≥8 were classified as excellent, 6 or 7 as good, 1 to 5 as poor and 0 were classified as zero level of malaria knowledge. Participants who were in the group of excellent and good score were categorised as having malaria awareness whilst for those who were in poor, and zero score was classified as unware of malaria [15,27]. The computation of the Malaria Awareness Index was conducted for each ethnicity independently.

### Socio-demographic and environmental factors by ethnic groups

Ethnicity was classified as Sumba, Atoni, Manggarai, and other ethnicities. For other ethnicities, it was combining many tribes including Sabu, Maumere, Nagakeo, Javaness, Bima, and Bugis. Their number was none significant in this study, therefore they were classified as other ethnicities. In each ethnic group, demographic information comprising gender, age, education level, main occupation, family size, household income, and socio-economic status (SES) was collected. Gender was categorised as male or female. Age was classified as 5 groups, less than 30 years old, 30 to 40 years, 40 to 50 years, 50 to 60 years and above 60 years of age. The level of education was categorised as no education, primary school level of education (grade 1 to 6), junior high school (grade 7 to 9), senior high school (grade 10 to 12), and diploma or above education level [25]. Main occupation was classified as farmer, housewife, entrepreneur, government or non-government workers, and others [25]. The SES group assessed according to the ownership of durable assets and housing characteristics was categorized as low, moderate, and high SES. The nearest health facilities were classified as village maternity posts, village health post, public health centres (PHC), and subsidiary PHC [28]. Distance to the nearest health facilities was categorized as < 1 kilo metres (KM), 1-2 KM, 2-3 KM, and ≥ 3 KM.

### Statistical Analyses

The participants’ sociodemographic, and environmental characteristics (including gender, age group, education level, SES, family size, the nearest health facilities, and distance to the nearest health facilities) in each ethnicity were reported using descriptive statistics. The association of malaria awareness with all covariates were evaluated using the chi-square test. The logistic regression method was applied to investigate the factors associated with malaria awareness among adults in rural ENTP. Three models of odd ratio (OR) with 95% confidence interval (CI) were developed; crude OR (95% CI), age and sex adjusted OR (95% CI), adjusted OR (95% CI) in a full multiple logistic regression model. The P value of 0.05 or less was deemed to be statistically significant. All analyses were conducted using the Statistical Package for Social Science (SPSS) version 27.

### Ethical Statement

This research was approved by Human Ethics Committee of the Swinburne University of Technology (reference number 20191428-1490) and the Indonesian Ministry of Health (reference letter: LB.02.01/2/KE.418/2019).

## RESULTS

### Socio-demographic characteristics of study participants by ethnic groups

In the total sample, approximately equal participants were from Manggarai (33.0%), Atoni (32.3%), and Sumba ethnicity (30.2%). It was only 4.5% from other ethnicities. The composition of participants was significantly different demographically for each ethnicity except for gender. The proportion of rural adults having no education in Manggarai ethnicity was the lowest (2.60%) of all ethnicities, whilst it was the highest in Sumba ethnicity (36.1%). Most of participants in Sumba and Manggarai ethnicity, 72.1% and 65.9% respectively were farmers, meanwhile it was housewife in Atoni ethnicity (47.6%). The majority of participants in each ethnicity was living in moderate socio-economic status as shown in Table 1.

**Table 1:** Socio-demographic characteristics of study participants in the East Nusa Tenggara Province, Indonesia (N = 1495).

| Characteristic | Ethnic Groups, n(%) |  |  |  | Total | P value |
| --- | --- | --- | --- | --- | --- | --- |
|  | Sumba | Atoni | Manggarai | Others |  |  |
| **Overall** | 452 (30.2) | 483 (32.3) | 493 (33.0) | 67 (4.5) | 1495 |  |
| **Gender** |  |  |  |  |  |  |
| Female | 232 (51.3) | 260 (53.8) | 234 (47.5) | 42 (62.7) | 768 (51.4) | 0.055 |
| Male | 220 (48.7) | 223 (46.2) | 259 (52.5) | 25 (37.3) | 727 (48.6) |  |
| **Age Group** |  |  |  |  |  |  |
| < 30 | 71 (15.7) | 63 (13.0) | 59 (12.0) | 12 (17.9) | 205 (13.7) | < 0.001 |
| 30 – 39 | 131 (29.0) | 100 (20.7) | 172 (34.9) | 15 (22.4) | 418 (28.0) |  |
| 40 – 49 | 123 (27.2) | 121 (25.1) | 110 (22.3) | 17 (25.4) | 371 (24.8) |  |
| 50 – 59 | 60 (13.3) | 128 (26.5) | 94 (19.1) | 13 (19.4) | 295 (19.7) |  |
| ≥ 60 | 67 (14.8) | 71 (14.7) | 58 (11.8) | 10 (14.9) | 206 (13.8) |  |
| **Level of education** |  |  |  |  |  |  |
| No Education | 163 (36.1) | 89 (18.4) | 13 (2.60) | 14 (20.9) | 279 (18.7) | < 0.001 |
| Primary School | 192 (42.5) | 199 (41.2) | 264 (53.5) | 23 (34.3) | 678 (45.4) |  |
| Junior High School | 40 (8.85) | 93 (19.3) | 86 (17.4) | 10 (14.9) | 229 (15.3) |  |
| Senior High School | 43 (9.51) | 82 (17.0) | 73 (14.8) | 12 (17.9) | 210 (14.0) |  |
| Diploma or above | 14 (3.10) | 20 (4.14) | 57 (11.6) | 8 (11.9) | 99 (6.62) |  |
| **Occupation** |  |  |  |  |  |  |
| Housewife | 70 (15.5) | 230 (47.6) | 76 (15.4) | 27 (40.3) | 403 (27.0) | < 0.001 |
| Farmer | 326 (72.1) | 160 (33.1) | 325 (65.9) | 20 (29.9) | 831 (55.6) |  |
| Entrepreneurs | 28 (6.19) | 3 (0.62) | 12 (2.43) | 5 (7.46) | 48 (3.21) |  |
| Other | 14 (3.10) | 38 (7.87) | 5 (1.01) | 5 (7.46) | 62 (4.15) |  |
| Govt. or non-govt. employment | 14 (3.10) | 52 (10.8) | 75 (15.2) | 10 (14.9) | 151 (10.1) |  |
| **Socio-Economic Status** |  |  |  |  |  |  |
| Low | 146 (32.3) | 103 (21.3) | 192 (38.9) | 8 (11.9) | 449 (30.0) | < 0.001 |
| Average | 263 (58.2) | 318 (65.8) | 239 (48.5) | 40 (59.7) | 860 (57.5) |  |
| High | 43 (9.51) | 62 (12.8) | 62 (12.6) | 19 (28.4) | 186 (12.4) |  |
| **Family size** |  |  |  |  |  |  |
| ≤ 4 | 187 (41.4) | 347 (71.8) | 235 (47.7) | 34 (50.7) | 803 (53.7) | < 0.001 |
| > 4 | 265 (58.6) | 136 (28.2) | 258 (52.3) | 33 (49.3) | 692 (46.3) |  |
| **‡HH Income in relation to PMW** |  |  |  |  |  |  |
| < PMW | 427 (94.5) | 439 (90.9) | 422 (85.6) | 54 (80.6) | 1342 (89.8) | < 0.001 |
| ≥ PMW | 25 (5.53) | 44 (9.11) | 71 (14.4) | 13 (19.4) | 153 (10.2) |  |
| **The nearest Health Service** |  |  |  |  |  |  |
| Village maternity posts | 128 (28.3) | 218 (45.1) | 16 (3.25) | 24 (35.8) | 386 (25.8) | < 0.001 |
| Village health Post | 32 (7.08) | 101 (20.9) | 164 (33.3) | 5 (7.46) | 302 (20.2) |  |
| Subsidiary PHC | 170 (37.6) | 53 (11.0) | 108 (21.9) | 7 (10.4) | 338 (22.6) |  |
| Public Health centres (PHC) | 122 (27.0) | 111 (23.0) | 205 (41.6) | 31 (46.3) | 469 (31.4) |  |
| **Distance to the Nearest Health Facilities** |  |  |  |  |  |  |
| < 1 Km | 164 (36.3) | 259 (53.6) | 132 (26.8) | 23 (34.3) | 578 (38.7) | < 0.001 |
| 1 – 2 Km | 79 (17.5) | 145 (30) | 164 (33.3) | 13 (19.4) | 400 (26.8) |  |
| 2 – 3 Km | 54 (11.9) | 53 (11.0) | 86 (17.4) | 10 (14.9) | 204 (13.6) |  |
| ≥ 3 Km | 155 (34.3) | 26 (5.38) | 111 (22.5) | 21 (31.3) | 313 (20.9) |  |

‡The Provincial Minimum Wages (PMW) in 2019 is IDR 1,795,000 monthly.

### Awareness of malaria by ethnic groups

Malaria awareness index (MAI) by ethnicity was presented in Table 2. Overall, MAI among ethnicities was significantly different (p value < 0.001). The highest MAI was in Manggarai ethnicity, accounting for 65.1% (95% confidence interval [CI]: 59.9 – 70.3), whilst it was the lowest in Sumba ethnicity (35%, 95% CI: 27.6 – 42.4). The MAI in Atoni and other ethnicities was also poor, accounting for 43.7%; 95% CI: 37.0 – 50.4, and 59.7%; 95% CI: 44.5 – 74.9, respectively.

**Table 2:**
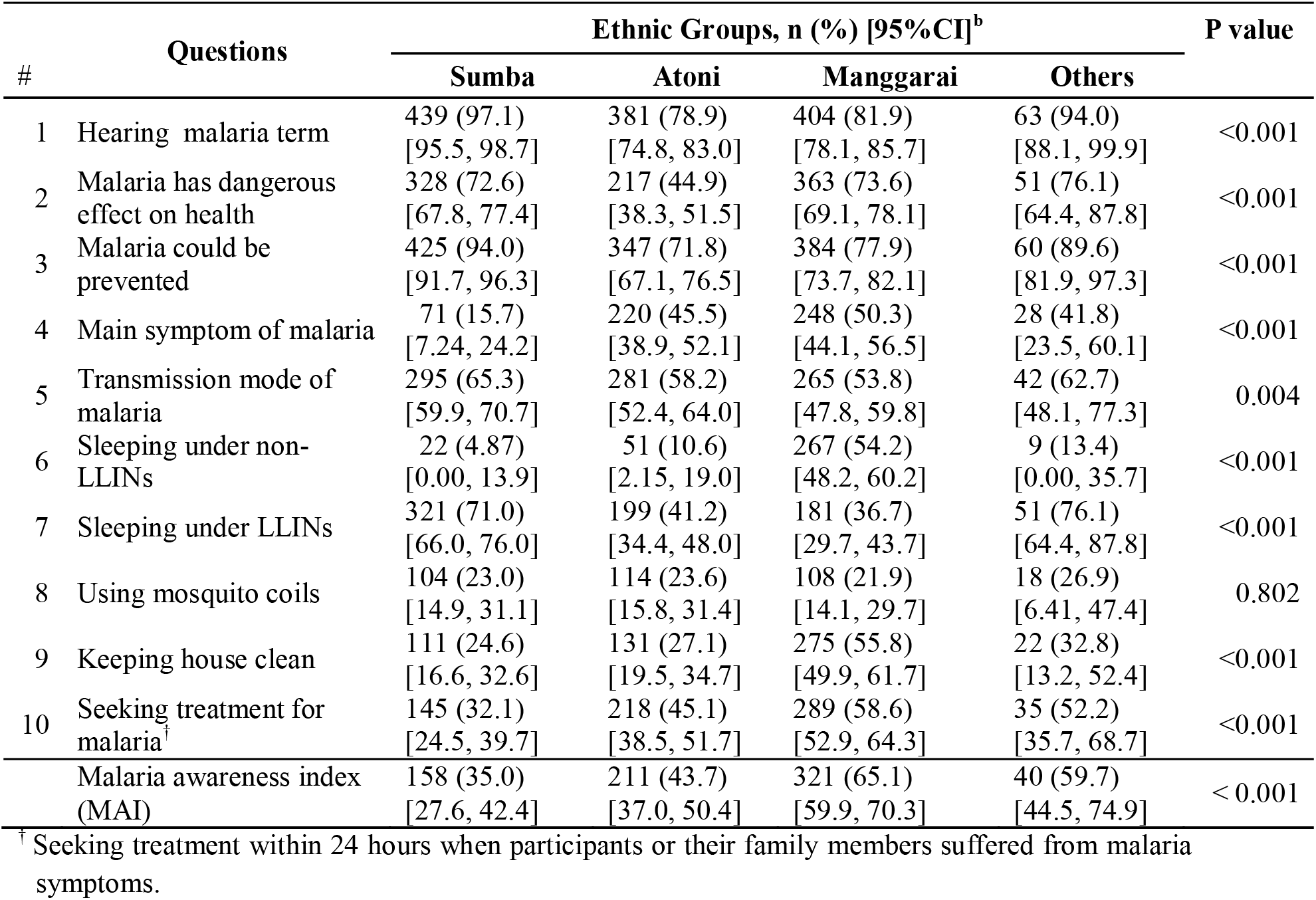
Distribution of malaria knowledge of rural adults in different ethnicities in the East Nusa Tenggara Province (ENTP), Indonesia.

### Awareness of malaria by different socio-demographic characteristics and environmental characteristics in each ethnicity

Malaria awareness index (MAI) by socio-demographic factors and environmental factors for each ethnicity was presented in Table 3. The MAI between male and female was significantly different in Atoni (p < 0.001) and Sumba (p = 0.046) ethnicity. In all ethnicities, except in other ethnicity, the MAI was significantly different amongst education level of participants and there was a trend in malaria awareness as the level of education was higher. In terms of main occupation, the MAI for housewife was the lowest of all other occupations for both Sumba and Atoni tribe, accounting for 32.9% and 34.3% respectively. The MAI between socio-economic status (SES) was different significantly particularly in Sumba and Atoni ethnicity and the MAI increased as SES was higher.

**Table 3:** Variation of Malaria awareness index (MAI) by socio-demographic and environmental factors of rural adults in East Nusa Tenggara Province Indonesia.

| Characteristic | Ethnic Groups |  |  |  |  |  |  |  |
| --- | --- | --- | --- | --- | --- | --- | --- | --- |
|  | Sumba |  | Atoni |  | Manggarai |  | Others |  |
|  | No at risk | MAI | No at risk | MAI | No at risk | MAI | No at risk | MAI |
| Total | 452 | 158 (35.0) | 483 | 211 (43.7) | 493 | 321 (65.1) | 67 | 40 (59.7) |
| **Gender** |  |  |  |  |  |  |  |  |
| Female | 232 | 71 (30.6) | 260 | 93 (35.8) | 234 | 152 (65.0) | 42 | 26 (61.9) |
| Male | 220 | 87 (39.5) | 223 | 118 (52.9) | 259 | 169 (65.3) | 25 | 14 (56.0) |
| P value |  | 0.046 |  | < 0.001 |  | 0.946 |  | 0.634 |
| **Age Group** |  |  |  |  |  |  |  |  |
| < 30 | 71 | 23 (32.4) | 63 | 31 (49.2) | 59 | 33 (55.9) | 12 | 5 (41.7) |
| 30 – 39 | 131 | 48 (36.6) | 100 | 56 (56.0) | 172 | 120 (69.8) | 15 | 11 (73.3) |
| 40 – 49 | 123 | 50 (40.7) | 121 | 53 (43.8) | 110 | 83 (75.5) | 17 | 12 (70.6) |
| 50 – 59 | 60 | 22 (36.7) | 128 | 42 (32.8) | 94 | 57 (60.6) | 13 | 7 (53.8) |
| > 60 | 67 | 15 (22.4) | 71 | 29 (40.8) | 58 | 28 (48.3) | 10 | 5 (50.0) |
| P value |  | 0.144 |  | 0.01 |  | 0.002 |  | 0.38 |
| **Level of Education** |  |  |  |  |  |  |  |  |
| No education | 163 | 36 (22.1) | 89 | 34 (38.2) | 13 | 5 (38.5) | 14 | 6 (42.9) |
| Primary school | 192 | 70 (36.5) | 199 | 56 (28.1) | 264 | 157 (59.5) | 23 | 11 (47.8) |
| Junior High School | 40 | 16 (40.0) | 93 | 56 (60.2) | 86 | 62 (72.1) | 10 | 7 (70.0) |
| Senior High School | 43 | 26 (60.5) | 82 | 49 (59.8) | 73 | 51 (69.9) | 12 | 8 (66.7) |
| Diploma or above | 14 | 10 (71.4) | 20 | 16 (80.0) | 57 | 46 (80.7) | 8 | 8 (100.0) |
| P value |  | < 0.001 |  | < 0.001 |  | 0.002 |  | 0.059 |
| **Occupation** |  |  |  |  |  |  |  |  |
| Housewife | 70 | 23 (32.9) | 230 | 79 (34.3) | 76 | 41 (53.9) | 27 | 16 (59.3) |
| Farmer | 326 | 108 (33.1) | 160 | 64 (40.0) | 325 | 217 (66.8) | 20 | 9 (45.0) |
| Entrepreneurs | 28 | 12 (42.9) | 3 | 2 (66.7) | 12 | 6 (50.0) | 5 | 3 (60.0) |
| Other | 14 | 7 (50.0) | 38 | 27 (71.1) | 5 | 2 (40.0) | 5 | 3 (60.0) |
| Govt. or non-govt. staff | 14 | 8 (57.1) | 52 | 39 (75.0) | 75 | 55 (73.3) | 10 | 9 (90.0) |
| P value |  | 0.214 |  | < 0.001 |  | 0.052 |  | 0.230 |
| **Socio-Economic Status** |  |  |  |  |  |  |  |  |
| Poor | 146 | 35 (24.0) | 103 | 13 (12.6) | 192 | 132 (68.8) | 8 | 3 (37.5) |
| Average | 263 | 100 (38.0) | 318 | 149 (46.9) | 239 | 151 (63.2) | 40 | 21 (52.5) |
| Rich | 43 | 23 (53.5) | 62 | 49 (79.0) | 62 | 38 (61.3) | 19 | 16 (84.2) |
| P value |  | < 0.001 |  | < 0.001 |  | 0.385 |  | 0.027 |
| **†HH Income in relation to PMW** |  |  |  |  |  |  |  |  |
| < PMW | 427 | 140 (32.8) | 439 | 185 (42.1) | 422 | 264 (62.6) | 54 | 28 (51.9) |
| >= PMW | 25 | 18 (72.0) | 44 | 26 (59.1) | 71 | 57 (80.3) | 13 | 12 (92.3) |
| P value |  | < 0.001 |  | 0.031 |  | 0.004 |  | 0.008 |
| **Family size** |  |  |  |  |  |  |  |  |
| < = 4 | 187 | 74 (39.6) | 347 | 148 (42.7) | 235 | 143 (60.9) | 34 | 22 (64.7) |
| > 4 | 265 | 84 (31.7) | 136 | 63 (46.3) | 258 | 178 (69.0) | 33 | 18 (54.5) |
| P value |  | 0.084 |  | 0.464 |  | 0.058 |  | 0.397 |
| **Distance to the nearest health facilities** |  |  |  |  |  |  |  |  |
| < 1 Km | 164 | 55 (33.5) | 259 | 87 (33.6) | 132 | 104 (78.8) | 23 | 15 (65.2) |
| 1 – 2 Km | 79 | 24 (30.4) | 145 | 86 (59.3) | 163 | 108 (65.9) | 13 | 7 (53.8) |
| 2 – 3 Km | 54 | 20 (37.0) | 53 | 30 (56.6) | 87 | 56 (65.1) | 10 | 6 (60.0) |
| >=3 Km | 155 | 59 (38.1) | 26 | 8 (30.8) | 111 | 53 (47.7) | 21 | 12 (57.1) |
| P value |  | 0.652 |  | < 0.001 |  | < 0.001 |  | 0.911 |
| **The nearest health service** |  |  |  |  |  |  |  |  |
| Village health Post | 32 | 4 (12.5) | 101 | 21 (20.8) | 164 | 103 (62.8) | 5 | 4 (80.0) |
| Village maternity posts | 128 | 57 (44.5) | 218 | 89 (40.8) | 16 | 10 (62.5) | 24 | 13 (54.2) |
| Subsidiary Public Health centres | 170 | 43 (25.3) | 53 | 29 (54.7) | 108 | 71 (65.7) | 7 | 4 (57.1) |
| Public Health centres | 122 | 54 (44.3) | 111 | 72 (64.9) | 205 | 137 (66.8) | 31 | 19 (61.3) |
| P value |  | < 0.001 |  | < 0.001 |  | 0.869 |  | 0.750 |

†The Provincial Minimum Wages (PMW) in 2019 is IDR 1,795,000 monthly.

**Table 4.**
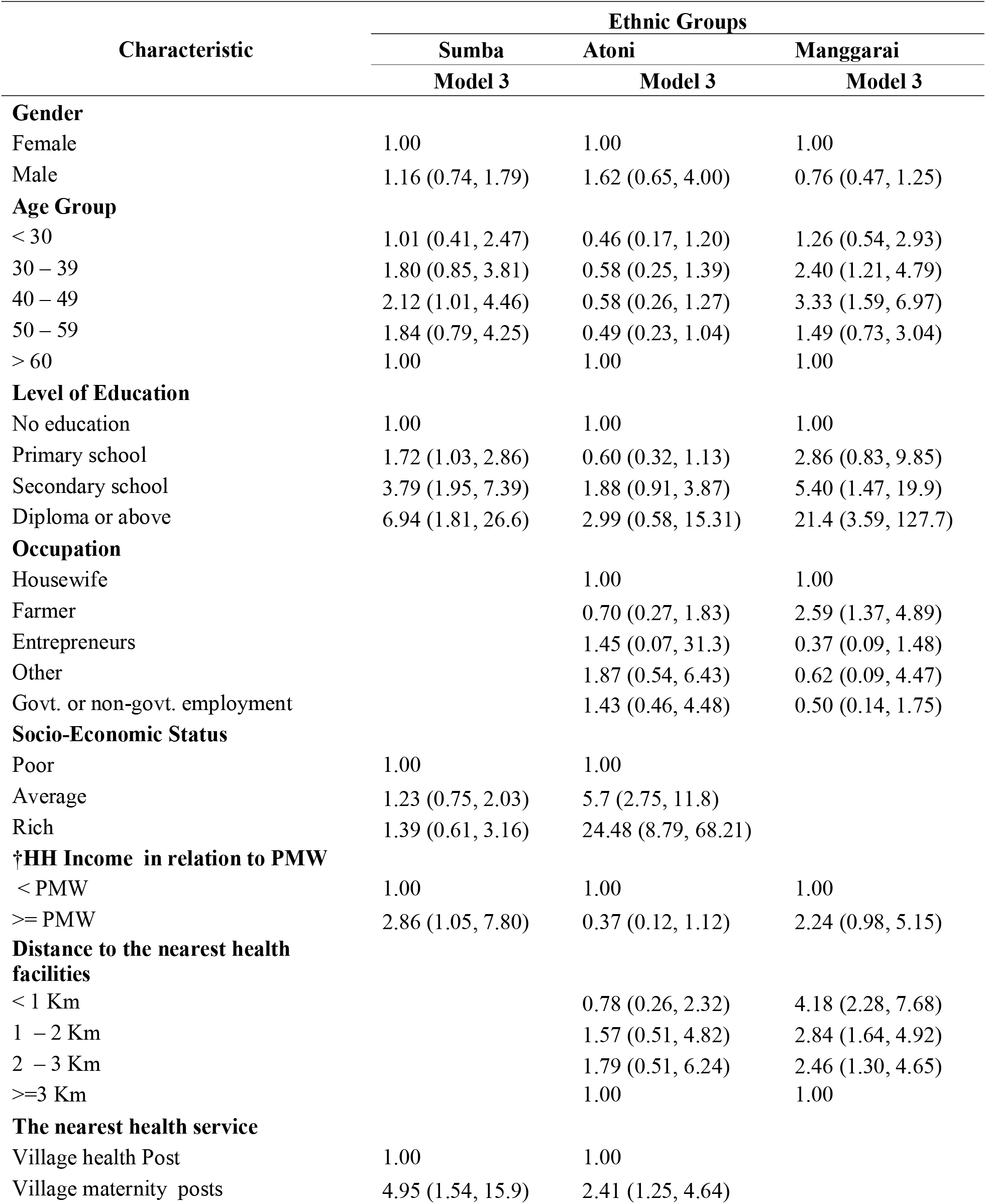
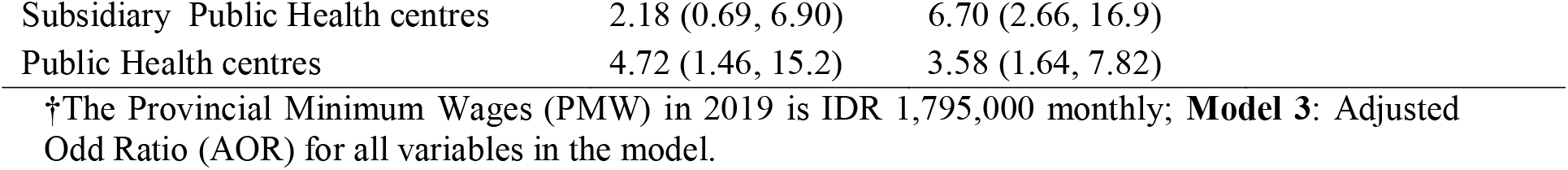
Multivariate binary logistic regression: factors associated with malaria awareness among rural adult in East Nusa Tenggara Province.

| Characteristic | Ethnic Groups |  |  |
| --- | --- | --- | --- |
|  | Sumba | Atoni | Manggarai |
|  | Model 3 | Model 3 | Model 3 |
| **Gender** |  |  |  |
| Female | 1.00 | 1.00 | 1.00 |
| Male | 1.16 (0.74, 1.79) | 1.62 (0.65, 4.00) | 0.76 (0.47, 1.25) |
| **Age Group** |  |  |  |
| < 30 | 1.01 (0.41, 2.47) | 0.46 (0.17, 1.20) | 1.26 (0.54, 2.93) |
| 30 – 39 | 1.80 (0.85, 3.81) | 0.58 (0.25, 1.39) | 2.40 (1.21, 4.79) |
| 40 – 49 | 2.12 (1.01, 4.46) | 0.58 (0.26, 1.27) | 3.33 (1.59, 6.97) |
| 50 – 59 | 1.84 (0.79, 4.25) | 0.49 (0.23, 1.04) | 1.49 (0.73, 3.04) |
| > 60 | 1.00 | 1.00 | 1.00 |
| **Level of Education** |  |  |  |
| No education | 1.00 | 1.00 | 1.00 |
| Primary school | 1.72 (1.03, 2.86) | 0.60 (0.32, 1.13) | 2.86 (0.83, 9.85) |
| Secondary school | 3.79 (1.95, 7.39) | 1.88 (0.91, 3.87) | 5.40 (1.47, 19.9) |
| Diploma or above | 6.94 (1.81, 26.6) | 2.99 (0.58, 15.31) | 21.4 (3.59, 127.7) |
| **Occupation** |  |  |  |
| Housewife |  | 1.00 | 1.00 |
| Farmer |  | 0.70 (0.27, 1.83) | 2.59 (1.37, 4.89) |
| Entrepreneurs |  | 1.45 (0.07, 31.3) | 0.37 (0.09, 1.48) |
| Other |  | 1.87 (0.54, 6.43) | 0.62 (0.09, 4.47) |
| Govt. or non-govt. employment |  | 1.43 (0.46, 4.48) | 0.50 (0.14, 1.75) |
| **Socio-Economic Status** |  |  |  |
| Poor | 1.00 | 1.00 |  |
| Average | 1.23 (0.75, 2.03) | 5.7 (2.75, 11.8) |  |
| Rich | 1.39 (0.61, 3.16) | 24.48 (8.79, 68.21) |  |
| **HH Income in relation to PMW** |  |  |  |
| < PMW | 1.00 | 1.00 | 1.00 |
| >= PMW | 2.86 (1.05, 7.80) | 0.37 (0.12, 1.12) | 2.24 (0.98, 5.15) |
| **Distance to the nearest health facilities** |  |  |  |
| < 1 Km |  | 0.78 (0.26, 2.32) | 4.18 (2.28, 7.68) |
| 1 – 2 Km |  | 1.57 (0.51, 4.82) | 2.84 (1.64, 4.92) |
| 2 – 3 Km |  | 1.79 (0.51, 6.24) | 2.46 (1.30, 4.65) |
| >=3 Km |  | 1.00 | 1.00 |
| **The nearest health service** |  |  |  |
| Village health Post | 1.00 | 1.00 |  |
| Village maternity posts | 4.95 (1.54, 15.9) | 2.41 (1.25, 4.64) |  |

|  |  |  |
| --- | --- | --- |
| Subsidiary Public Health centres | 2.18 (0.69, 6.90) | 6.70 (2.66, 16.9) |
| Public Health centres | 4.72 (1.46, 15.2) | 3.58 (1.64, 7.82) |

†The Provincial Minimum Wages (PMW) in 2019 is IDR 1,795,000 monthly; **Model 3:** Adjusted Odd Ratio (AOR) for all variables in the model.

### Factors associated with malaria awareness

After adjustment for all covariates, the following factors were associated with a low level of malaria awareness in Sumba ethnicity of ENTP: diploma or above education level compared to those with no education level (Adjusted odds ratio (AOR) 6.94, 95% confidence interval (CI): 1.81 – 26.6); living closest to village maternity post compared to those living closed to village health post (AOR 4.95, 95% CI: 1.54 – 15.9). Whilst in Atoni ethnicity, the MAI of rural adults in high SES was 24 times higher than those were in low SES (AOR 24.48, 95% CI: 8.79 – 68.21) and the MAI of participants living closest to Subsidiary Public Health centres was almost 7 times higher than those were around village health post (AOR 6.70, 95% CI: 2.66 – 16.9). Finally, in Manggarai ethnicity: diploma or above education level compared to those no education level (AOR 21.4, 95% CI: 3.59 – 127.7); living less than 1 kilometres from the nearest health facilities compared to those were living more than three kilometres from the closest health facilities (AOR: 4.18, 95% CI: 2.28 – 7.68); working as farmer compared to those as housewife (AOR: 2.59, 95% CI: 1.37 – 4.89).

## DISCUSSION

To our knowledge this is the first study focused on the diversity of ethnic groups in relation to malaria awareness in rural adults of Indonesia. The results shown that malaria awareness among these three groups was significantly different. The highest malaria awareness was in Manggarai ethnicity, whilst it was the lowest in Sumba ethnicity. The most important factors in malaria awareness in this rural community was education level. This was found particularly in Sumba and Manggarai ethnic group. The higher the education level of participants, the higher their malaria awareness. Whilst in Atoni ethnic group, the prominent factors associated with malaria awareness was SES of participants. The distance to the nearest health facilities, the nearest health facilities and main occupation were also associated with the awareness.

This study demonstrated that there was a significant discrepancy on malaria awareness among ethnicity. This result confirmed findings in other countries such as China [15] and Malawi [14] indicating that the disparity of malaria awareness existed amongst ethnicities in a community. The study shown that malaria awareness in Manggarai ethnicity was the highest, whereas it was the lowest in Sumba ethnicity. The disparity amongst ethnicities revealed the variation in terms of socio-demographic characteristic of those ethnicities [14]. In this study, the huge disparity of malaria awareness amongst ethnicities might be attributed to the different levels of education amongst ethnicities. This study shows that the proportion of adults with primary or no education in Sumba ethnicity was higher (79%) than that of in Manggarai ethnicity (56%). These situations might contribute to the poor health literacy in the community of Sumba ethnicity. The improvement of health literacy of community allows them to improve their understanding and application of the complexity of various health information on individual and population level [29].

This study shown that the improvement of malaria awareness was in line with the increase education level of participants particularly in Sumba and Manggarai ethnic group. This findings confirm the behavioural aspect of malaria studies in other rural settings and other countries such as Malawi [14], India [16], and Bangladesh [17] revealing that education level positively correlated with malaria awareness. This study indicated that in Manggarai ethnicity, malaria awareness of participants with at least a diploma level education was 21 times higher compared with those having no education and in Sumba ethnicity, it was almost seven times higher compared with those having no education. This looks like people with higher education have many chance to access multiple source of information [30] and they have capability to understand written information [31] supporting them to distinguish various terms of malaria. This study has shown only about 7% of rural adults in these ethnicities having diploma or above education level. This has made them particularly disadvantaged gaining knowledge of malaria and needs more attention from authority for spreading malaria education message to boost malaria elimination program in the region. Higher education is associated with high awareness of any kinds of diseases, this is established true [32].

In this study, we found that malaria awareness was significantly associated with the distance to the nearest health facilities particularly in Manggarai ethnicity. The reduction of malaria awareness was in line with the improving of distance to the nearest health facilities. This finding was consistent with studies in rural communities in China [15] and Malawi [33]. This situation may be attributed to health promotion education does not work properly in this province since the shortage of health workers in public health centres (PHC). Basic database of PHC in the ENTP revealed that the distribution of health workers in 381 public health centres is uneven ranging from 0 to 183 staff in each PHC and 33% of the total number of PHC does not be supported by the availability of general practitioner (GP) [34]. The total number of GPs in the province is 861 (indicating that one GP for 6,000 people) to provide malaria treatment [6].

Our study has further shown that the disparity of malaria awareness was evident amongst gender of participants particularly in Sumba and Altona ethnicity. Malaria awareness in male adults was higher compared with that of in female adults. This finding was consistent with studies on malaria awareness of rural communities in China [15] and Bangladesh [17]. Furthermore, this study also shows that the MAI of those working as housewife was the lowest of all other occupations. The low level of malaria awareness for females in the ENTP is unexpected by the authors because the Indonesia government has provided informative systems allowing women to improve their malaria knowledge. For instance, the distribution of long-lasting insecticide-treated nets (LLINs) was integrated with immunization and Vitamin A distribution activity at community health centres [35]. All those activities have been run mainly by females and the participants on those activities have been mostly female. Improving malaria awareness for women in the region is critical considering that they are role models for raising malaria awareness at home [36].

The finding of this study indicated that adults from low level of socio-economic status (SES) and low income had low level of malaria awareness. This is in line with study in Malawi [14] and Bangladesh [17]. This association might be due to the fact that adults from low SES were struggling to access health facilities leads to poor health information [37].

Community participation provides a significant role to progress towards malaria elimination [38]. To encourage the community participation actively, they must have a high awareness of the disease. This study has demonstrated the low level of malaria awareness of rural communities regardless of gender and age group. The improvement of malaria awareness of this community is critical to progress in malaria elimination. Promoting equal access to a malaria education program will reduce the gap on malaria awareness amongst level of the group. Finally, our study has shown that the Sumba ethnicity, possibly due to their low level of education was the most vulnerable group that should be prioritized to improve their malaria awareness. Given the low level of education those who are unable to read written posters or pamphlets and the high rate of children dropping out of school in the rural population of this province [39], the announcement of malaria education communication by loudspeaker might be suitable for this rural community as shown in other countries [40].

The present study applies a large sample size that includes adult aged 18 to 89 years of three different minority groups of Indonesia and comparative presentation of data allowing comprehensive understanding of malaria awareness in rural communities. However, there are some limitations of the study including data were collected over one period which was not possible for authors to follow-up with the participants to assess their perception change. Data was also merely captured from minority ethnic groups of one province which is hard to present the diversity of minority ethnic groups of Indonesia. The study needs to be repeated randomly in other parts of the country to capture a truly representative sample of minority groups of rural adults in terms of malaria awareness at the national level.

In conclusion, this study shows that malaria awareness is low and the disparity of malaria awareness among ethnicity was found in rural ENTP. The risk factors associated with the low level of this awareness included low levels of education, low socio-economic status, working as housewife, living more than three kilometres from the nearest health facility. Disparity on malaria awareness amongst gender, age group, education level, occupation, and distance to the nearest health facility was statistically significant within each ethnicity. Public health programs should focus on improving malaria awareness on these vulnerable groups and promoting equal access on a malaria awareness campaign to reduce the disparity of awareness amongst the group. Interventions to improve malaria awareness will support government’s expectation to achieve malaria free-zone by 2030.

## Data Availability

All data produced in the present study are available upon reasonable request to the authors

## SUPPLEMENTAL MATERIALS

Dataset: Database for study Ethnic variation and its association with Malaria Awareness in East Nusa Tenggara Province Indonesia.

## ACKNOWLEDGMENTS

We would like to thank the Australia Awards Scholarship for supporting this research and the Faculty of Health, Arts, and Design of the Swinburne University Technology for providing funding for primary data collection for this study. The funders had no role in the designing of the study, data collection, analysis, or interpretation of data, or writing the paper. We further would like to express our gratitude to the Health Ministry of Indonesia, the governor of ENTP, head of East Sumba, Belu, and East Manggarai district, nine head of sub-districts, and forty-nine village leaders for allowing conducted this research in their region.

## AUTHOR CONTRIBUTIONS

Conceptualization: RDG. Data curation: RDG. Formal analysis: RDG. Funding acquisition: RDG. Methodology: RDG. Project administration: RDG. Writing - original draft: RDG. Writing - review & editing: RDG, JK, FMAI.

