## Supplemental Table for "Ethnic Variation and its Association with Malaria Awareness: A Cross-sectional Study in the East Nusa Tenggara Province (Indonesia)"

| # | Res_ID | MES | G | Edu_1 | AG | SES | FZ | DTHF | HFC | IMWP | IIPL | Job | Edu2 | Eth | Q10 | Q1 | Q2 | Q3 | Q4 | Q5 | Q6 | Q7 | Q8 | Q9 | MKS | MA |
|---|---|---|---|---|---|---|---|---|---|---|---|---|---|---|---|---|---|---|---|---|---|---|---|---|---|---|
| 1 | 10111 | 3 | 1 | 3 | 4 | 3 | 2 | 2 | 1 | 1 | 1 | 2 | 2 | 3 | 1 | 1 | 1 | 1 | 1 | 1 | 1 | 0 | 1 | 0 | 4 | 1 |
| 2 | 10112 | 3 | 1 | 5 | 2 | 3 | 1 | 2 | 1 | 0 | 0 | 5 | 3 | 3 | 1 | 1 | 1 | 1 | 1 | 1 | 0 | 1 | 0 | 0 | 3 | 1 |
| 3 | 10113 | 3 | 1 | 2 | 3 | 2 | 1 | 2 | 1 | 0 | 0 | 2 | 1 | 3 | 1 | 1 | 1 | 1 | 1 | 1 | 0 | 1 | 0 | 0 | 3 | 1 |
| 4 | 10114 | 3 | 1 | 5 | 1 | 2 | 1 | 2 | 1 | 0 | 0 | 5 | 3 | 3 | 1 | 1 | 1 | 1 | 1 | 1 | 0 | 1 | 1 | 1 | 4 | 1 |
| 5 | 10115 | 3 | 1 | 3 | 4 | 2 | 1 | 2 | 1 | 1 | 1 | 5 | 2 | 3 | 1 | 1 | 1 | 1 | 0 | 1 | 1 | 0 | 0 | 0 | 3 | 1 |
| 6 | 10116 | 3 | 1 | 3 | 3 | 2 | 1 | 2 | 1 | 0 | 0 | 2 | 2 | 3 | 1 | 1 | 1 | 1 | 1 | 1 | 0 | 0 | 1 | 0 | 3 | 1 |
| 7 | 10117 | 3 | 1 | 5 | 1 | 2 | 1 | 2 | 1 | 0 | 0 | 2 | 3 | 3 | 1 | 1 | 1 | 1 | 1 | 1 | 0 | 1 | 1 | 1 | 4 | 1 |
| 8 | 10118 | 3 | 1 | 4 | 3 | 2 | 1 | 2 | 1 | 1 | 0 | 5 | 2 | 3 | 1 | 1 | 1 | 1 | 1 | 1 | 0 | 0 | 1 | 1 | 4 | 1 |
| 9 | 10119 | 3 | 1 | 3 | 3 | 2 | 1 | 2 | 1 | 0 | 0 | 2 | 2 | 3 | 1 | 1 | 1 | 1 | 1 | 1 | 0 | 0 | 0 | 1 | 3 | 1 |
| 10 | 101110 | 3 | 1 | 2 | 4 | 2 | 1 | 2 | 1 | 0 | 0 | 2 | 1 | 3 | 1 | 1 | 0 | 1 | 1 | 1 | 0 | 0 | 1 | 0 | 3 | 1 |
| 11 | 101111 | 3 | 1 | 2 | 3 | 2 | 2 | 2 | 1 | 0 | 0 | 2 | 1 | 3 | 1 | 1 | 0 | 1 | 1 | 1 | 0 | 1 | 0 | 0 | 3 | 1 |
| 12 | 101112 | 3 | 1 | 5 | 1 | 2 | 1 | 2 | 1 | 1 | 0 | 5 | 3 | 3 | 1 | 1 | 1 | 1 | 1 | 1 | 0 | 1 | 1 | 0 | 4 | 1 |
| 13 | 101113 | 3 | 1 | 4 | 4 | 2 | 1 | 2 | 2 | 1 | 1 | 5 | 2 | 3 | 1 | 1 | 0 | 1 | 1 | 1 | 0 | 0 | 1 | 0 | 3 | 1 |
| 14 | 101114 | 3 | 1 | 5 | 5 | 3 | 1 | 2 | 1 | 1 | 1 | 5 | 3 | 3 | 1 | 1 | 1 | 1 | 1 | 1 | 0 | 0 | 1 | 0 | 3 | 1 |
| 15 | 101115 | 3 | 0 | 1 | 5 | 1 | 1 | 2 | 1 | 0 | 0 | 2 | 1 | 3 | 1 | 0 | 0 | 0 | 0 | 0 | 0 | 0 | 0 | 0 | 2 | 0 |
| 16 | 101116 | 3 | 0 | 3 | 2 | 2 | 2 | 1 | 1 | 0 | 0 | 1 | 2 | 3 | 1 | 1 | 1 | 1 | 0 | 1 | 0 | 0 | 1 | 0 | 3 | 1 |
| 17 | 101117 | 3 | 1 | 5 | 3 | 3 | 1 | 1 | 1 | 1 | 1 | 5 | 3 | 3 | 0 | 1 | 1 | 1 | 1 | 1 | 1 | 0 | 1 | 0 | 3 | 1 |
| 18 | 101118 | 3 | 1 | 3 | 4 | 2 | 1 | 1 | 1 | 0 | 0 | 2 | 2 | 3 | 1 | 1 | 1 | 1 | 1 | 1 | 0 | 0 | 1 | 1 | 4 | 1 |
| 19 | 101119 | 3 | 1 | 3 | 2 | 2 | 2 | 1 | 1 | 0 | 0 | 2 | 2 | 3 | 1 | 1 | 1 | 1 | 0 | 1 | 0 | 0 | 0 | 1 | 3 | 1 |
| 20 | 101120 | 3 | 0 | 2 | 4 | 1 | 1 | 1 | 1 | 0 | 0 | 1 | 1 | 3 | 1 | 1 | 0 | 1 | 0 | 1 | 0 | 0 | 0 | 0 | 2 | 0 |
| 21 | 101121 | 3 | 1 | 3 | 3 | 1 | 1 | 1 | 1 | 0 | 0 | 2 | 2 | 3 | 1 | 1 | 1 | 1 | 0 | 1 | 0 | 0 | 0 | 0 | 2 | 0 |
| 22 | 101122 | 3 | 0 | 1 | 4 | 1 | 1 | 1 | 1 | 0 | 0 | 1 | 1 | 3 | 1 | 1 | 1 | 1 | 0 | 1 | 1 | 0 | 0 | 0 | 3 | 1 |
| 23 | 101123 | 3 | 0 | 3 | 4 | 2 | 1 | 1 | 1 | 0 | 0 | 1 | 2 | 3 | 1 | 1 | 1 | 1 | 1 | 1 | 0 | 1 | 0 | 0 | 3 | 1 |
| 24 | 101124 | 3 | 0 | 1 | 4 | 2 | 1 | 1 | 1 | 0 | 0 | 1 | 1 | 3 | 1 | 1 | 1 | 1 | 1 | 1 | 1 | 0 | 0 | 0 | 3 | 1 |
| 25 | 101125 | 3 | 1 | 1 | 5 | 2 | 1 | 1 | 1 | 0 | 0 | 2 | 1 | 3 | 1 | 1 | 1 | 1 | 0 | 0 | 0 | 0 | 1 | 0 | 2 | 0 |
| 26 | 101126 | 3 | 0 | 3 | 3 | 2 | 1 | 1 | 1 | 0 | 0 | 1 | 2 | 3 | 1 | 1 | 1 | 1 | 1 | 1 | 0 | 0 | 1 | 0 | 3 | 1 |
| 27 | 101127 | 3 | 1 | 3 | 3 | 2 | 1 | 1 | 1 | 0 | 0 | 2 | 2 | 3 | 1 | 1 | 1 | 1 | 1 | 1 | 1 | 0 | 0 | 0 | 3 | 1 |
| 28 | 101128 | 3 | 1 | 3 | 4 | 2 | 1 | 1 | 1 | 0 | 0 | 2 | 2 | 3 | 1 | 1 | 1 | 1 | 1 | 1 | 0 | 1 | 0 | 0 | 3 | 1 |
| 29 | 101129 | 3 | 0 | 4 | 2 | 1 | 1 | 1 | 1 | 0 | 0 | 1 | 2 | 3 | 1 | 1 | 0 | 1 | 1 | 1 | 0 | 0 | 1 | 0 | 3 | 1 |
| 30 | 101130 | 3 | 1 | 3 | 4 | 2 | 1 | 1 | 1 | 0 | 0 | 2 | 2 | 3 | 1 | 1 | 1 | 1 | 1 | 1 | 0 | 0 | 0 | 1 | 3 | 1 |
| 31 | 10121 | 3 | 0 | 3 | 1 | 2 | 1 | 3 | 3 | 0 | 0 | 1 | 2 | 3 | 1 | 1 | 1 | 1 | 1 | 1 | 0 | 1 | 0 | 1 | 4 | 1 |
| 32 | 10122 | 3 | 1 | 4 | 2 | 2 | 2 | 3 | 3 | 0 | 0 | 2 | 2 | 3 | 1 | 1 | 1 | 1 | 1 | 1 | 0 | 1 | 1 | 1 | 4 | 1 |
| 33 | 10123 | 3 | 0 | 2 | 2 | 2 | 2 | 3 | 3 | 0 | 0 | 1 | 1 | 3 | 1 | 1 | 1 | 1 | 1 | 1 | 0 | 1 | 1 | 1 | 4 | 1 |
| 34 | 10124 | 3 | 0 | 4 | 1 | 2 | 1 | 3 | 3 | 0 | 0 | 1 | 2 | 3 | 1 | 1 | 1 | 1 | 1 | 1 | 0 | 1 | 0 | 1 | 4 | 1 |
| 35 | 10125 | 3 | 0 | 4 | 1 | 2 | 1 | 3 | 3 | 0 | 0 | 1 | 2 | 3 | 1 | 1 | 0 | 1 | 0 | 1 | 0 | 1 | 0 | 0 | 2 | 0 |
| 36 | 10126 | 3 | 0 | 1 | 5 | 1 | 1 | 3 | 3 | 0 | 0 | 2 | 1 | 3 | 1 | 0 | 0 | 0 | 0 | 0 | 0 | 0 | 0 | 0 | 2 | 0 |
| 37 | 10127 | 3 | 1 | 1 | 5 | 1 | 1 | 3 | 3 | 0 | 0 | 2 | 1 | 3 | 0 | 1 | 0 | 1 | 1 | 1 | 0 | 1 | 1 | 1 | 3 | 1 |
| 38 | 10128 | 3 | 0 | 2 | 3 | 2 | 1 | 2 | 3 | 0 | 0 | 1 | 1 | 3 | 1 | 1 | 1 | 1 | 1 | 1 | 0 | 1 | 0 | 1 | 4 | 1 |
| 39 | 10129 | 3 | 1 | 1 | 3 | 2 | 1 | 2 | 3 | 0 | 0 | 2 | 1 | 3 | 0 | 1 | 1 | 1 | 1 | 1 | 0 | 1 | 0 | 1 | 3 | 1 |
| 40 | 101210 | 3 | 0 | 1 | 2 | 2 | 2 | 2 | 3 | 0 | 0 | 1 | 1 | 3 | 1 | 1 | 1 | 1 | 1 | 1 | 0 | 1 | 0 | 0 | 3 | 1 |
| 41 | 101211 | 3 | 0 | 1 | 4 | 2 | 1 | 2 | 3 | 0 | 0 | 1 | 1 | 3 | 1 | 1 | 1 | 1 | 1 | 1 | 1 | 1 | 0 | 1 | 4 | 1 |
| 42 | 101212 | 3 | 0 | 4 | 2 | 3 | 1 | 2 | 3 | 0 | 0 | 1 | 2 | 3 | 0 | 1 | 1 | 1 | 1 | 1 | 1 | 0 | 1 | 1 | 4 | 1 |
| 43 | 101213 | 3 | 1 | 2 | 5 | 2 | 1 | 2 | 3 | 0 | 0 | 2 | 1 | 3 | 0 | 1 | 1 | 1 | 1 | 1 | 1 | 0 | 1 | 0 | 3 | 1 |
| 44 | 101214 | 3 | 1 | 1 | 4 | 2 | 1 | 2 | 3 | 0 | 0 | 2 | 1 | 3 | 1 | 1 | 1 | 1 | 0 | 1 | 1 | 0 | 1 | 1 | 4 | 1 |
| 45 | 101215 | 3 | 0 | 1 | 1 | 2 | 1 | 2 | 3 | 0 | 0 | 1 | 1 | 3 | 0 | 0 | 0 | 0 | 0 | 0 | 0 | 0 | 0 | 0 | 1 | 0 |
| 46 | 101216 | 3 | 0 | 1 | 5 | 1 | 1 | 2 | 3 | 0 | 0 | 1 | 1 | 3 | 1 | 1 | 1 | 1 | 0 | 1 | 1 | 1 | 0 | 0 | 3 | 1 |
| 47 | 101217 | 3 | 1 | 2 | 2 | 2 | 1 | 2 | 3 | 0 | 0 | 5 | 1 | 3 | 1 | 1 | 1 | 1 | 1 | 1 | 1 | 0 | 1 | 0 | 4 | 1 |
| 48 | 101218 | 3 | 0 | 2 | 1 | 2 | 1 | 2 | 3 | 0 | 0 | 1 | 1 | 3 | 1 | 1 | 0 | 1 | 1 | 1 | 0 | 1 | 0 | 1 | 3 | 1 |
| 49 | 101219 | 3 | 1 | 3 | 2 | 2 | 1 | 3 | 3 | 0 | 0 | 4 | 2 | 3 | 1 | 1 | 1 | 1 | 1 | 1 | 0 | 0 | 1 | 1 | 4 | 1 |
| 50 | 101220 | 3 | 0 | 2 | 2 | 2 | 1 | 3 | 3 | 0 | 0 | 1 | 1 | 3 | 1 | 1 | 1 | 1 | 1 | 1 | 1 | 0 | 1 | 1 | 4 | 1 |
| 51 | 101221 | 3 | 1 | 3 | 2 | 2 | 1 | 3 | 3 | 0 | 0 | 4 | 2 | 3 | 1 | 1 | 1 | 1 | 1 | 1 | 0 | 0 | 1 | 1 | 4 | 1 |
| 52 | 101222 | 3 | 1 | 4 | 2 | 3 | 1 | 3 | 3 | 1 | 0 | 4 | 2 | 3 | 1 | 1 | 1 | 1 | 1 | 1 | 1 | 1 | 1 | 1 | 4 | 1 |
| 53 | 101223 | 3 | 1 | 1 | 4 | 2 | 1 | 3 | 3 | 0 | 0 | 4 | 1 | 3 | 1 | 1 | 1 | 1 | 1 | 1 | 0 | 1 | 1 | 1 | 4 | 1 |
| 54 | 101224 | 3 | 0 | 2 | 3 | 2 | 1 | 2 | 3 | 0 | 0 | 1 | 1 | 3 | 1 | 1 | 1 | 1 | 1 | 1 | 1 | 0 | 1 | 1 | 4 | 1 |
| 55 | 101225 | 3 | 1 | 3 | 1 | 2 | 1 | 3 | 3 | 0 | 0 | 4 | 2 | 3 | 1 | 1 | 1 | 1 | 1 | 1 | 0 | 1 | 1 | 1 | 4 | 1 |
| 56 | 101226 | 3 | 0 | 2 | 3 | 1 | 1 | 1 | 4 | 0 | 0 | 1 | 1 | 2 | 0 | 1 | 1 | 1 | 1 | 1 | 1 | 0 | 1 | 0 | 3 | 1 |
| 57 | 101227 | 3 | 1 | 3 | 2 | 2 | 1 | 1 | 3 | 0 | 0 | 4 | 2 | 2 | 1 | 1 | 1 | 1 | 1 | 1 | 1 | 0 | 1 | 1 | 4 | 1 |
| 58 | 101228 | 3 | 0 | 3 | 3 | 2 | 1 | 1 | 3 | 0 | 0 | 1 | 2 | 3 | 1 | 1 | 1 | 1 | 1 | 1 | 1 | 0 | 0 | 0 | 3 | 1 |
| 59 | 101229 | 3 | 1 | 4 | 2 | 2 | 1 | 1 | 3 | 1 | 0 | 1 | 2 | 3 | 1 | 1 | 1 | 1 | 1 | 1 | 1 | 1 | 1 | 1 | 4 | 1 |
| 60 | 101230 | 3 | 1 | 3 | 3 | 3 | 1 | 1 | 3 | 0 | 0 | 2 | 2 | 3 | 1 | 1 | 1 | 1 | 1 | 1 | 0 | 0 | 1 | 0 | 3 | 1 |
| 61 | 101231 | 3 | 1 | 1 | 5 | 1 | 1 | 1 | 3 | 0 | 0 | 2 | 1 | 3 | 1 | 0 | 0 | 0 | 0 | 0 | 0 | 0 | 0 | 0 | 2 | 0 |
| 62 | 101232 | 3 | 1 | 1 | 5 | 1 | 1 | 2 | 3 | 0 | 0 | 2 | 1 | 3 | 1 | 1 | 1 | 1 | 1 | 1 | 0 | 1 | 1 | 0 | 4 | 1 |
| 63 | 101233 | 3 | 1 | 1 | 2 | 1 | 1 | 1 | 3 | 0 | 0 | 2 | 1 | 3 | 1 | 1 | 1 | 1 | 1 | 1 | 0 | 0 | 1 | 0 | 3 | 1 |
| 64 | 101234 | 3 | 1 | 2 | 3 | 1 | 2 | 3 | 3 | 0 | 0 | 2 | 1 | 3 | 1 | 1 | 1 | 1 | 0 | 1 | 0 | 1 | 0 | 0 | 3 | 1 |
| 65 | 101235 | 3 | 1 | 1 | 5 | 1 | 1 | 3 | 3 | 0 | 0 | 2 | 1 | 3 | 1 | 1 | 1 | 1 | 1 | 1 | 0 | 1 | 0 | 0 | 3 | 1 |
| 66 | 10131 | 3 | 0 | 3 | 3 | 2 | 1 | 1 | 4 | 0 | 0 | 1 | 2 | 3 | 1 | 1 | 1 | 1 | 1 | 1 | 0 | 0 | 1 | 0 | 3 | 1 |
| 67 | 10132 | 3 | 1 | 4 | 1 | 2 | 1 | 1 | 4 | 0 | 0 | 5 | 2 | 3 | 1 | 1 | 1 | 1 | 1 | 1 | 0 | 1 | 0 | 0 | 3 | 1 |
| 68 | 10133 | 3 | 1 | 4 | 3 | 2 | 1 | 1 | 4 | 0 | 0 | 5 | 2 | 3 | 1 | 1 | 1 | 1 | 1 | 1 | 0 | 1 | 0 | 0 | 3 | 1 |
| 69 | 10134 | 3 | 0 | 4 | 2 | 2 | 1 | 1 | 4 | 1 | 0 | 1 | 2 | 3 | 1 | 1 | 1 | 1 | 1 | 1 | 0 | 0 | 1 | 0 | 3 | 1 |
| 70 | 10135 | 3 | 0 | 4 | 1 | 2 | 1 | 1 | 4 | 0 | 0 | 1 | 2 | 3 | 1 | 1 | 0 | 1 | 0 | 1 | 0 | 1 | 0 | 0 | 2 | 0 |
| 71 | 10136 | 3 | 0 | 4 | 3 | 2 | 1 | 1 | 4 | 0 | 0 | 1 | 2 | 3 | 1 | 1 | 1 | 1 | 1 | 1 | 0 | 1 | 0 | 0 | 3 | 1 |
| 72 | 10137 | 3 | 1 | 4 | 2 | 3 | 2 | 2 | 4 | 0 | 0 | 5 | 2 | 3 | 1 | 1 | 1 | 1 | 1 | 1 | 1 | 0 | 0 | 0 | 3 | 1 |
| 73 | 10138 | 3 | 0 | 4 | 2 | 2 | 1 | 2 | 4 | 1 | 0 | 1 | 2 | 3 | 1 | 1 | 1 | 1 | 1 | 1 | 0 | 0 | 1 | 0 | 3 | 1 |
| 74 | 10139 | 3 | 0 | 3 | 2 | 2 | 1 | 2 | 4 | 1 | 0 | 1 | 2 | 3 | 1 | 1 | 1 | 1 | 1 | 1 | 0 | 0 | 1 | 0 | 3 | 1 |
| 75 | 101310 | 3 | 1 | 4 | 2 | 2 | 1 | 2 | 4 | 0 | 0 | 5 | 2 | 3 | 1 | 1 | 1 | 1 | 1 | 1 | 0 | 0 | 1 | 0 | 3 | 1 |
| 76 | 101311 | 3 | 1 | 4 | 1 | 2 | 1 | 2 | 4 | 0 | 0 | 5 | 2 | 3 | 1 | 1 | 1 | 1 | 1 | 1 | 0 | 0 | 1 | 0 | 3 | 1 |
| 77 | 101312 | 3 | 0 | 4 | 2 | 2 | 1 | 2 | 4 | 0 | 0 | 1 | 2 | 3 | 1 | 1 | 1 | 1 | 1 | 1 | 1 | 0 | 0 | 0 | 3 | 1 |
| 78 | 101313 | 3 | 0 | 1 | 3 | 1 | 1 | 2 | 4 | 0 | 0 | 1 | 1 | 3 | 1 | 1 | 1 | 1 | 0 | 1 | 1 | 0 | 0 | 0 | 3 | 1 |
| 79 | 101314 | 3 | 1 | 4 | 2 | 2 | 1 | 2 | 4 | 0 | 0 | 2 | 2 | 3 | 1 | 1 | 1 | 1 | 1 | 1 | 0 | 0 | 1 | 0 | 3 | 1 |
| 80 | 101315 | 3 | 1 | 3 | 1 | 1 | 1 | 2 | 4 | 0 | 0 | 2 | 2 | 3 | 1 | 1 | 1 | 1 | 0 | 1 | 0 | 0 | 1 | 0 | 3 | 1 |
| 81 | 101316 | 3 | 0 | 3 | 3 | 2 | 1 | 2 | 4 | 0 | 0 | 1 | 2 | 3 | 1 | 1 | 1 | 1 | 1 | 1 | 1 | 0 | 1 | 0 | 4 | 1 |
| 82 | 101317 | 3 | 0 | 3 | 2 | 2 | 1 | 2 | 4 | 0 | 0 | 1 | 2 | 3 | 1 | 1 | 1 | 1 | 1 | 1 | 1 | 0 | 1 | 0 | 4 | 1 |
| 83 | 101318 | 3 | 0 | 4 | 4 | 2 | 2 | 2 | 4 | 0 | 0 | 1 | 2 | 3 | 1 | 1 | 1 | 1 | 1 | 1 | 0 | 0 | 1 | 0 | 3 | 1 |
| 84 | 101319 | 3 | 1 | 3 | 3 | 3 | 1 | 2 | 4 | 0 | 0 | 3 | 2 | 3 | 1 | 1 | 1 | 1 | 1 | 1 | 0 | 0 | 1 | 0 | 3 | 1 |
| 85 | 101320 | 3 | 1 | 2 | 5 | 2 | 1 | 2 | 4 | 0 | 0 | 2 | 1 | 3 | 1 | 1 | 1 | 1 | 1 | 1 | 0 | 0 | 1 | 0 | 3 | 1 |
| 86 | 101321 | 3 | 0 | 2 | 4 | 2 | 1 | 2 | 4 | 0 | 0 | 1 | 1 | 3 | 1 | 1 | 1 | 1 | 1 | 1 | 0 | 0 | 1 | 0 | 3 | 1 |
| 87 | 101322 | 3 | 1 | 3 | 2 | 2 | 1 | 2 | 4 | 0 | 0 | 2 | 2 | 3 | 1 | 1 | 1 | 1 | 1 | 1 | 0 | 0 | 1 | 0 | 3 | 1 |
| 88 | 101323 | 3 | 0 | 1 | 2 | 2 | 2 | 2 | 4 | 0 | 0 | 2 | 1 | 3 | 1 | 1 | 1 | 1 | 1 | 1 | 0 | 0 | 1 | 0 | 3 | 1 |
| 89 | 101324 | 3 | 1 | 2 | 4 | 2 | 1 | 2 | 4 | 0 | 0 | 2 | 1 | 3 | 1 | 1 | 1 | 1 | 1 | 1 | 0 | 0 | 1 | 0 | 3 | 1 |
| 90 | 101325 | 3 | 1 | 3 | 3 | 2 | 1 | 2 | 4 | 0 | 0 | 1 | 2 | 3 | 1 | 1 | 1 | 1 | 1 | 1 | 1 | 0 | 0 | 0 | 3 | 1 |
| 91 | 101326 | 3 | 0 | 1 | 1 | 2 | 1 | 2 | 4 | 0 | 0 | 1 | 1 | 3 | 1 | 1 | 1 | 1 | 1 | 1 | 0 | 0 | 1 | 0 | 3 | 1 |
| 92 | 101327 | 3 | 1 | 3 | 1 | 2 | 1 | 2 | 4 | 0 | 0 | 2 | 2 | 3 | 1 | 1 | 1 | 1 | 1 | 1 | 0 | 0 | 1 | 0 | 3 | 1 |
| 93 | 101328 | 3 | 1 | 1 | 5 | 2 | 1 | 2 | 4 | 0 | 0 | 2 | 1 | 3 | 1 | 1 | 1 | 1 | 1 | 1 | 0 | 0 | 1 | 0 | 3 | 1 |

| | | | | | | | | | | | | | | | | | | | | | | | | | | |
|---|---|---|---|---|---|---|---|---|---|---|---|---|---|---|---|---|---|---|---|---|---|---|---|---|---|---|
| 94 | 101329 | 3 | 0 | 4 | 1 | 2 | 1 | 2 | 4 | 0 | 0 | 1 | 2 | 3 | 1 | 1 | 1 | 1 | 1 | 1 | 0 | 0 | 1 | 0 | 3 | 1 |
| 95 | 101330 | 3 | 1 | 3 | 5 | 2 | 1 | 2 | 4 | 0 | 0 | 2 | 2 | 3 | 1 | 1 | 1 | 1 | 1 | 1 | 1 | 0 | 0 | 0 | 3 | 1 |
| 96 | 10141 | 3 | 1 | 5 | 2 | 3 | 1 | 1 | 4 | 0 | 0 | 5 | 3 | 3 | 1 | 1 | 1 | 1 | 0 | 0 | 0 | 1 | 1 | 0 | 3 | 1 |
| 97 | 10142 | 3 | 0 | 4 | 2 | 2 | 1 | 1 | 4 | 0 | 0 | 1 | 2 | 3 | 0 | 1 | 1 | 1 | 1 | 0 | 0 | 1 | 0 | 0 | 2 | 0 |
| 98 | 10143 | 3 | 1 | 2 | 5 | 2 | 1 | 1 | 4 | 1 | 1 | 5 | 1 | 3 | 0 | 1 | 1 | 1 | 0 | 1 | 0 | 1 | 1 | 0 | 3 | 1 |
| 99 | 10144 | 3 | 0 | 4 | 1 | 2 | 1 | 1 | 4 | 0 | 0 | 5 | 2 | 3 | 0 | 1 | 1 | 1 | 0 | 0 | 0 | 1 | 0 | 0 | 2 | 0 |
| 100 | 10145 | 3 | 1 | 1 | 4 | 1 | 2 | 1 | 4 | 0 | 0 | 2 | 1 | 3 | 0 | 0 | 0 | 0 | 0 | 0 | 0 | 0 | 0 | 0 | 1 | 0 |
| 101 | 10146 | 3 | 0 | 2 | 5 | 1 | 2 | 1 | 4 | 0 | 0 | 1 | 1 | 2 | 0 | 0 | 0 | 0 | 0 | 0 | 0 | 0 | 0 | 0 | 1 | 0 |
| 102 | 10147 | 3 | 1 | 4 | 2 | 2 | 2 | 1 | 4 | 0 | 0 | 5 | 2 | 3 | 0 | 1 | 0 | 1 | 0 | 0 | 0 | 1 | 0 | 0 | 2 | 0 |
| 103 | 10148 | 3 | 1 | 1 | 4 | 2 | 2 | 1 | 4 | 0 | 0 | 2 | 1 | 3 | 0 | 1 | 0 | 0 | 0 | 0 | 0 | 0 | 0 | 0 | 2 | 0 |
| 104 | 10149 | 3 | 0 | 2 | 3 | 3 | 2 | 2 | 4 | 0 | 0 | 1 | 1 | 3 | 1 | 1 | 0 | 1 | 0 | 0 | 1 | 0 | 0 | 0 | 2 | 0 |
| 105 | 101410 | 3 | 0 | 2 | 3 | 2 | 2 | 2 | 4 | 0 | 0 | 1 | 1 | 2 | 0 | 1 | 0 | 0 | 0 | 0 | 0 | 0 | 0 | 0 | 2 | 0 |
| 106 | 101411 | 3 | 1 | 1 | 5 | 1 | 1 | 2 | 4 | 0 | 0 | 5 | 1 | 3 | 0 | 0 | 0 | 0 | 0 | 0 | 0 | 0 | 0 | 0 | 1 | 0 |
| 107 | 101412 | 3 | 1 | 2 | 3 | 2 | 1 | 2 | 4 | 0 | 0 | 2 | 1 | 3 | 0 | 1 | 0 | 0 | 0 | 0 | 0 | 0 | 0 | 0 | 2 | 0 |
| 108 | 101413 | 3 | 1 | 4 | 2 | 2 | 1 | 2 | 4 | 0 | 0 | 2 | 2 | 3 | 1 | 1 | 0 | 1 | 0 | 0 | 0 | 1 | 0 | 1 | 2 | 0 |
| 109 | 101414 | 3 | 1 | 3 | 3 | 1 | 1 | 2 | 4 | 0 | 0 | 2 | 2 | 3 | 0 | 0 | 0 | 0 | 0 | 0 | 0 | 0 | 0 | 0 | 1 | 0 |
| 110 | 101415 | 3 | 1 | 1 | 4 | 1 | 1 | 2 | 4 | 0 | 0 | 2 | 1 | 3 | 0 | 0 | 0 | 0 | 0 | 0 | 0 | 0 | 0 | 0 | 1 | 0 |
| 111 | 101416 | 3 | 0 | 1 | 4 | 1 | 1 | 2 | 4 | 0 | 0 | 1 | 1 | 3 | 0 | 0 | 0 | 0 | 0 | 0 | 0 | 0 | 0 | 0 | 1 | 0 |
| 112 | 101417 | 3 | 1 | 2 | 3 | 2 | 1 | 2 | 4 | 0 | 0 | 2 | 1 | 3 | 0 | 1 | 0 | 1 | 0 | 0 | 0 | 1 | 0 | 1 | 2 | 0 |
| 113 | 101418 | 3 | 1 | 2 | 4 | 1 | 1 | 2 | 4 | 0 | 0 | 2 | 1 | 3 | 0 | 1 | 0 | 1 | 0 | 0 | 0 | 1 | 0 | 0 | 2 | 0 |
| 114 | 101419 | 3 | 1 | 2 | 3 | 2 | 1 | 2 | 4 | 0 | 0 | 2 | 1 | 3 | 0 | 1 | 1 | 1 | 0 | 0 | 1 | 0 | 0 | 0 | 2 | 0 |
| 115 | 101420 | 3 | 0 | 3 | 2 | 2 | 2 | 2 | 4 | 0 | 0 | 1 | 2 | 3 | 0 | 1 | 0 | 1 | 0 | 0 | 0 | 1 | 0 | 1 | 2 | 0 |
| 116 | 101421 | 3 | 1 | 2 | 4 | 2 | 1 | 2 | 4 | 0 | 0 | 2 | 1 | 3 | 0 | 0 | 0 | 0 | 0 | 0 | 0 | 0 | 0 | 0 | 1 | 0 |
| 117 | 101422 | 3 | 0 | 1 | 3 | 1 | 1 | 2 | 4 | 0 | 0 | 1 | 1 | 3 | 0 | 0 | 0 | 0 | 0 | 0 | 0 | 0 | 0 | 0 | 1 | 0 |
| 118 | 101423 | 3 | 1 | 2 | 4 | 2 | 1 | 2 | 4 | 0 | 0 | 2 | 1 | 3 | 0 | 1 | 0 | 1 | 0 | 0 | 0 | 0 | 0 | 1 | 2 | 0 |
| 119 | 101424 | 3 | 0 | 4 | 1 | 2 | 1 | 1 | 4 | 0 | 0 | 1 | 2 | 2 | 0 | 1 | 1 | 1 | 1 | 1 | 0 | 1 | 0 | 1 | 3 | 1 |
| 120 | 101425 | 3 | 1 | 4 | 1 | 2 | 1 | 1 | 4 | 0 | 0 | 4 | 2 | 3 | 0 | 1 | 1 | 1 | 0 | 0 | 0 | 1 | 0 | 1 | 2 | 0 |
| 121 | 101426 | 3 | 0 | 3 | 1 | 1 | 1 | 2 | 4 | 0 | 0 | 1 | 2 | 3 | 1 | 1 | 1 | 1 | 1 | 1 | 0 | 0 | 1 | 0 | 3 | 1 |
| 122 | 101427 | 3 | 0 | 3 | 3 | 2 | 1 | 2 | 4 | 0 | 0 | 1 | 2 | 3 | 0 | 1 | 1 | 1 | 0 | 1 | 1 | 1 | 0 | 0 | 3 | 1 |
| 123 | 101428 | 3 | 0 | 1 | 4 | 2 | 2 | 2 | 4 | 0 | 0 | 1 | 1 | 3 | 0 | 1 | 0 | 0 | 0 | 0 | 0 | 0 | 0 | 0 | 2 | 0 |
| 124 | 101429 | 3 | 1 | 3 | 2 | 2 | 1 | 2 | 4 | 0 | 0 | 2 | 2 | 3 | 0 | 1 | 0 | 1 | 0 | 0 | 0 | 1 | 0 | 1 | 2 | 0 |
| 125 | 101430 | 3 | 0 | 2 | 3 | 2 | 1 | 2 | 4 | 0 | 0 | 1 | 1 | 3 | 0 | 1 | 0 | 1 | 0 | 0 | 0 | 0 | 0 | 1 | 2 | 0 |
| 126 | 101431 | 3 | 1 | 2 | 4 | 2 | 2 | 2 | 4 | 0 | 0 | 2 | 1 | 3 | 0 | 1 | 1 | 1 | 0 | 1 | 1 | 1 | 0 | 1 | 3 | 1 |
| 127 | 101432 | 3 | 1 | 1 | 4 | 2 | 1 | 2 | 4 | 0 | 0 | 1 | 1 | 3 | 0 | 1 | 1 | 1 | 0 | 0 | 0 | 0 | 1 | 0 | 2 | 0 |
| 128 | 101433 | 3 | 0 | 3 | 1 | 2 | 1 | 2 | 4 | 0 | 0 | 1 | 2 | 3 | 0 | 1 | 0 | 1 | 1 | 1 | 0 | 1 | 0 | 1 | 3 | 1 |
| 129 | 101434 | 3 | 1 | 3 | 2 | 1 | 1 | 2 | 4 | 0 | 0 | 2 | 2 | 4 | 1 | 1 | 1 | 1 | 0 | 0 | 1 | 0 | 0 | 1 | 3 | 1 |
| 130 | 101435 | 3 | 1 | 3 | 2 | 2 | 1 | 2 | 4 | 0 | 0 | 4 | 2 | 3 | 0 | 1 | 1 | 1 | 1 | 1 | 0 | 1 | 0 | 0 | 3 | 1 |
| 131 | 10151 | 3 | 0 | 4 | 2 | 2 | 2 | 2 | 4 | 0 | 0 | 1 | 2 | 3 | 0 | 1 | 1 | 1 | 1 | 0 | 0 | 0 | 0 | 1 | 2 | 0 |
| 132 | 10152 | 3 | 1 | 4 | 4 | 2 | 1 | 2 | 4 | 0 | 0 | 4 | 2 | 3 | 0 | 1 | 1 | 1 | 1 | 1 | 1 | 0 | 0 | 0 | 3 | 1 |
| 133 | 10153 | 3 | 0 | 3 | 2 | 2 | 1 | 2 | 4 | 0 | 0 | 1 | 2 | 3 | 0 | 1 | 1 | 1 | 1 | 1 | 0 | 1 | 1 | 0 | 3 | 1 |
| 134 | 10154 | 3 | 0 | 4 | 1 | 2 | 2 | 2 | 4 | 1 | 0 | 5 | 2 | 3 | 0 | 1 | 1 | 1 | 1 | 1 | 0 | 0 | 0 | 1 | 3 | 1 |
| 135 | 10155 | 3 | 1 | 3 | 3 | 2 | 1 | 2 | 4 | 0 | 0 | 4 | 2 | 3 | 1 | 1 | 1 | 1 | 1 | 1 | 0 | 0 | 0 | 1 | 3 | 1 |
| 136 | 10156 | 3 | 1 | 3 | 3 | 2 | 1 | 2 | 4 | 0 | 0 | 5 | 2 | 3 | 0 | 1 | 1 | 1 | 0 | 1 | 0 | 0 | 0 | 1 | 2 | 0 |
| 137 | 10157 | 3 | 0 | 4 | 1 | 2 | 1 | 1 | 1 | 0 | 0 | 1 | 2 | 3 | 0 | 1 | 1 | 1 | 1 | 1 | 0 | 1 | 0 | 1 | 3 | 1 |
| 138 | 10158 | 3 | 0 | 4 | 2 | 2 | 1 | 1 | 1 | 0 | 0 | 1 | 2 | 3 | 0 | 1 | 1 | 1 | 1 | 1 | 0 | 1 | 0 | 1 | 3 | 1 |
| 139 | 10159 | 3 | 1 | 4 | 5 | 3 | 1 | 3 | 4 | 1 | 1 | 5 | 2 | 3 | 0 | 1 | 1 | 1 | 1 | 1 | 0 | 1 | 0 | 1 | 3 | 1 |
| 140 | 101510 | 3 | 1 | 4 | 1 | 2 | 1 | 1 | 1 | 0 | 0 | 4 | 2 | 3 | 0 | 1 | 1 | 1 | 1 | 1 | 0 | 1 | 0 | 1 | 3 | 1 |
| 141 | 101511 | 3 | 0 | 4 | 4 | 3 | 2 | 1 | 1 | 1 | 1 | 5 | 2 | 3 | 0 | 1 | 1 | 1 | 1 | 1 | 0 | 0 | 1 | 1 | 3 | 1 |
| 142 | 101512 | 3 | 0 | 5 | 2 | 1 | 1 | 1 | 1 | 0 | 0 | 1 | 3 | 3 | 0 | 1 | 1 | 1 | 1 | 1 | 0 | 0 | 0 | 1 | 3 | 1 |
| 143 | 101513 | 3 | 1 | 4 | 3 | 2 | 2 | 2 | 1 | 1 | 0 | 4 | 2 | 3 | 0 | 1 | 1 | 1 | 1 | 1 | 0 | 0 | 1 | 0 | 3 | 1 |
| 144 | 101514 | 3 | 1 | 3 | 5 | 2 | 1 | 2 | 1 | 0 | 0 | 4 | 2 | 3 | 1 | 1 | 1 | 1 | 0 | 1 | 0 | 1 | 0 | 1 | 3 | 1 |
| 145 | 101515 | 3 | 0 | 4 | 1 | 2 | 1 | 1 | 1 | 0 | 0 | 1 | 2 | 3 | 0 | 1 | 1 | 1 | 0 | 1 | 0 | 1 | 0 | 0 | 2 | 0 |
| 146 | 101516 | 3 | 1 | 2 | 2 | 2 | 1 | 1 | 1 | 0 | 0 | 4 | 1 | 3 | 0 | 1 | 1 | 1 | 1 | 0 | 0 | 0 | 1 | 0 | 2 | 0 |
| 147 | 101517 | 3 | 0 | 3 | 2 | 2 | 2 | 1 | 1 | 0 | 0 | 1 | 2 | 2 | 0 | 1 | 1 | 1 | 1 | 1 | 1 | 0 | 0 | 0 | 3 | 1 |
| 148 | 101518 | 3 | 1 | 4 | 3 | 2 | 1 | 1 | 1 | 0 | 0 | 4 | 2 | 3 | 0 | 1 | 1 | 1 | 1 | 1 | 0 | 1 | 0 | 0 | 3 | 1 |
| 149 | 101519 | 3 | 0 | 4 | 2 | 2 | 2 | 1 | 1 | 0 | 0 | 1 | 2 | 3 | 0 | 1 | 1 | 1 | 0 | 1 | 0 | 0 | 0 | 1 | 2 | 0 |
| 150 | 101520 | 3 | 1 | 4 | 4 | 3 | 2 | 2 | 1 | 1 | 1 | 4 | 2 | 3 | 0 | 1 | 1 | 1 | 1 | 1 | 0 | 0 | 1 | 0 | 3 | 1 |
| 151 | 101521 | 3 | 0 | 2 | 5 | 2 | 1 | 2 | 1 | 0 | 0 | 1 | 1 | 3 | 0 | 1 | 1 | 1 | 1 | 0 | 0 | 0 | 0 | 1 | 2 | 0 |
| 152 | 101522 | 3 | 1 | 3 | 5 | 2 | 1 | 2 | 1 | 1 | 1 | 5 | 2 | 3 | 0 | 1 | 1 | 1 | 1 | 1 | 0 | 1 | 0 | 0 | 3 | 1 |
| 153 | 101523 | 3 | 1 | 4 | 4 | 2 | 1 | 1 | 1 | 1 | 1 | 5 | 2 | 3 | 0 | 1 | 1 | 1 | 1 | 1 | 0 | 0 | 0 | 0 | 2 | 0 |
| 154 | 101524 | 3 | 0 | 4 | 3 | 3 | 1 | 1 | 1 | 1 | 1 | 5 | 2 | 3 | 0 | 1 | 0 | 1 | 1 | 1 | 0 | 0 | 0 | 1 | 2 | 0 |
| 155 | 101525 | 3 | 1 | 4 | 2 | 2 | 2 | 1 | 1 | 1 | 1 | 5 | 2 | 3 | 0 | 1 | 1 | 1 | 1 | 1 | 0 | 0 | 0 | 1 | 3 | 1 |
| 156 | 101526 | 3 | 1 | 4 | 2 | 2 | 1 | 1 | 1 | 1 | 1 | 5 | 2 | 3 | 0 | 1 | 0 | 1 | 1 | 0 | 0 | 0 | 1 | 1 | 2 | 0 |
| 157 | 101527 | 3 | 1 | 4 | 3 | 2 | 1 | 1 | 1 | 0 | 0 | 5 | 2 | 3 | 0 | 1 | 0 | 1 | 1 | 1 | 0 | 1 | 0 | 1 | 3 | 1 |
| 158 | 101528 | 3 | 1 | 2 | 4 | 2 | 1 | 1 | 1 | 1 | 1 | 2 | 1 | 3 | 0 | 1 | 1 | 1 | 1 | 0 | 0 | 0 | 1 | 0 | 2 | 0 |
| 159 | 101529 | 3 | 0 | 3 | 3 | 2 | 1 | 1 | 1 | 0 | 0 | 1 | 2 | 3 | 0 | 1 | 1 | 1 | 1 | 1 | 1 | 0 | 0 | 0 | 3 | 1 |
| 160 | 101530 | 3 | 0 | 4 | 3 | 2 | 1 | 1 | 1 | 0 | 0 | 1 | 2 | 3 | 0 | 1 | 1 | 1 | 1 | 1 | 0 | 1 | 0 | 1 | 3 | 1 |
| 161 | 10161 | 3 | 0 | 4 | 1 | 2 | 1 | 1 | 1 | 0 | 0 | 1 | 2 | 3 | 0 | 1 | 1 | 1 | 0 | 0 | 0 | 0 | 0 | 1 | 2 | 0 |
| 162 | 10162 | 3 | 0 | 2 | 1 | 2 | 1 | 2 | 2 | 1 | 0 | 1 | 1 | 3 | 0 | 1 | 0 | 1 | 1 | 0 | 0 | 0 | 0 | 1 | 2 | 0 |
| 163 | 10163 | 3 | 0 | 4 | 2 | 3 | 1 | 2 | 1 | 0 | 0 | 1 | 2 | 3 | 0 | 1 | 1 | 1 | 1 | 1 | 0 | 0 | 0 | 1 | 3 | 1 |
| 164 | 10164 | 3 | 1 | 5 | 4 | 3 | 2 | 2 | 1 | 1 | 1 | 5 | 3 | 3 | 1 | 1 | 1 | 1 | 1 | 0 | 0 | 0 | 0 | 0 | 2 | 0 |
| 165 | 10165 | 3 | 1 | 2 | 2 | 2 | 1 | 2 | 1 | 0 | 0 | 2 | 1 | 3 | 0 | 1 | 0 | 1 | 1 | 1 | 0 | 0 | 0 | 1 | 2 | 0 |
| 166 | 10166 | 3 | 1 | 2 | 5 | 2 | 1 | 2 | 1 | 1 | 1 | 2 | 1 | 3 | 0 | 1 | 1 | 1 | 1 | 0 | 0 | 0 | 0 | 0 | 2 | 0 |
| 167 | 10167 | 3 | 1 | 2 | 4 | 2 | 2 | 1 | 1 | 0 | 0 | 4 | 1 | 3 | 0 | 1 | 0 | 1 | 1 | 1 | 0 | 0 | 1 | 0 | 2 | 0 |
| 168 | 10168 | 3 | 1 | 2 | 1 | 2 | 1 | 1 | 1 | 0 | 0 | 2 | 1 | 3 | 0 | 1 | 0 | 1 | 0 | 0 | 0 | 0 | 1 | 0 | 2 | 0 |
| 169 | 10169 | 3 | 0 | 2 | 3 | 2 | 1 | 1 | 1 | 0 | 0 | 1 | 1 | 3 | 0 | 1 | 0 | 1 | 1 | 1 | 0 | 0 | 0 | 1 | 2 | 0 |
| 170 | 101610 | 3 | 0 | 1 | 3 | 2 | 2 | 1 | 1 | 0 | 0 | 1 | 1 | 3 | 0 | 1 | 1 | 1 | 0 | 1 | 0 | 0 | 0 | 0 | 2 | 0 |
| 171 | 101611 | 3 | 0 | 2 | 1 | 2 | 2 | 1 | 1 | 0 | 0 | 1 | 1 | 3 | 0 | 1 | 1 | 1 | 1 | 1 | 0 | 0 | 0 | 1 | 3 | 1 |
| 172 | 101612 | 3 | 0 | 2 | 4 | 2 | 1 | 1 | 1 | 0 | 0 | 1 | 1 | 3 | 0 | 1 | 0 | 1 | 0 | 1 | 0 | 0 | 0 | 0 | 2 | 0 |
| 173 | 101613 | 3 | 1 | 2 | 3 | 2 | 1 | 1 | 1 | 0 | 0 | 4 | 1 | 3 | 0 | 1 | 1 | 1 | 1 | 1 | 1 | 0 | 0 | 0 | 3 | 1 |
| 174 | 101614 | 3 | 0 | 2 | 3 | 3 | 2 | 1 | 1 | 0 | 0 | 1 | 1 | 3 | 0 | 1 | 1 | 1 | 1 | 1 | 0 | 0 | 0 | 1 | 3 | 1 |
| 175 | 101615 | 3 | 0 | 2 | 4 | 2 | 1 | 1 | 1 | 0 | 0 | 1 | 1 | 3 | 0 | 1 | 1 | 1 | 1 | 0 | 0 | 0 | 0 | 0 | 2 | 0 |
| 176 | 101616 | 3 | 0 | 4 | 1 | 2 | 1 | 1 | 1 | 0 | 0 | 1 | 2 | 3 | 0 | 1 | 1 | 1 | 0 | 0 | 0 | 0 | 0 | 1 | 2 | 0 |
| 177 | 101617 | 3 | 0 | 1 | 3 | 2 | 2 | 1 | 1 | 0 | 0 | 1 | 1 | 3 | 0 | 1 | 1 | 1 | 1 | 1 | 0 | 0 | 0 | 1 | 3 | 1 |
| 178 | 101618 | 3 | 1 | 2 | 5 | 1 | 1 | 1 | 1 | 0 | 0 | 2 | 1 | 3 | 0 | 1 | 1 | 1 | 0 | 0 | 0 | 0 | 0 | 1 | 2 | 0 |
| 179 | 101619 | 3 | 1 | 2 | 4 | 2 | 2 | 1 | 1 | 0 | 0 | 4 | 1 | 3 | 0 | 1 | 1 | 1 | 1 | 1 | 0 | 0 | 0 | 1 | 3 | 1 |
| 180 | 101620 | 3 | 0 | 3 | 1 | 2 | 2 | 1 | 1 | 0 | 0 | 1 | 2 | 3 | 0 | 1 | 0 | 1 | 0 | 1 | 0 | 0 | 0 | 1 | 2 | 0 |
| 181 | 101621 | 3 | 1 | 2 | 4 | 2 | 1 | 1 | 1 | 0 | 0 | 4 | 1 | 3 | 0 | 1 | 1 | 1 | 0 | 1 | 0 | 0 | 1 | 1 | 3 | 1 |
| 182 | 101622 | 3 | 0 | 3 | 2 | 2 | 1 | 1 | 1 | 0 | 0 | 1 | 2 | 3 | 0 | 1 | 1 | 1 | 1 | 1 | 1 | 0 | 0 | 0 | 3 | 1 |
| 183 | 101623 | 3 | 1 | 2 | 1 | 2 | 1 | 1 | 1 | 0 | 0 | 4 | 1 | 3 | 0 | 1 | 0 | 1 | 0 | 0 | 0 | 0 | 0 | 1 | 2 | 0 |
| 184 | 101624 | 3 | 0 | 2 | 5 | 1 | 1 | 1 | 1 | 0 | 0 | 1 | 1 | 3 | 0 | 1 | 1 | 1 | 0 | 0 | 0 | 0 | 1 | 0 | 2 | 0 |
| 185 | 101625 | 3 | 1 | 3 | 2 | 2 | 1 | 1 | 1 | 0 | 0 | 2 | 2 | 1 | 0 | 1 | 1 | 1 | 0 | 0 | 0 | 0 | 0 | 1 | 2 | 0 |
| 186 | 101626 | 3 | 1 | 2 | 4 | 2 | 2 | 1 | 1 | 0 | 0 | 4 | 1 | 3 | 0 | 1 | 1 | 1 | 0 | 1 | 0 | 0 | 0 | 1 | 2 | 0 |
| 187 | 101627 | 3 | 1 | 2 | 3 | 2 | 1 | 1 | 1 | 0 | 0 | 4 | 1 | 3 | 0 | 1 | 1 | 1 | 1 | 1 | 0 | 0 | 0 | 1 | 3 | 1 |

| | | | | | | | | | | | | | | | | | | | | | | | | | | |
|---|---|---|---|---|---|---|---|---|---|---|---|---|---|---|---|---|---|---|---|---|---|---|---|---|---|---|
| 188 | 101628 | 3 | 1 | 2 | 2 | 2 | 2 | 1 | 1 | 0 | 0 | 4 | 1 | 3 | 0 | 1 | 1 | 1 | 0 | 0 | 0 | 0 | 1 | 0 | 2 | 0 |
| 189 | 101629 | 3 | 1 | 3 | 3 | 2 | 1 | 1 | 1 | 0 | 0 | 4 | 2 | 3 | 0 | 1 | 1 | 1 | 1 | 1 | 0 | 0 | 0 | 1 | 3 | 1 |
| 190 | 101630 | 3 | 0 | 2 | 4 | 2 | 1 | 1 | 1 | 0 | 0 | 1 | 1 | 3 | 0 | 1 | 1 | 1 | 1 | 1 | 0 | 0 | 0 | 1 | 3 | 1 |
| 191 | 10271 | 3 | 0 | 1 | 3 | 2 | 2 | 1 | 1 | 0 | 0 | 1 | 1 | 3 | 0 | 1 | 0 | 1 | 0 | 0 | 0 | 1 | 0 | 0 | 2 | 0 |
| 192 | 10272 | 3 | 0 | 4 | 1 | 2 | 2 | 1 | 1 | 0 | 0 | 1 | 2 | 3 | 0 | 1 | 1 | 1 | 0 | 1 | 0 | 1 | 0 | 0 | 2 | 0 |
| 193 | 10273 | 3 | 0 | 2 | 2 | 2 | 2 | 3 | 1 | 0 | 0 | 1 | 1 | 3 | 0 | 1 | 1 | 1 | 0 | 0 | 0 | 1 | 0 | 0 | 2 | 0 |
| 194 | 10274 | 3 | 1 | 2 | 2 | 2 | 2 | 3 | 1 | 0 | 0 | 4 | 1 | 3 | 0 | 1 | 0 | 1 | 0 | 0 | 0 | 0 | 0 | 1 | 2 | 0 |
| 195 | 10275 | 3 | 0 | 2 | 4 | 1 | 1 | 3 | 1 | 0 | 0 | 1 | 1 | 3 | 1 | 1 | 0 | 1 | 0 | 0 | 0 | 1 | 0 | 1 | 2 | 0 |
| 196 | 10276 | 3 | 1 | 1 | 2 | 2 | 1 | 3 | 1 | 0 | 0 | 2 | 1 | 3 | 0 | 1 | 0 | 0 | 0 | 0 | 0 | 0 | 0 | 0 | 2 | 0 |
| 197 | 10277 | 3 | 0 | 3 | 1 | 2 | 2 | 3 | 1 | 0 | 0 | 1 | 2 | 3 | 0 | 1 | 0 | 1 | 1 | 1 | 0 | 1 | 0 | 1 | 3 | 1 |
| 198 | 10278 | 3 | 0 | 1 | 5 | 1 | 1 | 1 | 1 | 0 | 0 | 1 | 1 | 3 | 0 | 0 | 0 | 0 | 0 | 0 | 0 | 0 | 0 | 0 | 1 | 0 |
| 199 | 10279 | 3 | 1 | 2 | 4 | 2 | 2 | 3 | 1 | 0 | 0 | 2 | 1 | 3 | 1 | 1 | 1 | 1 | 1 | 1 | 0 | 1 | 1 | 1 | 4 | 1 |
| 200 | 102710 | 3 | 1 | 4 | 3 | 2 | 1 | 3 | 1 | 0 | 0 | 2 | 2 | 3 | 0 | 1 | 0 | 1 | 0 | 0 | 0 | 1 | 0 | 1 | 2 | 0 |
| 201 | 102711 | 3 | 1 | 1 | 4 | 2 | 1 | 3 | 1 | 0 | 0 | 2 | 1 | 3 | 0 | 1 | 0 | 0 | 0 | 0 | 0 | 0 | 0 | 0 | 2 | 0 |
| 202 | 102712 | 3 | 0 | 4 | 2 | 2 | 1 | 3 | 1 | 0 | 0 | 1 | 2 | 3 | 1 | 1 | 1 | 1 | 1 | 1 | 0 | 1 | 1 | 0 | 4 | 1 |
| 203 | 102713 | 3 | 0 | 2 | 4 | 2 | 1 | 3 | 1 | 0 | 0 | 1 | 1 | 3 | 0 | 0 | 0 | 0 | 0 | 0 | 0 | 0 | 0 | 0 | 1 | 0 |
| 204 | 102714 | 3 | 0 | 3 | 3 | 1 | 1 | 3 | 1 | 0 | 0 | 1 | 2 | 3 | 0 | 1 | 0 | 0 | 0 | 0 | 0 | 0 | 0 | 0 | 2 | 0 |
| 205 | 102715 | 3 | 1 | 1 | 5 | 1 | 1 | 3 | 1 | 0 | 0 | 2 | 1 | 3 | 0 | 0 | 0 | 0 | 0 | 0 | 0 | 0 | 0 | 0 | 1 | 0 |
| 206 | 102716 | 3 | 1 | 2 | 4 | 1 | 1 | 3 | 1 | 0 | 0 | 2 | 1 | 3 | 0 | 1 | 0 | 1 | 0 | 0 | 0 | 1 | 0 | 0 | 2 | 0 |
| 207 | 102717 | 3 | 0 | 2 | 4 | 1 | 1 | 3 | 1 | 0 | 0 | 1 | 1 | 3 | 0 | 0 | 0 | 0 | 0 | 0 | 0 | 0 | 0 | 0 | 1 | 0 |
| 208 | 102718 | 3 | 1 | 2 | 3 | 1 | 1 | 3 | 1 | 0 | 0 | 2 | 1 | 3 | 1 | 1 | 0 | 1 | 0 | 1 | 0 | 1 | 0 | 1 | 3 | 1 |
| 209 | 102719 | 3 | 0 | 1 | 4 | 1 | 1 | 3 | 1 | 0 | 0 | 2 | 1 | 3 | 0 | 1 | 1 | 1 | 1 | 0 | 0 | 0 | 0 | 1 | 2 | 0 |
| 210 | 102720 | 3 | 1 | 3 | 3 | 2 | 1 | 3 | 1 | 0 | 0 | 4 | 2 | 3 | 0 | 1 | 0 | 1 | 0 | 1 | 0 | 1 | 0 | 1 | 2 | 0 |
| 211 | 102721 | 3 | 1 | 2 | 4 | 2 | 1 | 3 | 1 | 0 | 0 | 2 | 1 | 3 | 1 | 1 | 1 | 1 | 0 | 1 | 0 | 1 | 1 | 1 | 4 | 1 |
| 212 | 102722 | 3 | 0 | 1 | 4 | 1 | 1 | 3 | 1 | 0 | 0 | 1 | 1 | 3 | 0 | 0 | 0 | 0 | 0 | 0 | 0 | 0 | 0 | 0 | 1 | 0 |
| 213 | 102723 | 3 | 0 | 2 | 2 | 1 | 1 | 3 | 1 | 0 | 0 | 1 | 1 | 3 | 0 | 1 | 0 | 0 | 0 | 0 | 0 | 0 | 0 | 0 | 2 | 0 |
| 214 | 102724 | 3 | 1 | 1 | 3 | 2 | 1 | 3 | 1 | 0 | 0 | 2 | 1 | 3 | 0 | 1 | 1 | 1 | 0 | 0 | 0 | 1 | 1 | 1 | 3 | 1 |
| 215 | 102725 | 3 | 0 | 2 | 2 | 1 | 2 | 3 | 1 | 0 | 0 | 1 | 1 | 3 | 0 | 1 | 0 | 1 | 0 | 0 | 0 | 1 | 0 | 1 | 2 | 0 |
| 216 | 102726 | 3 | 1 | 2 | 3 | 2 | 1 | 3 | 1 | 0 | 0 | 2 | 1 | 3 | 0 | 1 | 0 | 0 | 0 | 0 | 0 | 0 | 0 | 0 | 2 | 0 |
| 217 | 102727 | 3 | 1 | 2 | 2 | 2 | 1 | 2 | 1 | 0 | 0 | 2 | 1 | 3 | 1 | 1 | 0 | 1 | 0 | 0 | 0 | 1 | 0 | 1 | 2 | 0 |
| 218 | 102728 | 3 | 1 | 4 | 2 | 2 | 1 | 2 | 1 | 0 | 0 | 2 | 2 | 3 | 1 | 1 | 1 | 1 | 1 | 1 | 0 | 1 | 0 | 1 | 4 | 1 |
| 219 | 102729 | 3 | 1 | 3 | 2 | 2 | 2 | 3 | 1 | 0 | 0 | 4 | 2 | 3 | 0 | 1 | 0 | 1 | 0 | 1 | 0 | 1 | 1 | 1 | 3 | 1 |
| 220 | 102730 | 3 | 1 | 2 | 3 | 2 | 1 | 3 | 1 | 0 | 0 | 4 | 1 | 3 | 0 | 1 | 0 | 1 | 0 | 1 | 1 | 0 | 1 | 0 | 2 | 0 |
| 221 | 10281 | 3 | 1 | 2 | 3 | 2 | 1 | 4 | 1 | 0 | 0 | 2 | 1 | 3 | 0 | 1 | 1 | 1 | 0 | 0 | 0 | 1 | 0 | 0 | 2 | 0 |
| 222 | 10282 | 3 | 1 | 2 | 4 | 1 | 1 | 1 | 1 | 0 | 0 | 2 | 1 | 3 | 0 | 0 | 0 | 0 | 0 | 0 | 0 | 0 | 0 | 0 | 1 | 0 |
| 223 | 10283 | 3 | 0 | 1 | 4 | 1 | 1 | 4 | 1 | 0 | 0 | 1 | 1 | 3 | 0 | 1 | 1 | 1 | 0 | 0 | 0 | 0 | 0 | 0 | 2 | 0 |
| 224 | 10284 | 3 | 1 | 1 | 3 | 2 | 1 | 4 | 1 | 0 | 0 | 1 | 1 | 3 | 0 | 1 | 0 | 1 | 0 | 0 | 0 | 0 | 0 | 0 | 2 | 0 |
| 225 | 10285 | 3 | 1 | 2 | 4 | 2 | 1 | 4 | 1 | 0 | 0 | 2 | 1 | 3 | 1 | 1 | 0 | 1 | 0 | 1 | 0 | 1 | 0 | 0 | 2 | 0 |
| 226 | 10286 | 3 | 0 | 2 | 3 | 1 | 1 | 4 | 1 | 0 | 0 | 1 | 1 | 3 | 0 | 1 | 0 | 0 | 0 | 0 | 0 | 0 | 0 | 0 | 2 | 0 |
| 227 | 10287 | 3 | 0 | 1 | 5 | 1 | 1 | 4 | 1 | 0 | 0 | 1 | 1 | 3 | 0 | 0 | 0 | 0 | 0 | 0 | 0 | 0 | 0 | 0 | 1 | 0 |
| 228 | 10288 | 3 | 1 | 1 | 5 | 1 | 1 | 4 | 1 | 0 | 0 | 2 | 1 | 3 | 0 | 0 | 0 | 0 | 0 | 0 | 0 | 0 | 0 | 0 | 1 | 0 |
| 229 | 10289 | 3 | 1 | 3 | 1 | 1 | 1 | 4 | 1 | 0 | 0 | 2 | 2 | 3 | 0 | 1 | 0 | 1 | 0 | 1 | 0 | 1 | 0 | 0 | 2 | 0 |
| 230 | 102810 | 3 | 0 | 2 | 3 | 1 | 1 | 4 | 1 | 0 | 0 | 1 | 1 | 3 | 0 | 1 | 0 | 1 | 0 | 0 | 0 | 1 | 0 | 0 | 2 | 0 |
| 231 | 102811 | 3 | 1 | 3 | 3 | 2 | 1 | 4 | 1 | 0 | 0 | 2 | 2 | 3 | 1 | 1 | 0 | 1 | 1 | 1 | 0 | 1 | 0 | 0 | 3 | 1 |
| 232 | 102812 | 3 | 1 | 3 | 3 | 2 | 1 | 4 | 1 | 0 | 0 | 2 | 2 | 3 | 1 | 1 | 1 | 1 | 0 | 1 | 1 | 0 | 0 | 0 | 3 | 1 |
| 233 | 102813 | 3 | 1 | 2 | 4 | 1 | 1 | 4 | 1 | 0 | 0 | 2 | 1 | 3 | 0 | 1 | 0 | 1 | 0 | 0 | 0 | 0 | 1 | 0 | 2 | 0 |
| 234 | 102814 | 3 | 0 | 2 | 3 | 1 | 1 | 4 | 1 | 0 | 0 | 1 | 1 | 3 | 0 | 0 | 0 | 0 | 0 | 0 | 0 | 0 | 0 | 0 | 1 | 0 |
| 235 | 102815 | 3 | 1 | 1 | 5 | 1 | 1 | 4 | 1 | 0 | 0 | 2 | 1 | 3 | 0 | 0 | 0 | 0 | 0 | 0 | 0 | 0 | 0 | 0 | 1 | 0 |
| 236 | 102816 | 3 | 1 | 2 | 3 | 1 | 1 | 4 | 1 | 0 | 0 | 2 | 1 | 3 | 1 | 1 | 0 | 1 | 0 | 1 | 0 | 1 | 0 | 0 | 2 | 0 |
| 237 | 102817 | 3 | 0 | 2 | 4 | 1 | 1 | 4 | 1 | 0 | 0 | 1 | 1 | 3 | 1 | 0 | 0 | 0 | 0 | 0 | 0 | 0 | 0 | 0 | 2 | 0 |
| 238 | 102818 | 3 | 0 | 2 | 4 | 1 | 1 | 4 | 1 | 0 | 0 | 1 | 1 | 3 | 0 | 1 | 0 | 1 | 0 | 0 | 0 | 1 | 0 | 0 | 2 | 0 |
| 239 | 102819 | 3 | 1 | 2 | 3 | 1 | 1 | 4 | 1 | 0 | 0 | 2 | 1 | 3 | 0 | 1 | 0 | 1 | 0 | 0 | 0 | 1 | 0 | 1 | 2 | 0 |
| 240 | 102820 | 3 | 0 | 3 | 2 | 1 | 1 | 4 | 1 | 0 | 0 | 1 | 2 | 3 | 0 | 1 | 0 | 1 | 0 | 0 | 0 | 0 | 0 | 1 | 2 | 0 |
| 241 | 102821 | 3 | 1 | 3 | 2 | 3 | 1 | 1 | 1 | 0 | 0 | 2 | 2 | 3 | 0 | 1 | 1 | 1 | 1 | 1 | 1 | 1 | 0 | 1 | 4 | 1 |
| 242 | 102822 | 3 | 0 | 2 | 3 | 2 | 2 | 1 | 1 | 0 | 0 | 1 | 1 | 3 | 0 | 1 | 0 | 1 | 0 | 1 | 1 | 1 | 0 | 1 | 3 | 1 |
| 243 | 102823 | 3 | 0 | 2 | 3 | 3 | 1 | 1 | 1 | 0 | 0 | 1 | 1 | 3 | 1 | 1 | 1 | 1 | 1 | 1 | 0 | 1 | 0 | 0 | 3 | 1 |
| 244 | 102824 | 3 | 1 | 1 | 4 | 2 | 1 | 1 | 1 | 0 | 0 | 2 | 1 | 3 | 0 | 0 | 0 | 0 | 0 | 0 | 0 | 0 | 0 | 0 | 1 | 0 |
| 245 | 102825 | 3 | 1 | 2 | 4 | 2 | 1 | 1 | 1 | 0 | 0 | 2 | 1 | 3 | 1 | 1 | 0 | 1 | 0 | 0 | 0 | 0 | 0 | 0 | 2 | 0 |
| 246 | 102826 | 3 | 0 | 2 | 4 | 2 | 1 | 1 | 1 | 0 | 0 | 1 | 1 | 3 | 1 | 1 | 0 | 1 | 0 | 1 | 0 | 1 | 0 | 0 | 2 | 0 |
| 247 | 102827 | 3 | 0 | 2 | 5 | 3 | 1 | 1 | 1 | 0 | 0 | 1 | 1 | 3 | 1 | 1 | 1 | 1 | 1 | 1 | 1 | 1 | 1 | 0 | 4 | 1 |
| 248 | 102828 | 3 | 0 | 2 | 2 | 3 | 1 | 1 | 1 | 0 | 0 | 1 | 1 | 3 | 1 | 1 | 1 | 1 | 1 | 1 | 1 | 1 | 0 | 0 | 4 | 1 |
| 249 | 102829 | 3 | 1 | 1 | 5 | 3 | 1 | 1 | 1 | 0 | 0 | 2 | 1 | 3 | 1 | 1 | 0 | 1 | 0 | 1 | 1 | 1 | 1 | 0 | 3 | 1 |
| 250 | 102830 | 3 | 0 | 3 | 1 | 2 | 1 | 1 | 1 | 0 | 0 | 1 | 2 | 3 | 1 | 1 | 1 | 1 | 1 | 1 | 0 | 1 | 1 | 0 | 4 | 1 |
| 251 | 10291 | 3 | 1 | 2 | 5 | 2 | 1 | 2 | 1 | 0 | 0 | 2 | 1 | 3 | 1 | 1 | 0 | 1 | 0 | 1 | 0 | 1 | 1 | 0 | 3 | 1 |
| 252 | 10292 | 3 | 0 | 2 | 4 | 2 | 1 | 2 | 1 | 0 | 0 | 1 | 1 | 3 | 0 | 0 | 0 | 0 | 0 | 0 | 0 | 0 | 0 | 0 | 1 | 0 |
| 253 | 10293 | 3 | 0 | 1 | 5 | 2 | 1 | 2 | 1 | 0 | 0 | 1 | 1 | 3 | 0 | 0 | 0 | 0 | 0 | 0 | 0 | 0 | 0 | 0 | 1 | 0 |
| 254 | 10294 | 3 | 1 | 3 | 1 | 2 | 1 | 2 | 1 | 0 | 0 | 2 | 2 | 3 | 1 | 1 | 0 | 1 | 1 | 1 | 0 | 1 | 0 | 0 | 3 | 1 |
| 255 | 10295 | 3 | 1 | 3 | 2 | 2 | 1 | 2 | 1 | 0 | 0 | 2 | 2 | 3 | 1 | 1 | 0 | 1 | 0 | 0 | 0 | 0 | 0 | 1 | 2 | 0 |
| 256 | 10296 | 3 | 1 | 2 | 5 | 3 | 2 | 2 | 1 | 0 | 0 | 2 | 1 | 3 | 0 | 1 | 0 | 1 | 1 | 1 | 0 | 1 | 0 | 0 | 2 | 0 |
| 257 | 10297 | 3 | 1 | 2 | 3 | 2 | 1 | 2 | 1 | 0 | 0 | 2 | 1 | 3 | 0 | 0 | 0 | 0 | 0 | 0 | 0 | 0 | 0 | 0 | 1 | 0 |
| 258 | 10298 | 3 | 0 | 4 | 1 | 3 | 1 | 2 | 1 | 0 | 0 | 1 | 2 | 3 | 0 | 1 | 0 | 1 | 1 | 1 | 0 | 1 | 0 | 0 | 2 | 0 |
| 259 | 10299 | 3 | 1 | 2 | 4 | 2 | 1 | 2 | 1 | 0 | 0 | 2 | 1 | 3 | 0 | 1 | 0 | 1 | 0 | 1 | 0 | 0 | 0 | 0 | 2 | 0 |
| 260 | 102910 | 3 | 0 | 2 | 3 | 2 | 2 | 2 | 1 | 0 | 0 | 1 | 1 | 3 | 1 | 1 | 0 | 1 | 0 | 0 | 0 | 0 | 0 | 1 | 2 | 0 |
| 261 | 102911 | 3 | 1 | 3 | 3 | 2 | 1 | 2 | 1 | 0 | 0 | 2 | 2 | 3 | 0 | 1 | 0 | 1 | 0 | 1 | 0 | 0 | 0 | 1 | 2 | 0 |
| 262 | 102912 | 3 | 0 | 3 | 1 | 2 | 1 | 2 | 1 | 0 | 0 | 1 | 2 | 3 | 0 | 1 | 1 | 1 | 0 | 0 | 0 | 1 | 0 | 0 | 2 | 0 |
| 263 | 102913 | 3 | 1 | 2 | 3 | 2 | 1 | 2 | 1 | 0 | 0 | 2 | 1 | 3 | 0 | 1 | 1 | 1 | 1 | 1 | 1 | 1 | 1 | 1 | 4 | 1 |
| 264 | 102914 | 3 | 0 | 2 | 4 | 2 | 1 | 2 | 1 | 0 | 0 | 1 | 1 | 3 | 1 | 1 | 0 | 0 | 0 | 0 | 0 | 0 | 0 | 0 | 2 | 0 |
| 265 | 102915 | 3 | 0 | 1 | 4 | 2 | 1 | 2 | 1 | 0 | 0 | 1 | 1 | 3 | 0 | 1 | 0 | 0 | 0 | 0 | 0 | 0 | 0 | 0 | 2 | 0 |
| 266 | 102916 | 3 | 1 | 2 | 4 | 2 | 1 | 2 | 1 | 0 | 0 | 2 | 1 | 3 | 0 | 0 | 0 | 0 | 0 | 0 | 0 | 0 | 0 | 0 | 1 | 0 |
| 267 | 102917 | 3 | 1 | 2 | 3 | 2 | 1 | 2 | 1 | 0 | 0 | 2 | 1 | 3 | 1 | 1 | 0 | 1 | 0 | 0 | 0 | 0 | 0 | 0 | 2 | 0 |
| 268 | 102918 | 3 | 0 | 2 | 4 | 2 | 1 | 2 | 1 | 0 | 0 | 1 | 1 | 3 | 1 | 0 | 0 | 0 | 0 | 0 | 0 | 0 | 0 | 0 | 2 | 0 |
| 269 | 102919 | 3 | 0 | 2 | 4 | 2 | 1 | 2 | 1 | 0 | 0 | 1 | 1 | 3 | 0 | 0 | 0 | 0 | 0 | 0 | 0 | 0 | 0 | 0 | 1 | 0 |
| 270 | 102920 | 3 | 0 | 3 | 1 | 2 | 1 | 2 | 1 | 0 | 0 | 1 | 2 | 3 | 0 | 1 | 1 | 1 | 0 | 0 | 0 | 1 | 0 | 0 | 2 | 0 |
| 271 | 102921 | 3 | 0 | 2 | 1 | 2 | 1 | 2 | 1 | 0 | 0 | 1 | 1 | 3 | 0 | 1 | 0 | 1 | 1 | 1 | 0 | 1 | 0 | 0 | 2 | 0 |
| 272 | 102922 | 3 | 1 | 2 | 3 | 2 | 1 | 2 | 1 | 0 | 0 | 2 | 1 | 3 | 0 | 1 | 0 | 0 | 0 | 0 | 0 | 0 | 0 | 0 | 2 | 0 |
| 273 | 102923 | 3 | 1 | 2 | 3 | 2 | 1 | 2 | 1 | 0 | 0 | 2 | 1 | 3 | 1 | 1 | 0 | 1 | 0 | 0 | 0 | 0 | 0 | 0 | 2 | 0 |
| 274 | 102924 | 3 | 1 | 3 | 2 | 2 | 1 | 2 | 1 | 0 | 0 | 2 | 2 | 3 | 1 | 0 | 0 | 0 | 0 | 0 | 0 | 0 | 0 | 0 | 2 | 0 |
| 275 | 102925 | 3 | 0 | 4 | 2 | 3 | 1 | 2 | 1 | 0 | 0 | 1 | 2 | 3 | 0 | 1 | 1 | 1 | 1 | 1 | 0 | 1 | 1 | 1 | 4 | 1 |
| 276 | 102926 | 3 | 1 | 2 | 3 | 2 | 1 | 2 | 1 | 0 | 0 | 2 | 1 | 3 | 0 | 1 | 0 | 0 | 0 | 0 | 0 | 0 | 0 | 0 | 2 | 0 |
| 277 | 102927 | 3 | 1 | 2 | 3 | 2 | 1 | 2 | 1 | 0 | 0 | 2 | 1 | 3 | 1 | 0 | 0 | 0 | 0 | 0 | 0 | 0 | 0 | 0 | 2 | 0 |
| 278 | 102928 | 3 | 0 | 3 | 4 | 2 | 1 | 2 | 1 | 0 | 0 | 1 | 2 | 3 | 0 | 1 | 0 | 0 | 0 | 0 | 0 | 0 | 0 | 0 | 2 | 0 |
| 279 | 102929 | 3 | 0 | 2 | 4 | 2 | 1 | 2 | 1 | 0 | 0 | 1 | 1 | 3 | 0 | 1 | 0 | 0 | 1 | 0 | 0 | 0 | 0 | 0 | 2 | 0 |
| 280 | 102930 | 3 | 1 | 2 | 4 | 2 | 1 | 2 | 1 | 0 | 0 | 2 | 1 | 3 | 0 | 1 | 0 | 1 | 0 | 0 | 0 | 1 | 0 | 0 | 2 | 0 |
| 281 | 102101 | 3 | 0 | 4 | 1 | 2 | 2 | 1 | 1 | 0 | 0 | 1 | 2 | 3 | 1 | 1 | 1 | 1 | 1 | 1 | 1 | 1 | 1 | 0 | 4 | 1 |

| | | | | | | | | | | | | | | | | | | | | | | | | | | |
|---|---|---|---|---|---|---|---|---|---|---|---|---|---|---|---|---|---|---|---|---|---|---|---|---|---|---|
| 282 | 102102 | 3 | 0 | 3 | 2 | 2 | 1 | 1 | 1 | 0 | 0 | 1 | 2 | 3 | 0 | 1 | 1 | 1 | 1 | 1 | 1 | 1 | 0 | 0 | 3 | 1 |
| 283 | 102103 | 3 | 0 | 2 | 2 | 2 | 2 | 1 | 2 | 0 | 0 | 1 | 1 | 3 | 0 | 1 | 1 | 1 | 1 | 1 | 0 | 1 | 0 | 0 | 3 | 1 |
| 284 | 102104 | 3 | 0 | 1 | 5 | 2 | 2 | 1 | 1 | 0 | 0 | 2 | 1 | 3 | 0 | 1 | 0 | 1 | 1 | 1 | 1 | 0 | 1 | 0 | 3 | 1 |
| 285 | 102105 | 3 | 1 | 3 | 5 | 3 | 2 | 1 | 1 | 0 | 0 | 2 | 2 | 3 | 1 | 1 | 1 | 1 | 1 | 1 | 0 | 1 | 0 | 0 | 3 | 1 |
| 286 | 102106 | 3 | 1 | 1 | 4 | 2 | 2 | 2 | 4 | 0 | 0 | 2 | 1 | 3 | 0 | 1 | 0 | 1 | 1 | 1 | 0 | 1 | 0 | 0 | 2 | 0 |
| 287 | 102107 | 3 | 0 | 4 | 1 | 2 | 1 | 1 | 4 | 0 | 0 | 1 | 2 | 3 | 0 | 1 | 1 | 1 | 1 | 1 | 0 | 1 | 0 | 0 | 3 | 1 |
| 288 | 102108 | 3 | 0 | 2 | 2 | 2 | 2 | 1 | 1 | 0 | 0 | 1 | 1 | 3 | 0 | 1 | 1 | 1 | 1 | 1 | 0 | 1 | 0 | 0 | 3 | 1 |
| 289 | 102109 | 3 | 1 | 2 | 3 | 3 | 1 | 1 | 1 | 0 | 0 | 2 | 1 | 3 | 0 | 1 | 1 | 1 | 1 | 1 | 0 | 1 | 0 | 0 | 3 | 1 |
| 290 | 1021010 | 3 | 0 | 2 | 3 | 2 | 1 | 1 | 1 | 0 | 0 | 1 | 1 | 3 | 0 | 1 | 1 | 1 | 1 | 1 | 1 | 1 | 0 | 0 | 3 | 1 |
| 291 | 1021011 | 3 | 1 | 1 | 4 | 2 | 1 | 1 | 1 | 0 | 0 | 2 | 1 | 2 | 0 | 1 | 0 | 1 | 1 | 1 | 0 | 1 | 0 | 0 | 2 | 0 |
| 292 | 1021012 | 3 | 0 | 2 | 1 | 3 | 1 | 1 | 1 | 0 | 0 | 1 | 1 | 3 | 0 | 1 | 1 | 1 | 1 | 1 | 0 | 1 | 0 | 0 | 3 | 1 |
| 293 | 1021013 | 3 | 0 | 1 | 5 | 2 | 1 | 1 | 4 | 0 | 0 | 2 | 1 | 3 | 0 | 1 | 0 | 1 | 0 | 1 | 0 | 0 | 0 | 0 | 2 | 0 |
| 294 | 1021014 | 3 | 0 | 2 | 2 | 2 | 1 | 1 | 1 | 0 | 0 | 1 | 1 | 3 | 0 | 1 | 0 | 1 | 1 | 1 | 0 | 1 | 0 | 0 | 2 | 0 |
| 295 | 1021015 | 3 | 1 | 1 | 5 | 2 | 1 | 1 | 1 | 0 | 0 | 2 | 1 | 3 | 1 | 1 | 1 | 1 | 1 | 1 | 0 | 1 | 0 | 0 | 3 | 1 |
| 296 | 1021016 | 3 | 1 | 2 | 4 | 2 | 2 | 1 | 4 | 0 | 0 | 2 | 1 | 3 | 0 | 1 | 0 | 1 | 1 | 1 | 1 | 0 | 0 | 0 | 2 | 0 |
| 297 | 1021017 | 3 | 1 | 3 | 2 | 2 | 2 | 4 | 2 | 0 | 0 | 4 | 2 | 3 | 0 | 1 | 1 | 1 | 1 | 1 | 0 | 1 | 0 | 0 | 3 | 1 |
| 298 | 1021018 | 3 | 1 | 1 | 2 | 2 | 2 | 1 | 1 | 0 | 0 | 2 | 1 | 3 | 0 | 1 | 0 | 1 | 1 | 1 | 1 | 1 | 0 | 0 | 3 | 1 |
| 299 | 1021019 | 3 | 1 | 1 | 4 | 2 | 1 | 1 | 1 | 0 | 0 | 2 | 1 | 3 | 0 | 1 | 0 | 1 | 1 | 1 | 0 | 1 | 0 | 0 | 2 | 0 |
| 300 | 1021020 | 3 | 1 | 2 | 5 | 2 | 1 | 1 | 1 | 0 | 0 | 2 | 1 | 2 | 0 | 1 | 0 | 1 | 1 | 1 | 0 | 1 | 0 | 0 | 2 | 0 |
| 301 | 1021021 | 3 | 1 | 1 | 4 | 2 | 1 | 1 | 1 | 0 | 0 | 2 | 1 | 3 | 0 | 1 | 0 | 1 | 1 | 1 | 0 | 0 | 1 | 0 | 2 | 0 |
| 302 | 1021022 | 3 | 1 | 1 | 3 | 2 | 1 | 1 | 1 | 0 | 0 | 2 | 1 | 3 | 0 | 0 | 0 | 0 | 0 | 0 | 0 | 0 | 0 | 0 | 1 | 0 |
| 303 | 1021023 | 3 | 1 | 2 | 4 | 3 | 2 | 1 | 4 | 1 | 0 | 5 | 1 | 3 | 0 | 1 | 1 | 1 | 1 | 1 | 0 | 0 | 1 | 0 | 3 | 1 |
| 304 | 1021024 | 3 | 1 | 1 | 3 | 2 | 1 | 1 | 1 | 0 | 0 | 2 | 1 | 3 | 0 | 1 | 1 | 1 | 1 | 1 | 0 | 1 | 0 | 0 | 3 | 1 |
| 305 | 1021025 | 3 | 1 | 2 | 4 | 2 | 2 | 2 | 4 | 0 | 0 | 5 | 1 | 3 | 0 | 1 | 1 | 1 | 1 | 1 | 0 | 1 | 0 | 0 | 3 | 1 |
| 306 | 1021026 | 3 | 1 | 2 | 4 | 2 | 1 | 1 | 1 | 0 | 0 | 2 | 1 | 3 | 0 | 1 | 0 | 1 | 1 | 1 | 1 | 0 | 0 | 0 | 2 | 0 |
| 307 | 1021027 | 3 | 0 | 1 | 5 | 1 | 1 | 1 | 1 | 0 | 0 | 2 | 1 | 3 | 0 | 1 | 0 | 0 | 0 | 0 | 0 | 0 | 0 | 0 | 2 | 0 |
| 308 | 1021028 | 3 | 0 | 1 | 5 | 2 | 2 | 2 | 4 | 0 | 0 | 2 | 1 | 3 | 1 | 1 | 1 | 0 | 1 | 1 | 0 | 0 | 0 | 1 | 3 | 1 |
| 309 | 1021029 | 3 | 0 | 1 | 5 | 2 | 1 | 1 | 1 | 0 | 0 | 2 | 1 | 3 | 1 | 1 | 0 | 1 | 1 | 1 | 0 | 1 | 0 | 0 | 3 | 1 |
| 310 | 1021030 | 3 | 0 | 2 | 5 | 3 | 2 | 1 | 1 | 0 | 0 | 2 | 1 | 3 | 1 | 1 | 0 | 1 | 1 | 1 | 0 | 1 | 0 | 0 | 3 | 1 |
| 311 | 1021031 | 3 | 0 | 3 | 2 | 2 | 1 | 4 | 2 | 0 | 0 | 1 | 2 | 3 | 0 | 1 | 1 | 1 | 0 | 1 | 0 | 1 | 0 | 1 | 3 | 1 |
| 312 | 1021032 | 3 | 0 | 4 | 2 | 3 | 2 | 1 | 4 | 0 | 0 | 1 | 2 | 3 | 0 | 1 | 1 | 1 | 1 | 1 | 0 | 1 | 0 | 0 | 3 | 1 |
| 313 | 1021033 | 3 | 0 | 3 | 2 | 2 | 1 | 1 | 1 | 0 | 0 | 1 | 2 | 3 | 1 | 1 | 1 | 1 | 1 | 1 | 0 | 1 | 0 | 0 | 3 | 1 |
| 314 | 102111 | 3 | 0 | 2 | 3 | 1 | 1 | 1 | 2 | 0 | 0 | 2 | 1 | 3 | 0 | 0 | 0 | 0 | 0 | 0 | 0 | 0 | 0 | 0 | 1 | 0 |
| 315 | 102112 | 3 | 1 | 2 | 3 | 2 | 1 | 1 | 1 | 0 | 0 | 3 | 1 | 3 | 0 | 1 | 0 | 1 | 0 | 1 | 0 | 1 | 0 | 0 | 2 | 0 |
| 316 | 102113 | 3 | 0 | 1 | 4 | 2 | 1 | 1 | 2 | 0 | 0 | 2 | 1 | 3 | 0 | 1 | 0 | 1 | 1 | 1 | 0 | 1 | 0 | 0 | 2 | 0 |
| 317 | 102114 | 3 | 0 | 3 | 3 | 2 | 1 | 3 | 4 | 0 | 0 | 1 | 2 | 3 | 0 | 1 | 0 | 1 | 0 | 1 | 0 | 1 | 0 | 0 | 2 | 0 |
| 318 | 102115 | 3 | 1 | 2 | 2 | 2 | 1 | 3 | 4 | 0 | 0 | 4 | 1 | 3 | 1 | 1 | 0 | 1 | 1 | 0 | 0 | 1 | 0 | 0 | 2 | 0 |
| 319 | 102116 | 3 | 0 | 2 | 2 | 2 | 2 | 3 | 4 | 0 | 0 | 1 | 1 | 2 | 0 | 1 | 1 | 1 | 1 | 0 | 0 | 1 | 0 | 0 | 2 | 0 |
| 320 | 102117 | 3 | 1 | 5 | 2 | 3 | 1 | 1 | 4 | 1 | 0 | 5 | 3 | 3 | 1 | 1 | 1 | 1 | 1 | 1 | 0 | 1 | 1 | 1 | 4 | 1 |
| 321 | 102118 | 3 | 1 | 3 | 1 | 2 | 2 | 4 | 4 | 0 | 0 | 5 | 2 | 3 | 0 | 1 | 1 | 1 | 1 | 1 | 0 | 1 | 1 | 1 | 4 | 1 |
| 322 | 102119 | 3 | 1 | 4 | 5 | 2 | 1 | 1 | 2 | 0 | 0 | 4 | 2 | 3 | 1 | 1 | 1 | 1 | 1 | 1 | 0 | 1 | 1 | 1 | 4 | 1 |
| 323 | 1021110 | 3 | 0 | 1 | 4 | 2 | 1 | 2 | 4 | 0 | 0 | 2 | 1 | 3 | 0 | 0 | 0 | 0 | 0 | 0 | 0 | 0 | 0 | 0 | 1 | 0 |
| 324 | 1021111 | 3 | 1 | 4 | 2 | 2 | 1 | 2 | 4 | 0 | 0 | 4 | 2 | 3 | 0 | 1 | 1 | 1 | 1 | 0 | 0 | 1 | 0 | 0 | 2 | 0 |
| 325 | 1021112 | 3 | 1 | 3 | 2 | 2 | 1 | 2 | 4 | 0 | 0 | 2 | 2 | 3 | 1 | 1 | 1 | 1 | 0 | 1 | 0 | 1 | 0 | 1 | 3 | 1 |
| 326 | 1021113 | 3 | 1 | 1 | 1 | 2 | 1 | 1 | 2 | 0 | 0 | 4 | 1 | 2 | 0 | 1 | 0 | 0 | 0 | 0 | 0 | 0 | 0 | 0 | 2 | 0 |
| 327 | 1021114 | 3 | 0 | 1 | 3 | 1 | 1 | 1 | 2 | 0 | 0 | 2 | 1 | 3 | 0 | 1 | 0 | 0 | 0 | 0 | 0 | 0 | 0 | 0 | 2 | 0 |
| 328 | 1021115 | 3 | 0 | 2 | 3 | 2 | 1 | 1 | 2 | 0 | 0 | 1 | 1 | 3 | 0 | 1 | 1 | 1 | 0 | 1 | 0 | 1 | 0 | 0 | 2 | 0 |
| 329 | 1021116 | 3 | 0 | 2 | 3 | 2 | 2 | 1 | 1 | 0 | 0 | 1 | 1 | 3 | 0 | 1 | 0 | 1 | 1 | 1 | 0 | 1 | 0 | 0 | 2 | 0 |
| 330 | 1021117 | 3 | 1 | 2 | 5 | 2 | 1 | 3 | 4 | 1 | 1 | 5 | 1 | 2 | 1 | 1 | 1 | 1 | 1 | 1 | 0 | 1 | 0 | 0 | 3 | 1 |
| 331 | 1021118 | 3 | 0 | 2 | 4 | 2 | 1 | 3 | 4 | 0 | 0 | 1 | 1 | 3 | 1 | 1 | 1 | 1 | 1 | 1 | 0 | 1 | 0 | 1 | 4 | 1 |
| 332 | 1021119 | 3 | 0 | 2 | 4 | 2 | 2 | 3 | 4 | 0 | 0 | 1 | 1 | 3 | 1 | 1 | 0 | 1 | 1 | 1 | 0 | 1 | 0 | 0 | 3 | 1 |
| 333 | 1021120 | 3 | 1 | 1 | 3 | 2 | 2 | 2 | 4 | 0 | 0 | 2 | 1 | 3 | 0 | 1 | 0 | 1 | 1 | 1 | 0 | 1 | 0 | 1 | 3 | 1 |
| 334 | 1021121 | 3 | 1 | 1 | 5 | 1 | 1 | 1 | 4 | 0 | 0 | 2 | 1 | 3 | 0 | 0 | 0 | 0 | 0 | 0 | 0 | 0 | 0 | 0 | 1 | 0 |
| 335 | 1021122 | 3 | 1 | 1 | 5 | 3 | 2 | 2 | 4 | 0 | 0 | 2 | 1 | 3 | 0 | 1 | 1 | 1 | 1 | 1 | 0 | 1 | 0 | 0 | 3 | 1 |
| 336 | 1021123 | 3 | 0 | 2 | 5 | 1 | 1 | 3 | 4 | 0 | 0 | 2 | 1 | 3 | 0 | 1 | 0 | 1 | 1 | 1 | 0 | 0 | 0 | 0 | 2 | 0 |
| 337 | 1021124 | 3 | 1 | 1 | 4 | 2 | 2 | 4 | 4 | 0 | 0 | 4 | 1 | 3 | 0 | 1 | 1 | 1 | 1 | 1 | 0 | 1 | 0 | 0 | 3 | 1 |
| 338 | 1021125 | 3 | 1 | 3 | 2 | 3 | 1 | 2 | 4 | 0 | 0 | 4 | 2 | 3 | 0 | 1 | 1 | 1 | 1 | 1 | 0 | 1 | 0 | 0 | 3 | 1 |
| 339 | 1021126 | 3 | 0 | 2 | 4 | 3 | 2 | 3 | 4 | 0 | 0 | 5 | 1 | 3 | 0 | 1 | 1 | 1 | 1 | 1 | 0 | 1 | 0 | 0 | 3 | 1 |
| 340 | 1021127 | 3 | 1 | 1 | 3 | 2 | 1 | 2 | 4 | 0 | 0 | 4 | 1 | 3 | 1 | 1 | 1 | 1 | 1 | 1 | 0 | 0 | 1 | 1 | 4 | 1 |
| 341 | 1021128 | 3 | 1 | 4 | 3 | 3 | 2 | 1 | 4 | 0 | 0 | 5 | 2 | 3 | 1 | 1 | 1 | 1 | 1 | 1 | 0 | 0 | 1 | 1 | 4 | 1 |
| 342 | 1021129 | 3 | 1 | 4 | 3 | 3 | 2 | 1 | 4 | 0 | 0 | 5 | 2 | 3 | 0 | 1 | 1 | 1 | 1 | 1 | 0 | 0 | 1 | 1 | 3 | 1 |
| 343 | 1021130 | 3 | 1 | 4 | 3 | 3 | 1 | 3 | 4 | 0 | 0 | 5 | 2 | 3 | 1 | 1 | 1 | 1 | 1 | 1 | 0 | 1 | 1 | 1 | 4 | 1 |
| 344 | 1021131 | 3 | 0 | 4 | 2 | 2 | 1 | 3 | 4 | 0 | 0 | 5 | 2 | 3 | 1 | 1 | 1 | 1 | 1 | 1 | 0 | 1 | 1 | 1 | 4 | 1 |
| 345 | 1021132 | 3 | 0 | 4 | 3 | 3 | 2 | 2 | 4 | 0 | 0 | 1 | 2 | 3 | 0 | 1 | 1 | 1 | 1 | 1 | 0 | 1 | 1 | 1 | 4 | 1 |
| 346 | 1021133 | 3 | 0 | 2 | 4 | 3 | 1 | 3 | 4 | 0 | 0 | 5 | 1 | 3 | 1 | 1 | 1 | 1 | 1 | 1 | 0 | 1 | 0 | 0 | 3 | 1 |
| 347 | 1021134 | 3 | 1 | 4 | 4 | 3 | 2 | 2 | 4 | 0 | 0 | 5 | 2 | 3 | 1 | 1 | 1 | 1 | 1 | 1 | 0 | 1 | 1 | 1 | 4 | 1 |
| 348 | 1021135 | 3 | 1 | 5 | 2 | 3 | 1 | 2 | 4 | 0 | 0 | 5 | 3 | 2 | 1 | 1 | 1 | 1 | 1 | 1 | 0 | 1 | 1 | 1 | 4 | 1 |
| 349 | 1021136 | 3 | 0 | 4 | 2 | 2 | 2 | 4 | 4 | 0 | 0 | 1 | 2 | 3 | 1 | 1 | 1 | 1 | 1 | 1 | 0 | 1 | 1 | 1 | 4 | 1 |
| 350 | 1021137 | 3 | 0 | 2 | 5 | 2 | 2 | 3 | 4 | 0 | 0 | 2 | 1 | 3 | 1 | 1 | 1 | 1 | 1 | 1 | 0 | 1 | 0 | 0 | 3 | 1 |
| 351 | 1021138 | 3 | 1 | 5 | 2 | 3 | 1 | 2 | 4 | 0 | 0 | 5 | 3 | 2 | 1 | 1 | 1 | 1 | 1 | 1 | 0 | 1 | 1 | 1 | 4 | 1 |
| 352 | 1021139 | 3 | 0 | 2 | 4 | 2 | 1 | 2 | 4 | 0 | 0 | 1 | 1 | 3 | 1 | 1 | 0 | 1 | 1 | 1 | 0 | 0 | 1 | 0 | 3 | 1 |
| 353 | 102121 | 3 | 0 | 1 | 5 | 2 | 1 | 1 | 2 | 0 | 0 | 2 | 1 | 3 | 0 | 0 | 0 | 0 | 0 | 0 | 0 | 0 | 0 | 0 | 1 | 0 |
| 354 | 102122 | 3 | 0 | 2 | 4 | 3 | 1 | 1 | 1 | 0 | 0 | 2 | 1 | 3 | 1 | 1 | 0 | 1 | 1 | 1 | 0 | 1 | 0 | 0 | 3 | 1 |
| 355 | 102123 | 3 | 1 | 5 | 5 | 3 | 1 | 1 | 2 | 0 | 0 | 5 | 3 | 2 | 1 | 1 | 0 | 1 | 1 | 1 | 0 | 1 | 1 | 0 | 3 | 1 |
| 356 | 102124 | 3 | 1 | 1 | 4 | 2 | 1 | 1 | 1 | 0 | 0 | 2 | 1 | 3 | 0 | 0 | 0 | 0 | 0 | 0 | 0 | 0 | 0 | 0 | 1 | 0 |
| 357 | 102125 | 3 | 0 | 3 | 3 | 2 | 2 | 1 | 4 | 0 | 0 | 1 | 2 | 3 | 0 | 1 | 1 | 1 | 0 | 1 | 0 | 1 | 0 | 0 | 2 | 0 |
| 358 | 102126 | 3 | 1 | 2 | 3 | 2 | 1 | 1 | 2 | 0 | 0 | 2 | 1 | 3 | 0 | 1 | 1 | 1 | 0 | 0 | 0 | 0 | 0 | 0 | 2 | 0 |
| 359 | 102127 | 3 | 0 | 1 | 3 | 2 | 2 | 1 | 1 | 0 | 0 | 1 | 1 | 3 | 0 | 1 | 0 | 1 | 1 | 1 | 0 | 1 | 0 | 1 | 3 | 1 |
| 360 | 102128 | 3 | 1 | 3 | 3 | 3 | 1 | 1 | 4 | 0 | 0 | 2 | 2 | 3 | 0 | 1 | 0 | 1 | 1 | 1 | 0 | 1 | 0 | 0 | 2 | 0 |
| 361 | 102129 | 3 | 0 | 4 | 1 | 3 | 1 | 1 | 4 | 0 | 0 | 1 | 2 | 3 | 0 | 1 | 0 | 1 | 1 | 1 | 0 | 1 | 1 | 1 | 3 | 1 |
| 362 | 1021210 | 3 | 1 | 2 | 3 | 3 | 2 | 1 | 2 | 0 | 0 | 2 | 1 | 3 | 1 | 1 | 1 | 1 | 1 | 1 | 0 | 1 | 0 | 0 | 3 | 1 |
| 363 | 1021211 | 3 | 0 | 5 | 1 | 3 | 1 | 3 | 2 | 0 | 0 | 5 | 3 | 3 | 1 | 1 | 1 | 1 | 1 | 1 | 0 | 1 | 1 | 1 | 4 | 1 |
| 364 | 1021212 | 3 | 1 | 2 | 2 | 3 | 2 | 3 | 2 | 0 | 0 | 2 | 1 | 3 | 0 | 1 | 1 | 1 | 1 | 1 | 0 | 1 | 0 | 0 | 3 | 1 |
| 365 | 1021213 | 3 | 0 | 2 | 5 | 2 | 2 | 2 | 2 | 0 | 0 | 1 | 1 | 3 | 1 | 1 | 1 | 1 | 1 | 1 | 0 | 1 | 1 | 1 | 4 | 1 |
| 366 | 1021214 | 3 | 0 | 2 | 4 | 2 | 2 | 2 | 2 | 0 | 0 | 1 | 1 | 3 | 0 | 1 | 1 | 1 | 1 | 1 | 0 | 1 | 1 | 1 | 4 | 1 |
| 367 | 1021215 | 3 | 0 | 2 | 4 | 2 | 2 | 2 | 2 | 0 | 0 | 1 | 1 | 3 | 0 | 1 | 1 | 1 | 1 | 1 | 0 | 1 | 0 | 0 | 3 | 1 |
| 368 | 1021216 | 3 | 1 | 1 | 5 | 2 | 2 | 2 | 2 | 0 | 0 | 2 | 1 | 3 | 0 | 1 | 1 | 1 | 1 | 1 | 0 | 1 | 0 | 0 | 3 | 1 |
| 369 | 1021217 | 3 | 0 | 2 | 4 | 2 | 1 | 1 | 2 | 0 | 0 | 1 | 1 | 3 | 0 | 1 | 1 | 1 | 1 | 1 | 0 | 1 | 0 | 0 | 3 | 1 |
| 370 | 1021218 | 3 | 1 | 3 | 1 | 3 | 1 | 1 | 2 | 0 | 0 | 4 | 2 | 3 | 0 | 1 | 0 | 1 | 1 | 1 | 0 | 1 | 0 | 1 | 3 | 1 |
| 371 | 1021219 | 3 | 1 | 3 | 3 | 2 | 2 | 1 | 2 | 0 | 0 | 2 | 2 | 3 | 1 | 1 | 0 | 1 | 1 | 1 | 0 | 1 | 1 | 1 | 4 | 1 |
| 372 | 1021220 | 3 | 1 | 2 | 4 | 2 | 1 | 1 | 2 | 0 | 0 | 2 | 1 | 3 | 0 | 1 | 1 | 1 | 1 | 1 | 0 | 1 | 1 | 1 | 4 | 1 |
| 373 | 1021221 | 3 | 1 | 2 | 5 | 3 | 1 | 1 | 1 | 0 | 0 | 2 | 1 | 3 | 1 | 1 | 0 | 1 | 1 | 1 | 0 | 1 | 1 | 1 | 4 | 1 |
| 374 | 1021222 | 3 | 0 | 1 | 4 | 2 | 2 | 2 | 2 | 0 | 0 | 1 | 1 | 3 | 1 | 1 | 1 | 1 | 1 | 1 | 0 | 1 | 0 | 0 | 3 | 1 |
| 375 | 1021223 | 3 | 1 | 5 | 2 | 3 | 1 | 1 | 4 | 0 | 0 | 5 | 3 | 3 | 0 | 1 | 1 | 1 | 1 | 1 | 0 | 1 | 1 | 1 | 4 | 1 |

| | | | | | | | | | | | | | | | | | | | | | | | | | | |
|---|---|---|---|---|---|---|---|---|---|---|---|---|---|---|---|---|---|---|---|---|---|---|---|---|---|---|
| 376 | 1021224 | 3 | 0 | 5 | 4 | 3 | 2 | 1 | 4 | 1 | 1 | 5 | 3 | 3 | 0 | 1 | 1 | 1 | 1 | 1 | 0 | 1 | 1 | 1 | 4 | 1 |
| 377 | 1021225 | 3 | 0 | 2 | 5 | 2 | 2 | 1 | 2 | 0 | 0 | 1 | 1 | 3 | 0 | 1 | 1 | 1 | 1 | 1 | 0 | 1 | 1 | 1 | 4 | 1 |
| 378 | 1021226 | 3 | 0 | 1 | 4 | 2 | 1 | 2 | 2 | 0 | 0 | 1 | 1 | 3 | 0 | 1 | 0 | 0 | 1 | 1 | 0 | 0 | 0 | 0 | 2 | 0 |
| 379 | 1021227 | 3 | 1 | 1 | 2 | 2 | 1 | 2 | 2 | 0 | 0 | 4 | 1 | 3 | 1 | 1 | 1 | 1 | 1 | 1 | 0 | 1 | 1 | 1 | 4 | 1 |
| 380 | 1021228 | 3 | 1 | 1 | 5 | 2 | 2 | 1 | 2 | 0 | 0 | 2 | 1 | 3 | 0 | 1 | 0 | 1 | 1 | 1 | 0 | 1 | 0 | 0 | 2 | 0 |
| 381 | 1021229 | 3 | 0 | 2 | 2 | 2 | 2 | 1 | 4 | 0 | 0 | 1 | 1 | 4 | 0 | 1 | 1 | 1 | 1 | 1 | 0 | 1 | 1 | 0 | 3 | 1 |
| 382 | 1021230 | 3 | 1 | 2 | 2 | 2 | 2 | 1 | 2 | 0 | 0 | 2 | 1 | 3 | 0 | 1 | 0 | 1 | 1 | 1 | 0 | 1 | 0 | 0 | 2 | 0 |
| 383 | 1021231 | 3 | 1 | 1 | 3 | 2 | 2 | 1 | 2 | 0 | 0 | 4 | 1 | 3 | 0 | 1 | 1 | 1 | 1 | 1 | 0 | 1 | 0 | 1 | 3 | 1 |
| 384 | 1021232 | 3 | 1 | 1 | 4 | 2 | 1 | 1 | 2 | 0 | 0 | 4 | 1 | 2 | 0 | 1 | 1 | 1 | 1 | 1 | 0 | 1 | 1 | 1 | 4 | 1 |
| 385 | 1021233 | 3 | 1 | 5 | 1 | 3 | 1 | 1 | 2 | 1 | 1 | 3 | 3 | 3 | 0 | 1 | 1 | 1 | 1 | 1 | 0 | 1 | 1 | 1 | 4 | 1 |
| 386 | 1021234 | 3 | 0 | 1 | 5 | 2 | 2 | 2 | 4 | 0 | 0 | 1 | 1 | 2 | 1 | 1 | 1 | 1 | 1 | 1 | 0 | 1 | 0 | 0 | 3 | 1 |
| 387 | 1021235 | 3 | 0 | 5 | 2 | 2 | 2 | 2 | 4 | 0 | 0 | 5 | 3 | 3 | 1 | 1 | 1 | 1 | 1 | 1 | 0 | 1 | 1 | 1 | 4 | 1 |
| 388 | 103141 | 3 | 1 | 3 | 4 | 2 | 1 | 1 | 2 | 1 | 1 | 2 | 2 | 3 | 1 | 0 | 0 | 0 | 0 | 0 | 0 | 0 | 0 | 0 | 2 | 0 |
| 389 | 103142 | 3 | 1 | 3 | 2 | 2 | 2 | 1 | 2 | 0 | 0 | 2 | 2 | 3 | 0 | 0 | 0 | 0 | 0 | 0 | 0 | 0 | 0 | 0 | 1 | 0 |
| 390 | 103143 | 3 | 1 | 3 | 5 | 2 | 2 | 1 | 2 | 1 | 0 | 2 | 2 | 3 | 0 | 0 | 0 | 0 | 0 | 0 | 0 | 0 | 0 | 0 | 1 | 0 |
| 391 | 103144 | 3 | 1 | 3 | 3 | 2 | 2 | 1 | 2 | 0 | 0 | 2 | 2 | 3 | 1 | 1 | 0 | 1 | 0 | 1 | 0 | 1 | 0 | 0 | 2 | 0 |
| 392 | 103145 | 3 | 1 | 3 | 3 | 2 | 1 | 1 | 2 | 0 | 0 | 2 | 2 | 3 | 1 | 0 | 0 | 0 | 0 | 0 | 0 | 0 | 0 | 0 | 2 | 0 |
| 393 | 103146 | 3 | 1 | 1 | 5 | 2 | 1 | 1 | 2 | 0 | 0 | 2 | 1 | 3 | 1 | 0 | 0 | 0 | 0 | 0 | 0 | 0 | 0 | 0 | 2 | 0 |
| 394 | 103147 | 3 | 1 | 1 | 5 | 1 | 2 | 1 | 2 | 0 | 0 | 2 | 1 | 3 | 1 | 0 | 0 | 0 | 0 | 0 | 0 | 0 | 0 | 0 | 2 | 0 |
| 395 | 103148 | 3 | 1 | 3 | 4 | 2 | 2 | 1 | 2 | 0 | 0 | 2 | 2 | 3 | 1 | 0 | 0 | 0 | 0 | 0 | 0 | 0 | 0 | 0 | 2 | 0 |
| 396 | 103149 | 3 | 1 | 2 | 3 | 2 | 2 | 1 | 2 | 1 | 0 | 2 | 1 | 3 | 1 | 0 | 0 | 0 | 0 | 0 | 0 | 0 | 0 | 0 | 2 | 0 |
| 397 | 1031410 | 3 | 1 | 4 | 4 | 2 | 1 | 1 | 2 | 0 | 0 | 2 | 2 | 3 | 1 | 1 | 0 | 1 | 0 | 1 | 0 | 1 | 0 | 0 | 2 | 0 |
| 398 | 1031411 | 3 | 0 | 2 | 5 | 2 | 1 | 1 | 3 | 1 | 1 | 1 | 1 | 3 | 1 | 0 | 0 | 0 | 0 | 0 | 0 | 0 | 0 | 0 | 2 | 0 |
| 399 | 1031412 | 3 | 0 | 2 | 3 | 1 | 1 | 1 | 3 | 0 | 0 | 1 | 1 | 3 | 1 | 0 | 0 | 0 | 0 | 0 | 0 | 0 | 0 | 0 | 2 | 0 |
| 400 | 1031413 | 3 | 0 | 1 | 4 | 2 | 1 | 1 | 3 | 0 | 0 | 1 | 1 | 3 | 1 | 0 | 0 | 0 | 0 | 0 | 0 | 0 | 0 | 0 | 2 | 0 |
| 401 | 1031414 | 3 | 0 | 2 | 3 | 2 | 1 | 1 | 3 | 0 | 0 | 1 | 1 | 3 | 0 | 0 | 0 | 0 | 0 | 0 | 0 | 0 | 0 | 0 | 1 | 0 |
| 402 | 1031415 | 3 | 0 | 1 | 3 | 2 | 1 | 1 | 3 | 0 | 0 | 1 | 1 | 3 | 1 | 1 | 0 | 1 | 0 | 1 | 0 | 1 | 0 | 0 | 2 | 0 |
| 403 | 1031416 | 3 | 0 | 2 | 3 | 2 | 1 | 1 | 3 | 0 | 0 | 1 | 1 | 3 | 1 | 0 | 0 | 0 | 0 | 0 | 0 | 0 | 0 | 0 | 2 | 0 |
| 404 | 1031417 | 3 | 0 | 2 | 5 | 1 | 1 | 1 | 3 | 0 | 0 | 1 | 1 | 3 | 1 | 0 | 0 | 0 | 0 | 0 | 0 | 0 | 0 | 0 | 2 | 0 |
| 405 | 1031418 | 3 | 0 | 2 | 5 | 2 | 2 | 1 | 3 | 0 | 0 | 1 | 1 | 3 | 1 | 0 | 0 | 0 | 0 | 0 | 0 | 0 | 0 | 0 | 2 | 0 |
| 406 | 1031419 | 3 | 0 | 2 | 4 | 1 | 1 | 1 | 3 | 0 | 0 | 1 | 1 | 3 | 1 | 0 | 0 | 0 | 0 | 0 | 0 | 0 | 0 | 0 | 2 | 0 |
| 407 | 1031420 | 3 | 0 | 2 | 5 | 2 | 2 | 1 | 3 | 0 | 0 | 1 | 1 | 3 | 1 | 0 | 0 | 0 | 0 | 0 | 0 | 0 | 0 | 0 | 2 | 0 |
| 408 | 1031421 | 3 | 0 | 2 | 4 | 1 | 1 | 1 | 3 | 0 | 0 | 1 | 1 | 3 | 1 | 0 | 0 | 0 | 0 | 0 | 0 | 0 | 0 | 0 | 2 | 0 |
| 409 | 1031422 | 3 | 0 | 2 | 4 | 1 | 1 | 1 | 3 | 0 | 0 | 1 | 1 | 3 | 1 | 0 | 0 | 0 | 0 | 0 | 0 | 0 | 0 | 0 | 2 | 0 |
| 410 | 1031423 | 3 | 0 | 2 | 1 | 1 | 1 | 1 | 3 | 0 | 0 | 1 | 1 | 3 | 1 | 1 | 0 | 1 | 0 | 1 | 0 | 1 | 0 | 0 | 2 | 0 |
| 411 | 1031424 | 3 | 0 | 1 | 4 | 2 | 2 | 1 | 3 | 0 | 0 | 1 | 1 | 3 | 1 | 0 | 0 | 0 | 0 | 0 | 0 | 0 | 0 | 0 | 2 | 0 |
| 412 | 1031425 | 3 | 1 | 1 | 4 | 1 | 1 | 1 | 3 | 0 | 0 | 2 | 1 | 3 | 1 | 0 | 0 | 0 | 0 | 0 | 0 | 0 | 0 | 0 | 2 | 0 |
| 413 | 1031426 | 3 | 0 | 2 | 2 | 1 | 1 | 1 | 3 | 0 | 0 | 1 | 1 | 3 | 1 | 0 | 0 | 0 | 0 | 0 | 0 | 0 | 0 | 0 | 2 | 0 |
| 414 | 1031427 | 3 | 0 | 2 | 3 | 1 | 1 | 1 | 3 | 0 | 0 | 1 | 1 | 3 | 1 | 1 | 0 | 1 | 0 | 1 | 0 | 1 | 0 | 0 | 2 | 0 |
| 415 | 1031428 | 3 | 0 | 4 | 1 | 1 | 1 | 1 | 3 | 0 | 0 | 1 | 2 | 3 | 1 | 0 | 0 | 0 | 0 | 0 | 0 | 0 | 0 | 0 | 2 | 0 |
| 416 | 1031429 | 3 | 0 | 3 | 4 | 2 | 1 | 1 | 3 | 0 | 0 | 1 | 2 | 3 | 0 | 1 | 0 | 1 | 0 | 1 | 0 | 1 | 0 | 0 | 2 | 0 |
| 417 | 1031430 | 3 | 0 | 2 | 3 | 1 | 1 | 1 | 3 | 0 | 0 | 1 | 1 | 3 | 1 | 0 | 0 | 0 | 0 | 0 | 0 | 0 | 0 | 0 | 2 | 0 |
| 418 | 103131 | 3 | 0 | 2 | 3 | 2 | 1 | 1 | 2 | 0 | 0 | 1 | 1 | 3 | 1 | 1 | 0 | 1 | 0 | 1 | 0 | 1 | 0 | 0 | 2 | 0 |
| 419 | 103132 | 3 | 0 | 5 | 2 | 2 | 2 | 4 | 2 | 0 | 0 | 5 | 3 | 3 | 1 | 1 | 0 | 1 | 0 | 1 | 0 | 1 | 0 | 0 | 2 | 0 |
| 420 | 103133 | 3 | 0 | 2 | 2 | 2 | 2 | 1 | 2 | 0 | 0 | 1 | 1 | 3 | 0 | 1 | 0 | 1 | 0 | 0 | 0 | 0 | 0 | 0 | 2 | 0 |
| 421 | 103134 | 3 | 0 | 3 | 2 | 3 | 2 | 1 | 2 | 0 | 0 | 1 | 2 | 3 | 1 | 1 | 0 | 1 | 0 | 1 | 0 | 1 | 0 | 0 | 2 | 0 |
| 422 | 103135 | 3 | 0 | 5 | 2 | 3 | 2 | 1 | 2 | 0 | 0 | 5 | 3 | 3 | 1 | 1 | 0 | 1 | 0 | 1 | 0 | 1 | 0 | 0 | 2 | 0 |
| 423 | 103136 | 3 | 1 | 2 | 4 | 1 | 1 | 1 | 2 | 0 | 0 | 2 | 1 | 3 | 0 | 1 | 0 | 0 | 0 | 0 | 0 | 0 | 0 | 0 | 2 | 0 |
| 424 | 103137 | 3 | 0 | 2 | 3 | 1 | 2 | 1 | 2 | 0 | 0 | 1 | 1 | 3 | 0 | 1 | 0 | 1 | 0 | 1 | 0 | 1 | 0 | 0 | 2 | 0 |
| 425 | 103138 | 3 | 0 | 2 | 2 | 1 | 2 | 1 | 2 | 0 | 0 | 1 | 1 | 3 | 1 | 0 | 0 | 0 | 0 | 0 | 0 | 0 | 0 | 0 | 2 | 0 |
| 426 | 103139 | 3 | 0 | 4 | 3 | 1 | 1 | 1 | 2 | 0 | 0 | 1 | 2 | 3 | 1 | 1 | 0 | 1 | 0 | 1 | 0 | 1 | 0 | 0 | 2 | 0 |
| 427 | 1031310 | 3 | 1 | 2 | 5 | 1 | 1 | 1 | 2 | 0 | 0 | 2 | 1 | 3 | 0 | 1 | 0 | 0 | 0 | 0 | 0 | 0 | 0 | 0 | 2 | 0 |
| 428 | 1031311 | 3 | 0 | 2 | 4 | 2 | 1 | 1 | 2 | 0 | 0 | 1 | 1 | 3 | 0 | 1 | 0 | 1 | 0 | 1 | 0 | 1 | 0 | 0 | 2 | 0 |
| 429 | 1031312 | 3 | 1 | 2 | 4 | 1 | 1 | 1 | 2 | 0 | 0 | 5 | 1 | 3 | 0 | 0 | 0 | 0 | 0 | 0 | 0 | 0 | 0 | 0 | 1 | 0 |
| 430 | 1031313 | 3 | 1 | 4 | 3 | 2 | 2 | 1 | 2 | 0 | 0 | 2 | 2 | 3 | 0 | 1 | 0 | 1 | 0 | 1 | 1 | 0 | 0 | 0 | 2 | 0 |
| 431 | 1031314 | 3 | 1 | 4 | 2 | 2 | 2 | 1 | 2 | 0 | 0 | 2 | 2 | 3 | 0 | 1 | 0 | 1 | 1 | 1 | 1 | 0 | 0 | 0 | 2 | 0 |
| 432 | 1031315 | 3 | 0 | 2 | 4 | 1 | 1 | 1 | 2 | 0 | 0 | 1 | 1 | 3 | 0 | 1 | 0 | 0 | 0 | 0 | 0 | 0 | 0 | 0 | 2 | 0 |
| 433 | 1031316 | 3 | 0 | 2 | 4 | 1 | 1 | 1 | 2 | 0 | 0 | 1 | 1 | 3 | 0 | 1 | 0 | 1 | 0 | 1 | 1 | 0 | 0 | 0 | 2 | 0 |
| 434 | 1031317 | 3 | 1 | 2 | 2 | 2 | 2 | 1 | 2 | 0 | 0 | 2 | 1 | 3 | 1 | 0 | 0 | 0 | 0 | 0 | 0 | 0 | 0 | 0 | 2 | 0 |
| 435 | 1031318 | 3 | 0 | 2 | 3 | 1 | 1 | 1 | 2 | 0 | 0 | 2 | 1 | 3 | 0 | 1 | 0 | 0 | 0 | 0 | 0 | 0 | 0 | 0 | 2 | 0 |
| 436 | 1031319 | 3 | 0 | 4 | 1 | 3 | 1 | 1 | 2 | 0 | 0 | 5 | 2 | 3 | 0 | 1 | 0 | 1 | 0 | 0 | 0 | 0 | 0 | 1 | 2 | 0 |
| 437 | 1031320 | 3 | 0 | 4 | 1 | 2 | 1 | 1 | 2 | 0 | 0 | 1 | 2 | 3 | 1 | 1 | 0 | 1 | 0 | 0 | 0 | 0 | 0 | 1 | 2 | 0 |
| 438 | 1031321 | 3 | 0 | 2 | 5 | 1 | 1 | 1 | 2 | 0 | 0 | 1 | 1 | 3 | 0 | 0 | 0 | 0 | 0 | 0 | 0 | 0 | 0 | 0 | 1 | 0 |
| 439 | 1031322 | 3 | 0 | 2 | 5 | 2 | 1 | 1 | 2 | 0 | 0 | 1 | 1 | 3 | 0 | 0 | 0 | 0 | 0 | 0 | 0 | 0 | 0 | 0 | 1 | 0 |
| 440 | 1031323 | 3 | 0 | 2 | 2 | 2 | 2 | 1 | 2 | 0 | 0 | 1 | 1 | 3 | 0 | 0 | 0 | 0 | 0 | 0 | 0 | 0 | 0 | 0 | 1 | 0 |
| 441 | 1031324 | 3 | 0 | 2 | 3 | 2 | 1 | 1 | 2 | 0 | 0 | 1 | 1 | 3 | 0 | 1 | 0 | 1 | 0 | 1 | 0 | 1 | 0 | 0 | 2 | 0 |
| 442 | 1031325 | 3 | 0 | 2 | 4 | 2 | 1 | 1 | 2 | 0 | 0 | 1 | 1 | 3 | 0 | 1 | 0 | 1 | 0 | 1 | 0 | 1 | 0 | 0 | 2 | 0 |
| 443 | 1031326 | 3 | 0 | 2 | 2 | 2 | 2 | 1 | 2 | 0 | 0 | 1 | 1 | 3 | 1 | 1 | 0 | 0 | 0 | 0 | 0 | 0 | 0 | 0 | 2 | 0 |
| 444 | 1031327 | 3 | 0 | 2 | 3 | 2 | 2 | 1 | 2 | 0 | 0 | 1 | 1 | 3 | 0 | 1 | 0 | 0 | 0 | 0 | 0 | 0 | 0 | 0 | 2 | 0 |
| 445 | 1031328 | 3 | 0 | 2 | 5 | 1 | 1 | 1 | 2 | 0 | 0 | 1 | 1 | 3 | 0 | 0 | 0 | 0 | 0 | 0 | 0 | 0 | 0 | 0 | 1 | 0 |
| 446 | 1031329 | 3 | 0 | 2 | 3 | 1 | 2 | 1 | 2 | 0 | 0 | 1 | 1 | 3 | 0 | 1 | 0 | 1 | 0 | 0 | 0 | 0 | 0 | 1 | 2 | 0 |
| 447 | 1031330 | 3 | 0 | 2 | 3 | 2 | 1 | 1 | 2 | 0 | 0 | 1 | 1 | 3 | 0 | 0 | 0 | 0 | 0 | 0 | 0 | 0 | 0 | 0 | 1 | 0 |
| 448 | 103151 | 3 | 0 | 1 | 4 | 1 | 1 | 1 | 2 | 0 | 0 | 1 | 1 | 3 | 0 | 0 | 0 | 0 | 0 | 0 | 0 | 0 | 0 | 0 | 1 | 0 |
| 449 | 103152 | 3 | 0 | 2 | 4 | 2 | 1 | 1 | 2 | 0 | 0 | 1 | 1 | 3 | 0 | 1 | 0 | 1 | 0 | 0 | 0 | 0 | 0 | 0 | 2 | 0 |
| 450 | 103153 | 3 | 0 | 2 | 4 | 2 | 2 | 1 | 2 | 0 | 0 | 1 | 1 | 3 | 1 | 0 | 0 | 0 | 0 | 0 | 0 | 0 | 0 | 0 | 2 | 0 |
| 451 | 103154 | 3 | 0 | 2 | 1 | 2 | 2 | 1 | 2 | 0 | 0 | 1 | 1 | 3 | 0 | 0 | 0 | 0 | 0 | 0 | 0 | 0 | 0 | 0 | 1 | 0 |
| 452 | 103155 | 3 | 0 | 2 | 2 | 2 | 1 | 1 | 2 | 0 | 0 | 1 | 1 | 3 | 0 | 1 | 0 | 1 | 0 | 0 | 0 | 1 | 0 | 0 | 2 | 0 |
| 453 | 103156 | 3 | 1 | 2 | 4 | 2 | 1 | 1 | 2 | 0 | 0 | 2 | 1 | 3 | 1 | 0 | 0 | 0 | 0 | 0 | 0 | 0 | 0 | 0 | 2 | 0 |
| 454 | 103157 | 3 | 0 | 4 | 1 | 1 | 1 | 1 | 2 | 0 | 0 | 1 | 2 | 3 | 1 | 0 | 0 | 0 | 0 | 0 | 0 | 0 | 0 | 0 | 2 | 0 |
| 455 | 103158 | 3 | 0 | 2 | 4 | 1 | 1 | 1 | 2 | 0 | 0 | 1 | 1 | 3 | 1 | 0 | 0 | 0 | 0 | 0 | 0 | 0 | 0 | 0 | 2 | 0 |
| 456 | 103159 | 3 | 0 | 2 | 5 | 2 | 1 | 1 | 2 | 1 | 1 | 1 | 1 | 3 | 0 | 0 | 0 | 0 | 0 | 0 | 0 | 0 | 0 | 0 | 1 | 0 |
| 457 | 1031510 | 3 | 0 | 2 | 3 | 2 | 2 | 1 | 2 | 0 | 0 | 1 | 1 | 3 | 1 | 0 | 0 | 0 | 0 | 0 | 0 | 0 | 0 | 0 | 2 | 0 |
| 458 | 1031511 | 3 | 0 | 3 | 2 | 2 | 2 | 1 | 2 | 0 | 0 | 1 | 2 | 3 | 1 | 0 | 0 | 0 | 0 | 0 | 0 | 0 | 0 | 0 | 2 | 0 |
| 459 | 1031512 | 3 | 0 | 2 | 2 | 1 | 2 | 1 | 2 | 0 | 0 | 1 | 1 | 3 | 0 | 0 | 0 | 0 | 0 | 0 | 0 | 0 | 0 | 0 | 1 | 0 |
| 460 | 1031513 | 3 | 0 | 4 | 1 | 1 | 2 | 1 | 2 | 0 | 0 | 1 | 2 | 3 | 1 | 0 | 0 | 0 | 0 | 0 | 0 | 0 | 0 | 0 | 2 | 0 |
| 461 | 1031514 | 3 | 0 | 3 | 1 | 1 | 1 | 1 | 2 | 0 | 0 | 1 | 2 | 3 | 0 | 0 | 0 | 0 | 0 | 0 | 0 | 0 | 0 | 0 | 1 | 0 |
| 462 | 1031515 | 3 | 0 | 4 | 1 | 1 | 1 | 1 | 2 | 0 | 0 | 1 | 2 | 3 | 1 | 0 | 0 | 0 | 0 | 0 | 0 | 0 | 0 | 0 | 2 | 0 |
| 463 | 1031516 | 3 | 0 | 3 | 2 | 1 | 2 | 1 | 2 | 0 | 0 | 1 | 2 | 3 | 0 | 1 | 0 | 1 | 0 | 1 | 0 | 1 | 0 | 0 | 2 | 0 |
| 464 | 1031517 | 3 | 0 | 2 | 4 | 1 | 1 | 1 | 2 | 0 | 0 | 1 | 1 | 3 | 0 | 0 | 0 | 0 | 0 | 0 | 0 | 0 | 0 | 0 | 1 | 0 |
| 465 | 1031518 | 3 | 0 | 3 | 4 | 2 | 2 | 1 | 2 | 0 | 0 | 1 | 2 | 3 | 1 | 1 | 0 | 1 | 0 | 1 | 0 | 1 | 0 | 0 | 2 | 0 |
| 466 | 1031519 | 3 | 0 | 4 | 1 | 1 | 1 | 1 | 2 | 0 | 0 | 1 | 2 | 3 | 1 | 1 | 0 | 1 | 0 | 1 | 0 | 1 | 0 | 0 | 2 | 0 |
| 467 | 1031520 | 3 | 1 | 2 | 4 | 2 | 1 | 1 | 2 | 0 | 0 | 2 | 1 | 3 | 0 | 0 | 0 | 0 | 0 | 0 | 0 | 0 | 0 | 0 | 1 | 0 |
| 468 | 1031521 | 3 | 0 | 1 | 4 | 1 | 1 | 1 | 2 | 0 | 0 | 1 | 1 | 3 | 0 | 0 | 0 | 0 | 0 | 0 | 0 | 0 | 0 | 0 | 1 | 0 |
| 469 | 1031522 | 3 | 1 | 2 | 5 | 2 | 2 | 1 | 2 | 0 | 0 | 2 | 1 | 3 | 0 | 0 | 0 | 0 | 0 | 0 | 0 | 0 | 0 | 0 | 1 | 0 |

| | | | | | | | | | | | | | | | | | | | | | | | | | | |
|---|---|---|---|---|---|---|---|---|---|---|---|---|---|---|---|---|---|---|---|---|---|---|---|---|---|---|
| 470 | 1031523 | 3 | 1 | 2 | 5 | 3 | 2 | 1 | 2 | 1 | 0 | 2 | 1 | 3 | 1 | 1 | 0 | 1 | 0 | 1 | 0 | 1 | 0 | 0 | 2 | 0 |
| 471 | 1031524 | 3 | 0 | 1 | 5 | 2 | 1 | 1 | 2 | 0 | 0 | 1 | 1 | 3 | 0 | 0 | 0 | 0 | 0 | 0 | 0 | 0 | 0 | 0 | 1 | 0 |
| 472 | 1031525 | 3 | 1 | 1 | 4 | 2 | 1 | 1 | 2 | 0 | 0 | 2 | 1 | 3 | 0 | 0 | 0 | 0 | 0 | 0 | 0 | 0 | 0 | 0 | 1 | 0 |
| 473 | 1031526 | 3 | 0 | 1 | 5 | 1 | 1 | 1 | 2 | 0 | 0 | 1 | 1 | 3 | 0 | 0 | 0 | 0 | 0 | 0 | 0 | 0 | 0 | 0 | 1 | 0 |
| 474 | 1031527 | 3 | 0 | 2 | 4 | 2 | 1 | 1 | 2 | 0 | 0 | 1 | 1 | 3 | 0 | 0 | 0 | 0 | 0 | 0 | 0 | 0 | 0 | 0 | 1 | 0 |
| 475 | 1031528 | 3 | 0 | 3 | 1 | 2 | 2 | 1 | 2 | 0 | 0 | 1 | 2 | 3 | 1 | 1 | 0 | 1 | 0 | 1 | 0 | 1 | 0 | 0 | 2 | 0 |
| 476 | 1031529 | 3 | 0 | 2 | 3 | 2 | 1 | 1 | 2 | 0 | 0 | 1 | 1 | 3 | 0 | 1 | 0 | 1 | 0 | 1 | 0 | 1 | 0 | 0 | 2 | 0 |
| 477 | 1031530 | 3 | 0 | 5 | 2 | 3 | 1 | 1 | 2 | 0 | 0 | 5 | 3 | 3 | 1 | 1 | 0 | 1 | 0 | 1 | 0 | 1 | 0 | 0 | 2 | 0 |
| 478 | 103161 | 3 | 0 | 2 | 3 | 2 | 2 | 1 | 1 | 0 | 0 | 1 | 1 | 3 | 1 | 0 | 0 | 0 | 0 | 0 | 0 | 0 | 0 | 0 | 2 | 0 |
| 479 | 103162 | 3 | 0 | 3 | 3 | 2 | 2 | 1 | 1 | 0 | 0 | 1 | 2 | 3 | 1 | 0 | 0 | 0 | 0 | 0 | 0 | 0 | 0 | 0 | 2 | 0 |
| 480 | 103163 | 3 | 0 | 3 | 2 | 3 | 2 | 1 | 1 | 0 | 0 | 1 | 2 | 3 | 1 | 1 | 0 | 0 | 0 | 0 | 0 | 0 | 0 | 0 | 2 | 0 |
| 481 | 103164 | 3 | 0 | 3 | 2 | 2 | 1 | 1 | 1 | 0 | 0 | 1 | 2 | 3 | 1 | 1 | 0 | 0 | 0 | 0 | 0 | 0 | 0 | 0 | 2 | 0 |
| 482 | 103165 | 3 | 0 | 4 | 4 | 1 | 1 | 1 | 1 | 0 | 0 | 1 | 2 | 3 | 1 | 1 | 0 | 0 | 0 | 0 | 0 | 0 | 0 | 0 | 2 | 0 |
| 483 | 103166 | 3 | 0 | 2 | 4 | 1 | 1 | 1 | 1 | 0 | 0 | 1 | 1 | 3 | 1 | 1 | 0 | 0 | 0 | 0 | 0 | 0 | 0 | 0 | 2 | 0 |
| 484 | 103167 | 3 | 0 | 2 | 2 | 1 | 2 | 1 | 1 | 0 | 0 | 1 | 1 | 3 | 1 | 0 | 0 | 0 | 0 | 0 | 0 | 0 | 0 | 0 | 2 | 0 |
| 485 | 103168 | 3 | 0 | 2 | 5 | 2 | 1 | 1 | 1 | 0 | 0 | 1 | 1 | 3 | 1 | 1 | 0 | 1 | 0 | 1 | 0 | 1 | 0 | 0 | 2 | 0 |
| 486 | 103169 | 3 | 0 | 2 | 5 | 1 | 2 | 1 | 1 | 0 | 0 | 1 | 1 | 3 | 1 | 0 | 0 | 0 | 0 | 0 | 0 | 0 | 0 | 0 | 2 | 0 |
| 487 | 1031610 | 3 | 0 | 4 | 1 | 2 | 1 | 1 | 1 | 0 | 0 | 1 | 2 | 3 | 0 | 0 | 0 | 0 | 0 | 0 | 0 | 0 | 0 | 0 | 1 | 0 |
| 488 | 1031611 | 3 | 0 | 2 | 1 | 1 | 2 | 1 | 1 | 0 | 0 | 1 | 1 | 3 | 0 | 1 | 0 | 0 | 0 | 0 | 0 | 0 | 0 | 0 | 2 | 0 |
| 489 | 1031612 | 3 | 0 | 2 | 2 | 2 | 1 | 1 | 1 | 1 | 1 | 1 | 1 | 3 | 0 | 1 | 0 | 0 | 0 | 0 | 0 | 0 | 0 | 0 | 2 | 0 |
| 490 | 1031613 | 3 | 0 | 2 | 3 | 2 | 2 | 1 | 1 | 0 | 0 | 1 | 1 | 3 | 0 | 1 | 0 | 0 | 0 | 0 | 0 | 0 | 0 | 0 | 2 | 0 |
| 491 | 1031614 | 3 | 0 | 2 | 3 | 1 | 1 | 1 | 1 | 0 | 0 | 1 | 1 | 3 | 0 | 1 | 0 | 0 | 0 | 0 | 0 | 0 | 0 | 0 | 2 | 0 |
| 492 | 1031615 | 3 | 0 | 2 | 5 | 1 | 2 | 1 | 1 | 0 | 0 | 1 | 1 | 3 | 1 | 1 | 0 | 0 | 0 | 0 | 0 | 0 | 0 | 0 | 2 | 0 |
| 493 | 1031616 | 3 | 0 | 2 | 4 | 1 | 2 | 1 | 1 | 0 | 0 | 1 | 1 | 3 | 0 | 0 | 0 | 0 | 0 | 0 | 0 | 0 | 0 | 0 | 1 | 0 |
| 494 | 1031617 | 3 | 0 | 2 | 4 | 2 | 2 | 1 | 1 | 1 | 1 | 1 | 1 | 3 | 1 | 0 | 0 | 0 | 0 | 0 | 0 | 0 | 0 | 0 | 2 | 0 |
| 495 | 1031618 | 3 | 0 | 2 | 4 | 1 | 2 | 1 | 1 | 0 | 0 | 1 | 1 | 3 | 0 | 0 | 0 | 0 | 0 | 0 | 0 | 0 | 0 | 0 | 1 | 0 |
| 496 | 1031619 | 3 | 0 | 1 | 5 | 2 | 2 | 1 | 1 | 1 | 1 | 1 | 1 | 3 | 0 | 0 | 0 | 0 | 0 | 0 | 0 | 0 | 0 | 0 | 1 | 0 |
| 497 | 1031620 | 3 | 0 | 3 | 3 | 2 | 1 | 1 | 1 | 0 | 0 | 1 | 2 | 3 | 1 | 0 | 0 | 0 | 0 | 0 | 0 | 0 | 0 | 0 | 2 | 0 |
| 498 | 1031621 | 3 | 0 | 5 | 1 | 3 | 1 | 1 | 1 | 1 | 1 | 5 | 3 | 3 | 0 | 1 | 0 | 1 | 0 | 1 | 1 | 0 | 1 | 1 | 3 | 1 |
| 499 | 1031622 | 3 | 0 | 4 | 4 | 2 | 2 | 1 | 1 | 1 | 1 | 1 | 2 | 3 | 1 | 0 | 0 | 0 | 0 | 0 | 0 | 0 | 0 | 0 | 2 | 0 |
| 500 | 1031623 | 3 | 0 | 3 | 4 | 3 | 1 | 1 | 1 | 1 | 1 | 1 | 2 | 3 | 1 | 1 | 0 | 1 | 0 | 1 | 0 | 1 | 0 | 0 | 2 | 0 |
| 501 | 204171 | 1 | 0 | 4 | 1 | 2 | 2 | 4 | 4 | 0 | 0 | 1 | 2 | 1 | 1 | 1 | 1 | 1 | 0 | 1 | 0 | 1 | 1 | 0 | 3 | 1 |
| 502 | 204172 | 1 | 0 | 1 | 3 | 2 | 1 | 4 | 4 | 0 | 0 | 2 | 1 | 1 | 1 | 1 | 1 | 1 | 0 | 1 | 0 | 1 | 0 | 0 | 3 | 1 |
| 503 | 204173 | 1 | 0 | 2 | 1 | 2 | 1 | 4 | 4 | 0 | 0 | 1 | 1 | 1 | 0 | 1 | 1 | 1 | 0 | 1 | 0 | 0 | 0 | 1 | 2 | 0 |
| 504 | 204174 | 1 | 1 | 2 | 2 | 2 | 2 | 4 | 4 | 0 | 0 | 2 | 1 | 1 | 0 | 1 | 0 | 1 | 0 | 1 | 0 | 1 | 0 | 1 | 2 | 0 |
| 505 | 204175 | 1 | 0 | 4 | 1 | 2 | 1 | 4 | 4 | 0 | 0 | 2 | 2 | 1 | 0 | 1 | 1 | 1 | 1 | 1 | 0 | 1 | 0 | 0 | 3 | 1 |
| 506 | 204176 | 1 | 1 | 2 | 5 | 2 | 2 | 4 | 4 | 0 | 0 | 2 | 1 | 1 | 0 | 1 | 1 | 1 | 0 | 1 | 1 | 0 | 0 | 0 | 2 | 0 |
| 507 | 204177 | 1 | 1 | 2 | 4 | 1 | 2 | 1 | 4 | 0 | 0 | 2 | 1 | 1 | 1 | 1 | 0 | 1 | 1 | 1 | 0 | 1 | 0 | 0 | 3 | 1 |
| 508 | 204178 | 1 | 1 | 1 | 2 | 2 | 1 | 4 | 4 | 0 | 0 | 2 | 1 | 1 | 0 | 1 | 0 | 1 | 0 | 1 | 0 | 1 | 0 | 1 | 2 | 0 |
| 509 | 204179 | 1 | 0 | 2 | 1 | 2 | 2 | 2 | 4 | 0 | 0 | 1 | 1 | 1 | 0 | 1 | 0 | 1 | 1 | 1 | 0 | 1 | 0 | 0 | 2 | 0 |
| 510 | 2041710 | 1 | 0 | 5 | 1 | 2 | 2 | 1 | 4 | 0 | 0 | 1 | 3 | 1 | 1 | 1 | 0 | 1 | 1 | 1 | 0 | 0 | 1 | 1 | 3 | 1 |
| 511 | 2041711 | 1 | 1 | 2 | 4 | 2 | 2 | 4 | 4 | 0 | 0 | 2 | 1 | 1 | 0 | 1 | 0 | 1 | 0 | 1 | 0 | 1 | 1 | 0 | 2 | 0 |
| 512 | 2041712 | 1 | 1 | 2 | 4 | 2 | 2 | 4 | 4 | 0 | 0 | 4 | 1 | 1 | 0 | 1 | 1 | 1 | 0 | 1 | 0 | 1 | 0 | 1 | 3 | 1 |
| 513 | 2041713 | 1 | 0 | 2 | 4 | 2 | 1 | 4 | 4 | 0 | 0 | 1 | 1 | 1 | 1 | 1 | 1 | 1 | 1 | 1 | 0 | 1 | 1 | 0 | 4 | 1 |
| 514 | 2041714 | 1 | 1 | 2 | 5 | 1 | 1 | 3 | 4 | 0 | 0 | 2 | 1 | 1 | 0 | 1 | 1 | 1 | 1 | 0 | 0 | 1 | 0 | 1 | 3 | 1 |
| 515 | 2041715 | 1 | 1 | 2 | 5 | 2 | 1 | 1 | 4 | 0 | 0 | 2 | 1 | 1 | 0 | 1 | 0 | 1 | 0 | 0 | 0 | 1 | 0 | 0 | 2 | 0 |
| 516 | 2041716 | 1 | 1 | 2 | 5 | 2 | 2 | 1 | 4 | 0 | 0 | 2 | 1 | 1 | 0 | 1 | 0 | 1 | 0 | 0 | 0 | 1 | 0 | 0 | 2 | 0 |
| 517 | 2041717 | 1 | 1 | 3 | 4 | 3 | 2 | 1 | 4 | 1 | 1 | 5 | 2 | 1 | 1 | 1 | 1 | 1 | 1 | 1 | 0 | 1 | 1 | 1 | 4 | 1 |
| 518 | 2041718 | 1 | 0 | 1 | 4 | 2 | 2 | 2 | 4 | 0 | 0 | 1 | 1 | 1 | 0 | 1 | 0 | 1 | 0 | 1 | 0 | 1 | 1 | 0 | 2 | 0 |
| 519 | 2041719 | 1 | 0 | 2 | 3 | 2 | 2 | 1 | 4 | 0 | 0 | 1 | 1 | 1 | 0 | 1 | 0 | 1 | 1 | 0 | 0 | 1 | 1 | 0 | 2 | 0 |
| 520 | 2041720 | 1 | 1 | 2 | 5 | 2 | 2 | 1 | 4 | 0 | 0 | 2 | 1 | 1 | 0 | 1 | 1 | 1 | 0 | 1 | 0 | 0 | 1 | 1 | 3 | 1 |
| 521 | 2041721 | 1 | 1 | 1 | 4 | 2 | 2 | 1 | 4 | 0 | 0 | 2 | 1 | 1 | 0 | 1 | 0 | 1 | 0 | 1 | 0 | 1 | 0 | 0 | 2 | 0 |
| 522 | 2041722 | 1 | 1 | 1 | 2 | 2 | 2 | 1 | 4 | 0 | 0 | 2 | 1 | 1 | 0 | 1 | 0 | 1 | 0 | 1 | 0 | 1 | 1 | 0 | 2 | 0 |
| 523 | 2041723 | 1 | 0 | 3 | 2 | 2 | 2 | 1 | 4 | 0 | 0 | 1 | 2 | 1 | 1 | 1 | 0 | 1 | 0 | 1 | 0 | 0 | 0 | 0 | 2 | 0 |
| 524 | 2041724 | 1 | 0 | 1 | 5 | 2 | 1 | 1 | 4 | 0 | 0 | 1 | 1 | 1 | 0 | 1 | 1 | 1 | 1 | 1 | 0 | 1 | 0 | 0 | 3 | 1 |
| 525 | 2041725 | 1 | 1 | 1 | 5 | 2 | 2 | 2 | 4 | 0 | 0 | 2 | 1 | 1 | 1 | 1 | 1 | 1 | 0 | 0 | 0 | 1 | 0 | 0 | 2 | 0 |
| 526 | 2041726 | 1 | 1 | 2 | 2 | 2 | 1 | 1 | 4 | 0 | 0 | 2 | 1 | 1 | 0 | 1 | 0 | 1 | 0 | 0 | 0 | 1 | 0 | 0 | 2 | 0 |
| 527 | 2041727 | 1 | 1 | 1 | 5 | 2 | 2 | 2 | 4 | 0 | 0 | 2 | 1 | 1 | 0 | 1 | 0 | 1 | 0 | 1 | 0 | 1 | 1 | 0 | 2 | 0 |
| 528 | 2041728 | 1 | 1 | 2 | 3 | 3 | 2 | 1 | 4 | 0 | 0 | 2 | 1 | 1 | 1 | 1 | 1 | 1 | 1 | 1 | 0 | 1 | 0 | 1 | 4 | 1 |
| 529 | 2041729 | 1 | 1 | 2 | 2 | 2 | 2 | 3 | 4 | 0 | 0 | 2 | 1 | 1 | 0 | 1 | 0 | 1 | 0 | 1 | 0 | 1 | 0 | 0 | 2 | 0 |
| 530 | 2041730 | 1 | 0 | 2 | 4 | 1 | 1 | 3 | 4 | 0 | 0 | 2 | 1 | 1 | 0 | 1 | 0 | 1 | 1 | 1 | 0 | 1 | 0 | 0 | 2 | 0 |
| 531 | 2041731 | 1 | 1 | 2 | 1 | 2 | 1 | 3 | 4 | 0 | 0 | 2 | 1 | 1 | 1 | 1 | 1 | 1 | 0 | 1 | 0 | 1 | 0 | 0 | 3 | 1 |
| 532 | 2041732 | 1 | 1 | 5 | 2 | 2 | 1 | 4 | 4 | 0 | 0 | 2 | 3 | 1 | 0 | 1 | 1 | 1 | 1 | 0 | 0 | 1 | 0 | 0 | 2 | 0 |
| 533 | 2041733 | 1 | 1 | 3 | 2 | 2 | 1 | 4 | 4 | 0 | 0 | 2 | 2 | 1 | 0 | 1 | 1 | 1 | 1 | 0 | 0 | 1 | 0 | 1 | 3 | 1 |
| 534 | 2041734 | 1 | 1 | 2 | 4 | 1 | 1 | 4 | 4 | 0 | 0 | 2 | 1 | 1 | 0 | 1 | 1 | 1 | 1 | 0 | 0 | 1 | 0 | 1 | 3 | 1 |
| 535 | 2041735 | 1 | 0 | 2 | 3 | 2 | 2 | 1 | 4 | 0 | 0 | 1 | 1 | 1 | 0 | 1 | 0 | 1 | 1 | 1 | 0 | 0 | 0 | 1 | 2 | 0 |
| 536 | 204181 | 1 | 1 | 2 | 3 | 3 | 2 | 2 | 1 | 0 | 0 | 2 | 1 | 1 | 0 | 1 | 1 | 1 | 0 | 1 | 0 | 1 | 0 | 1 | 3 | 1 |
| 537 | 204182 | 1 | 1 | 2 | 4 | 1 | 2 | 1 | 1 | 0 | 0 | 2 | 1 | 1 | 0 | 1 | 1 | 1 | 0 | 0 | 0 | 0 | 0 | 0 | 2 | 0 |
| 538 | 204183 | 1 | 1 | 2 | 3 | 2 | 2 | 3 | 1 | 0 | 0 | 2 | 1 | 1 | 0 | 1 | 1 | 1 | 0 | 1 | 0 | 1 | 1 | 0 | 3 | 1 |
| 539 | 204184 | 1 | 1 | 2 | 4 | 2 | 1 | 2 | 1 | 0 | 0 | 2 | 1 | 1 | 0 | 1 | 0 | 1 | 0 | 1 | 0 | 1 | 1 | 0 | 2 | 0 |
| 540 | 204185 | 1 | 0 | 1 | 5 | 2 | 1 | 1 | 1 | 0 | 0 | 2 | 1 | 1 | 0 | 1 | 0 | 1 | 0 | 0 | 0 | 1 | 0 | 0 | 2 | 0 |
| 541 | 204186 | 1 | 0 | 1 | 5 | 2 | 1 | 1 | 2 | 0 | 0 | 2 | 1 | 1 | 0 | 1 | 0 | 1 | 0 | 1 | 0 | 1 | 0 | 0 | 2 | 0 |
| 542 | 204187 | 1 | 1 | 5 | 2 | 2 | 2 | 2 | 4 | 0 | 0 | 2 | 3 | 1 | 1 | 1 | 1 | 1 | 1 | 1 | 0 | 1 | 1 | 0 | 4 | 1 |
| 543 | 204188 | 1 | 1 | 4 | 1 | 2 | 1 | 2 | 1 | 0 | 0 | 2 | 2 | 1 | 0 | 1 | 1 | 1 | 0 | 0 | 0 | 1 | 0 | 0 | 2 | 0 |
| 544 | 204189 | 1 | 1 | 2 | 4 | 2 | 1 | 1 | 1 | 0 | 0 | 2 | 1 | 1 | 0 | 1 | 1 | 1 | 0 | 0 | 0 | 1 | 0 | 0 | 2 | 0 |
| 545 | 2041810 | 1 | 1 | 3 | 5 | 1 | 2 | 2 | 1 | 0 | 0 | 2 | 2 | 1 | 1 | 1 | 1 | 1 | 0 | 1 | 0 | 1 | 1 | 0 | 3 | 1 |
| 546 | 2041811 | 1 | 1 | 4 | 4 | 3 | 2 | 2 | 4 | 0 | 0 | 2 | 2 | 1 | 0 | 1 | 0 | 1 | 0 | 1 | 0 | 1 | 0 | 1 | 2 | 0 |
| 547 | 2041812 | 1 | 0 | 2 | 3 | 3 | 1 | 1 | 1 | 0 | 0 | 2 | 1 | 1 | 1 | 1 | 0 | 1 | 1 | 0 | 1 | 1 | 0 | 0 | 3 | 1 |
| 548 | 2041813 | 1 | 0 | 2 | 3 | 2 | 1 | 2 | 4 | 0 | 0 | 2 | 1 | 1 | 0 | 1 | 1 | 1 | 0 | 1 | 0 | 1 | 0 | 0 | 2 | 0 |
| 549 | 2041814 | 1 | 1 | 4 | 3 | 2 | 1 | 4 | 3 | 0 | 0 | 2 | 2 | 1 | 1 | 1 | 1 | 1 | 1 | 1 | 1 | 1 | 0 | 0 | 4 | 1 |
| 550 | 2041815 | 1 | 1 | 5 | 1 | 1 | 1 | 4 | 4 | 0 | 0 | 2 | 3 | 1 | 0 | 1 | 1 | 1 | 1 | 1 | 1 | 1 | 1 | 0 | 4 | 1 |
| 551 | 2041816 | 1 | 1 | 2 | 3 | 2 | 1 | 4 | 4 | 0 | 0 | 2 | 1 | 1 | 0 | 1 | 0 | 1 | 0 | 1 | 0 | 0 | 0 | 1 | 2 | 0 |
| 552 | 2041817 | 1 | 1 | 2 | 4 | 2 | 2 | 4 | 4 | 0 | 0 | 2 | 1 | 1 | 1 | 1 | 0 | 1 | 0 | 0 | 0 | 0 | 0 | 0 | 2 | 0 |
| 553 | 2041818 | 1 | 1 | 2 | 5 | 2 | 2 | 1 | 1 | 0 | 0 | 2 | 1 | 1 | 0 | 1 | 0 | 1 | 0 | 0 | 0 | 0 | 0 | 1 | 2 | 0 |
| 554 | 2041819 | 1 | 1 | 5 | 2 | 2 | 1 | 4 | 4 | 1 | 1 | 5 | 3 | 1 | 1 | 1 | 1 | 1 | 0 | 0 | 0 | 0 | 0 | 1 | 2 | 0 |
| 555 | 2041820 | 1 | 0 | 1 | 4 | 3 | 2 | 1 | 1 | 0 | 0 | 2 | 1 | 1 | 0 | 1 | 0 | 1 | 0 | 0 | 0 | 0 | 0 | 1 | 2 | 0 |
| 556 | 2041821 | 1 | 1 | 3 | 3 | 2 | 2 | 2 | 4 | 0 | 0 | 2 | 2 | 1 | 0 | 1 | 1 | 1 | 0 | 1 | 0 | 1 | 0 | 1 | 3 | 1 |
| 557 | 2041822 | 1 | 1 | 1 | 4 | 2 | 1 | 4 | 4 | 0 | 0 | 2 | 1 | 1 | 0 | 1 | 1 | 1 | 1 | 1 | 1 | 0 | 1 | 0 | 3 | 1 |
| 558 | 2041823 | 1 | 1 | 1 | 5 | 2 | 2 | 1 | 1 | 0 | 0 | 2 | 1 | 1 | 0 | 1 | 0 | 1 | 0 | 1 | 0 | 1 | 0 | 0 | 2 | 0 |
| 559 | 2041824 | 1 | 1 | 4 | 1 | 2 | 1 | 1 | 1 | 0 | 0 | 2 | 2 | 1 | 1 | 1 | 1 | 1 | 1 | 1 | 0 | 1 | 1 | 1 | 4 | 1 |
| 560 | 2041825 | 1 | 0 | 3 | 1 | 2 | 1 | 1 | 1 | 0 | 0 | 1 | 2 | 1 | 0 | 1 | 1 | 1 | 0 | 1 | 0 | 1 | 0 | 0 | 2 | 0 |
| 561 | 2041826 | 1 | 1 | 2 | 3 | 2 | 2 | 1 | 1 | 0 | 0 | 2 | 1 | 1 | 0 | 1 | 0 | 1 | 0 | 1 | 0 | 1 | 0 | 1 | 2 | 0 |
| 562 | 2041827 | 1 | 1 | 2 | 5 | 2 | 1 | 1 | 1 | 0 | 0 | 2 | 1 | 1 | 0 | 1 | 0 | 0 | 0 | 1 | 0 | 0 | 0 | 0 | 2 | 0 |
| 563 | 2041828 | 1 | 1 | 1 | 5 | 2 | 2 | 4 | 4 | 0 | 0 | 2 | 1 | 1 | 0 | 1 | 0 | 1 | 0 | 1 | 0 | 1 | 0 | 0 | 2 | 0 |

| | | | | | | | | | | | | | | | | | | | | | | | | | | |
|---|---|---|---|---|---|---|---|---|---|---|---|---|---|---|---|---|---|---|---|---|---|---|---|---|---|---|
| 564 | 2041829 | 1 | 1 | 4 | 1 | 2 | 1 | 4 | 4 | 0 | 0 | 2 | 2 | 1 | 0 | 1 | 1 | 1 | 1 | 1 | 0 | 1 | 0 | 1 | 3 | 1 |
| 565 | 2041830 | 1 | 1 | 2 | 4 | 2 | 2 | 1 | 1 | 0 | 0 | 1 | 1 | 1 | 0 | 1 | 0 | 1 | 0 | 0 | 0 | 1 | 0 | 0 | 2 | 0 |
| 566 | 204191 | 1 | 1 | 4 | 2 | 2 | 2 | 4 | 3 | 0 | 0 | 2 | 2 | 1 | 1 | 1 | 1 | 1 | 0 | 1 | 0 | 0 | 1 | 0 | 3 | 1 |
| 567 | 204192 | 1 | 1 | 5 | 1 | 2 | 1 | 2 | 1 | 0 | 0 | 2 | 3 | 1 | 0 | 1 | 1 | 1 | 0 | 1 | 0 | 0 | 1 | 1 | 3 | 1 |
| 568 | 204193 | 1 | 0 | 2 | 2 | 1 | 1 | 4 | 3 | 0 | 0 | 2 | 1 | 1 | 0 | 1 | 1 | 1 | 0 | 0 | 0 | 0 | 1 | 1 | 2 | 0 |
| 569 | 204194 | 1 | 1 | 1 | 5 | 1 | 1 | 4 | 3 | 0 | 0 | 2 | 1 | 1 | 0 | 1 | 1 | 1 | 0 | 1 | 0 | 0 | 0 | 0 | 2 | 0 |
| 570 | 204195 | 1 | 0 | 2 | 3 | 2 | 2 | 3 | 1 | 0 | 0 | 2 | 1 | 1 | 1 | 1 | 1 | 1 | 0 | 1 | 0 | 0 | 1 | 0 | 3 | 1 |
| 571 | 204196 | 1 | 0 | 2 | 4 | 1 | 1 | 2 | 1 | 0 | 0 | 2 | 1 | 1 | 0 | 1 | 1 | 1 | 0 | 1 | 0 | 0 | 0 | 0 | 2 | 0 |
| 572 | 204197 | 1 | 0 | 1 | 5 | 1 | 1 | 2 | 1 | 0 | 0 | 2 | 1 | 1 | 0 | 1 | 0 | 1 | 0 | 1 | 0 | 0 | 1 | 0 | 2 | 0 |
| 573 | 204198 | 1 | 0 | 4 | 3 | 2 | 1 | 2 | 1 | 0 | 0 | 1 | 2 | 1 | 0 | 1 | 1 | 1 | 0 | 1 | 0 | 0 | 1 | 1 | 3 | 1 |
| 574 | 204199 | 1 | 0 | 1 | 5 | 1 | 1 | 4 | 1 | 0 | 0 | 2 | 1 | 1 | 0 | 0 | 0 | 0 | 0 | 0 | 0 | 0 | 0 | 0 | 1 | 0 |
| 575 | 2041910 | 1 | 0 | 2 | 3 | 1 | 2 | 4 | 3 | 0 | 0 | 2 | 1 | 1 | 0 | 1 | 1 | 1 | 0 | 1 | 0 | 0 | 0 | 1 | 2 | 0 |
| 576 | 2041911 | 1 | 0 | 2 | 3 | 2 | 2 | 2 | 1 | 0 | 0 | 2 | 1 | 1 | 0 | 1 | 1 | 1 | 0 | 0 | 0 | 0 | 0 | 0 | 2 | 0 |
| 577 | 2041912 | 1 | 1 | 3 | 2 | 2 | 1 | 4 | 3 | 0 | 0 | 2 | 2 | 1 | 0 | 1 | 1 | 1 | 0 | 1 | 0 | 0 | 1 | 1 | 3 | 1 |
| 578 | 2041913 | 1 | 1 | 1 | 5 | 1 | 1 | 4 | 3 | 0 | 0 | 2 | 1 | 1 | 0 | 1 | 1 | 1 | 0 | 1 | 0 | 0 | 1 | 0 | 2 | 0 |
| 579 | 2041914 | 1 | 1 | 1 | 5 | 1 | 1 | 4 | 3 | 0 | 0 | 2 | 1 | 1 | 0 | 1 | 1 | 1 | 1 | 1 | 0 | 0 | 1 | 0 | 3 | 1 |
| 580 | 2041915 | 1 | 0 | 2 | 3 | 1 | 1 | 2 | 3 | 0 | 0 | 2 | 1 | 1 | 0 | 1 | 1 | 1 | 0 | 1 | 0 | 0 | 1 | 0 | 2 | 0 |
| 581 | 2041916 | 1 | 1 | 2 | 2 | 2 | 1 | 4 | 3 | 0 | 0 | 2 | 1 | 1 | 0 | 1 | 1 | 1 | 0 | 1 | 0 | 0 | 1 | 1 | 3 | 1 |
| 582 | 2041917 | 1 | 1 | 2 | 2 | 1 | 2 | 4 | 3 | 0 | 0 | 2 | 1 | 1 | 0 | 1 | 1 | 1 | 0 | 1 | 0 | 0 | 0 | 1 | 2 | 0 |
| 583 | 2041918 | 1 | 1 | 2 | 3 | 2 | 2 | 3 | 3 | 0 | 0 | 2 | 1 | 1 | 0 | 1 | 0 | 1 | 0 | 1 | 0 | 0 | 1 | 0 | 2 | 0 |
| 584 | 2041919 | 1 | 1 | 2 | 3 | 1 | 1 | 4 | 3 | 0 | 0 | 2 | 1 | 1 | 0 | 1 | 1 | 1 | 0 | 1 | 0 | 0 | 1 | 1 | 3 | 1 |
| 585 | 2041920 | 1 | 0 | 1 | 5 | 1 | 1 | 4 | 3 | 0 | 0 | 2 | 1 | 1 | 0 | 1 | 1 | 1 | 0 | 1 | 0 | 0 | 0 | 0 | 2 | 0 |
| 586 | 2041921 | 1 | 0 | 2 | 3 | 2 | 2 | 4 | 3 | 0 | 0 | 2 | 1 | 1 | 1 | 1 | 1 | 1 | 0 | 1 | 0 | 0 | 1 | 1 | 3 | 1 |
| 587 | 2041922 | 1 | 1 | 2 | 4 | 1 | 1 | 3 | 3 | 0 | 0 | 2 | 1 | 1 | 0 | 1 | 0 | 1 | 0 | 1 | 0 | 0 | 0 | 1 | 2 | 0 |
| 588 | 2041923 | 1 | 1 | 1 | 5 | 1 | 1 | 4 | 3 | 0 | 0 | 2 | 1 | 1 | 0 | 1 | 0 | 1 | 0 | 1 | 0 | 0 | 0 | 1 | 2 | 0 |
| 589 | 2041924 | 1 | 1 | 2 | 2 | 2 | 2 | 4 | 3 | 0 | 0 | 2 | 1 | 1 | 0 | 1 | 1 | 1 | 0 | 1 | 0 | 0 | 0 | 1 | 2 | 0 |
| 590 | 2041925 | 1 | 1 | 2 | 2 | 1 | 1 | 4 | 3 | 0 | 0 | 2 | 1 | 1 | 1 | 1 | 0 | 1 | 0 | 1 | 0 | 0 | 0 | 0 | 2 | 0 |
| 591 | 2041926 | 1 | 0 | 3 | 2 | 1 | 1 | 4 | 3 | 0 | 0 | 2 | 2 | 1 | 1 | 1 | 1 | 1 | 0 | 1 | 0 | 0 | 1 | 1 | 3 | 1 |
| 592 | 2041927 | 1 | 1 | 1 | 4 | 2 | 2 | 4 | 3 | 0 | 0 | 2 | 1 | 1 | 0 | 1 | 1 | 1 | 0 | 1 | 0 | 1 | 1 | 0 | 3 | 1 |
| 593 | 2041928 | 1 | 0 | 1 | 4 | 1 | 1 | 4 | 3 | 0 | 0 | 2 | 1 | 1 | 1 | 1 | 0 | 1 | 0 | 1 | 0 | 0 | 0 | 1 | 2 | 0 |
| 594 | 2041929 | 1 | 0 | 4 | 1 | 1 | 1 | 4 | 3 | 0 | 0 | 2 | 2 | 1 | 0 | 1 | 1 | 1 | 1 | 1 | 0 | 0 | 1 | 1 | 3 | 1 |
| 595 | 2041930 | 1 | 1 | 2 | 2 | 1 | 1 | 4 | 1 | 0 | 0 | 2 | 1 | 1 | 0 | 1 | 1 | 1 | 0 | 1 | 0 | 0 | 1 | 1 | 3 | 1 |
| 596 | 204201 | 1 | 1 | 4 | 2 | 2 | 1 | 4 | 1 | 0 | 0 | 2 | 2 | 1 | 0 | 1 | 1 | 1 | 0 | 1 | 0 | 0 | 1 | 1 | 3 | 1 |
| 597 | 204202 | 1 | 0 | 4 | 1 | 2 | 1 | 4 | 1 | 0 | 0 | 2 | 2 | 1 | 0 | 1 | 1 | 1 | 0 | 1 | 0 | 0 | 1 | 0 | 2 | 0 |
| 598 | 204203 | 1 | 1 | 5 | 3 | 2 | 1 | 4 | 1 | 0 | 0 | 5 | 3 | 1 | 0 | 1 | 1 | 1 | 0 | 1 | 0 | 0 | 1 | 1 | 3 | 1 |
| 599 | 204204 | 1 | 1 | 2 | 2 | 1 | 1 | 4 | 1 | 0 | 0 | 2 | 1 | 1 | 0 | 1 | 1 | 1 | 0 | 1 | 0 | 0 | 1 | 1 | 3 | 1 |
| 600 | 204205 | 1 | 1 | 2 | 3 | 2 | 2 | 2 | 1 | 0 | 0 | 2 | 1 | 1 | 0 | 1 | 1 | 1 | 0 | 1 | 0 | 0 | 0 | 0 | 2 | 0 |
| 601 | 204206 | 1 | 1 | 1 | 5 | 1 | 1 | 2 | 1 | 0 | 0 | 2 | 1 | 1 | 0 | 0 | 0 | 0 | 0 | 0 | 0 | 0 | 0 | 0 | 1 | 0 |
| 602 | 204207 | 1 | 1 | 2 | 3 | 1 | 2 | 1 | 1 | 0 | 0 | 2 | 1 | 1 | 0 | 1 | 1 | 1 | 0 | 1 | 0 | 0 | 0 | 0 | 2 | 0 |
| 603 | 204208 | 1 | 1 | 3 | 1 | 2 | 1 | 2 | 1 | 0 | 0 | 2 | 2 | 1 | 0 | 1 | 1 | 1 | 0 | 1 | 0 | 0 | 0 | 0 | 2 | 0 |
| 604 | 204209 | 1 | 0 | 2 | 3 | 1 | 1 | 2 | 1 | 0 | 0 | 1 | 1 | 1 | 0 | 1 | 1 | 1 | 0 | 0 | 0 | 0 | 0 | 0 | 2 | 0 |
| 605 | 2042010 | 1 | 0 | 2 | 3 | 1 | 2 | 2 | 1 | 0 | 0 | 3 | 1 | 1 | 0 | 1 | 1 | 1 | 1 | 1 | 0 | 0 | 1 | 1 | 3 | 1 |
| 606 | 2042011 | 1 | 1 | 1 | 5 | 1 | 1 | 3 | 1 | 0 | 0 | 2 | 1 | 1 | 0 | 1 | 1 | 1 | 0 | 1 | 0 | 0 | 0 | 0 | 2 | 0 |
| 607 | 2042012 | 1 | 0 | 1 | 5 | 1 | 1 | 3 | 1 | 0 | 0 | 2 | 1 | 1 | 0 | 0 | 0 | 0 | 0 | 0 | 0 | 0 | 0 | 0 | 1 | 0 |
| 608 | 2042013 | 1 | 1 | 2 | 3 | 2 | 2 | 4 | 1 | 0 | 0 | 2 | 1 | 1 | 0 | 1 | 1 | 1 | 0 | 1 | 0 | 0 | 1 | 0 | 2 | 0 |
| 609 | 2042014 | 1 | 0 | 1 | 4 | 1 | 2 | 4 | 1 | 0 | 0 | 2 | 1 | 1 | 0 | 1 | 1 | 1 | 0 | 1 | 0 | 0 | 0 | 1 | 2 | 0 |
| 610 | 2042015 | 1 | 1 | 2 | 2 | 2 | 1 | 2 | 1 | 0 | 0 | 2 | 1 | 1 | 0 | 1 | 1 | 1 | 0 | 1 | 0 | 0 | 0 | 1 | 2 | 0 |
| 611 | 2042016 | 1 | 0 | 2 | 2 | 1 | 1 | 2 | 1 | 0 | 0 | 1 | 1 | 1 | 0 | 1 | 1 | 1 | 0 | 1 | 0 | 0 | 1 | 1 | 3 | 1 |
| 612 | 2042017 | 1 | 0 | 2 | 2 | 3 | 2 | 1 | 1 | 0 | 0 | 2 | 1 | 1 | 0 | 1 | 1 | 1 | 0 | 0 | 0 | 0 | 1 | 0 | 2 | 0 |
| 613 | 2042018 | 1 | 1 | 2 | 4 | 2 | 2 | 4 | 4 | 0 | 0 | 2 | 1 | 1 | 0 | 1 | 1 | 1 | 0 | 1 | 0 | 0 | 1 | 1 | 3 | 1 |
| 614 | 2042019 | 1 | 1 | 2 | 3 | 2 | 2 | 4 | 1 | 0 | 0 | 2 | 1 | 1 | 0 | 1 | 1 | 1 | 1 | 1 | 0 | 0 | 1 | 1 | 3 | 1 |
| 615 | 2042020 | 1 | 0 | 1 | 5 | 1 | 1 | 4 | 1 | 0 | 0 | 2 | 1 | 1 | 0 | 1 | 1 | 1 | 0 | 1 | 0 | 0 | 1 | 0 | 2 | 0 |
| 616 | 2042021 | 1 | 1 | 2 | 3 | 2 | 1 | 4 | 1 | 0 | 0 | 2 | 1 | 1 | 0 | 1 | 1 | 1 | 0 | 1 | 1 | 0 | 1 | 1 | 3 | 1 |
| 617 | 2042022 | 1 | 1 | 1 | 5 | 1 | 1 | 4 | 1 | 0 | 0 | 2 | 1 | 1 | 0 | 1 | 0 | 1 | 0 | 1 | 0 | 0 | 1 | 1 | 2 | 0 |
| 618 | 2042023 | 1 | 1 | 2 | 2 | 2 | 2 | 4 | 1 | 0 | 0 | 2 | 1 | 1 | 1 | 1 | 1 | 1 | 0 | 1 | 0 | 0 | 1 | 1 | 3 | 1 |
| 619 | 2042024 | 1 | 1 | 4 | 1 | 2 | 1 | 4 | 1 | 0 | 0 | 2 | 2 | 1 | 0 | 1 | 1 | 1 | 0 | 1 | 0 | 0 | 0 | 0 | 2 | 0 |
| 620 | 2042025 | 1 | 1 | 2 | 3 | 1 | 2 | 4 | 1 | 0 | 0 | 2 | 1 | 1 | 0 | 1 | 0 | 1 | 0 | 1 | 0 | 0 | 1 | 1 | 2 | 0 |
| 621 | 205211 | 1 | 0 | 2 | 3 | 1 | 2 | 1 | 3 | 0 | 0 | 2 | 1 | 1 | 0 | 1 | 1 | 1 | 0 | 0 | 0 | 1 | 0 | 0 | 2 | 0 |
| 622 | 205212 | 1 | 1 | 2 | 3 | 3 | 2 | 1 | 3 | 0 | 0 | 2 | 1 | 1 | 1 | 1 | 1 | 1 | 0 | 1 | 0 | 1 | 0 | 0 | 3 | 1 |
| 623 | 205213 | 1 | 0 | 2 | 5 | 1 | 2 | 1 | 3 | 0 | 0 | 2 | 1 | 1 | 0 | 1 | 0 | 1 | 0 | 0 | 0 | 1 | 0 | 0 | 2 | 0 |
| 624 | 205214 | 1 | 0 | 2 | 3 | 1 | 1 | 2 | 3 | 0 | 0 | 2 | 1 | 1 | 0 | 1 | 1 | 1 | 0 | 1 | 0 | 1 | 0 | 0 | 2 | 0 |
| 625 | 205215 | 1 | 1 | 4 | 4 | 1 | 1 | 3 | 3 | 0 | 0 | 2 | 2 | 1 | 0 | 1 | 1 | 1 | 0 | 0 | 0 | 1 | 0 | 0 | 2 | 0 |
| 626 | 205216 | 1 | 0 | 1 | 4 | 1 | 1 | 3 | 3 | 0 | 0 | 2 | 1 | 1 | 0 | 1 | 1 | 1 | 0 | 0 | 0 | 1 | 0 | 0 | 2 | 0 |
| 627 | 205217 | 1 | 0 | 2 | 2 | 1 | 2 | 3 | 3 | 0 | 0 | 2 | 1 | 1 | 0 | 1 | 1 | 1 | 0 | 1 | 0 | 1 | 0 | 0 | 2 | 0 |
| 628 | 205218 | 1 | 1 | 4 | 2 | 1 | 2 | 3 | 3 | 0 | 0 | 2 | 2 | 1 | 1 | 1 | 1 | 1 | 1 | 1 | 0 | 1 | 0 | 0 | 3 | 1 |
| 629 | 205219 | 1 | 1 | 2 | 5 | 1 | 2 | 4 | 3 | 0 | 0 | 2 | 1 | 1 | 1 | 1 | 1 | 1 | 0 | 0 | 0 | 1 | 0 | 0 | 2 | 0 |
| 630 | 2052110 | 1 | 0 | 2 | 3 | 2 | 2 | 2 | 3 | 0 | 0 | 2 | 1 | 1 | 1 | 1 | 1 | 1 | 0 | 0 | 0 | 1 | 0 | 0 | 2 | 0 |
| 631 | 2052111 | 1 | 0 | 2 | 4 | 1 | 1 | 1 | 3 | 0 | 0 | 2 | 1 | 1 | 0 | 1 | 1 | 1 | 0 | 0 | 0 | 1 | 0 | 0 | 2 | 0 |
| 632 | 2052112 | 1 | 1 | 2 | 3 | 2 | 1 | 3 | 3 | 0 | 0 | 2 | 1 | 1 | 0 | 1 | 1 | 1 | 0 | 1 | 0 | 1 | 0 | 0 | 2 | 0 |
| 633 | 2052113 | 1 | 0 | 1 | 3 | 2 | 1 | 2 | 3 | 0 | 0 | 2 | 1 | 1 | 0 | 1 | 1 | 1 | 0 | 1 | 0 | 1 | 0 | 0 | 2 | 0 |
| 634 | 2052114 | 1 | 0 | 2 | 2 | 1 | 2 | 2 | 3 | 0 | 0 | 2 | 1 | 1 | 1 | 1 | 1 | 1 | 0 | 0 | 0 | 1 | 0 | 0 | 2 | 0 |
| 635 | 2052115 | 1 | 0 | 5 | 1 | 2 | 1 | 1 | 3 | 0 | 0 | 5 | 3 | 1 | 1 | 1 | 0 | 1 | 0 | 1 | 0 | 1 | 0 | 0 | 2 | 0 |
| 636 | 2052116 | 1 | 1 | 2 | 3 | 1 | 2 | 4 | 3 | 0 | 0 | 2 | 1 | 1 | 0 | 1 | 0 | 1 | 0 | 1 | 0 | 1 | 0 | 0 | 2 | 0 |
| 637 | 2052117 | 1 | 1 | 2 | 2 | 2 | 1 | 1 | 3 | 0 | 0 | 2 | 1 | 1 | 1 | 1 | 1 | 1 | 0 | 0 | 0 | 1 | 0 | 0 | 2 | 0 |
| 638 | 2052118 | 1 | 0 | 2 | 3 | 1 | 2 | 1 | 3 | 0 | 0 | 2 | 1 | 1 | 0 | 1 | 1 | 1 | 0 | 0 | 0 | 1 | 0 | 0 | 2 | 0 |
| 639 | 2052119 | 1 | 0 | 2 | 3 | 1 | 2 | 1 | 3 | 0 | 0 | 2 | 1 | 1 | 1 | 1 | 1 | 1 | 0 | 0 | 0 | 1 | 0 | 0 | 2 | 0 |
| 640 | 2052120 | 1 | 1 | 1 | 5 | 1 | 1 | 1 | 3 | 0 | 0 | 2 | 1 | 1 | 0 | 1 | 1 | 1 | 1 | 0 | 0 | 1 | 0 | 0 | 2 | 0 |
| 641 | 2052121 | 1 | 0 | 2 | 3 | 2 | 2 | 1 | 3 | 0 | 0 | 2 | 1 | 1 | 0 | 1 | 1 | 1 | 0 | 0 | 0 | 1 | 0 | 0 | 2 | 0 |
| 642 | 2052122 | 1 | 1 | 2 | 3 | 1 | 2 | 1 | 1 | 0 | 0 | 2 | 1 | 1 | 0 | 0 | 0 | 0 | 0 | 0 | 0 | 0 | 0 | 0 | 1 | 0 |
| 643 | 2052123 | 1 | 0 | 2 | 3 | 2 | 2 | 1 | 3 | 0 | 0 | 2 | 1 | 1 | 1 | 1 | 1 | 1 | 0 | 0 | 0 | 1 | 0 | 0 | 2 | 0 |
| 644 | 2052124 | 1 | 0 | 2 | 3 | 1 | 2 | 1 | 3 | 0 | 0 | 2 | 1 | 1 | 0 | 1 | 1 | 1 | 0 | 0 | 0 | 1 | 0 | 0 | 2 | 0 |
| 645 | 2052125 | 1 | 1 | 2 | 2 | 1 | 2 | 4 | 1 | 0 | 0 | 2 | 1 | 1 | 0 | 1 | 1 | 1 | 0 | 0 | 0 | 1 | 0 | 0 | 2 | 0 |
| 646 | 2052126 | 1 | 1 | 4 | 2 | 2 | 2 | 2 | 3 | 0 | 0 | 2 | 2 | 1 | 1 | 1 | 1 | 1 | 0 | 1 | 0 | 1 | 0 | 0 | 3 | 1 |
| 647 | 2052127 | 1 | 0 | 2 | 3 | 1 | 2 | 2 | 3 | 0 | 0 | 2 | 1 | 1 | 0 | 1 | 1 | 1 | 0 | 0 | 0 | 1 | 0 | 0 | 2 | 0 |
| 648 | 2052128 | 1 | 0 | 3 | 3 | 2 | 2 | 2 | 3 | 0 | 0 | 2 | 2 | 2 | 1 | 1 | 1 | 1 | 0 | 0 | 0 | 1 | 0 | 0 | 2 | 0 |
| 649 | 2052129 | 1 | 1 | 2 | 4 | 2 | 1 | 1 | 2 | 0 | 0 | 2 | 1 | 1 | 0 | 1 | 1 | 1 | 0 | 0 | 0 | 1 | 0 | 0 | 2 | 0 |
| 650 | 2052130 | 1 | 0 | 1 | 5 | 1 | 1 | 3 | 3 | 0 | 0 | 2 | 1 | 1 | 0 | 0 | 0 | 0 | 0 | 0 | 0 | 0 | 0 | 0 | 1 | 0 |
| 651 | 205221 | 1 | 0 | 1 | 2 | 2 | 2 | 4 | 4 | 0 | 0 | 2 | 1 | 1 | 1 | 1 | 1 | 1 | 0 | 1 | 0 | 1 | 0 | 0 | 3 | 1 |
| 652 | 205222 | 1 | 1 | 1 | 3 | 2 | 2 | 4 | 4 | 0 | 0 | 2 | 1 | 1 | 0 | 1 | 1 | 1 | 0 | 1 | 0 | 1 | 0 | 0 | 2 | 0 |
| 653 | 205223 | 1 | 1 | 1 | 1 | 1 | 2 | 4 | 4 | 0 | 0 | 2 | 1 | 1 | 0 | 1 | 1 | 1 | 0 | 1 | 0 | 1 | 0 | 0 | 2 | 0 |
| 654 | 205224 | 1 | 0 | 1 | 1 | 2 | 1 | 4 | 4 | 0 | 0 | 3 | 1 | 2 | 0 | 1 | 1 | 1 | 0 | 1 | 0 | 1 | 0 | 0 | 2 | 0 |
| 655 | 205225 | 1 | 1 | 3 | 2 | 1 | 2 | 4 | 4 | 0 | 0 | 2 | 2 | 1 | 0 | 1 | 1 | 1 | 0 | 1 | 0 | 1 | 0 | 0 | 2 | 0 |
| 656 | 205226 | 1 | 1 | 2 | 3 | 1 | 1 | 4 | 4 | 0 | 0 | 2 | 1 | 1 | 0 | 1 | 1 | 1 | 0 | 1 | 0 | 1 | 0 | 0 | 2 | 0 |
| 657 | 205227 | 1 | 0 | 1 | 4 | 2 | 2 | 4 | 4 | 0 | 0 | 2 | 1 | 1 | 0 | 1 | 1 | 1 | 0 | 1 | 1 | 0 | 0 | 0 | 2 | 0 |

| | | | | | | | | | | | | | | | | | | | | | | | | | | |
|---|---|---|---|---|---|---|---|---|---|---|---|---|---|---|---|---|---|---|---|---|---|---|---|---|---|---|
| 658 | 205228 | 1 | 0 | 1 | 4 | 1 | 2 | 4 | 3 | 0 | 0 | 2 | 1 | 1 | 0 | 1 | 1 | 1 | 1 | 1 | 0 | 1 | 0 | 0 | 3 | 1 |
| 659 | 205229 | 1 | 0 | 4 | 1 | 2 | 2 | 2 | 3 | 0 | 0 | 2 | 2 | 1 | 0 | 1 | 1 | 1 | 0 | 1 | 0 | 1 | 0 | 0 | 2 | 0 |
| 660 | 2052210 | 1 | 1 | 1 | 5 | 2 | 2 | 1 | 3 | 0 | 0 | 4 | 1 | 1 | 0 | 1 | 1 | 1 | 0 | 1 | 0 | 1 | 0 | 0 | 2 | 0 |
| 661 | 2052211 | 1 | 0 | 2 | 3 | 2 | 2 | 1 | 3 | 0 | 0 | 2 | 1 | 1 | 1 | 1 | 1 | 1 | 0 | 1 | 0 | 1 | 0 | 0 | 3 | 1 |
| 662 | 2052212 | 1 | 0 | 4 | 1 | 2 | 1 | 1 | 3 | 0 | 0 | 2 | 2 | 1 | 1 | 1 | 1 | 1 | 0 | 1 | 0 | 1 | 0 | 0 | 3 | 1 |
| 663 | 2052213 | 1 | 0 | 3 | 4 | 2 | 2 | 2 | 3 | 0 | 0 | 1 | 2 | 2 | 0 | 1 | 1 | 1 | 0 | 1 | 0 | 1 | 0 | 0 | 2 | 0 |
| 664 | 2052214 | 1 | 0 | 2 | 4 | 2 | 2 | 2 | 3 | 0 | 0 | 1 | 1 | 2 | 0 | 1 | 1 | 1 | 0 | 1 | 0 | 1 | 0 | 0 | 2 | 0 |
| 665 | 2052215 | 1 | 0 | 3 | 3 | 2 | 2 | 1 | 3 | 0 | 0 | 2 | 2 | 1 | 1 | 1 | 1 | 1 | 0 | 1 | 0 | 1 | 0 | 0 | 3 | 1 |
| 666 | 2052216 | 1 | 0 | 1 | 2 | 1 | 2 | 2 | 3 | 0 | 0 | 2 | 1 | 1 | 1 | 1 | 1 | 1 | 0 | 1 | 0 | 1 | 0 | 0 | 3 | 1 |
| 667 | 2052217 | 1 | 0 | 2 | 2 | 1 | 2 | 3 | 3 | 0 | 0 | 2 | 1 | 1 | 1 | 1 | 1 | 1 | 0 | 1 | 0 | 1 | 0 | 0 | 3 | 1 |
| 668 | 2052218 | 1 | 0 | 2 | 1 | 1 | 1 | 4 | 3 | 0 | 0 | 2 | 1 | 1 | 0 | 1 | 1 | 1 | 0 | 1 | 0 | 1 | 0 | 0 | 2 | 0 |
| 669 | 2052219 | 1 | 0 | 1 | 5 | 1 | 2 | 4 | 3 | 0 | 0 | 2 | 1 | 1 | 0 | 1 | 1 | 1 | 0 | 1 | 0 | 1 | 0 | 0 | 2 | 0 |
| 670 | 2052220 | 1 | 0 | 2 | 2 | 2 | 2 | 4 | 3 | 0 | 0 | 2 | 1 | 1 | 0 | 1 | 1 | 1 | 0 | 1 | 0 | 1 | 0 | 0 | 2 | 0 |
| 671 | 2052221 | 1 | 0 | 2 | 3 | 2 | 2 | 4 | 3 | 0 | 0 | 2 | 1 | 1 | 0 | 1 | 1 | 1 | 0 | 1 | 0 | 1 | 0 | 0 | 2 | 0 |
| 672 | 2052222 | 1 | 0 | 1 | 2 | 3 | 2 | 4 | 2 | 0 | 0 | 3 | 1 | 1 | 0 | 1 | 1 | 1 | 0 | 1 | 0 | 1 | 0 | 0 | 2 | 0 |
| 673 | 2052223 | 1 | 0 | 1 | 2 | 1 | 2 | 4 | 3 | 0 | 0 | 2 | 1 | 1 | 0 | 1 | 0 | 1 | 0 | 1 | 1 | 1 | 0 | 0 | 2 | 0 |
| 674 | 2052224 | 1 | 1 | 2 | 5 | 2 | 1 | 4 | 3 | 0 | 0 | 2 | 1 | 1 | 0 | 1 | 1 | 1 | 0 | 1 | 0 | 1 | 0 | 0 | 2 | 0 |
| 675 | 2052225 | 1 | 0 | 1 | 3 | 2 | 1 | 1 | 3 | 0 | 0 | 2 | 1 | 1 | 0 | 1 | 1 | 1 | 0 | 1 | 0 | 1 | 0 | 0 | 2 | 0 |
| 676 | 2052226 | 1 | 0 | 1 | 4 | 2 | 2 | 1 | 3 | 0 | 0 | 1 | 1 | 1 | 1 | 1 | 1 | 1 | 0 | 1 | 0 | 1 | 0 | 0 | 3 | 1 |
| 677 | 2052227 | 1 | 0 | 1 | 2 | 1 | 2 | 1 | 3 | 0 | 0 | 1 | 1 | 1 | 1 | 1 | 0 | 1 | 0 | 0 | 0 | 1 | 0 | 0 | 2 | 0 |
| 678 | 2052228 | 1 | 0 | 1 | 1 | 1 | 2 | 2 | 4 | 0 | 0 | 2 | 1 | 1 | 0 | 1 | 1 | 1 | 0 | 0 | 0 | 1 | 0 | 0 | 2 | 0 |
| 679 | 2052229 | 1 | 0 | 3 | 5 | 2 | 1 | 2 | 4 | 0 | 0 | 2 | 2 | 2 | 1 | 1 | 1 | 1 | 0 | 1 | 0 | 1 | 0 | 0 | 3 | 1 |
| 680 | 2052230 | 1 | 0 | 1 | 2 | 1 | 2 | 1 | 3 | 0 | 0 | 2 | 1 | 1 | 0 | 1 | 1 | 1 | 0 | 1 | 0 | 1 | 0 | 0 | 2 | 0 |
| 681 | 205231 | 1 | 0 | 5 | 1 | 2 | 1 | 1 | 3 | 0 | 0 | 5 | 3 | 1 | 0 | 1 | 1 | 1 | 0 | 1 | 0 | 1 | 0 | 0 | 2 | 0 |
| 682 | 205232 | 1 | 0 | 1 | 3 | 2 | 2 | 1 | 3 | 0 | 0 | 1 | 1 | 1 | 0 | 1 | 1 | 1 | 0 | 1 | 0 | 1 | 0 | 0 | 2 | 0 |
| 683 | 205233 | 1 | 0 | 4 | 1 | 2 | 2 | 1 | 3 | 0 | 0 | 2 | 2 | 1 | 1 | 1 | 1 | 1 | 0 | 1 | 0 | 1 | 0 | 0 | 3 | 1 |
| 684 | 205234 | 1 | 0 | 4 | 3 | 2 | 1 | 1 | 3 | 0 | 0 | 2 | 2 | 1 | 0 | 1 | 1 | 1 | 0 | 1 | 0 | 1 | 0 | 0 | 2 | 0 |
| 685 | 205235 | 1 | 1 | 3 | 2 | 3 | 1 | 1 | 3 | 1 | 0 | 3 | 2 | 1 | 0 | 1 | 1 | 1 | 0 | 1 | 0 | 0 | 1 | 0 | 2 | 0 |
| 686 | 205236 | 1 | 0 | 2 | 3 | 2 | 1 | 1 | 3 | 0 | 0 | 2 | 1 | 1 | 0 | 1 | 1 | 1 | 0 | 1 | 0 | 1 | 0 | 0 | 2 | 0 |
| 687 | 205237 | 1 | 0 | 2 | 2 | 2 | 2 | 2 | 4 | 0 | 0 | 3 | 1 | 1 | 1 | 1 | 1 | 1 | 0 | 1 | 0 | 1 | 0 | 0 | 3 | 1 |
| 688 | 205238 | 1 | 1 | 1 | 3 | 2 | 2 | 3 | 4 | 0 | 0 | 2 | 1 | 1 | 0 | 1 | 1 | 1 | 0 | 1 | 0 | 1 | 0 | 0 | 2 | 0 |
| 689 | 205239 | 1 | 0 | 3 | 1 | 1 | 2 | 4 | 4 | 0 | 0 | 1 | 2 | 2 | 0 | 1 | 1 | 1 | 0 | 0 | 0 | 1 | 0 | 0 | 2 | 0 |
| 690 | 2052310 | 1 | 1 | 1 | 3 | 2 | 2 | 4 | 4 | 0 | 0 | 2 | 1 | 1 | 0 | 1 | 1 | 1 | 0 | 1 | 0 | 1 | 0 | 0 | 2 | 0 |
| 691 | 2052311 | 1 | 1 | 1 | 2 | 1 | 2 | 2 | 4 | 0 | 0 | 4 | 1 | 1 | 0 | 1 | 1 | 1 | 0 | 1 | 0 | 1 | 0 | 0 | 2 | 0 |
| 692 | 2052312 | 1 | 1 | 1 | 3 | 2 | 2 | 3 | 4 | 0 | 0 | 4 | 1 | 1 | 0 | 1 | 1 | 1 | 0 | 1 | 0 | 1 | 0 | 0 | 2 | 0 |
| 693 | 2052313 | 1 | 0 | 2 | 3 | 2 | 1 | 4 | 4 | 0 | 0 | 2 | 1 | 1 | 0 | 1 | 1 | 1 | 0 | 1 | 0 | 1 | 0 | 0 | 2 | 0 |
| 694 | 2052314 | 1 | 0 | 5 | 3 | 2 | 2 | 4 | 4 | 0 | 0 | 5 | 3 | 1 | 1 | 1 | 1 | 1 | 0 | 1 | 0 | 1 | 0 | 0 | 3 | 1 |
| 695 | 2052315 | 1 | 0 | 1 | 2 | 2 | 2 | 2 | 4 | 0 | 0 | 2 | 1 | 1 | 0 | 1 | 1 | 1 | 0 | 1 | 0 | 1 | 0 | 0 | 2 | 0 |
| 696 | 2052316 | 1 | 1 | 1 | 5 | 2 | 2 | 3 | 4 | 0 | 0 | 2 | 1 | 1 | 0 | 1 | 1 | 1 | 0 | 1 | 0 | 1 | 0 | 0 | 2 | 0 |
| 697 | 2052317 | 1 | 0 | 1 | 1 | 2 | 1 | 4 | 4 | 0 | 0 | 2 | 1 | 1 | 0 | 1 | 0 | 1 | 1 | 1 | 0 | 1 | 0 | 0 | 2 | 0 |
| 698 | 2052318 | 1 | 1 | 4 | 5 | 3 | 2 | 4 | 4 | 1 | 1 | 5 | 2 | 1 | 1 | 1 | 1 | 1 | 0 | 1 | 0 | 1 | 0 | 0 | 3 | 1 |
| 699 | 2052319 | 1 | 1 | 5 | 5 | 3 | 2 | 1 | 4 | 0 | 0 | 5 | 3 | 1 | 1 | 1 | 1 | 1 | 0 | 1 | 0 | 1 | 0 | 0 | 3 | 1 |
| 700 | 2052320 | 1 | 0 | 2 | 2 | 3 | 2 | 1 | 3 | 1 | 1 | 3 | 1 | 1 | 0 | 1 | 1 | 1 | 0 | 1 | 0 | 1 | 0 | 0 | 2 | 0 |
| 701 | 2052321 | 1 | 1 | 4 | 5 | 2 | 2 | 1 | 3 | 0 | 0 | 2 | 2 | 1 | 0 | 1 | 1 | 1 | 0 | 1 | 0 | 1 | 0 | 0 | 2 | 0 |
| 702 | 2052322 | 1 | 0 | 1 | 4 | 2 | 2 | 1 | 3 | 0 | 0 | 2 | 1 | 1 | 0 | 1 | 1 | 1 | 0 | 1 | 0 | 1 | 0 | 0 | 2 | 0 |
| 703 | 2052323 | 1 | 1 | 1 | 5 | 2 | 2 | 1 | 3 | 0 | 0 | 2 | 1 | 1 | 0 | 1 | 1 | 1 | 0 | 1 | 1 | 1 | 0 | 0 | 3 | 1 |
| 704 | 2052324 | 1 | 0 | 1 | 1 | 1 | 2 | 1 | 3 | 0 | 0 | 2 | 1 | 1 | 0 | 1 | 1 | 1 | 0 | 1 | 0 | 1 | 0 | 0 | 2 | 0 |
| 705 | 2052325 | 1 | 1 | 1 | 3 | 2 | 2 | 1 | 3 | 0 | 0 | 2 | 1 | 1 | 0 | 1 | 1 | 1 | 0 | 1 | 0 | 1 | 0 | 0 | 2 | 0 |
| 706 | 2052326 | 1 | 0 | 2 | 1 | 2 | 2 | 1 | 3 | 0 | 0 | 2 | 1 | 1 | 0 | 1 | 1 | 1 | 0 | 1 | 0 | 1 | 0 | 0 | 2 | 0 |
| 707 | 2052327 | 1 | 1 | 1 | 5 | 1 | 1 | 1 | 3 | 0 | 0 | 2 | 1 | 1 | 0 | 1 | 1 | 1 | 0 | 1 | 0 | 1 | 0 | 0 | 2 | 0 |
| 708 | 2052328 | 1 | 1 | 2 | 3 | 2 | 2 | 1 | 3 | 0 | 0 | 2 | 1 | 1 | 1 | 1 | 1 | 1 | 0 | 1 | 0 | 1 | 0 | 0 | 3 | 1 |
| 709 | 2052329 | 1 | 1 | 1 | 1 | 2 | 1 | 1 | 4 | 0 | 0 | 2 | 1 | 1 | 0 | 1 | 1 | 1 | 0 | 1 | 0 | 1 | 0 | 0 | 2 | 0 |
| 710 | 2052330 | 1 | 0 | 1 | 1 | 1 | 2 | 1 | 4 | 0 | 0 | 1 | 1 | 1 | 1 | 1 | 1 | 0 | 0 | 0 | 0 | 0 | 0 | 0 | 2 | 0 |
| 711 | 205241 | 1 | 0 | 4 | 1 | 2 | 1 | 4 | 4 | 0 | 0 | 1 | 2 | 2 | 1 | 1 | 1 | 1 | 0 | 1 | 0 | 0 | 0 | 0 | 2 | 0 |
| 712 | 205242 | 1 | 0 | 2 | 3 | 3 | 2 | 3 | 4 | 0 | 0 | 2 | 1 | 1 | 1 | 1 | 1 | 1 | 0 | 1 | 0 | 0 | 0 | 0 | 2 | 0 |
| 713 | 205243 | 1 | 0 | 4 | 3 | 3 | 1 | 4 | 4 | 1 | 1 | 1 | 2 | 2 | 1 | 1 | 1 | 1 | 0 | 1 | 0 | 0 | 0 | 1 | 3 | 1 |
| 714 | 205244 | 1 | 0 | 3 | 3 | 3 | 2 | 3 | 4 | 1 | 1 | 1 | 2 | 2 | 1 | 1 | 1 | 1 | 0 | 1 | 1 | 0 | 0 | 0 | 3 | 1 |
| 715 | 205245 | 1 | 1 | 4 | 3 | 2 | 2 | 4 | 4 | 1 | 1 | 2 | 2 | 2 | 1 | 1 | 1 | 1 | 0 | 1 | 0 | 1 | 1 | 0 | 3 | 1 |
| 716 | 205246 | 1 | 0 | 5 | 4 | 3 | 2 | 4 | 4 | 1 | 1 | 5 | 3 | 2 | 1 | 1 | 1 | 1 | 0 | 1 | 0 | 1 | 0 | 1 | 3 | 1 |
| 717 | 205247 | 1 | 1 | 3 | 4 | 3 | 2 | 4 | 4 | 0 | 0 | 2 | 2 | 1 | 1 | 1 | 1 | 1 | 1 | 1 | 0 | 1 | 0 | 1 | 4 | 1 |
| 718 | 205248 | 1 | 1 | 2 | 3 | 2 | 2 | 4 | 4 | 0 | 0 | 4 | 1 | 1 | 0 | 1 | 1 | 1 | 0 | 1 | 0 | 1 | 0 | 0 | 2 | 0 |
| 719 | 205249 | 1 | 0 | 2 | 2 | 2 | 2 | 4 | 4 | 0 | 0 | 1 | 1 | 2 | 0 | 1 | 1 | 1 | 1 | 1 | 0 | 1 | 0 | 0 | 3 | 1 |
| 720 | 2052410 | 1 | 0 | 2 | 2 | 3 | 2 | 4 | 4 | 0 | 0 | 1 | 1 | 1 | 0 | 1 | 1 | 0 | 0 | 1 | 0 | 0 | 0 | 0 | 2 | 0 |
| 721 | 2052411 | 1 | 1 | 1 | 2 | 3 | 1 | 4 | 4 | 1 | 0 | 4 | 1 | 1 | 1 | 1 | 1 | 1 | 1 | 1 | 0 | 1 | 0 | 0 | 3 | 1 |
| 722 | 2052412 | 1 | 0 | 3 | 4 | 3 | 1 | 4 | 4 | 1 | 1 | 3 | 2 | 2 | 1 | 1 | 1 | 1 | 1 | 1 | 0 | 1 | 0 | 0 | 3 | 1 |
| 723 | 2052413 | 1 | 1 | 3 | 5 | 2 | 2 | 4 | 4 | 0 | 0 | 4 | 2 | 1 | 1 | 1 | 1 | 1 | 1 | 1 | 0 | 0 | 0 | 1 | 3 | 1 |
| 724 | 2052414 | 1 | 1 | 2 | 4 | 3 | 2 | 4 | 4 | 0 | 0 | 2 | 1 | 2 | 0 | 1 | 1 | 1 | 0 | 1 | 0 | 1 | 0 | 0 | 2 | 0 |
| 725 | 2052415 | 1 | 0 | 4 | 3 | 3 | 2 | 2 | 4 | 1 | 1 | 5 | 2 | 1 | 1 | 1 | 1 | 1 | 1 | 1 | 0 | 1 | 0 | 0 | 3 | 1 |
| 726 | 2052416 | 1 | 1 | 1 | 4 | 2 | 2 | 3 | 4 | 0 | 0 | 2 | 1 | 1 | 1 | 1 | 1 | 1 | 1 | 1 | 0 | 0 | 0 | 0 | 3 | 1 |
| 727 | 2052417 | 1 | 0 | 5 | 3 | 3 | 1 | 3 | 4 | 0 | 0 | 5 | 3 | 2 | 1 | 1 | 1 | 1 | 1 | 1 | 0 | 1 | 0 | 1 | 4 | 1 |
| 728 | 2052418 | 1 | 1 | 1 | 3 | 2 | 2 | 4 | 4 | 0 | 0 | 5 | 1 | 1 | 0 | 1 | 1 | 0 | 0 | 1 | 0 | 0 | 0 | 0 | 2 | 0 |
| 729 | 2052419 | 1 | 0 | 1 | 1 | 2 | 2 | 2 | 4 | 0 | 0 | 1 | 1 | 1 | 0 | 1 | 1 | 1 | 0 | 1 | 0 | 1 | 0 | 0 | 2 | 0 |
| 730 | 2052420 | 1 | 0 | 1 | 2 | 3 | 2 | 3 | 4 | 1 | 1 | 1 | 1 | 1 | 0 | 1 | 1 | 1 | 0 | 1 | 0 | 1 | 0 | 0 | 2 | 0 |
| 731 | 2052421 | 1 | 1 | 1 | 5 | 2 | 2 | 4 | 4 | 0 | 0 | 2 | 1 | 1 | 0 | 1 | 1 | 1 | 0 | 1 | 0 | 0 | 0 | 0 | 2 | 0 |
| 732 | 2052422 | 1 | 1 | 3 | 1 | 3 | 2 | 4 | 4 | 0 | 0 | 5 | 2 | 2 | 1 | 1 | 1 | 1 | 0 | 1 | 0 | 0 | 1 | 0 | 3 | 1 |
| 733 | 2052423 | 1 | 0 | 4 | 1 | 2 | 1 | 4 | 4 | 0 | 0 | 1 | 2 | 2 | 0 | 1 | 1 | 1 | 1 | 1 | 0 | 1 | 0 | 0 | 3 | 1 |
| 734 | 2052424 | 1 | 0 | 2 | 2 | 2 | 1 | 4 | 4 | 0 | 0 | 1 | 1 | 2 | 1 | 1 | 1 | 1 | 0 | 1 | 0 | 1 | 0 | 0 | 3 | 1 |
| 735 | 2052425 | 1 | 1 | 2 | 3 | 3 | 2 | 4 | 4 | 0 | 0 | 4 | 1 | 1 | 0 | 1 | 1 | 1 | 0 | 1 | 0 | 1 | 0 | 0 | 2 | 0 |
| 736 | 2052426 | 1 | 1 | 2 | 3 | 3 | 2 | 4 | 4 | 0 | 0 | 4 | 1 | 1 | 1 | 1 | 1 | 1 | 1 | 1 | 0 | 1 | 1 | 0 | 4 | 1 |
| 737 | 2052427 | 1 | 0 | 3 | 2 | 3 | 1 | 4 | 4 | 1 | 1 | 1 | 2 | 1 | 0 | 1 | 1 | 1 | 0 | 1 | 0 | 1 | 1 | 0 | 3 | 1 |
| 738 | 2052428 | 1 | 1 | 4 | 3 | 3 | 1 | 4 | 4 | 1 | 1 | 4 | 2 | 2 | 0 | 1 | 1 | 1 | 0 | 1 | 0 | 1 | 0 | 0 | 2 | 0 |
| 739 | 2052429 | 1 | 0 | 1 | 4 | 3 | 2 | 2 | 4 | 0 | 0 | 1 | 1 | 1 | 0 | 1 | 0 | 1 | 1 | 1 | 0 | 1 | 0 | 0 | 2 | 0 |
| 740 | 2052430 | 1 | 0 | 1 | 5 | 2 | 1 | 4 | 4 | 0 | 0 | 3 | 1 | 2 | 0 | 1 | 0 | 1 | 0 | 0 | 0 | 1 | 0 | 0 | 2 | 0 |
| 741 | 205251 | 1 | 0 | 2 | 2 | 2 | 1 | 1 | 2 | 0 | 0 | 2 | 1 | 1 | 1 | 1 | 1 | 1 | 0 | 1 | 0 | 1 | 0 | 0 | 3 | 1 |
| 742 | 205252 | 1 | 0 | 1 | 3 | 1 | 1 | 1 | 2 | 0 | 0 | 2 | 1 | 1 | 0 | 1 | 1 | 1 | 0 | 1 | 0 | 1 | 0 | 0 | 2 | 0 |
| 743 | 205253 | 1 | 0 | 4 | 2 | 2 | 2 | 1 | 2 | 0 | 0 | 1 | 2 | 1 | 0 | 1 | 1 | 1 | 0 | 1 | 0 | 1 | 1 | 0 | 3 | 1 |
| 744 | 205254 | 1 | 1 | 2 | 5 | 2 | 1 | 1 | 2 | 0 | 0 | 2 | 1 | 1 | 0 | 1 | 1 | 1 | 0 | 1 | 0 | 0 | 0 | 0 | 2 | 0 |
| 745 | 205255 | 1 | 0 | 1 | 2 | 1 | 2 | 1 | 2 | 0 | 0 | 3 | 1 | 1 | 0 | 1 | 1 | 1 | 0 | 1 | 0 | 1 | 0 | 0 | 2 | 0 |
| 746 | 205256 | 1 | 0 | 2 | 2 | 2 | 2 | 1 | 2 | 0 | 0 | 1 | 1 | 1 | 0 | 1 | 1 | 1 | 0 | 1 | 0 | 1 | 0 | 0 | 2 | 0 |
| 747 | 205257 | 1 | 0 | 2 | 2 | 2 | 2 | 1 | 2 | 0 | 0 | 2 | 1 | 1 | 0 | 1 | 1 | 1 | 0 | 1 | 0 | 1 | 0 | 0 | 2 | 0 |
| 748 | 205258 | 1 | 1 | 1 | 2 | 1 | 1 | 1 | 2 | 0 | 0 | 2 | 1 | 1 | 0 | 1 | 1 | 1 | 0 | 0 | 0 | 1 | 0 | 0 | 2 | 0 |
| 749 | 205259 | 1 | 0 | 1 | 3 | 1 | 2 | 1 | 2 | 0 | 0 | 2 | 1 | 2 | 1 | 1 | 1 | 1 | 0 | 1 | 0 | 1 | 0 | 0 | 3 | 1 |
| 750 | 2052510 | 1 | 1 | 1 | 5 | 1 | 1 | 1 | 2 | 0 | 0 | 2 | 1 | 1 | 0 | 1 | 1 | 1 | 0 | 0 | 0 | 1 | 0 | 0 | 2 | 0 |
| 751 | 2052511 | 1 | 1 | 2 | 2 | 1 | 2 | 1 | 2 | 0 | 0 | 2 | 1 | 1 | 0 | 1 | 1 | 1 | 0 | 1 | 0 | 1 | 0 | 0 | 2 | 0 |

| | | | | | | | | | | | | | | | | | | | | | | | | | | |
|---|---|---|---|---|---|---|---|---|---|---|---|---|---|---|---|---|---|---|---|---|---|---|---|---|---|---|
| 752 | 2052512 | 1 | 0 | 2 | 1 | 1 | 2 | 1 | 2 | 0 | 0 | 2 | 1 | 1 | 0 | 1 | 1 | 1 | 0 | 1 | 0 | 1 | 0 | 0 | 2 | 0 |
| 753 | 2052513 | 1 | 0 | 1 | 2 | 1 | 2 | 1 | 2 | 0 | 0 | 2 | 1 | 1 | 0 | 1 | 1 | 1 | 0 | 0 | 0 | 0 | 0 | 0 | 2 | 0 |
| 754 | 2052514 | 1 | 0 | 4 | 2 | 2 | 2 | 1 | 2 | 0 | 0 | 2 | 2 | 1 | 0 | 1 | 1 | 1 | 0 | 1 | 0 | 1 | 0 | 0 | 2 | 0 |
| 755 | 2052515 | 1 | 0 | 2 | 2 | 2 | 2 | 1 | 2 | 1 | 0 | 2 | 1 | 1 | 0 | 1 | 1 | 1 | 0 | 1 | 0 | 1 | 0 | 0 | 2 | 0 |
| 756 | 2052516 | 1 | 0 | 4 | 2 | 2 | 2 | 1 | 2 | 0 | 0 | 3 | 2 | 1 | 1 | 1 | 1 | 1 | 0 | 1 | 0 | 1 | 1 | 0 | 3 | 1 |
| 757 | 2052517 | 1 | 0 | 1 | 2 | 1 | 1 | 2 | 2 | 0 | 0 | 2 | 1 | 1 | 0 | 1 | 1 | 1 | 0 | 1 | 0 | 1 | 0 | 0 | 2 | 0 |
| 758 | 2052518 | 1 | 0 | 1 | 2 | 1 | 2 | 2 | 2 | 0 | 0 | 2 | 1 | 1 | 0 | 1 | 1 | 1 | 0 | 1 | 0 | 1 | 0 | 0 | 2 | 0 |
| 759 | 2052519 | 1 | 1 | 1 | 2 | 1 | 2 | 1 | 2 | 0 | 0 | 2 | 1 | 1 | 0 | 1 | 1 | 1 | 0 | 1 | 0 | 1 | 0 | 0 | 2 | 0 |
| 760 | 2052520 | 1 | 0 | 1 | 1 | 2 | 2 | 1 | 2 | 0 | 0 | 2 | 1 | 1 | 0 | 1 | 1 | 1 | 0 | 0 | 0 | 1 | 0 | 0 | 2 | 0 |
| 761 | 2052521 | 1 | 0 | 1 | 3 | 1 | 2 | 1 | 3 | 0 | 0 | 2 | 1 | 1 | 0 | 1 | 1 | 1 | 0 | 1 | 0 | 1 | 0 | 0 | 2 | 0 |
| 762 | 2052522 | 1 | 0 | 1 | 2 | 2 | 2 | 1 | 2 | 0 | 0 | 2 | 1 | 1 | 0 | 1 | 1 | 1 | 0 | 1 | 0 | 1 | 0 | 0 | 2 | 0 |
| 763 | 2052523 | 1 | 0 | 1 | 3 | 2 | 2 | 1 | 3 | 0 | 0 | 2 | 1 | 1 | 0 | 1 | 1 | 1 | 0 | 1 | 0 | 1 | 0 | 0 | 2 | 0 |
| 764 | 2052524 | 1 | 0 | 1 | 3 | 2 | 2 | 1 | 3 | 0 | 0 | 2 | 1 | 1 | 0 | 1 | 1 | 1 | 0 | 1 | 0 | 1 | 0 | 0 | 2 | 0 |
| 765 | 2052525 | 1 | 0 | 2 | 3 | 3 | 2 | 2 | 3 | 0 | 0 | 3 | 1 | 1 | 0 | 1 | 1 | 1 | 0 | 1 | 0 | 1 | 0 | 0 | 2 | 0 |
| 766 | 2052526 | 1 | 0 | 2 | 2 | 1 | 2 | 2 | 3 | 0 | 0 | 2 | 1 | 1 | 0 | 1 | 1 | 1 | 0 | 1 | 0 | 1 | 0 | 0 | 2 | 0 |
| 767 | 2052527 | 1 | 0 | 1 | 1 | 2 | 1 | 3 | 3 | 0 | 0 | 1 | 1 | 1 | 0 | 1 | 1 | 1 | 0 | 1 | 0 | 1 | 0 | 0 | 2 | 0 |
| 768 | 2052528 | 1 | 1 | 1 | 3 | 2 | 2 | 3 | 3 | 0 | 0 | 2 | 1 | 1 | 0 | 1 | 1 | 1 | 0 | 1 | 0 | 1 | 0 | 0 | 2 | 0 |
| 769 | 2052529 | 1 | 0 | 1 | 3 | 1 | 2 | 1 | 2 | 0 | 0 | 2 | 1 | 1 | 1 | 1 | 1 | 1 | 0 | 1 | 0 | 1 | 0 | 0 | 3 | 1 |
| 770 | 2052530 | 1 | 1 | 3 | 3 | 2 | 2 | 2 | 2 | 0 | 0 | 3 | 2 | 1 | 0 | 1 | 1 | 1 | 0 | 1 | 0 | 1 | 0 | 0 | 2 | 0 |
| 771 | 206261 | 1 | 1 | 2 | 3 | 2 | 1 | 2 | 3 | 0 | 0 | 2 | 1 | 1 | 1 | 1 | 1 | 1 | 0 | 1 | 0 | 1 | 0 | 0 | 3 | 1 |
| 772 | 206262 | 1 | 1 | 2 | 1 | 1 | 1 | 2 | 3 | 0 | 0 | 2 | 1 | 1 | 0 | 1 | 1 | 1 | 0 | 1 | 0 | 1 | 0 | 0 | 2 | 0 |
| 773 | 206263 | 1 | 1 | 3 | 3 | 2 | 2 | 1 | 3 | 0 | 0 | 2 | 2 | 1 | 1 | 1 | 0 | 1 | 0 | 1 | 0 | 1 | 0 | 0 | 2 | 0 |
| 774 | 206264 | 1 | 1 | 4 | 2 | 2 | 2 | 1 | 3 | 0 | 0 | 2 | 2 | 1 | 1 | 1 | 1 | 1 | 0 | 1 | 0 | 1 | 0 | 1 | 3 | 1 |
| 775 | 206265 | 1 | 1 | 2 | 4 | 2 | 2 | 1 | 3 | 0 | 0 | 2 | 1 | 1 | 1 | 1 | 1 | 1 | 0 | 1 | 0 | 1 | 0 | 0 | 3 | 1 |
| 776 | 206266 | 1 | 0 | 2 | 1 | 1 | 1 | 3 | 3 | 0 | 0 | 2 | 1 | 1 | 0 | 1 | 0 | 1 | 0 | 1 | 0 | 1 | 0 | 0 | 2 | 0 |
| 777 | 206267 | 1 | 1 | 3 | 2 | 2 | 1 | 2 | 3 | 0 | 0 | 2 | 2 | 1 | 1 | 1 | 0 | 1 | 0 | 1 | 0 | 0 | 0 | 1 | 2 | 0 |
| 778 | 206268 | 1 | 0 | 4 | 1 | 2 | 2 | 3 | 3 | 0 | 0 | 1 | 2 | 1 | 1 | 1 | 0 | 1 | 0 | 0 | 0 | 1 | 0 | 0 | 2 | 0 |
| 779 | 206269 | 1 | 0 | 2 | 4 | 2 | 2 | 4 | 3 | 0 | 0 | 2 | 1 | 1 | 1 | 1 | 1 | 1 | 0 | 0 | 0 | 1 | 0 | 0 | 2 | 0 |
| 780 | 2062610 | 1 | 0 | 2 | 1 | 2 | 2 | 4 | 3 | 0 | 0 | 2 | 1 | 1 | 0 | 1 | 0 | 1 | 0 | 1 | 0 | 1 | 0 | 0 | 2 | 0 |
| 781 | 2062611 | 1 | 0 | 1 | 2 | 1 | 1 | 4 | 3 | 0 | 0 | 2 | 1 | 1 | 1 | 1 | 1 | 1 | 0 | 1 | 0 | 1 | 0 | 0 | 3 | 1 |
| 782 | 2062612 | 1 | 0 | 4 | 1 | 1 | 2 | 4 | 3 | 0 | 0 | 2 | 2 | 1 | 1 | 1 | 1 | 1 | 0 | 1 | 0 | 1 | 0 | 1 | 3 | 1 |
| 783 | 2062613 | 1 | 0 | 1 | 3 | 2 | 2 | 4 | 3 | 0 | 0 | 2 | 1 | 1 | 0 | 1 | 0 | 1 | 0 | 1 | 0 | 1 | 0 | 0 | 2 | 0 |
| 784 | 2062614 | 1 | 0 | 2 | 3 | 2 | 1 | 4 | 3 | 1 | 1 | 2 | 1 | 1 | 1 | 1 | 1 | 1 | 1 | 1 | 0 | 1 | 0 | 0 | 3 | 1 |
| 785 | 2062615 | 1 | 1 | 1 | 4 | 1 | 1 | 3 | 3 | 0 | 0 | 2 | 1 | 1 | 0 | 1 | 0 | 0 | 0 | 0 | 0 | 0 | 0 | 0 | 2 | 0 |
| 786 | 2062616 | 1 | 1 | 1 | 3 | 1 | 1 | 4 | 3 | 0 | 0 | 2 | 1 | 1 | 0 | 1 | 1 | 1 | 0 | 0 | 0 | 1 | 0 | 0 | 2 | 0 |
| 787 | 2062617 | 1 | 0 | 3 | 1 | 1 | 2 | 3 | 3 | 0 | 0 | 5 | 2 | 1 | 0 | 1 | 1 | 1 | 0 | 0 | 0 | 1 | 0 | 0 | 2 | 0 |
| 788 | 2062618 | 1 | 1 | 1 | 5 | 2 | 2 | 3 | 3 | 0 | 0 | 2 | 1 | 1 | 0 | 1 | 1 | 1 | 1 | 1 | 0 | 1 | 1 | 0 | 3 | 1 |
| 789 | 2062619 | 1 | 0 | 2 | 2 | 2 | 2 | 4 | 3 | 0 | 0 | 2 | 1 | 1 | 0 | 1 | 0 | 1 | 0 | 1 | 0 | 1 | 0 | 1 | 2 | 0 |
| 790 | 2062620 | 1 | 0 | 1 | 2 | 2 | 2 | 2 | 3 | 0 | 0 | 2 | 1 | 1 | 0 | 1 | 1 | 1 | 0 | 0 | 0 | 1 | 0 | 0 | 2 | 0 |
| 791 | 2062621 | 1 | 0 | 1 | 2 | 1 | 2 | 2 | 3 | 0 | 0 | 1 | 1 | 1 | 1 | 1 | 0 | 1 | 0 | 1 | 0 | 0 | 0 | 0 | 2 | 0 |
| 792 | 2062622 | 1 | 0 | 1 | 2 | 3 | 2 | 1 | 3 | 0 | 0 | 3 | 1 | 1 | 1 | 1 | 1 | 1 | 0 | 0 | 0 | 1 | 0 | 0 | 2 | 0 |
| 793 | 2062623 | 1 | 0 | 1 | 4 | 2 | 1 | 2 | 3 | 0 | 0 | 2 | 1 | 1 | 0 | 1 | 1 | 1 | 0 | 0 | 0 | 0 | 0 | 0 | 2 | 0 |
| 794 | 2062624 | 1 | 1 | 2 | 3 | 2 | 2 | 1 | 3 | 0 | 0 | 2 | 1 | 1 | 1 | 1 | 1 | 1 | 0 | 1 | 0 | 1 | 0 | 1 | 3 | 1 |
| 795 | 2062625 | 1 | 1 | 3 | 3 | 2 | 1 | 2 | 3 | 0 | 0 | 2 | 2 | 1 | 0 | 1 | 0 | 1 | 0 | 0 | 0 | 1 | 0 | 0 | 2 | 0 |
| 796 | 206271 | 1 | 0 | 1 | 4 | 1 | 2 | 1 | 3 | 0 | 0 | 2 | 1 | 1 | 0 | 1 | 0 | 1 | 0 | 0 | 0 | 1 | 1 | 0 | 2 | 0 |
| 797 | 206272 | 1 | 0 | 4 | 1 | 3 | 1 | 1 | 3 | 0 | 0 | 2 | 2 | 1 | 0 | 1 | 1 | 1 | 0 | 1 | 0 | 1 | 0 | 0 | 2 | 0 |
| 798 | 206273 | 1 | 0 | 2 | 2 | 2 | 2 | 1 | 3 | 0 | 0 | 2 | 1 | 1 | 0 | 1 | 0 | 1 | 0 | 0 | 0 | 1 | 0 | 0 | 2 | 0 |
| 799 | 206274 | 1 | 0 | 2 | 1 | 3 | 1 | 1 | 3 | 0 | 0 | 1 | 1 | 1 | 0 | 1 | 0 | 1 | 0 | 0 | 0 | 0 | 0 | 0 | 2 | 0 |
| 800 | 206275 | 1 | 1 | 1 | 1 | 1 | 2 | 1 | 3 | 0 | 0 | 2 | 1 | 1 | 0 | 1 | 0 | 1 | 0 | 0 | 0 | 0 | 0 | 0 | 2 | 0 |
| 801 | 206276 | 1 | 0 | 2 | 3 | 2 | 2 | 4 | 3 | 0 | 0 | 2 | 1 | 1 | 0 | 1 | 1 | 1 | 0 | 0 | 0 | 1 | 1 | 0 | 2 | 0 |
| 802 | 206277 | 1 | 1 | 4 | 1 | 2 | 1 | 4 | 3 | 0 | 0 | 2 | 2 | 1 | 1 | 1 | 0 | 1 | 0 | 1 | 0 | 1 | 0 | 0 | 2 | 0 |
| 803 | 206278 | 1 | 0 | 4 | 2 | 2 | 1 | 4 | 3 | 0 | 0 | 2 | 2 | 1 | 0 | 1 | 1 | 1 | 0 | 1 | 0 | 1 | 0 | 1 | 3 | 1 |
| 804 | 206279 | 1 | 1 | 2 | 3 | 1 | 2 | 4 | 3 | 0 | 0 | 2 | 1 | 1 | 1 | 1 | 0 | 1 | 0 | 1 | 0 | 1 | 0 | 1 | 3 | 1 |
| 805 | 2062710 | 1 | 0 | 2 | 2 | 1 | 1 | 4 | 3 | 0 | 0 | 2 | 1 | 1 | 1 | 1 | 0 | 1 | 0 | 1 | 0 | 1 | 0 | 1 | 3 | 1 |
| 806 | 2062711 | 1 | 0 | 1 | 3 | 1 | 2 | 2 | 1 | 0 | 0 | 2 | 1 | 1 | 0 | 1 | 0 | 1 | 0 | 1 | 0 | 1 | 1 | 1 | 3 | 1 |
| 807 | 2062712 | 1 | 0 | 1 | 2 | 2 | 1 | 4 | 3 | 0 | 0 | 2 | 1 | 1 | 1 | 1 | 0 | 1 | 0 | 0 | 0 | 1 | 0 | 0 | 2 | 0 |
| 808 | 2062713 | 1 | 1 | 3 | 1 | 2 | 2 | 4 | 3 | 0 | 0 | 2 | 2 | 1 | 1 | 1 | 0 | 1 | 1 | 1 | 0 | 0 | 0 | 0 | 2 | 0 |
| 809 | 2062714 | 1 | 1 | 2 | 1 | 2 | 1 | 4 | 3 | 0 | 0 | 2 | 1 | 1 | 0 | 1 | 0 | 1 | 0 | 0 | 0 | 0 | 0 | 0 | 2 | 0 |
| 810 | 2062715 | 1 | 1 | 1 | 1 | 2 | 2 | 4 | 3 | 0 | 0 | 2 | 1 | 1 | 1 | 1 | 0 | 0 | 0 | 0 | 0 | 0 | 0 | 0 | 2 | 0 |
| 811 | 2062716 | 1 | 1 | 3 | 2 | 1 | 2 | 4 | 3 | 0 | 0 | 2 | 2 | 1 | 1 | 1 | 0 | 1 | 0 | 1 | 0 | 0 | 0 | 0 | 2 | 0 |
| 812 | 2062717 | 1 | 0 | 3 | 1 | 2 | 1 | 4 | 3 | 0 | 0 | 2 | 2 | 1 | 0 | 1 | 1 | 1 | 0 | 1 | 0 | 1 | 0 | 0 | 2 | 0 |
| 813 | 2062718 | 1 | 1 | 1 | 1 | 1 | 1 | 4 | 3 | 0 | 0 | 2 | 1 | 1 | 0 | 1 | 0 | 0 | 0 | 0 | 0 | 0 | 0 | 0 | 2 | 0 |
| 814 | 2062719 | 1 | 0 | 2 | 2 | 2 | 1 | 4 | 3 | 0 | 0 | 2 | 1 | 1 | 0 | 1 | 0 | 1 | 0 | 1 | 0 | 1 | 0 | 1 | 2 | 0 |
| 815 | 2062720 | 1 | 0 | 3 | 1 | 2 | 1 | 4 | 3 | 0 | 0 | 1 | 2 | 1 | 0 | 1 | 0 | 1 | 0 | 1 | 0 | 1 | 0 | 0 | 2 | 0 |
| 816 | 2062721 | 1 | 1 | 1 | 4 | 1 | 2 | 4 | 3 | 0 | 0 | 2 | 1 | 1 | 0 | 1 | 1 | 1 | 0 | 0 | 0 | 1 | 0 | 0 | 2 | 0 |
| 817 | 2062722 | 1 | 0 | 3 | 2 | 1 | 2 | 4 | 3 | 0 | 0 | 2 | 2 | 1 | 1 | 1 | 1 | 1 | 0 | 0 | 0 | 1 | 0 | 0 | 2 | 0 |
| 818 | 2062723 | 1 | 1 | 2 | 3 | 1 | 2 | 4 | 3 | 0 | 0 | 2 | 1 | 1 | 1 | 1 | 0 | 1 | 1 | 1 | 0 | 1 | 0 | 1 | 3 | 1 |
| 819 | 2062724 | 1 | 0 | 2 | 2 | 2 | 2 | 4 | 3 | 0 | 0 | 2 | 1 | 1 | 1 | 1 | 1 | 1 | 0 | 0 | 0 | 1 | 0 | 0 | 2 | 0 |
| 820 | 2062725 | 1 | 1 | 2 | 2 | 2 | 2 | 4 | 3 | 0 | 0 | 2 | 1 | 1 | 1 | 1 | 0 | 1 | 0 | 1 | 0 | 1 | 0 | 1 | 3 | 1 |
| 821 | 2062726 | 1 | 1 | 1 | 1 | 1 | 1 | 2 | 1 | 0 | 0 | 2 | 1 | 1 | 0 | 1 | 0 | 1 | 0 | 1 | 0 | 1 | 1 | 1 | 3 | 1 |
| 822 | 2062727 | 1 | 1 | 1 | 3 | 2 | 2 | 2 | 3 | 0 | 0 | 2 | 1 | 1 | 0 | 1 | 0 | 0 | 0 | 0 | 0 | 0 | 0 | 0 | 2 | 0 |
| 823 | 2062728 | 1 | 0 | 2 | 1 | 2 | 2 | 2 | 3 | 0 | 0 | 2 | 1 | 1 | 0 | 1 | 0 | 1 | 0 | 0 | 0 | 0 | 0 | 0 | 2 | 0 |
| 824 | 2062729 | 1 | 0 | 1 | 3 | 2 | 2 | 2 | 3 | 0 | 0 | 2 | 1 | 1 | 0 | 1 | 0 | 1 | 0 | 1 | 0 | 1 | 0 | 1 | 2 | 0 |
| 825 | 2062730 | 1 | 0 | 2 | 2 | 1 | 2 | 2 | 3 | 0 | 0 | 1 | 1 | 1 | 0 | 1 | 0 | 1 | 0 | 1 | 0 | 1 | 0 | 1 | 2 | 0 |
| 826 | 206281 | 1 | 0 | 3 | 3 | 2 | 2 | 3 | 3 | 0 | 0 | 1 | 2 | 1 | 0 | 1 | 0 | 1 | 0 | 0 | 0 | 1 | 0 | 0 | 2 | 0 |
| 827 | 206282 | 1 | 1 | 4 | 2 | 2 | 2 | 2 | 3 | 0 | 0 | 2 | 2 | 1 | 0 | 1 | 1 | 1 | 0 | 0 | 0 | 1 | 0 | 0 | 2 | 0 |
| 828 | 206283 | 1 | 1 | 2 | 3 | 2 | 2 | 1 | 3 | 0 | 0 | 2 | 1 | 1 | 1 | 1 | 1 | 1 | 1 | 0 | 0 | 1 | 0 | 0 | 3 | 1 |
| 829 | 206284 | 1 | 1 | 2 | 3 | 2 | 2 | 1 | 3 | 0 | 0 | 2 | 1 | 1 | 0 | 1 | 1 | 1 | 1 | 0 | 0 | 0 | 0 | 0 | 2 | 0 |
| 830 | 206285 | 1 | 0 | 2 | 2 | 2 | 2 | 1 | 3 | 0 | 0 | 3 | 1 | 1 | 0 | 1 | 1 | 1 | 0 | 0 | 0 | 1 | 1 | 0 | 2 | 0 |
| 831 | 206286 | 1 | 0 | 2 | 5 | 2 | 2 | 1 | 3 | 0 | 0 | 1 | 1 | 1 | 0 | 1 | 1 | 1 | 0 | 0 | 0 | 1 | 0 | 0 | 2 | 0 |
| 832 | 206287 | 1 | 0 | 2 | 2 | 2 | 2 | 1 | 3 | 0 | 0 | 3 | 1 | 1 | 0 | 1 | 1 | 1 | 0 | 1 | 0 | 0 | 1 | 0 | 2 | 0 |
| 833 | 206288 | 1 | 0 | 1 | 2 | 2 | 2 | 4 | 2 | 0 | 0 | 1 | 1 | 1 | 0 | 1 | 1 | 1 | 0 | 0 | 0 | 1 | 0 | 0 | 2 | 0 |
| 834 | 206289 | 1 | 0 | 4 | 1 | 2 | 2 | 4 | 2 | 0 | 0 | 1 | 2 | 1 | 0 | 1 | 0 | 1 | 0 | 0 | 0 | 1 | 0 | 0 | 2 | 0 |
| 835 | 2062810 | 1 | 1 | 1 | 2 | 2 | 2 | 4 | 2 | 0 | 0 | 2 | 1 | 1 | 0 | 1 | 1 | 1 | 0 | 0 | 0 | 0 | 0 | 0 | 2 | 0 |
| 836 | 2062811 | 1 | 1 | 1 | 4 | 1 | 2 | 4 | 2 | 0 | 0 | 2 | 1 | 1 | 0 | 1 | 1 | 1 | 0 | 0 | 0 | 1 | 0 | 0 | 2 | 0 |
| 837 | 2062812 | 1 | 1 | 1 | 2 | 1 | 2 | 4 | 2 | 0 | 0 | 2 | 1 | 1 | 0 | 1 | 1 | 1 | 0 | 0 | 0 | 1 | 0 | 0 | 2 | 0 |
| 838 | 2062813 | 1 | 0 | 3 | 1 | 1 | 1 | 4 | 2 | 0 | 0 | 3 | 2 | 1 | 0 | 1 | 0 | 1 | 0 | 0 | 0 | 1 | 0 | 0 | 2 | 0 |
| 839 | 2062814 | 1 | 1 | 4 | 4 | 2 | 2 | 1 | 1 | 0 | 0 | 2 | 2 | 2 | 0 | 1 | 1 | 1 | 1 | 0 | 0 | 1 | 1 | 1 | 3 | 1 |
| 840 | 2062815 | 1 | 0 | 2 | 3 | 1 | 2 | 3 | 1 | 0 | 0 | 1 | 1 | 1 | 0 | 1 | 0 | 1 | 0 | 0 | 0 | 1 | 0 | 0 | 2 | 0 |
| 841 | 2062816 | 1 | 1 | 2 | 1 | 2 | 1 | 2 | 1 | 0 | 0 | 2 | 1 | 2 | 0 | 1 | 1 | 1 | 0 | 0 | 0 | 0 | 0 | 1 | 2 | 0 |
| 842 | 2062817 | 1 | 0 | 2 | 4 | 2 | 2 | 1 | 1 | 0 | 0 | 1 | 1 | 1 | 1 | 1 | 0 | 1 | 0 | 0 | 0 | 1 | 0 | 0 | 2 | 0 |
| 843 | 2062818 | 1 | 0 | 1 | 3 | 2 | 2 | 4 | 2 | 0 | 0 | 1 | 1 | 1 | 0 | 1 | 1 | 1 | 1 | 0 | 0 | 1 | 0 | 0 | 2 | 0 |
| 844 | 2062819 | 1 | 0 | 2 | 4 | 2 | 2 | 2 | 1 | 0 | 0 | 5 | 1 | 2 | 0 | 1 | 0 | 1 | 1 | 0 | 0 | 1 | 0 | 0 | 2 | 0 |
| 845 | 2062820 | 1 | 1 | 1 | 4 | 3 | 2 | 4 | 1 | 0 | 0 | 2 | 1 | 2 | 0 | 1 | 1 | 1 | 0 | 0 | 0 | 1 | 0 | 1 | 2 | 0 |

| | | | | | | | | | | | | | | | | | | | | | | | | | | |
|---|---|---|---|---|---|---|---|---|---|---|---|---|---|---|---|---|---|---|---|---|---|---|---|---|---|---|
| 846 | 2062821 | 1 | 0 | 1 | 3 | 2 | 2 | 1 | 2 | 0 | 0 | 2 | 1 | 2 | 0 | 1 | 1 | 1 | 0 | 1 | 0 | 1 | 0 | 1 | 3 | 1 |
| 847 | 2062822 | 1 | 1 | 2 | 3 | 3 | 2 | 2 | 1 | 0 | 0 | 2 | 1 | 2 | 1 | 1 | 0 | 1 | 1 | 1 | 1 | 1 | 1 | 0 | 4 | 1 |
| 848 | 2062823 | 1 | 1 | 1 | 3 | 3 | 2 | 1 | 1 | 0 | 0 | 2 | 1 | 1 | 0 | 1 | 1 | 1 | 1 | 0 | 1 | 1 | 0 | 0 | 3 | 1 |
| 849 | 2062824 | 1 | 0 | 1 | 3 | 2 | 2 | 1 | 1 | 0 | 0 | 2 | 1 | 2 | 1 | 1 | 1 | 1 | 1 | 0 | 0 | 1 | 0 | 0 | 3 | 1 |
| 850 | 2062825 | 1 | 0 | 2 | 2 | 2 | 2 | 1 | 1 | 0 | 0 | 1 | 1 | 2 | 1 | 1 | 1 | 1 | 0 | 0 | 0 | 1 | 0 | 0 | 2 | 0 |
| 851 | 2062826 | 1 | 1 | 1 | 3 | 1 | 1 | 1 | 1 | 0 | 0 | 2 | 1 | 2 | 0 | 1 | 0 | 1 | 0 | 0 | 0 | 1 | 0 | 0 | 2 | 0 |
| 852 | 2062827 | 1 | 1 | 4 | 3 | 3 | 2 | 2 | 1 | 0 | 0 | 3 | 2 | 1 | 0 | 1 | 0 | 1 | 0 | 0 | 0 | 1 | 0 | 0 | 2 | 0 |
| 853 | 2062828 | 1 | 1 | 2 | 5 | 2 | 1 | 3 | 3 | 0 | 0 | 2 | 1 | 1 | 0 | 1 | 1 | 1 | 1 | 1 | 0 | 1 | 1 | 0 | 3 | 1 |
| 854 | 2062829 | 1 | 1 | 1 | 5 | 2 | 2 | 1 | 3 | 0 | 0 | 2 | 1 | 1 | 0 | 1 | 1 | 1 | 0 | 0 | 0 | 1 | 0 | 0 | 2 | 0 |
| 855 | 2062830 | 1 | 1 | 4 | 4 | 3 | 2 | 1 | 1 | 0 | 0 | 2 | 2 | 2 | 1 | 1 | 1 | 1 | 1 | 0 | 1 | 1 | 0 | 0 | 3 | 1 |
| 856 | 206291 | 1 | 1 | 1 | 5 | 2 | 1 | 1 | 1 | 0 | 0 | 2 | 1 | 1 | 0 | 1 | 1 | 1 | 0 | 0 | 1 | 1 | 0 | 1 | 3 | 1 |
| 857 | 206292 | 1 | 0 | 2 | 3 | 2 | 2 | 1 | 1 | 0 | 0 | 2 | 1 | 1 | 1 | 1 | 1 | 1 | 0 | 0 | 0 | 1 | 0 | 1 | 3 | 1 |
| 858 | 206293 | 1 | 0 | 1 | 3 | 2 | 1 | 2 | 1 | 0 | 0 | 3 | 1 | 1 | 1 | 1 | 1 | 1 | 0 | 1 | 0 | 1 | 0 | 1 | 3 | 1 |
| 859 | 206294 | 1 | 1 | 2 | 2 | 2 | 2 | 3 | 1 | 0 | 0 | 2 | 1 | 1 | 0 | 1 | 1 | 1 | 0 | 1 | 0 | 1 | 0 | 1 | 3 | 1 |
| 860 | 206295 | 1 | 1 | 1 | 5 | 2 | 2 | 2 | 1 | 1 | 0 | 2 | 1 | 1 | 0 | 1 | 0 | 1 | 0 | 0 | 0 | 1 | 0 | 1 | 2 | 0 |
| 861 | 206296 | 1 | 0 | 1 | 4 | 2 | 2 | 3 | 1 | 0 | 0 | 2 | 1 | 1 | 0 | 1 | 1 | 1 | 0 | 0 | 1 | 1 | 0 | 0 | 2 | 0 |
| 862 | 206297 | 1 | 1 | 1 | 5 | 2 | 2 | 4 | 1 | 0 | 0 | 2 | 1 | 1 | 0 | 1 | 1 | 1 | 0 | 0 | 1 | 1 | 0 | 1 | 3 | 1 |
| 863 | 206298 | 1 | 1 | 1 | 5 | 1 | 1 | 4 | 1 | 0 | 0 | 2 | 1 | 1 | 0 | 0 | 0 | 0 | 0 | 0 | 0 | 0 | 0 | 0 | 1 | 0 |
| 864 | 206299 | 1 | 0 | 2 | 4 | 3 | 2 | 4 | 1 | 0 | 0 | 2 | 1 | 1 | 1 | 1 | 1 | 1 | 1 | 1 | 1 | 1 | 1 | 0 | 4 | 1 |
| 865 | 2062910 | 1 | 1 | 1 | 5 | 3 | 1 | 4 | 1 | 0 | 0 | 2 | 1 | 1 | 0 | 0 | 0 | 0 | 0 | 0 | 0 | 0 | 0 | 0 | 1 | 0 |
| 866 | 2062911 | 1 | 0 | 2 | 2 | 2 | 2 | 4 | 1 | 1 | 1 | 1 | 1 | 2 | 0 | 1 | 0 | 1 | 1 | 1 | 0 | 1 | 0 | 1 | 3 | 1 |
| 867 | 2062912 | 1 | 0 | 1 | 3 | 2 | 2 | 4 | 1 | 0 | 0 | 2 | 1 | 2 | 0 | 1 | 0 | 1 | 0 | 0 | 0 | 1 | 0 | 0 | 2 | 0 |
| 868 | 2062913 | 1 | 0 | 1 | 2 | 2 | 2 | 1 | 1 | 0 | 0 | 2 | 1 | 1 | 1 | 1 | 1 | 1 | 0 | 0 | 0 | 1 | 1 | 0 | 3 | 1 |
| 869 | 2062914 | 1 | 0 | 1 | 3 | 2 | 2 | 1 | 1 | 0 | 0 | 2 | 1 | 1 | 0 | 1 | 1 | 1 | 0 | 1 | 1 | 1 | 0 | 0 | 3 | 1 |
| 870 | 2062915 | 1 | 1 | 5 | 1 | 2 | 2 | 1 | 1 | 1 | 1 | 4 | 3 | 1 | 1 | 1 | 1 | 1 | 0 | 1 | 1 | 1 | 0 | 1 | 4 | 1 |
| 871 | 2062916 | 1 | 1 | 4 | 2 | 3 | 2 | 1 | 1 | 1 | 1 | 5 | 2 | 1 | 0 | 1 | 1 | 1 | 1 | 0 | 0 | 1 | 1 | 0 | 3 | 1 |
| 872 | 2062917 | 1 | 1 | 1 | 5 | 2 | 2 | 3 | 1 | 0 | 0 | 2 | 1 | 1 | 1 | 1 | 0 | 1 | 0 | 1 | 0 | 1 | 0 | 0 | 2 | 0 |
| 873 | 2062918 | 1 | 0 | 3 | 2 | 2 | 2 | 2 | 1 | 0 | 0 | 1 | 2 | 1 | 0 | 1 | 0 | 1 | 0 | 1 | 0 | 1 | 0 | 0 | 2 | 0 |
| 874 | 2062919 | 1 | 0 | 1 | 3 | 2 | 2 | 1 | 1 | 0 | 0 | 1 | 1 | 1 | 0 | 1 | 1 | 1 | 0 | 1 | 0 | 1 | 0 | 0 | 2 | 0 |
| 875 | 2062920 | 1 | 0 | 4 | 1 | 3 | 2 | 1 | 1 | 0 | 0 | 3 | 2 | 1 | 0 | 1 | 0 | 1 | 1 | 1 | 0 | 1 | 1 | 0 | 3 | 1 |
| 876 | 2062921 | 1 | 1 | 2 | 3 | 3 | 2 | 1 | 1 | 0 | 0 | 2 | 1 | 1 | 1 | 1 | 0 | 1 | 0 | 0 | 0 | 1 | 0 | 0 | 2 | 0 |
| 877 | 2062922 | 1 | 0 | 1 | 2 | 2 | 2 | 2 | 1 | 0 | 0 | 1 | 1 | 1 | 0 | 1 | 1 | 1 | 0 | 0 | 0 | 0 | 1 | 1 | 2 | 0 |
| 878 | 2062923 | 1 | 0 | 4 | 3 | 2 | 2 | 3 | 1 | 0 | 0 | 1 | 2 | 1 | 0 | 1 | 1 | 1 | 1 | 1 | 0 | 1 | 0 | 0 | 3 | 1 |
| 879 | 2062924 | 1 | 1 | 1 | 4 | 3 | 2 | 1 | 1 | 0 | 0 | 2 | 1 | 1 | 0 | 1 | 1 | 1 | 0 | 0 | 1 | 1 | 0 | 0 | 2 | 0 |
| 880 | 2062925 | 1 | 1 | 1 | 2 | 2 | 2 | 1 | 1 | 0 | 0 | 2 | 1 | 1 | 0 | 1 | 0 | 1 | 0 | 1 | 0 | 1 | 0 | 0 | 2 | 0 |
| 881 | 2062926 | 1 | 0 | 2 | 2 | 2 | 2 | 1 | 1 | 0 | 0 | 1 | 1 | 1 | 0 | 1 | 0 | 1 | 1 | 1 | 0 | 1 | 0 | 0 | 2 | 0 |
| 882 | 2062927 | 1 | 0 | 2 | 2 | 2 | 2 | 1 | 1 | 0 | 0 | 1 | 1 | 1 | 1 | 1 | 0 | 1 | 1 | 1 | 0 | 1 | 1 | 0 | 3 | 1 |
| 883 | 2062928 | 1 | 0 | 1 | 2 | 2 | 2 | 2 | 1 | 0 | 0 | 1 | 1 | 1 | 1 | 1 | 0 | 1 | 0 | 1 | 0 | 1 | 0 | 0 | 2 | 0 |
| 884 | 2062929 | 1 | 0 | 1 | 2 | 2 | 2 | 3 | 1 | 0 | 0 | 1 | 1 | 1 | 0 | 1 | 0 | 1 | 0 | 0 | 0 | 0 | 0 | 0 | 2 | 0 |
| 885 | 2062930 | 1 | 1 | 2 | 4 | 2 | 2 | 3 | 1 | 0 | 0 | 2 | 1 | 1 | 0 | 1 | 1 | 0 | 0 | 0 | 0 | 0 | 0 | 0 | 2 | 0 |
| 886 | 206301 | 1 | 0 | 2 | 5 | 2 | 1 | 1 | 1 | 0 | 0 | 3 | 1 | 1 | 1 | 1 | 1 | 1 | 0 | 1 | 0 | 1 | 1 | 0 | 3 | 1 |
| 887 | 206302 | 1 | 0 | 1 | 4 | 2 | 1 | 2 | 1 | 0 | 0 | 3 | 1 | 1 | 1 | 1 | 1 | 1 | 1 | 1 | 0 | 1 | 1 | 0 | 4 | 1 |
| 888 | 206303 | 1 | 0 | 4 | 5 | 1 | 1 | 1 | 1 | 0 | 0 | 1 | 2 | 2 | 0 | 0 | 0 | 0 | 0 | 0 | 0 | 0 | 0 | 0 | 1 | 0 |
| 889 | 206304 | 1 | 0 | 4 | 1 | 2 | 1 | 4 | 1 | 1 | 1 | 1 | 2 | 2 | 1 | 1 | 1 | 1 | 0 | 0 | 0 | 1 | 1 | 1 | 3 | 1 |
| 890 | 206305 | 1 | 0 | 1 | 5 | 2 | 1 | 2 | 1 | 0 | 0 | 3 | 1 | 1 | 0 | 0 | 0 | 0 | 0 | 0 | 0 | 0 | 0 | 0 | 1 | 0 |
| 891 | 206306 | 1 | 1 | 3 | 4 | 2 | 2 | 1 | 1 | 0 | 0 | 2 | 2 | 1 | 1 | 1 | 1 | 1 | 0 | 1 | 1 | 1 | 1 | 1 | 4 | 1 |
| 892 | 206307 | 1 | 1 | 3 | 3 | 2 | 1 | 1 | 1 | 0 | 0 | 4 | 2 | 1 | 1 | 1 | 1 | 1 | 1 | 1 | 0 | 1 | 1 | 0 | 4 | 1 |
| 893 | 206308 | 1 | 0 | 1 | 5 | 2 | 1 | 3 | 1 | 0 | 0 | 1 | 1 | 1 | 1 | 0 | 0 | 0 | 0 | 0 | 0 | 0 | 0 | 0 | 2 | 0 |
| 894 | 206309 | 1 | 0 | 5 | 2 | 2 | 1 | 1 | 1 | 1 | 1 | 5 | 3 | 1 | 1 | 1 | 1 | 1 | 0 | 1 | 0 | 1 | 1 | 1 | 4 | 1 |
| 895 | 2063010 | 1 | 1 | 1 | 3 | 3 | 1 | 3 | 1 | 1 | 0 | 4 | 1 | 2 | 1 | 1 | 1 | 1 | 0 | 0 | 0 | 1 | 1 | 1 | 3 | 1 |
| 896 | 2063011 | 1 | 1 | 2 | 4 | 3 | 1 | 1 | 1 | 1 | 1 | 2 | 1 | 1 | 1 | 1 | 1 | 1 | 1 | 0 | 0 | 1 | 1 | 1 | 4 | 1 |
| 897 | 2063012 | 1 | 0 | 2 | 2 | 2 | 1 | 1 | 1 | 0 | 0 | 1 | 1 | 2 | 1 | 1 | 1 | 1 | 0 | 1 | 0 | 1 | 1 | 0 | 3 | 1 |
| 898 | 2063013 | 1 | 0 | 5 | 1 | 3 | 1 | 4 | 1 | 0 | 0 | 1 | 3 | 2 | 1 | 1 | 1 | 1 | 0 | 0 | 0 | 1 | 1 | 0 | 3 | 1 |
| 899 | 2063014 | 1 | 0 | 4 | 5 | 3 | 1 | 2 | 1 | 1 | 1 | 3 | 2 | 2 | 1 | 1 | 1 | 1 | 1 | 0 | 0 | 1 | 1 | 1 | 4 | 1 |
| 900 | 2063015 | 1 | 1 | 1 | 5 | 2 | 1 | 3 | 1 | 0 | 0 | 2 | 1 | 2 | 1 | 0 | 0 | 0 | 0 | 0 | 0 | 0 | 0 | 0 | 2 | 0 |
| 901 | 2063016 | 1 | 0 | 2 | 3 | 2 | 1 | 4 | 1 | 0 | 0 | 1 | 1 | 1 | 1 | 1 | 1 | 1 | 0 | 1 | 0 | 1 | 1 | 1 | 4 | 1 |
| 902 | 2063017 | 1 | 1 | 3 | 4 | 3 | 1 | 4 | 1 | 1 | 1 | 4 | 2 | 1 | 1 | 1 | 1 | 1 | 1 | 0 | 0 | 1 | 1 | 1 | 4 | 1 |
| 903 | 2063018 | 1 | 0 | 4 | 3 | 2 | 1 | 3 | 1 | 1 | 0 | 1 | 2 | 1 | 1 | 1 | 1 | 1 | 0 | 1 | 0 | 1 | 1 | 1 | 4 | 1 |
| 904 | 2063019 | 1 | 0 | 2 | 3 | 1 | 1 | 1 | 1 | 0 | 0 | 1 | 1 | 2 | 1 | 1 | 1 | 1 | 0 | 1 | 0 | 1 | 0 | 0 | 3 | 1 |
| 905 | 2063020 | 1 | 0 | 2 | 2 | 3 | 1 | 1 | 1 | 1 | 0 | 3 | 1 | 2 | 1 | 1 | 1 | 1 | 0 | 0 | 1 | 1 | 0 | 1 | 3 | 1 |
| 906 | 2063021 | 1 | 0 | 2 | 1 | 2 | 1 | 4 | 1 | 0 | 0 | 3 | 1 | 1 | 1 | 1 | 1 | 1 | 0 | 1 | 0 | 1 | 1 | 0 | 3 | 1 |
| 907 | 2063022 | 1 | 1 | 2 | 2 | 2 | 1 | 1 | 1 | 0 | 0 | 2 | 1 | 1 | 1 | 1 | 1 | 1 | 1 | 1 | 0 | 1 | 1 | 0 | 4 | 1 |
| 908 | 2063023 | 1 | 1 | 2 | 2 | 2 | 1 | 1 | 1 | 0 | 0 | 2 | 1 | 1 | 1 | 1 | 0 | 1 | 0 | 1 | 0 | 1 | 1 | 1 | 3 | 1 |
| 909 | 2063024 | 1 | 1 | 2 | 3 | 2 | 1 | 3 | 1 | 0 | 0 | 2 | 1 | 1 | 1 | 1 | 0 | 1 | 1 | 1 | 0 | 0 | 1 | 0 | 3 | 1 |
| 910 | 2063025 | 1 | 0 | 2 | 5 | 2 | 1 | 1 | 1 | 0 | 0 | 2 | 1 | 1 | 0 | 0 | 0 | 0 | 0 | 0 | 0 | 0 | 0 | 0 | 1 | 0 |
| 911 | 206311 | 1 | 0 | 2 | 1 | 2 | 1 | 4 | 4 | 0 | 0 | 1 | 1 | 1 | 1 | 1 | 1 | 1 | 0 | 0 | 0 | 1 | 1 | 0 | 3 | 1 |
| 912 | 206312 | 1 | 0 | 3 | 2 | 2 | 1 | 4 | 4 | 0 | 0 | 1 | 2 | 1 | 1 | 1 | 1 | 1 | 0 | 0 | 0 | 1 | 1 | 1 | 3 | 1 |
| 913 | 206313 | 1 | 1 | 2 | 2 | 2 | 2 | 3 | 4 | 0 | 0 | 2 | 1 | 1 | 0 | 0 | 0 | 0 | 0 | 0 | 0 | 0 | 0 | 0 | 1 | 0 |
| 914 | 206314 | 1 | 1 | 1 | 3 | 2 | 1 | 4 | 4 | 0 | 0 | 2 | 1 | 1 | 1 | 1 | 1 | 1 | 1 | 0 | 0 | 1 | 1 | 1 | 4 | 1 |
| 915 | 206315 | 1 | 0 | 2 | 2 | 2 | 1 | 2 | 4 | 1 | 0 | 1 | 1 | 1 | 1 | 1 | 1 | 1 | 0 | 0 | 0 | 1 | 1 | 0 | 3 | 1 |
| 916 | 206316 | 1 | 0 | 5 | 1 | 3 | 1 | 4 | 4 | 0 | 0 | 1 | 3 | 1 | 1 | 1 | 1 | 1 | 0 | 1 | 0 | 1 | 0 | 0 | 3 | 1 |
| 917 | 206317 | 1 | 0 | 1 | 2 | 2 | 1 | 4 | 4 | 1 | 0 | 3 | 1 | 1 | 1 | 1 | 1 | 1 | 1 | 1 | 0 | 1 | 1 | 1 | 4 | 1 |
| 918 | 206318 | 1 | 1 | 1 | 3 | 2 | 1 | 1 | 4 | 1 | 0 | 2 | 1 | 1 | 1 | 1 | 1 | 1 | 1 | 0 | 0 | 1 | 1 | 0 | 3 | 1 |
| 919 | 206319 | 1 | 0 | 1 | 5 | 1 | 1 | 1 | 4 | 0 | 0 | 1 | 1 | 1 | 0 | 0 | 0 | 0 | 0 | 0 | 0 | 0 | 0 | 0 | 1 | 0 |
| 920 | 2063110 | 1 | 1 | 2 | 2 | 2 | 1 | 3 | 4 | 1 | 0 | 2 | 1 | 1 | 1 | 1 | 1 | 1 | 1 | 1 | 0 | 1 | 0 | 1 | 4 | 1 |
| 921 | 2063111 | 1 | 0 | 1 | 3 | 2 | 1 | 4 | 4 | 0 | 0 | 3 | 1 | 1 | 1 | 1 | 1 | 1 | 0 | 1 | 0 | 1 | 1 | 1 | 4 | 1 |
| 922 | 2063112 | 1 | 0 | 1 | 2 | 2 | 1 | 3 | 4 | 0 | 0 | 1 | 1 | 1 | 1 | 1 | 1 | 1 | 0 | 0 | 0 | 1 | 0 | 0 | 2 | 0 |
| 923 | 2063113 | 1 | 1 | 2 | 2 | 2 | 1 | 2 | 4 | 0 | 0 | 2 | 1 | 1 | 1 | 1 | 1 | 1 | 0 | 0 | 0 | 1 | 1 | 1 | 3 | 1 |
| 924 | 2063114 | 1 | 1 | 2 | 3 | 2 | 1 | 2 | 4 | 1 | 0 | 2 | 1 | 1 | 1 | 1 | 1 | 1 | 1 | 1 | 0 | 1 | 1 | 0 | 4 | 1 |
| 925 | 2063115 | 1 | 0 | 2 | 3 | 2 | 1 | 4 | 4 | 0 | 0 | 1 | 1 | 1 | 1 | 0 | 0 | 0 | 0 | 0 | 0 | 0 | 0 | 0 | 2 | 0 |
| 926 | 2063116 | 1 | 1 | 2 | 2 | 2 | 1 | 3 | 4 | 1 | 0 | 2 | 1 | 1 | 1 | 1 | 1 | 1 | 1 | 1 | 0 | 1 | 1 | 0 | 4 | 1 |
| 927 | 2063117 | 1 | 0 | 3 | 3 | 2 | 1 | 4 | 4 | 0 | 0 | 1 | 2 | 2 | 1 | 1 | 1 | 1 | 0 | 1 | 0 | 1 | 0 | 1 | 3 | 1 |
| 928 | 2063118 | 1 | 1 | 2 | 3 | 2 | 1 | 3 | 4 | 0 | 0 | 3 | 1 | 1 | 0 | 1 | 1 | 1 | 1 | 1 | 0 | 1 | 1 | 0 | 3 | 1 |
| 929 | 2063119 | 1 | 0 | 2 | 2 | 2 | 1 | 3 | 4 | 0 | 0 | 1 | 1 | 1 | 1 | 1 | 1 | 1 | 0 | 0 | 0 | 1 | 1 | 0 | 3 | 1 |
| 930 | 2063120 | 1 | 1 | 2 | 1 | 2 | 1 | 1 | 4 | 0 | 0 | 2 | 1 | 1 | 1 | 1 | 1 | 1 | 1 | 1 | 0 | 1 | 1 | 0 | 4 | 1 |
| 931 | 2063121 | 1 | 1 | 2 | 2 | 2 | 1 | 4 | 4 | 0 | 0 | 2 | 1 | 1 | 1 | 1 | 1 | 1 | 0 | 0 | 0 | 1 | 0 | 1 | 3 | 1 |
| 932 | 2063122 | 1 | 1 | 1 | 4 | 2 | 1 | 3 | 4 | 0 | 0 | 2 | 1 | 1 | 1 | 1 | 1 | 1 | 0 | 1 | 0 | 1 | 1 | 1 | 4 | 1 |
| 933 | 2063123 | 1 | 0 | 1 | 2 | 2 | 1 | 3 | 4 | 0 | 0 | 1 | 1 | 1 | 1 | 1 | 1 | 1 | 1 | 1 | 0 | 1 | 0 | 1 | 4 | 1 |
| 934 | 2063124 | 1 | 0 | 1 | 1 | 3 | 1 | 1 | 4 | 0 | 0 | 1 | 1 | 1 | 1 | 1 | 1 | 1 | 0 | 0 | 0 | 1 | 1 | 1 | 3 | 1 |
| 935 | 2063125 | 1 | 1 | 2 | 2 | 3 | 2 | 4 | 4 | 0 | 0 | 2 | 1 | 1 | 1 | 1 | 1 | 1 | 0 | 0 | 0 | 1 | 1 | 1 | 3 | 1 |
| 936 | 206321 | 1 | 0 | 1 | 2 | 1 | 2 | 4 | 4 | 0 | 0 | 1 | 1 | 1 | 0 | 1 | 1 | 1 | 0 | 0 | 0 | 1 | 0 | 0 | 2 | 0 |
| 937 | 206322 | 1 | 0 | 1 | 2 | 1 | 1 | 4 | 4 | 0 | 0 | 3 | 1 | 1 | 0 | 1 | 1 | 1 | 0 | 1 | 0 | 1 | 0 | 0 | 2 | 0 |
| 938 | 206323 | 1 | 0 | 2 | 1 | 2 | 1 | 4 | 4 | 0 | 0 | 1 | 1 | 1 | 0 | 1 | 0 | 1 | 0 | 1 | 0 | 1 | 0 | 1 | 2 | 0 |
| 939 | 206324 | 1 | 1 | 2 | 4 | 1 | 1 | 4 | 4 | 0 | 0 | 2 | 1 | 1 | 0 | 1 | 1 | 1 | 0 | 0 | 0 | 1 | 0 | 1 | 2 | 0 |

| | | | | | | | | | | | | | | | | | | | | | | | | |
|---|---|---|---|---|---|---|---|---|---|---|---|---|---|---|---|---|---|---|---|---|---|---|---|---|
| 940 | 206325 | 1 | 0 | 3 | 1 | 2 | 1 | 4 | 4 | 0 | 0 | 1 | 2 | 1 | 0 | 1 | 1 | 0 | 0 | 0 | 0 | 0 | 0 | 0 | 2 | 0 |
| 941 | 206326 | 1 | 0 | 1 | 2 | 1 | 2 | 4 | 4 | 0 | 0 | 2 | 1 | 1 | 0 | 1 | 0 | 1 | 0 | 0 | 0 | 1 | 0 | 0 | 2 | 0 |
| 942 | 206327 | 1 | 0 | 1 | 5 | 1 | 2 | 4 | 4 | 0 | 0 | 2 | 1 | 1 | 0 | 1 | 1 | 1 | 0 | 0 | 0 | 1 | 0 | 0 | 2 | 0 |
| 943 | 206328 | 1 | 0 | 1 | 3 | 1 | 2 | 4 | 4 | 0 | 0 | 1 | 1 | 1 | 0 | 1 | 1 | 1 | 0 | 1 | 0 | 1 | 0 | 0 | 2 | 0 |
| 944 | 206329 | 1 | 0 | 3 | 1 | 3 | 2 | 1 | 1 | 0 | 0 | 1 | 2 | 1 | 0 | 1 | 1 | 1 | 1 | 0 | 1 | 0 | 0 | 1 | 3 | 1 |
| 945 | 2063210 | 1 | 0 | 3 | 1 | 1 | 2 | 1 | 1 | 0 | 0 | 3 | 2 | 1 | 0 | 1 | 1 | 1 | 0 | 1 | 1 | 0 | 1 | 1 | 3 | 1 |
| 946 | 2063211 | 1 | 1 | 1 | 4 | 1 | 2 | 1 | 1 | 0 | 0 | 2 | 1 | 1 | 0 | 1 | 1 | 1 | 0 | 0 | 1 | 0 | 1 | 0 | 2 | 0 |
| 947 | 2063212 | 1 | 1 | 2 | 3 | 2 | 2 | 1 | 1 | 1 | 1 | 2 | 1 | 1 | 0 | 1 | 0 | 0 | 0 | 1 | 0 | 0 | 0 | 0 | 2 | 0 |
| 948 | 2063213 | 1 | 0 | 1 | 3 | 2 | 2 | 1 | 1 | 0 | 0 | 2 | 1 | 1 | 0 | 1 | 1 | 1 | 0 | 1 | 0 | 1 | 1 | 1 | 3 | 1 |
| 949 | 2063214 | 1 | 0 | 2 | 2 | 2 | 2 | 4 | 4 | 0 | 0 | 1 | 1 | 2 | 0 | 1 | 1 | 1 | 0 | 1 | 0 | 1 | 0 | 0 | 2 | 0 |
| 950 | 2063215 | 1 | 0 | 3 | 1 | 2 | 2 | 4 | 4 | 0 | 0 | 5 | 2 | 1 | 0 | 1 | 1 | 1 | 0 | 1 | 0 | 1 | 0 | 0 | 2 | 0 |
| 951 | 2063216 | 1 | 0 | 1 | 4 | 2 | 1 | 4 | 4 | 0 | 0 | 3 | 1 | 1 | 0 | 1 | 1 | 1 | 0 | 0 | 0 | 1 | 0 | 0 | 2 | 0 |
| 952 | 2063217 | 1 | 1 | 2 | 2 | 2 | 2 | 2 | 1 | 0 | 0 | 2 | 1 | 1 | 1 | 1 | 0 | 1 | 0 | 1 | 0 | 1 | 1 | 1 | 3 | 1 |
| 953 | 2063218 | 1 | 1 | 4 | 1 | 2 | 2 | 2 | 1 | 0 | 0 | 2 | 2 | 1 | 0 | 1 | 0 | 1 | 0 | 1 | 1 | 0 | 0 | 0 | 2 | 0 |
| 954 | 2063219 | 1 | 0 | 2 | 5 | 2 | 2 | 1 | 1 | 0 | 0 | 2 | 1 | 1 | 0 | 1 | 0 | 0 | 0 | 0 | 0 | 0 | 0 | 0 | 2 | 0 |
| 955 | 2063220 | 1 | 0 | 1 | 2 | 2 | 2 | 1 | 1 | 0 | 0 | 1 | 1 | 1 | 0 | 1 | 0 | 0 | 0 | 1 | 0 | 0 | 0 | 0 | 2 | 0 |
| 956 | 2063221 | 1 | 1 | 2 | 2 | 2 | 1 | 4 | 1 | 0 | 0 | 2 | 1 | 1 | 0 | 1 | 1 | 1 | 0 | 1 | 0 | 1 | 0 | 0 | 2 | 0 |
| 957 | 2063222 | 1 | 0 | 2 | 3 | 3 | 2 | 1 | 1 | 0 | 0 | 1 | 1 | 1 | 0 | 1 | 0 | 1 | 0 | 1 | 0 | 1 | 0 | 0 | 2 | 0 |
| 958 | 2063223 | 1 | 0 | 2 | 2 | 2 | 2 | 1 | 4 | 0 | 0 | 1 | 1 | 1 | 0 | 1 | 0 | 1 | 0 | 1 | 0 | 1 | 0 | 0 | 2 | 0 |
| 959 | 2063224 | 1 | 0 | 1 | 5 | 2 | 1 | 1 | 1 | 0 | 0 | 3 | 1 | 1 | 0 | 1 | 1 | 1 | 0 | 1 | 0 | 0 | 0 | 0 | 2 | 0 |
| 960 | 2063225 | 1 | 0 | 1 | 3 | 1 | 2 | 4 | 4 | 0 | 0 | 3 | 1 | 1 | 0 | 1 | 1 | 1 | 0 | 0 | 0 | 0 | 0 | 0 | 2 | 0 |
| 961 | 2063226 | 1 | 1 | 2 | 2 | 1 | 1 | 2 | 1 | 0 | 0 | 2 | 1 | 1 | 0 | 1 | 0 | 1 | 0 | 1 | 0 | 1 | 0 | 1 | 2 | 0 |
| 962 | 2063227 | 1 | 1 | 3 | 3 | 2 | 2 | 2 | 1 | 0 | 0 | 2 | 2 | 1 | 0 | 1 | 0 | 1 | 0 | 1 | 0 | 1 | 0 | 1 | 2 | 0 |
| 963 | 2063228 | 1 | 1 | 3 | 1 | 2 | 2 | 2 | 1 | 0 | 0 | 2 | 2 | 1 | 0 | 1 | 0 | 1 | 0 | 1 | 0 | 0 | 0 | 0 | 2 | 0 |
| 964 | 2063229 | 1 | 0 | 4 | 1 | 3 | 2 | 1 | 1 | 0 | 0 | 1 | 2 | 1 | 1 | 1 | 1 | 1 | 0 | 1 | 0 | 1 | 0 | 1 | 3 | 1 |
| 965 | 2063230 | 1 | 1 | 1 | 4 | 1 | 1 | 4 | 4 | 0 | 0 | 2 | 1 | 1 | 0 | 1 | 1 | 0 | 0 | 0 | 0 | 0 | 0 | 0 | 2 | 0 |
| 966 | 206331 | 1 | 1 | 1 | 3 | 1 | 1 | 4 | 3 | 0 | 0 | 2 | 1 | 1 | 0 | 1 | 1 | 1 | 0 | 0 | 0 | 0 | 0 | 0 | 2 | 0 |
| 967 | 206332 | 1 | 1 | 1 | 3 | 1 | 2 | 4 | 3 | 0 | 0 | 2 | 1 | 1 | 0 | 1 | 1 | 1 | 0 | 0 | 0 | 1 | 0 | 0 | 2 | 0 |
| 968 | 206333 | 1 | 0 | 2 | 2 | 1 | 2 | 4 | 3 | 0 | 0 | 2 | 1 | 1 | 0 | 1 | 1 | 1 | 0 | 0 | 0 | 1 | 1 | 1 | 3 | 1 |
| 969 | 206334 | 1 | 1 | 3 | 5 | 1 | 1 | 3 | 3 | 0 | 0 | 2 | 2 | 1 | 0 | 1 | 1 | 1 | 0 | 1 | 0 | 0 | 0 | 0 | 2 | 0 |
| 970 | 206335 | 1 | 1 | 2 | 3 | 1 | 2 | 2 | 1 | 0 | 0 | 2 | 1 | 1 | 1 | 1 | 1 | 1 | 1 | 1 | 0 | 1 | 0 | 0 | 3 | 1 |
| 971 | 206336 | 1 | 1 | 2 | 2 | 2 | 2 | 1 | 1 | 0 | 0 | 2 | 1 | 1 | 1 | 1 | 1 | 1 | 0 | 0 | 0 | 1 | 0 | 0 | 2 | 0 |
| 972 | 206337 | 1 | 0 | 1 | 3 | 1 | 2 | 1 | 1 | 0 | 0 | 2 | 1 | 1 | 1 | 1 | 1 | 1 | 0 | 1 | 0 | 1 | 0 | 0 | 3 | 1 |
| 973 | 206338 | 1 | 0 | 2 | 4 | 1 | 2 | 1 | 1 | 0 | 0 | 1 | 1 | 1 | 1 | 1 | 1 | 1 | 0 | 0 | 0 | 1 | 1 | 0 | 3 | 1 |
| 974 | 206339 | 1 | 1 | 2 | 3 | 2 | 2 | 2 | 1 | 0 | 0 | 2 | 1 | 1 | 1 | 1 | 1 | 1 | 0 | 0 | 0 | 1 | 0 | 1 | 3 | 1 |
| 975 | 2063310 | 1 | 0 | 1 | 2 | 1 | 2 | 1 | 1 | 0 | 0 | 2 | 1 | 1 | 0 | 1 | 1 | 1 | 0 | 1 | 0 | 1 | 0 | 0 | 2 | 0 |
| 976 | 2063311 | 1 | 1 | 2 | 5 | 1 | 1 | 1 | 1 | 0 | 0 | 2 | 1 | 1 | 0 | 1 | 1 | 1 | 0 | 0 | 0 | 1 | 0 | 0 | 2 | 0 |
| 977 | 2063312 | 1 | 1 | 1 | 2 | 1 | 1 | 1 | 1 | 0 | 0 | 4 | 1 | 1 | 0 | 1 | 1 | 1 | 0 | 0 | 0 | 0 | 0 | 0 | 2 | 0 |
| 978 | 2063313 | 1 | 1 | 2 | 2 | 2 | 1 | 1 | 3 | 0 | 0 | 2 | 1 | 1 | 0 | 1 | 1 | 1 | 0 | 0 | 0 | 0 | 0 | 0 | 2 | 0 |
| 979 | 2063314 | 1 | 0 | 2 | 4 | 1 | 1 | 2 | 4 | 0 | 0 | 2 | 1 | 1 | 1 | 1 | 1 | 1 | 0 | 1 | 0 | 1 | 0 | 0 | 3 | 1 |
| 980 | 2063315 | 1 | 1 | 2 | 4 | 2 | 1 | 1 | 3 | 0 | 0 | 2 | 1 | 1 | 1 | 1 | 1 | 1 | 0 | 0 | 0 | 1 | 0 | 0 | 2 | 0 |
| 981 | 2063316 | 1 | 0 | 2 | 1 | 1 | 2 | 1 | 3 | 0 | 0 | 2 | 1 | 1 | 0 | 1 | 0 | 1 | 1 | 0 | 0 | 0 | 0 | 0 | 2 | 0 |
| 982 | 2063317 | 1 | 0 | 1 | 2 | 1 | 2 | 1 | 3 | 0 | 0 | 2 | 1 | 1 | 1 | 1 | 1 | 1 | 0 | 1 | 0 | 1 | 0 | 0 | 3 | 1 |
| 983 | 2063318 | 1 | 0 | 1 | 4 | 1 | 2 | 1 | 3 | 0 | 0 | 2 | 1 | 1 | 1 | 1 | 1 | 1 | 0 | 1 | 0 | 1 | 0 | 0 | 3 | 1 |
| 984 | 2063319 | 1 | 1 | 1 | 2 | 1 | 2 | 1 | 3 | 0 | 0 | 2 | 1 | 1 | 1 | 1 | 1 | 1 | 0 | 0 | 0 | 1 | 0 | 0 | 2 | 0 |
| 985 | 2063320 | 1 | 1 | 2 | 3 | 2 | 2 | 1 | 3 | 0 | 0 | 2 | 1 | 1 | 0 | 1 | 1 | 1 | 0 | 1 | 0 | 1 | 0 | 1 | 3 | 1 |
| 986 | 2063321 | 1 | 0 | 4 | 3 | 1 | 2 | 2 | 3 | 0 | 0 | 2 | 2 | 1 | 1 | 1 | 1 | 1 | 0 | 0 | 0 | 0 | 0 | 1 | 2 | 0 |
| 987 | 2063322 | 1 | 0 | 2 | 1 | 1 | 1 | 1 | 3 | 0 | 0 | 2 | 1 | 1 | 0 | 1 | 1 | 1 | 0 | 0 | 0 | 1 | 0 | 0 | 2 | 0 |
| 988 | 2063323 | 1 | 1 | 4 | 2 | 2 | 2 | 3 | 3 | 0 | 0 | 2 | 2 | 1 | 0 | 1 | 1 | 1 | 0 | 1 | 0 | 1 | 0 | 1 | 3 | 1 |
| 989 | 2063324 | 1 | 1 | 2 | 4 | 1 | 1 | 3 | 3 | 0 | 0 | 2 | 1 | 1 | 0 | 1 | 1 | 1 | 0 | 0 | 0 | 1 | 0 | 0 | 2 | 0 |
| 990 | 2063325 | 1 | 0 | 2 | 5 | 1 | 1 | 3 | 3 | 0 | 0 | 2 | 1 | 1 | 0 | 1 | 1 | 1 | 1 | 0 | 0 | 1 | 0 | 0 | 2 | 0 |
| 991 | 2063326 | 1 | 1 | 1 | 5 | 1 | 1 | 4 | 3 | 0 | 0 | 4 | 1 | 1 | 0 | 1 | 1 | 1 | 0 | 0 | 0 | 0 | 0 | 0 | 2 | 0 |
| 992 | 2063327 | 1 | 0 | 2 | 5 | 1 | 1 | 1 | 1 | 0 | 0 | 2 | 1 | 1 | 1 | 1 | 1 | 1 | 0 | 1 | 0 | 1 | 0 | 1 | 3 | 1 |
| 993 | 2063328 | 1 | 1 | 1 | 3 | 1 | 2 | 1 | 1 | 0 | 0 | 2 | 1 | 1 | 1 | 1 | 1 | 1 | 0 | 1 | 0 | 1 | 0 | 1 | 3 | 1 |
| 994 | 2063329 | 1 | 0 | 1 | 2 | 2 | 2 | 1 | 3 | 0 | 0 | 2 | 1 | 1 | 0 | 1 | 1 | 1 | 0 | 1 | 0 | 0 | 0 | 0 | 2 | 0 |
| 995 | 2063330 | 1 | 0 | 2 | 1 | 1 | 1 | 1 | 3 | 0 | 0 | 2 | 1 | 1 | 1 | 1 | 1 | 1 | 0 | 0 | 0 | 0 | 0 | 0 | 2 | 0 |
| 996 | 307341 | 2 | 1 | 4 | 5 | 3 | 2 | 1 | 4 | 1 | 1 | 5 | 2 | 4 | 1 | 1 | 1 | 1 | 0 | 0 | 1 | 0 | 1 | 0 | 3 | 1 |
| 997 | 307342 | 2 | 1 | 5 | 5 | 2 | 1 | 1 | 4 | 1 | 1 | 5 | 3 | 4 | 1 | 1 | 1 | 1 | 0 | 1 | 0 | 0 | 1 | 0 | 3 | 1 |
| 998 | 307343 | 2 | 0 | 3 | 2 | 1 | 2 | 2 | 4 | 0 | 0 | 1 | 2 | 4 | 1 | 1 | 1 | 1 | 0 | 0 | 1 | 0 | 1 | 0 | 3 | 1 |
| 999 | 307344 | 2 | 1 | 2 | 4 | 2 | 1 | 1 | 4 | 0 | 0 | 2 | 1 | 4 | 1 | 1 | 1 | 1 | 0 | 0 | 1 | 0 | 0 | 1 | 3 | 1 |
| 1000 | 307345 | 2 | 1 | 3 | 4 | 2 | 2 | 1 | 4 | 0 | 0 | 2 | 2 | 4 | 1 | 1 | 1 | 1 | 0 | 0 | 1 | 0 | 1 | 1 | 3 | 1 |
| 1001 | 307346 | 2 | 1 | 5 | 4 | 2 | 2 | 1 | 4 | 1 | 1 | 5 | 3 | 4 | 1 | 1 | 1 | 1 | 0 | 0 | 1 | 0 | 1 | 0 | 3 | 1 |
| 1002 | 307347 | 2 | 0 | 4 | 3 | 2 | 2 | 3 | 4 | 1 | 1 | 1 | 2 | 4 | 1 | 1 | 1 | 1 | 0 | 0 | 1 | 0 | 0 | 1 | 3 | 1 |
| 1003 | 307348 | 2 | 1 | 4 | 2 | 1 | 2 | 3 | 4 | 0 | 0 | 5 | 2 | 4 | 1 | 1 | 1 | 1 | 0 | 1 | 1 | 0 | 0 | 1 | 3 | 1 |
| 1004 | 307349 | 2 | 1 | 3 | 3 | 2 | 1 | 3 | 4 | 0 | 0 | 2 | 2 | 4 | 1 | 1 | 1 | 1 | 0 | 0 | 1 | 0 | 0 | 1 | 3 | 1 |
| 1005 | 3073410 | 2 | 0 | 3 | 2 | 1 | 2 | 3 | 4 | 0 | 0 | 1 | 2 | 4 | 1 | 1 | 1 | 1 | 1 | 0 | 1 | 0 | 0 | 1 | 3 | 1 |
| 1006 | 3073411 | 2 | 1 | 3 | 1 | 2 | 2 | 3 | 4 | 0 | 0 | 2 | 2 | 4 | 1 | 1 | 1 | 1 | 0 | 0 | 1 | 0 | 1 | 0 | 3 | 1 |
| 1007 | 3073412 | 2 | 0 | 5 | 1 | 3 | 1 | 3 | 4 | 1 | 1 | 5 | 3 | 4 | 1 | 1 | 1 | 1 | 0 | 0 | 1 | 0 | 0 | 1 | 3 | 1 |
| 1008 | 3073413 | 2 | 0 | 2 | 1 | 1 | 2 | 3 | 4 | 0 | 0 | 1 | 1 | 4 | 1 | 1 | 0 | 1 | 0 | 0 | 1 | 0 | 0 | 1 | 2 | 0 |
| 1009 | 3073414 | 2 | 1 | 3 | 5 | 2 | 2 | 3 | 4 | 0 | 0 | 2 | 2 | 4 | 1 | 0 | 0 | 0 | 0 | 0 | 0 | 0 | 0 | 0 | 2 | 0 |
| 1010 | 3073415 | 2 | 1 | 2 | 4 | 1 | 2 | 3 | 4 | 0 | 0 | 2 | 1 | 4 | 1 | 1 | 1 | 1 | 0 | 1 | 1 | 0 | 0 | 1 | 3 | 1 |
| 1011 | 3073416 | 2 | 1 | 3 | 2 | 1 | 1 | 3 | 4 | 0 | 0 | 2 | 2 | 4 | 1 | 1 | 1 | 1 | 0 | 0 | 1 | 0 | 0 | 1 | 3 | 1 |
| 1012 | 3073417 | 2 | 1 | 3 | 2 | 1 | 2 | 3 | 4 | 0 | 0 | 2 | 2 | 4 | 1 | 1 | 1 | 1 | 0 | 0 | 1 | 0 | 0 | 1 | 3 | 1 |
| 1013 | 3073418 | 2 | 1 | 2 | 4 | 1 | 1 | 3 | 4 | 0 | 0 | 2 | 1 | 4 | 1 | 0 | 0 | 0 | 0 | 0 | 0 | 0 | 0 | 0 | 2 | 0 |
| 1014 | 3073419 | 2 | 1 | 3 | 2 | 1 | 2 | 3 | 4 | 0 | 0 | 2 | 2 | 4 | 1 | 1 | 0 | 1 | 0 | 0 | 1 | 0 | 0 | 1 | 2 | 0 |
| 1015 | 3073420 | 2 | 1 | 2 | 2 | 1 | 2 | 3 | 4 | 0 | 0 | 2 | 1 | 4 | 1 | 1 | 1 | 1 | 0 | 0 | 1 | 0 | 1 | 0 | 3 | 1 |
| 1016 | 3073421 | 2 | 1 | 2 | 3 | 2 | 1 | 3 | 4 | 0 | 0 | 2 | 1 | 4 | 1 | 1 | 1 | 1 | 0 | 0 | 1 | 0 | 1 | 0 | 3 | 1 |
| 1017 | 3073422 | 2 | 1 | 2 | 3 | 2 | 2 | 3 | 4 | 0 | 0 | 2 | 1 | 4 | 1 | 1 | 1 | 1 | 0 | 0 | 1 | 0 | 1 | 0 | 3 | 1 |
| 1018 | 3073423 | 2 | 1 | 4 | 4 | 2 | 2 | 3 | 4 | 0 | 0 | 5 | 2 | 4 | 1 | 1 | 0 | 1 | 0 | 0 | 1 | 0 | 1 | 0 | 2 | 0 |
| 1019 | 3073424 | 2 | 1 | 5 | 2 | 3 | 2 | 3 | 4 | 1 | 1 | 5 | 3 | 4 | 1 | 1 | 1 | 1 | 0 | 0 | 1 | 0 | 1 | 1 | 3 | 1 |
| 1020 | 3073425 | 2 | 1 | 2 | 3 | 1 | 2 | 3 | 4 | 0 | 0 | 2 | 1 | 4 | 1 | 1 | 1 | 1 | 0 | 0 | 1 | 0 | 1 | 0 | 3 | 1 |
| 1021 | 3073426 | 2 | 1 | 2 | 4 | 1 | 1 | 3 | 4 | 0 | 0 | 2 | 1 | 4 | 1 | 1 | 1 | 1 | 0 | 0 | 1 | 0 | 1 | 0 | 3 | 1 |
| 1022 | 3073427 | 2 | 1 | 2 | 4 | 1 | 2 | 3 | 4 | 0 | 0 | 2 | 1 | 4 | 1 | 1 | 0 | 1 | 1 | 1 | 1 | 0 | 1 | 0 | 3 | 1 |
| 1023 | 3073428 | 2 | 1 | 5 | 1 | 3 | 2 | 3 | 4 | 1 | 1 | 5 | 3 | 4 | 1 | 1 | 1 | 1 | 0 | 1 | 1 | 0 | 0 | 1 | 3 | 1 |
| 1024 | 3073429 | 2 | 1 | 4 | 2 | 2 | 1 | 3 | 4 | 0 | 0 | 2 | 2 | 4 | 1 | 1 | 1 | 1 | 1 | 0 | 1 | 0 | 1 | 0 | 3 | 1 |
| 1025 | 3073430 | 2 | 1 | 4 | 4 | 1 | 1 | 3 | 4 | 0 | 0 | 2 | 2 | 4 | 1 | 0 | 0 | 0 | 0 | 0 | 0 | 0 | 0 | 0 | 2 | 0 |
| 1026 | 3073431 | 2 | 1 | 2 | 4 | 1 | 1 | 3 | 4 | 0 | 0 | 2 | 1 | 4 | 1 | 1 | 1 | 1 | 1 | 0 | 1 | 0 | 0 | 1 | 3 | 1 |
| 1027 | 3073432 | 2 | 1 | 5 | 2 | 2 | 2 | 3 | 4 | 1 | 1 | 5 | 3 | 4 | 1 | 1 | 1 | 1 | 0 | 0 | 1 | 0 | 1 | 0 | 3 | 1 |
| 1028 | 307351 | 2 | 0 | 5 | 2 | 3 | 2 | 2 | 4 | 1 | 1 | 5 | 3 | 4 | 1 | 1 | 1 | 1 | 1 | 1 | 1 | 0 | 0 | 1 | 4 | 1 |
| 1029 | 307352 | 2 | 1 | 2 | 2 | 1 | 2 | 2 | 4 | 0 | 0 | 2 | 1 | 4 | 1 | 1 | 1 | 1 | 1 | 1 | 1 | 0 | 0 | 1 | 4 | 1 |
| 1030 | 307353 | 2 | 1 | 2 | 2 | 1 | 2 | 2 | 4 | 0 | 0 | 2 | 1 | 4 | 1 | 1 | 0 | 1 | 0 | 0 | 1 | 0 | 0 | 1 | 2 | 0 |
| 1031 | 307354 | 2 | 0 | 2 | 4 | 1 | 2 | 2 | 4 | 0 | 0 | 1 | 1 | 4 | 1 | 1 | 1 | 1 | 1 | 1 | 1 | 0 | 0 | 1 | 4 | 1 |
| 1032 | 307355 | 2 | 1 | 3 | 3 | 2 | 2 | 2 | 4 | 0 | 0 | 2 | 2 | 4 | 1 | 1 | 1 | 1 | 1 | 1 | 1 | 1 | 0 | 1 | 4 | 1 |
| 1033 | 307356 | 2 | 0 | 2 | 2 | 1 | 2 | 2 | 4 | 0 | 0 | 1 | 1 | 4 | 1 | 1 | 1 | 1 | 0 | 0 | 1 | 0 | 0 | 1 | 3 | 1 |

| | | | | | | | | | | | | | | | | | | | | | | | | | | |
|---|---|---|---|---|---|---|---|---|---|---|---|---|---|---|---|---|---|---|---|---|---|---|---|---|---|---|
| 1034 | 307357 | 2 | 0 | 2 | 5 | 1 | 1 | 2 | 4 | 0 | 0 | 1 | 1 | 4 | 1 | 1 | 1 | 1 | 0 | 0 | 1 | 0 | 0 | 1 | 3 | 1 |
| 1035 | 307358 | 2 | 0 | 2 | 4 | 1 | 1 | 2 | 4 | 0 | 0 | 1 | 1 | 4 | 1 | 1 | 1 | 1 | 0 | 0 | 1 | 0 | 0 | 1 | 3 | 1 |
| 1036 | 307359 | 2 | 0 | 3 | 2 | 1 | 1 | 2 | 4 | 0 | 0 | 1 | 2 | 4 | 1 | 1 | 1 | 1 | 0 | 1 | 1 | 0 | 0 | 1 | 3 | 1 |
| 1037 | 3073510 | 2 | 1 | 3 | 2 | 1 | 2 | 2 | 4 | 0 | 0 | 2 | 2 | 4 | 1 | 1 | 1 | 1 | 0 | 1 | 1 | 0 | 0 | 1 | 3 | 1 |
| 1038 | 3073511 | 2 | 1 | 2 | 5 | 2 | 1 | 2 | 4 | 0 | 0 | 2 | 1 | 4 | 1 | 1 | 1 | 1 | 1 | 1 | 1 | 0 | 0 | 1 | 4 | 1 |
| 1039 | 3073512 | 2 | 0 | 4 | 1 | 2 | 2 | 2 | 4 | 1 | 1 | 1 | 2 | 4 | 1 | 1 | 1 | 1 | 0 | 1 | 1 | 1 | 0 | 1 | 4 | 1 |
| 1040 | 3073513 | 2 | 1 | 2 | 3 | 1 | 2 | 2 | 4 | 0 | 0 | 2 | 1 | 4 | 1 | 1 | 1 | 1 | 0 | 0 | 1 | 0 | 0 | 1 | 3 | 1 |
| 1041 | 3073514 | 2 | 1 | 3 | 3 | 1 | 2 | 2 | 4 | 0 | 0 | 2 | 2 | 4 | 1 | 1 | 1 | 1 | 1 | 1 | 1 | 0 | 0 | 1 | 4 | 1 |
| 1042 | 3073515 | 2 | 0 | 2 | 3 | 1 | 2 | 2 | 4 | 0 | 0 | 1 | 1 | 4 | 1 | 1 | 1 | 1 | 1 | 1 | 1 | 0 | 0 | 1 | 4 | 1 |
| 1043 | 3073516 | 2 | 0 | 2 | 3 | 1 | 2 | 2 | 4 | 0 | 0 | 1 | 1 | 4 | 1 | 1 | 1 | 1 | 1 | 1 | 1 | 0 | 0 | 1 | 4 | 1 |
| 1044 | 3073517 | 2 | 0 | 2 | 4 | 1 | 2 | 2 | 4 | 0 | 0 | 1 | 1 | 4 | 1 | 1 | 1 | 1 | 0 | 0 | 1 | 0 | 0 | 1 | 3 | 1 |
| 1045 | 3073518 | 2 | 0 | 3 | 2 | 1 | 1 | 2 | 4 | 0 | 0 | 1 | 2 | 4 | 1 | 1 | 1 | 1 | 0 | 0 | 1 | 0 | 0 | 0 | 2 | 0 |
| 1046 | 3073519 | 2 | 0 | 3 | 2 | 1 | 2 | 2 | 4 | 0 | 0 | 1 | 2 | 4 | 1 | 1 | 1 | 1 | 0 | 0 | 1 | 0 | 0 | 0 | 2 | 0 |
| 1047 | 3073520 | 2 | 1 | 2 | 2 | 1 | 2 | 2 | 4 | 0 | 0 | 2 | 1 | 4 | 1 | 1 | 1 | 1 | 1 | 0 | 1 | 0 | 0 | 1 | 3 | 1 |
| 1048 | 3073521 | 2 | 0 | 5 | 2 | 2 | 1 | 1 | 4 | 1 | 1 | 5 | 3 | 4 | 1 | 1 | 1 | 1 | 0 | 0 | 1 | 0 | 0 | 1 | 3 | 1 |
| 1049 | 3073522 | 2 | 1 | 5 | 2 | 2 | 1 | 1 | 4 | 1 | 1 | 5 | 3 | 4 | 1 | 1 | 1 | 1 | 1 | 0 | 1 | 0 | 0 | 1 | 3 | 1 |
| 1050 | 3073523 | 2 | 0 | 3 | 3 | 2 | 2 | 2 | 4 | 0 | 0 | 1 | 2 | 4 | 1 | 0 | 0 | 0 | 0 | 0 | 0 | 0 | 0 | 0 | 2 | 0 |
| 1051 | 3073524 | 2 | 1 | 5 | 3 | 3 | 2 | 2 | 4 | 1 | 1 | 5 | 3 | 4 | 1 | 1 | 1 | 1 | 1 | 0 | 1 | 0 | 0 | 1 | 3 | 1 |
| 1052 | 3073525 | 2 | 1 | 5 | 2 | 3 | 1 | 1 | 4 | 1 | 1 | 5 | 3 | 4 | 1 | 1 | 1 | 1 | 1 | 0 | 1 | 0 | 0 | 1 | 3 | 1 |
| 1053 | 3073526 | 2 | 1 | 4 | 4 | 3 | 1 | 2 | 4 | 1 | 1 | 5 | 2 | 4 | 1 | 1 | 1 | 1 | 1 | 1 | 1 | 0 | 1 | 0 | 4 | 1 |
| 1054 | 3073527 | 2 | 1 | 5 | 1 | 2 | 1 | 1 | 4 | 1 | 1 | 5 | 3 | 4 | 1 | 1 | 1 | 1 | 0 | 0 | 1 | 0 | 0 | 1 | 3 | 1 |
| 1055 | 3073528 | 2 | 1 | 4 | 3 | 3 | 2 | 2 | 4 | 1 | 1 | 5 | 2 | 4 | 1 | 1 | 1 | 1 | 0 | 0 | 1 | 0 | 0 | 1 | 3 | 1 |
| 1056 | 3073529 | 2 | 1 | 5 | 2 | 3 | 1 | 1 | 4 | 1 | 1 | 5 | 3 | 4 | 1 | 1 | 1 | 1 | 0 | 1 | 1 | 0 | 0 | 1 | 3 | 1 |
| 1057 | 3073530 | 2 | 1 | 2 | 2 | 2 | 1 | 1 | 4 | 0 | 0 | 2 | 1 | 4 | 1 | 1 | 1 | 1 | 0 | 1 | 1 | 0 | 0 | 1 | 3 | 1 |
| 1058 | 3073531 | 2 | 1 | 3 | 2 | 2 | 2 | 1 | 4 | 0 | 0 | 2 | 2 | 4 | 1 | 1 | 1 | 1 | 0 | 0 | 1 | 0 | 1 | 1 | 3 | 1 |
| 1059 | 3073532 | 2 | 1 | 4 | 5 | 2 | 1 | 1 | 4 | 1 | 0 | 5 | 2 | 4 | 1 | 1 | 0 | 1 | 0 | 0 | 0 | 0 | 1 | 1 | 2 | 0 |
| 1060 | 3073533 | 2 | 1 | 5 | 2 | 2 | 1 | 1 | 4 | 1 | 1 | 5 | 3 | 4 | 1 | 1 | 1 | 1 | 1 | 0 | 1 | 0 | 1 | 0 | 3 | 1 |
| 1061 | 307361 | 2 | 1 | 4 | 2 | 2 | 2 | 1 | 3 | 1 | 1 | 2 | 2 | 4 | 1 | 1 | 1 | 1 | 1 | 1 | 1 | 1 | 0 | 0 | 4 | 1 |
| 1062 | 307362 | 2 | 1 | 5 | 2 | 3 | 1 | 1 | 3 | 1 | 1 | 5 | 3 | 4 | 1 | 1 | 1 | 1 | 1 | 1 | 1 | 0 | 1 | 0 | 4 | 1 |
| 1063 | 307363 | 2 | 1 | 5 | 5 | 2 | 1 | 1 | 3 | 1 | 1 | 5 | 3 | 4 | 0 | 1 | 0 | 1 | 0 | 1 | 0 | 1 | 0 | 1 | 2 | 0 |
| 1064 | 307364 | 2 | 1 | 2 | 5 | 2 | 1 | 1 | 3 | 0 | 0 | 2 | 1 | 4 | 1 | 1 | 1 | 1 | 1 | 1 | 1 | 1 | 0 | 1 | 4 | 1 |
| 1065 | 307365 | 2 | 0 | 3 | 3 | 2 | 2 | 1 | 3 | 0 | 0 | 1 | 2 | 4 | 1 | 1 | 1 | 1 | 1 | 1 | 1 | 1 | 0 | 0 | 4 | 1 |
| 1066 | 307366 | 2 | 1 | 4 | 4 | 2 | 2 | 1 | 3 | 0 | 0 | 2 | 2 | 4 | 0 | 1 | 1 | 1 | 1 | 1 | 0 | 1 | 0 | 1 | 3 | 1 |
| 1067 | 307367 | 2 | 1 | 2 | 5 | 1 | 1 | 4 | 3 | 0 | 0 | 2 | 1 | 4 | 1 | 1 | 1 | 1 | 1 | 1 | 1 | 0 | 0 | 0 | 3 | 1 |
| 1068 | 307368 | 2 | 1 | 5 | 4 | 3 | 2 | 1 | 3 | 1 | 1 | 5 | 3 | 2 | 1 | 1 | 1 | 1 | 1 | 1 | 0 | 1 | 1 | 1 | 4 | 1 |
| 1069 | 307369 | 2 | 0 | 5 | 4 | 3 | 2 | 1 | 3 | 1 | 1 | 5 | 3 | 4 | 1 | 1 | 1 | 1 | 1 | 1 | 0 | 1 | 1 | 1 | 4 | 1 |
| 1070 | 3073610 | 2 | 0 | 2 | 2 | 2 | 1 | 1 | 3 | 0 | 0 | 1 | 1 | 4 | 0 | 1 | 0 | 1 | 1 | 1 | 0 | 1 | 0 | 0 | 2 | 0 |
| 1071 | 3073611 | 2 | 0 | 2 | 4 | 2 | 1 | 1 | 3 | 0 | 0 | 2 | 1 | 4 | 1 | 1 | 1 | 1 | 1 | 1 | 1 | 1 | 0 | 1 | 4 | 1 |
| 1072 | 3073612 | 2 | 1 | 5 | 3 | 3 | 1 | 2 | 3 | 0 | 0 | 5 | 3 | 4 | 1 | 1 | 1 | 1 | 1 | 1 | 1 | 1 | 1 | 1 | 4 | 1 |
| 1073 | 3073613 | 2 | 0 | 4 | 2 | 2 | 1 | 2 | 3 | 1 | 0 | 5 | 2 | 4 | 0 | 1 | 1 | 1 | 1 | 1 | 0 | 1 | 1 | 1 | 4 | 1 |
| 1074 | 3073614 | 2 | 1 | 2 | 5 | 1 | 1 | 2 | 3 | 0 | 0 | 2 | 1 | 4 | 0 | 1 | 1 | 1 | 1 | 1 | 1 | 1 | 1 | 1 | 4 | 1 |
| 1075 | 3073615 | 2 | 0 | 4 | 3 | 3 | 1 | 2 | 3 | 1 | 1 | 5 | 2 | 4 | 1 | 1 | 0 | 1 | 1 | 1 | 1 | 1 | 0 | 1 | 4 | 1 |
| 1076 | 3073616 | 2 | 1 | 2 | 4 | 1 | 2 | 2 | 3 | 0 | 0 | 2 | 1 | 4 | 0 | 1 | 1 | 1 | 1 | 1 | 0 | 1 | 0 | 1 | 3 | 1 |
| 1077 | 3073617 | 2 | 1 | 5 | 4 | 3 | 1 | 2 | 3 | 1 | 1 | 5 | 3 | 4 | 1 | 1 | 1 | 1 | 1 | 1 | 0 | 1 | 1 | 1 | 4 | 1 |
| 1078 | 3073618 | 2 | 1 | 4 | 2 | 3 | 1 | 2 | 3 | 0 | 0 | 5 | 2 | 4 | 0 | 1 | 1 | 1 | 1 | 1 | 0 | 1 | 1 | 1 | 4 | 1 |
| 1079 | 3073619 | 2 | 1 | 2 | 5 | 2 | 1 | 2 | 3 | 0 | 0 | 2 | 1 | 4 | 0 | 1 | 1 | 1 | 1 | 1 | 0 | 1 | 0 | 0 | 3 | 1 |
| 1080 | 3073620 | 2 | 1 | 2 | 2 | 2 | 2 | 2 | 3 | 0 | 0 | 2 | 1 | 4 | 1 | 1 | 1 | 1 | 1 | 1 | 1 | 1 | 1 | 1 | 4 | 1 |
| 1081 | 3073621 | 2 | 1 | 4 | 2 | 2 | 2 | 2 | 3 | 0 | 0 | 2 | 2 | 4 | 0 | 1 | 1 | 1 | 1 | 1 | 0 | 1 | 1 | 0 | 3 | 1 |
| 1082 | 3073622 | 2 | 0 | 3 | 3 | 2 | 2 | 4 | 3 | 0 | 0 | 2 | 2 | 4 | 0 | 1 | 1 | 1 | 1 | 1 | 0 | 1 | 1 | 0 | 3 | 1 |
| 1083 | 3073623 | 2 | 1 | 4 | 5 | 2 | 1 | 4 | 3 | 0 | 0 | 2 | 2 | 4 | 0 | 1 | 1 | 1 | 0 | 1 | 0 | 1 | 0 | 0 | 2 | 0 |
| 1084 | 3073624 | 2 | 1 | 3 | 4 | 1 | 2 | 4 | 3 | 0 | 0 | 2 | 2 | 4 | 0 | 1 | 1 | 1 | 1 | 1 | 0 | 1 | 1 | 0 | 3 | 1 |
| 1085 | 3073625 | 2 | 0 | 2 | 5 | 2 | 1 | 4 | 3 | 0 | 0 | 1 | 1 | 4 | 0 | 1 | 1 | 1 | 1 | 1 | 0 | 1 | 1 | 0 | 3 | 1 |
| 1086 | 3073626 | 2 | 1 | 4 | 4 | 2 | 2 | 4 | 3 | 0 | 0 | 2 | 2 | 4 | 0 | 1 | 0 | 1 | 1 | 1 | 0 | 1 | 0 | 0 | 2 | 0 |
| 1087 | 3073627 | 2 | 0 | 2 | 3 | 1 | 1 | 4 | 3 | 0 | 0 | 1 | 1 | 4 | 1 | 1 | 1 | 1 | 1 | 1 | 1 | 0 | 1 | 1 | 4 | 1 |
| 1088 | 3073628 | 2 | 1 | 2 | 5 | 2 | 2 | 4 | 3 | 0 | 0 | 2 | 1 | 4 | 0 | 1 | 1 | 1 | 0 | 1 | 0 | 1 | 0 | 0 | 2 | 0 |
| 1089 | 3073629 | 2 | 1 | 3 | 2 | 2 | 2 | 4 | 3 | 0 | 0 | 2 | 2 | 4 | 0 | 1 | 0 | 1 | 1 | 1 | 0 | 1 | 1 | 0 | 3 | 1 |
| 1090 | 3073630 | 2 | 1 | 3 | 4 | 1 | 2 | 2 | 3 | 0 | 0 | 2 | 2 | 4 | 0 | 1 | 1 | 1 | 1 | 1 | 0 | 1 | 1 | 0 | 3 | 1 |
| 1091 | 307371 | 2 | 0 | 4 | 1 | 2 | 1 | 3 | 1 | 0 | 0 | 1 | 2 | 4 | 1 | 1 | 0 | 1 | 0 | 0 | 0 | 0 | 0 | 0 | 2 | 0 |
| 1092 | 307372 | 2 | 0 | 4 | 2 | 2 | 1 | 3 | 1 | 0 | 0 | 1 | 2 | 4 | 1 | 1 | 1 | 1 | 1 | 1 | 0 | 1 | 1 | 0 | 4 | 1 |
| 1093 | 307373 | 2 | 0 | 2 | 2 | 2 | 2 | 2 | 1 | 0 | 0 | 1 | 1 | 4 | 1 | 1 | 0 | 1 | 0 | 1 | 0 | 0 | 0 | 0 | 2 | 0 |
| 1094 | 307374 | 2 | 0 | 2 | 3 | 2 | 2 | 1 | 1 | 0 | 0 | 1 | 1 | 4 | 0 | 0 | 0 | 0 | 0 | 0 | 0 | 0 | 0 | 0 | 1 | 0 |
| 1095 | 307375 | 2 | 0 | 2 | 2 | 1 | 2 | 4 | 1 | 0 | 0 | 2 | 1 | 4 | 0 | 1 | 1 | 1 | 1 | 1 | 0 | 1 | 1 | 0 | 3 | 1 |
| 1096 | 307376 | 2 | 0 | 4 | 1 | 1 | 1 | 4 | 1 | 0 | 0 | 2 | 2 | 4 | 0 | 1 | 0 | 1 | 1 | 1 | 0 | 1 | 1 | 1 | 3 | 1 |
| 1097 | 307377 | 2 | 1 | 2 | 2 | 2 | 2 | 1 | 1 | 1 | 1 | 2 | 1 | 4 | 1 | 1 | 0 | 1 | 1 | 1 | 1 | 1 | 0 | 1 | 4 | 1 |
| 1098 | 307378 | 2 | 0 | 2 | 5 | 2 | 1 | 4 | 1 | 0 | 0 | 1 | 1 | 4 | 0 | 0 | 0 | 0 | 0 | 0 | 0 | 0 | 0 | 0 | 1 | 0 |
| 1099 | 307379 | 2 | 1 | 2 | 2 | 2 | 2 | 1 | 1 | 0 | 0 | 2 | 1 | 4 | 1 | 1 | 1 | 1 | 1 | 1 | 0 | 1 | 0 | 1 | 4 | 1 |
| 1100 | 3073710 | 2 | 0 | 3 | 3 | 2 | 1 | 1 | 1 | 0 | 0 | 1 | 2 | 4 | 1 | 1 | 1 | 1 | 1 | 1 | 0 | 1 | 0 | 1 | 4 | 1 |
| 1101 | 3073711 | 2 | 1 | 2 | 1 | 1 | 1 | 4 | 1 | 0 | 0 | 2 | 1 | 4 | 0 | 0 | 0 | 0 | 0 | 0 | 0 | 0 | 0 | 0 | 1 | 0 |
| 1102 | 3073712 | 2 | 1 | 1 | 5 | 1 | 1 | 4 | 1 | 0 | 0 | 2 | 1 | 4 | 0 | 0 | 0 | 0 | 0 | 0 | 0 | 0 | 0 | 0 | 1 | 0 |
| 1103 | 3073713 | 2 | 1 | 2 | 1 | 1 | 1 | 3 | 1 | 0 | 0 | 2 | 1 | 2 | 0 | 1 | 0 | 1 | 0 | 1 | 0 | 0 | 0 | 0 | 2 | 0 |
| 1104 | 3073714 | 2 | 1 | 4 | 4 | 2 | 2 | 3 | 1 | 0 | 0 | 2 | 2 | 4 | 0 | 1 | 0 | 1 | 1 | 1 | 1 | 1 | 0 | 0 | 3 | 1 |
| 1105 | 3073715 | 2 | 0 | 3 | 2 | 3 | 2 | 2 | 1 | 1 | 0 | 3 | 2 | 4 | 1 | 1 | 1 | 1 | 1 | 1 | 1 | 0 | 1 | 1 | 4 | 1 |
| 1106 | 3073716 | 2 | 1 | 2 | 2 | 2 | 1 | 3 | 4 | 0 | 0 | 2 | 1 | 4 | 1 | 1 | 1 | 1 | 1 | 1 | 0 | 0 | 0 | 0 | 3 | 1 |
| 1107 | 3073717 | 2 | 0 | 3 | 1 | 1 | 1 | 4 | 4 | 0 | 0 | 1 | 2 | 4 | 0 | 1 | 0 | 1 | 0 | 0 | 0 | 0 | 0 | 0 | 2 | 0 |
| 1108 | 3073718 | 2 | 0 | 3 | 2 | 2 | 2 | 4 | 4 | 0 | 0 | 1 | 2 | 4 | 1 | 0 | 0 | 0 | 0 | 0 | 0 | 0 | 0 | 0 | 2 | 0 |
| 1109 | 3073719 | 2 | 1 | 2 | 5 | 1 | 1 | 4 | 4 | 0 | 0 | 2 | 1 | 4 | 1 | 0 | 0 | 0 | 0 | 0 | 0 | 0 | 0 | 0 | 2 | 0 |
| 1110 | 3073720 | 2 | 0 | 3 | 2 | 1 | 1 | 4 | 4 | 0 | 0 | 1 | 2 | 4 | 0 | 1 | 0 | 1 | 0 | 0 | 0 | 0 | 1 | 1 | 2 | 0 |
| 1111 | 3073721 | 2 | 1 | 2 | 5 | 2 | 2 | 3 | 4 | 0 | 0 | 2 | 1 | 4 | 0 | 1 | 0 | 1 | 0 | 0 | 0 | 0 | 0 | 0 | 2 | 0 |
| 1112 | 3073722 | 2 | 0 | 2 | 2 | 1 | 1 | 4 | 4 | 0 | 0 | 1 | 1 | 4 | 0 | 1 | 0 | 1 | 0 | 0 | 0 | 0 | 0 | 0 | 2 | 0 |
| 1113 | 3073723 | 2 | 0 | 2 | 3 | 2 | 2 | 4 | 4 | 0 | 0 | 1 | 1 | 4 | 1 | 1 | 1 | 1 | 1 | 1 | 0 | 1 | 0 | 1 | 4 | 1 |
| 1114 | 3073724 | 2 | 0 | 2 | 1 | 2 | 1 | 2 | 4 | 0 | 0 | 1 | 1 | 4 | 1 | 1 | 1 | 1 | 1 | 1 | 1 | 1 | 1 | 0 | 4 | 1 |
| 1115 | 3073725 | 2 | 0 | 3 | 4 | 2 | 1 | 3 | 4 | 0 | 0 | 3 | 2 | 4 | 1 | 1 | 0 | 1 | 0 | 0 | 0 | 0 | 0 | 0 | 2 | 0 |
| 1116 | 3073726 | 2 | 1 | 2 | 2 | 2 | 1 | 3 | 4 | 0 | 0 | 4 | 1 | 4 | 1 | 0 | 0 | 0 | 0 | 0 | 0 | 0 | 0 | 0 | 2 | 0 |
| 1117 | 3073727 | 2 | 1 | 2 | 2 | 2 | 2 | 3 | 4 | 0 | 0 | 4 | 1 | 4 | 1 | 1 | 1 | 1 | 1 | 1 | 0 | 1 | 0 | 0 | 3 | 1 |
| 1118 | 3073728 | 2 | 0 | 2 | 1 | 1 | 2 | 2 | 4 | 0 | 0 | 1 | 1 | 4 | 0 | 0 | 0 | 0 | 0 | 0 | 0 | 0 | 0 | 0 | 1 | 0 |
| 1119 | 3073729 | 2 | 0 | 3 | 2 | 2 | 2 | 3 | 4 | 0 | 0 | 1 | 2 | 4 | 1 | 0 | 0 | 0 | 0 | 0 | 0 | 0 | 0 | 0 | 2 | 0 |
| 1120 | 3073730 | 2 | 0 | 2 | 1 | 1 | 1 | 2 | 4 | 0 | 0 | 1 | 1 | 3 | 1 | 0 | 0 | 0 | 0 | 0 | 0 | 0 | 0 | 0 | 2 | 0 |
| 1121 | 307381 | 2 | 0 | 2 | 2 | 2 | 1 | 1 | 4 | 0 | 0 | 2 | 1 | 4 | 1 | 1 | 1 | 1 | 1 | 1 | 1 | 0 | 0 | 1 | 4 | 1 |
| 1122 | 307382 | 2 | 1 | 3 | 4 | 2 | 2 | 1 | 4 | 0 | 0 | 2 | 2 | 4 | 1 | 1 | 1 | 1 | 1 | 1 | 1 | 0 | 0 | 1 | 4 | 1 |
| 1123 | 307383 | 2 | 0 | 3 | 2 | 2 | 2 | 1 | 4 | 0 | 0 | 2 | 2 | 4 | 1 | 1 | 1 | 1 | 1 | 1 | 1 | 0 | 0 | 1 | 4 | 1 |
| 1124 | 307384 | 2 | 1 | 2 | 3 | 2 | 2 | 1 | 4 | 0 | 0 | 2 | 1 | 4 | 1 | 1 | 1 | 1 | 1 | 1 | 1 | 0 | 0 | 0 | 3 | 1 |
| 1125 | 307385 | 2 | 0 | 4 | 1 | 2 | 2 | 1 | 4 | 0 | 0 | 2 | 2 | 4 | 1 | 1 | 1 | 1 | 1 | 1 | 1 | 0 | 1 | 1 | 4 | 1 |
| 1126 | 307386 | 2 | 1 | 2 | 3 | 2 | 2 | 1 | 4 | 0 | 0 | 2 | 1 | 4 | 1 | 1 | 1 | 1 | 1 | 1 | 1 | 1 | 0 | 1 | 4 | 1 |
| 1127 | 307387 | 2 | 0 | 3 | 1 | 2 | 2 | 1 | 4 | 0 | 0 | 2 | 2 | 4 | 1 | 1 | 1 | 1 | 1 | 0 | 1 | 0 | 1 | 1 | 4 | 1 |

| | | | | | | | | | | | | | | | | | | | | | | | | | | |
|---|---|---|---|---|---|---|---|---|---|---|---|---|---|---|---|---|---|---|---|---|---|---|---|---|---|---|
| 1128 | 307388 | 2 | 1 | 3 | 3 | 2 | 1 | 1 | 4 | 0 | 0 | 2 | 2 | 4 | 0 | 1 | 1 | 1 | 1 | 1 | 1 | 0 | 0 | 1 | 3 | 1 |
| 1129 | 307389 | 2 | 1 | 4 | 3 | 2 | 1 | 1 | 4 | 0 | 0 | 2 | 2 | 4 | 0 | 1 | 1 | 1 | 1 | 1 | 1 | 0 | 0 | 1 | 3 | 1 |
| 1130 | 3073810 | 2 | 0 | 4 | 2 | 2 | 1 | 1 | 4 | 1 | 0 | 3 | 2 | 4 | 1 | 1 | 1 | 1 | 1 | 1 | 1 | 0 | 0 | 1 | 4 | 1 |
| 1131 | 3073811 | 2 | 0 | 5 | 2 | 2 | 2 | 1 | 4 | 1 | 0 | 5 | 3 | 4 | 1 | 1 | 1 | 1 | 1 | 1 | 1 | 0 | 0 | 1 | 4 | 1 |
| 1132 | 3073812 | 2 | 1 | 3 | 2 | 2 | 2 | 1 | 4 | 0 | 0 | 2 | 2 | 4 | 1 | 1 | 1 | 1 | 1 | 1 | 1 | 0 | 0 | 1 | 4 | 1 |
| 1133 | 3073813 | 2 | 0 | 2 | 3 | 2 | 1 | 1 | 4 | 0 | 0 | 2 | 1 | 4 | 1 | 1 | 1 | 1 | 1 | 1 | 1 | 0 | 0 | 0 | 3 | 1 |
| 1134 | 3073814 | 2 | 1 | 5 | 2 | 2 | 2 | 1 | 4 | 0 | 0 | 5 | 3 | 4 | 0 | 1 | 1 | 1 | 1 | 1 | 1 | 0 | 1 | 1 | 4 | 1 |
| 1135 | 3073815 | 2 | 0 | 5 | 2 | 3 | 1 | 1 | 4 | 1 | 0 | 5 | 3 | 4 | 0 | 1 | 1 | 1 | 1 | 1 | 1 | 0 | 0 | 1 | 3 | 1 |
| 1136 | 3073816 | 2 | 1 | 2 | 2 | 1 | 2 | 3 | 4 | 0 | 0 | 2 | 1 | 4 | 1 | 1 | 1 | 1 | 1 | 1 | 1 | 0 | 1 | 1 | 4 | 1 |
| 1137 | 3073817 | 2 | 0 | 5 | 2 | 3 | 1 | 1 | 4 | 1 | 0 | 5 | 3 | 4 | 1 | 1 | 1 | 1 | 1 | 1 | 1 | 0 | 1 | 1 | 4 | 1 |
| 1138 | 3073818 | 2 | 1 | 3 | 4 | 2 | 2 | 2 | 4 | 0 | 0 | 2 | 2 | 4 | 1 | 1 | 1 | 1 | 1 | 1 | 1 | 0 | 1 | 1 | 4 | 1 |
| 1139 | 3073819 | 2 | 0 | 4 | 1 | 3 | 1 | 1 | 4 | 1 | 0 | 3 | 2 | 4 | 1 | 1 | 1 | 1 | 1 | 1 | 1 | 0 | 1 | 1 | 4 | 1 |
| 1140 | 3073820 | 2 | 1 | 4 | 3 | 2 | 2 | 2 | 4 | 0 | 0 | 2 | 2 | 4 | 0 | 1 | 1 | 1 | 1 | 1 | 1 | 0 | 0 | 1 | 3 | 1 |
| 1141 | 3073821 | 2 | 0 | 5 | 2 | 2 | 1 | 4 | 4 | 0 | 0 | 5 | 3 | 4 | 1 | 1 | 1 | 1 | 1 | 1 | 1 | 0 | 1 | 1 | 4 | 1 |
| 1142 | 3073822 | 2 | 1 | 2 | 4 | 2 | 2 | 3 | 4 | 0 | 0 | 2 | 1 | 4 | 1 | 1 | 1 | 1 | 1 | 1 | 1 | 1 | 1 | 1 | 4 | 1 |
| 1143 | 3073823 | 2 | 0 | 3 | 2 | 2 | 2 | 2 | 4 | 0 | 0 | 2 | 2 | 4 | 1 | 1 | 1 | 1 | 1 | 1 | 1 | 0 | 1 | 1 | 4 | 1 |
| 1144 | 3073824 | 2 | 1 | 2 | 4 | 2 | 2 | 3 | 4 | 0 | 0 | 2 | 1 | 4 | 1 | 1 | 1 | 1 | 1 | 1 | 1 | 0 | 0 | 1 | 4 | 1 |
| 1145 | 3073825 | 2 | 0 | 4 | 1 | 2 | 1 | 3 | 4 | 0 | 0 | 2 | 2 | 4 | 1 | 1 | 1 | 1 | 1 | 1 | 1 | 0 | 1 | 1 | 4 | 1 |
| 1146 | 3073826 | 2 | 0 | 2 | 4 | 2 | 1 | 3 | 4 | 0 | 0 | 1 | 1 | 4 | 1 | 1 | 1 | 1 | 1 | 1 | 1 | 1 | 1 | 1 | 4 | 1 |
| 1147 | 3073827 | 2 | 1 | 2 | 4 | 2 | 2 | 3 | 4 | 0 | 0 | 2 | 1 | 2 | 1 | 1 | 1 | 1 | 1 | 1 | 1 | 1 | 0 | 1 | 4 | 1 |
| 1148 | 3073828 | 2 | 1 | 4 | 4 | 3 | 2 | 2 | 4 | 1 | 1 | 5 | 2 | 4 | 1 | 1 | 1 | 1 | 1 | 1 | 1 | 0 | 1 | 1 | 4 | 1 |
| 1149 | 3073829 | 2 | 0 | 2 | 2 | 2 | 2 | 4 | 4 | 0 | 0 | 2 | 1 | 4 | 1 | 1 | 1 | 1 | 0 | 1 | 1 | 0 | 1 | 1 | 4 | 1 |
| 1150 | 3073830 | 2 | 1 | 2 | 4 | 2 | 2 | 3 | 4 | 0 | 0 | 2 | 1 | 4 | 1 | 1 | 1 | 1 | 1 | 1 | 1 | 1 | 1 | 0 | 4 | 1 |
| 1151 | 307391 | 2 | 1 | 4 | 3 | 2 | 2 | 3 | 1 | 1 | 1 | 5 | 2 | 4 | 1 | 1 | 1 | 1 | 1 | 1 | 1 | 0 | 0 | 1 | 4 | 1 |
| 1152 | 307392 | 2 | 1 | 4 | 1 | 2 | 2 | 3 | 2 | 0 | 0 | 2 | 2 | 4 | 1 | 1 | 1 | 1 | 1 | 1 | 1 | 0 | 0 | 1 | 4 | 1 |
| 1153 | 307393 | 2 | 1 | 2 | 2 | 2 | 2 | 3 | 2 | 0 | 0 | 2 | 1 | 4 | 0 | 1 | 1 | 1 | 1 | 1 | 1 | 0 | 0 | 1 | 3 | 1 |
| 1154 | 307394 | 2 | 0 | 2 | 2 | 2 | 2 | 3 | 2 | 0 | 0 | 1 | 1 | 4 | 0 | 0 | 0 | 0 | 0 | 0 | 0 | 0 | 0 | 0 | 1 | 0 |
| 1155 | 307395 | 2 | 1 | 4 | 1 | 2 | 1 | 3 | 2 | 0 | 0 | 2 | 2 | 4 | 1 | 0 | 0 | 0 | 0 | 0 | 0 | 0 | 0 | 0 | 2 | 0 |
| 1156 | 307396 | 2 | 0 | 2 | 2 | 2 | 2 | 2 | 3 | 0 | 0 | 1 | 1 | 4 | 1 | 0 | 0 | 0 | 0 | 0 | 0 | 0 | 0 | 0 | 2 | 0 |
| 1157 | 307397 | 2 | 1 | 2 | 3 | 2 | 2 | 2 | 3 | 0 | 0 | 2 | 1 | 4 | 1 | 0 | 0 | 0 | 0 | 0 | 0 | 0 | 0 | 0 | 2 | 0 |
| 1158 | 307398 | 2 | 1 | 3 | 4 | 2 | 1 | 3 | 2 | 0 | 0 | 2 | 2 | 4 | 0 | 0 | 0 | 0 | 0 | 0 | 0 | 0 | 0 | 0 | 1 | 0 |
| 1159 | 307399 | 2 | 0 | 3 | 4 | 1 | 1 | 2 | 2 | 0 | 0 | 1 | 2 | 4 | 0 | 1 | 0 | 1 | 0 | 0 | 0 | 0 | 0 | 0 | 2 | 0 |
| 1160 | 3073910 | 2 | 1 | 2 | 4 | 2 | 2 | 2 | 2 | 0 | 0 | 2 | 1 | 4 | 1 | 1 | 1 | 1 | 1 | 1 | 1 | 0 | 1 | 1 | 4 | 1 |
| 1161 | 3073911 | 2 | 1 | 3 | 5 | 1 | 1 | 2 | 2 | 0 | 0 | 2 | 2 | 4 | 1 | 1 | 1 | 1 | 1 | 0 | 0 | 0 | 0 | 0 | 2 | 0 |
| 1162 | 3073912 | 2 | 0 | 5 | 1 | 2 | 1 | 1 | 2 | 0 | 0 | 5 | 3 | 4 | 1 | 1 | 1 | 1 | 1 | 0 | 1 | 0 | 1 | 1 | 4 | 1 |
| 1163 | 3073913 | 2 | 1 | 5 | 3 | 2 | 1 | 1 | 2 | 1 | 0 | 5 | 3 | 4 | 1 | 1 | 1 | 1 | 0 | 1 | 1 | 0 | 1 | 1 | 4 | 1 |
| 1164 | 3073914 | 2 | 0 | 2 | 2 | 1 | 2 | 2 | 2 | 0 | 0 | 1 | 1 | 4 | 0 | 0 | 0 | 0 | 0 | 0 | 0 | 0 | 0 | 0 | 1 | 0 |
| 1165 | 3073915 | 2 | 0 | 3 | 1 | 2 | 1 | 1 | 2 | 0 | 0 | 1 | 2 | 4 | 0 | 0 | 0 | 0 | 0 | 0 | 0 | 0 | 0 | 0 | 1 | 0 |
| 1166 | 3073916 | 2 | 0 | 2 | 2 | 1 | 2 | 2 | 2 | 0 | 0 | 2 | 1 | 4 | 0 | 0 | 0 | 0 | 0 | 0 | 0 | 0 | 0 | 0 | 1 | 0 |
| 1167 | 3073917 | 2 | 0 | 2 | 2 | 1 | 2 | 1 | 2 | 0 | 0 | 1 | 1 | 4 | 0 | 0 | 0 | 0 | 0 | 0 | 0 | 0 | 0 | 0 | 1 | 0 |
| 1168 | 3073918 | 2 | 1 | 2 | 4 | 1 | 1 | 2 | 2 | 0 | 0 | 2 | 1 | 4 | 0 | 0 | 0 | 0 | 0 | 0 | 0 | 0 | 0 | 0 | 1 | 0 |
| 1169 | 3073919 | 2 | 1 | 2 | 4 | 1 | 2 | 1 | 2 | 0 | 0 | 2 | 1 | 4 | 0 | 0 | 0 | 0 | 0 | 0 | 0 | 0 | 0 | 0 | 1 | 0 |
| 1170 | 3073920 | 2 | 0 | 2 | 2 | 2 | 1 | 2 | 2 | 0 | 0 | 1 | 1 | 4 | 0 | 0 | 0 | 0 | 0 | 0 | 0 | 0 | 0 | 0 | 1 | 0 |
| 1171 | 3073921 | 2 | 1 | 2 | 2 | 1 | 1 | 2 | 2 | 0 | 0 | 2 | 1 | 4 | 0 | 1 | 1 | 1 | 1 | 1 | 1 | 0 | 0 | 1 | 3 | 1 |
| 1172 | 3073922 | 2 | 1 | 2 | 5 | 1 | 2 | 2 | 2 | 0 | 0 | 2 | 1 | 4 | 0 | 1 | 1 | 1 | 1 | 1 | 1 | 0 | 0 | 1 | 3 | 1 |
| 1173 | 3073923 | 2 | 1 | 2 | 2 | 1 | 1 | 2 | 2 | 0 | 0 | 2 | 1 | 4 | 0 | 0 | 0 | 0 | 0 | 0 | 0 | 0 | 0 | 0 | 1 | 0 |
| 1174 | 3073924 | 2 | 0 | 2 | 2 | 2 | 2 | 2 | 2 | 0 | 0 | 1 | 1 | 4 | 1 | 0 | 0 | 0 | 0 | 0 | 0 | 0 | 0 | 0 | 2 | 0 |
| 1175 | 3073925 | 2 | 1 | 2 | 5 | 1 | 1 | 2 | 2 | 0 | 0 | 2 | 1 | 4 | 1 | 0 | 0 | 0 | 0 | 0 | 0 | 0 | 0 | 0 | 2 | 0 |
| 1176 | 3073926 | 2 | 0 | 2 | 2 | 1 | 1 | 2 | 2 | 0 | 0 | 1 | 1 | 4 | 0 | 1 | 1 | 1 | 0 | 1 | 1 | 0 | 0 | 1 | 3 | 1 |
| 1177 | 3073927 | 2 | 0 | 2 | 2 | 2 | 2 | 2 | 2 | 0 | 0 | 1 | 1 | 4 | 0 | 1 | 1 | 1 | 1 | 1 | 1 | 0 | 0 | 1 | 3 | 1 |
| 1178 | 3073928 | 2 | 0 | 2 | 3 | 2 | 1 | 2 | 4 | 0 | 0 | 1 | 1 | 4 | 0 | 0 | 0 | 0 | 0 | 0 | 0 | 0 | 0 | 0 | 1 | 0 |
| 1179 | 3073929 | 2 | 0 | 2 | 3 | 1 | 1 | 2 | 2 | 0 | 0 | 2 | 1 | 4 | 1 | 0 | 0 | 0 | 0 | 0 | 0 | 0 | 0 | 0 | 2 | 0 |
| 1180 | 3073930 | 2 | 1 | 4 | 2 | 2 | 2 | 3 | 2 | 0 | 0 | 5 | 2 | 4 | 0 | 0 | 0 | 0 | 0 | 0 | 0 | 0 | 0 | 0 | 1 | 0 |
| 1181 | 307401 | 2 | 0 | 3 | 3 | 2 | 1 | 4 | 2 | 0 | 0 | 2 | 2 | 4 | 0 | 1 | 1 | 1 | 1 | 1 | 1 | 0 | 0 | 0 | 3 | 1 |
| 1182 | 307402 | 2 | 1 | 2 | 3 | 1 | 1 | 4 | 2 | 0 | 0 | 2 | 1 | 4 | 0 | 1 | 1 | 1 | 0 | 1 | 1 | 1 | 0 | 0 | 3 | 1 |
| 1183 | 307403 | 2 | 0 | 2 | 2 | 1 | 2 | 4 | 2 | 0 | 0 | 2 | 1 | 4 | 1 | 1 | 1 | 1 | 1 | 0 | 1 | 1 | 0 | 1 | 4 | 1 |
| 1184 | 307404 | 2 | 1 | 3 | 3 | 2 | 2 | 4 | 2 | 0 | 0 | 2 | 2 | 4 | 1 | 1 | 1 | 1 | 1 | 1 | 1 | 1 | 0 | 0 | 4 | 1 |
| 1185 | 307405 | 2 | 1 | 2 | 3 | 2 | 2 | 4 | 2 | 0 | 0 | 2 | 1 | 4 | 1 | 1 | 1 | 1 | 0 | 1 | 1 | 0 | 0 | 1 | 3 | 1 |
| 1186 | 307406 | 2 | 1 | 3 | 3 | 2 | 2 | 4 | 2 | 0 | 0 | 2 | 2 | 4 | 1 | 1 | 1 | 1 | 1 | 1 | 1 | 1 | 0 | 0 | 4 | 1 |
| 1187 | 307407 | 2 | 0 | 2 | 2 | 2 | 2 | 4 | 2 | 0 | 0 | 2 | 1 | 4 | 0 | 1 | 1 | 1 | 1 | 0 | 1 | 1 | 0 | 0 | 3 | 1 |
| 1188 | 307408 | 2 | 0 | 4 | 1 | 2 | 2 | 4 | 2 | 0 | 0 | 2 | 2 | 4 | 0 | 1 | 1 | 1 | 1 | 1 | 1 | 1 | 0 | 0 | 3 | 1 |
| 1189 | 307409 | 2 | 0 | 3 | 3 | 2 | 2 | 4 | 2 | 0 | 0 | 2 | 2 | 4 | 1 | 1 | 1 | 1 | 1 | 1 | 1 | 0 | 0 | 1 | 4 | 1 |
| 1190 | 3074010 | 2 | 1 | 5 | 1 | 2 | 1 | 4 | 2 | 0 | 0 | 5 | 3 | 4 | 1 | 1 | 1 | 1 | 1 | 1 | 1 | 0 | 0 | 1 | 4 | 1 |
| 1191 | 3074011 | 2 | 1 | 2 | 3 | 1 | 1 | 1 | 2 | 0 | 0 | 2 | 1 | 4 | 0 | 1 | 0 | 1 | 1 | 1 | 1 | 0 | 0 | 1 | 3 | 1 |
| 1192 | 3074012 | 2 | 1 | 5 | 2 | 3 | 1 | 1 | 2 | 1 | 0 | 5 | 3 | 4 | 0 | 1 | 1 | 1 | 1 | 1 | 1 | 0 | 0 | 1 | 3 | 1 |
| 1193 | 3074013 | 2 | 0 | 4 | 1 | 2 | 1 | 1 | 2 | 0 | 0 | 2 | 2 | 4 | 0 | 1 | 1 | 1 | 1 | 1 | 1 | 0 | 1 | 1 | 4 | 1 |
| 1194 | 3074014 | 2 | 0 | 3 | 2 | 2 | 2 | 1 | 2 | 0 | 0 | 2 | 2 | 4 | 1 | 1 | 1 | 1 | 1 | 1 | 1 | 0 | 1 | 1 | 4 | 1 |
| 1195 | 3074015 | 2 | 0 | 4 | 2 | 2 | 1 | 1 | 2 | 0 | 0 | 2 | 2 | 4 | 1 | 1 | 1 | 1 | 1 | 1 | 1 | 1 | 0 | 1 | 4 | 1 |
| 1196 | 3074016 | 2 | 1 | 5 | 2 | 3 | 2 | 1 | 2 | 1 | 1 | 5 | 3 | 4 | 1 | 1 | 1 | 1 | 1 | 1 | 1 | 1 | 0 | 1 | 4 | 1 |
| 1197 | 3074017 | 2 | 0 | 2 | 2 | 1 | 2 | 1 | 2 | 0 | 0 | 2 | 1 | 4 | 1 | 1 | 1 | 1 | 1 | 1 | 1 | 1 | 0 | 1 | 4 | 1 |
| 1198 | 3074018 | 2 | 1 | 2 | 2 | 1 | 1 | 3 | 2 | 0 | 0 | 2 | 1 | 4 | 1 | 1 | 1 | 1 | 1 | 1 | 1 | 1 | 0 | 1 | 4 | 1 |
| 1199 | 3074019 | 2 | 1 | 3 | 1 | 1 | 2 | 3 | 2 | 0 | 0 | 2 | 2 | 4 | 1 | 1 | 1 | 1 | 1 | 1 | 1 | 0 | 0 | 1 | 4 | 1 |
| 1200 | 3074020 | 2 | 1 | 2 | 1 | 1 | 1 | 1 | 2 | 0 | 0 | 2 | 1 | 4 | 1 | 1 | 1 | 1 | 1 | 1 | 1 | 0 | 0 | 1 | 4 | 1 |
| 1201 | 3074021 | 2 | 0 | 3 | 2 | 2 | 2 | 4 | 2 | 1 | 0 | 2 | 2 | 4 | 1 | 1 | 1 | 1 | 1 | 1 | 1 | 1 | 0 | 1 | 4 | 1 |
| 1202 | 3074022 | 2 | 0 | 2 | 2 | 1 | 2 | 3 | 2 | 0 | 0 | 2 | 1 | 4 | 1 | 1 | 1 | 1 | 1 | 1 | 1 | 0 | 0 | 1 | 4 | 1 |
| 1203 | 3074023 | 2 | 0 | 3 | 2 | 2 | 2 | 3 | 2 | 0 | 0 | 2 | 2 | 4 | 1 | 1 | 1 | 1 | 1 | 1 | 1 | 1 | 0 | 1 | 4 | 1 |
| 1204 | 3074024 | 2 | 1 | 2 | 5 | 2 | 2 | 3 | 2 | 0 | 0 | 2 | 1 | 4 | 1 | 1 | 1 | 1 | 1 | 1 | 1 | 1 | 1 | 1 | 4 | 1 |
| 1205 | 3074025 | 2 | 0 | 2 | 3 | 1 | 2 | 3 | 2 | 0 | 0 | 2 | 1 | 4 | 1 | 1 | 1 | 1 | 1 | 1 | 1 | 0 | 1 | 1 | 4 | 1 |
| 1206 | 3074026 | 2 | 0 | 4 | 2 | 2 | 1 | 3 | 2 | 0 | 0 | 2 | 2 | 4 | 0 | 1 | 1 | 1 | 1 | 1 | 1 | 0 | 0 | 1 | 3 | 1 |
| 1207 | 3074027 | 2 | 1 | 3 | 1 | 2 | 1 | 3 | 2 | 0 | 0 | 2 | 2 | 4 | 1 | 1 | 1 | 1 | 1 | 0 | 1 | 0 | 0 | 1 | 3 | 1 |
| 1208 | 3074028 | 2 | 0 | 2 | 3 | 2 | 2 | 3 | 2 | 0 | 0 | 2 | 1 | 4 | 1 | 1 | 1 | 1 | 1 | 0 | 1 | 0 | 0 | 1 | 3 | 1 |
| 1209 | 3074029 | 2 | 1 | 3 | 2 | 2 | 2 | 3 | 2 | 0 | 0 | 2 | 2 | 4 | 1 | 1 | 1 | 1 | 1 | 0 | 1 | 1 | 0 | 1 | 4 | 1 |
| 1210 | 3074030 | 2 | 1 | 3 | 3 | 1 | 1 | 1 | 2 | 0 | 0 | 2 | 2 | 4 | 1 | 1 | 1 | 1 | 1 | 1 | 1 | 0 | 0 | 1 | 4 | 1 |
| 1211 | 3074031 | 2 | 1 | 2 | 2 | 2 | 2 | 2 | 2 | 0 | 0 | 2 | 1 | 4 | 1 | 1 | 1 | 1 | 1 | 1 | 1 | 0 | 0 | 1 | 4 | 1 |
| 1212 | 3074032 | 2 | 0 | 4 | 1 | 2 | 1 | 1 | 2 | 0 | 0 | 2 | 2 | 4 | 1 | 1 | 1 | 1 | 1 | 0 | 1 | 1 | 0 | 1 | 4 | 1 |
| 1213 | 3074033 | 2 | 0 | 5 | 1 | 2 | 1 | 1 | 2 | 0 | 0 | 5 | 3 | 4 | 1 | 1 | 1 | 1 | 0 | 1 | 1 | 0 | 0 | 1 | 3 | 1 |
| 1214 | 3074034 | 2 | 1 | 4 | 4 | 1 | 1 | 1 | 2 | 0 | 0 | 2 | 2 | 4 | 0 | 1 | 1 | 1 | 0 | 1 | 1 | 0 | 0 | 1 | 3 | 1 |
| 1215 | 3074035 | 2 | 0 | 5 | 2 | 2 | 1 | 1 | 2 | 1 | 0 | 5 | 3 | 4 | 0 | 1 | 1 | 1 | 1 | 1 | 1 | 0 | 0 | 1 | 3 | 1 |
| 1216 | 307411 | 2 | 0 | 2 | 2 | 2 | 2 | 2 | 3 | 0 | 0 | 1 | 1 | 2 | 1 | 1 | 1 | 1 | 1 | 1 | 0 | 1 | 1 | 1 | 4 | 1 |
| 1217 | 307412 | 2 | 0 | 2 | 4 | 2 | 1 | 2 | 3 | 0 | 0 | 1 | 1 | 4 | 1 | 1 | 1 | 1 | 0 | 1 | 0 | 1 | 1 | 1 | 4 | 1 |
| 1218 | 307413 | 2 | 0 | 2 | 3 | 2 | 2 | 3 | 3 | 0 | 0 | 1 | 1 | 4 | 1 | 1 | 1 | 1 | 1 | 1 | 0 | 1 | 1 | 1 | 4 | 1 |
| 1219 | 307414 | 2 | 0 | 4 | 1 | 1 | 2 | 3 | 3 | 0 | 0 | 1 | 2 | 4 | 1 | 1 | 1 | 1 | 1 | 1 | 0 | 1 | 1 | 1 | 4 | 1 |
| 1220 | 307415 | 2 | 0 | 2 | 2 | 2 | 2 | 2 | 2 | 0 | 0 | 1 | 1 | 4 | 1 | 1 | 1 | 1 | 1 | 1 | 0 | 1 | 1 | 0 | 4 | 1 |
| 1221 | 307416 | 2 | 1 | 2 | 3 | 2 | 2 | 2 | 2 | 0 | 0 | 2 | 1 | 4 | 0 | 1 | 1 | 1 | 1 | 1 | 1 | 1 | 1 | 1 | 4 | 1 |

| | | | | | | | | | | | | | | | | | | | | | | | | | | |
|---|---|---|---|---|---|---|---|---|---|---|---|---|---|---|---|---|---|---|---|---|---|---|---|---|---|---|
| 1222 | 307417 | 2 | 0 | 2 | 1 | 2 | 2 | 2 | 3 | 0 | 0 | 1 | 1 | 4 | 1 | 1 | 1 | 1 | 1 | 1 | 0 | 1 | 1 | 0 | 4 | 1 |
| 1223 | 307418 | 2 | 0 | 4 | 2 | 2 | 1 | 3 | 3 | 0 | 0 | 3 | 2 | 4 | 0 | 1 | 1 | 1 | 1 | 1 | 0 | 1 | 1 | 1 | 4 | 1 |
| 1224 | 307419 | 2 | 0 | 5 | 2 | 2 | 2 | 3 | 3 | 0 | 0 | 5 | 3 | 2 | 0 | 1 | 1 | 1 | 1 | 1 | 1 | 1 | 1 | 1 | 4 | 1 |
| 1225 | 3074110 | 2 | 0 | 2 | 2 | 2 | 2 | 2 | 3 | 0 | 0 | 1 | 1 | 4 | 0 | 1 | 1 | 1 | 1 | 1 | 1 | 1 | 1 | 1 | 4 | 1 |
| 1226 | 3074111 | 2 | 1 | 3 | 1 | 2 | 2 | 2 | 3 | 0 | 0 | 2 | 2 | 4 | 0 | 1 | 1 | 1 | 1 | 1 | 0 | 1 | 1 | 1 | 4 | 1 |
| 1227 | 3074112 | 2 | 0 | 4 | 2 | 1 | 2 | 2 | 3 | 0 | 0 | 1 | 2 | 4 | 0 | 1 | 1 | 1 | 1 | 1 | 1 | 1 | 1 | 1 | 4 | 1 |
| 1228 | 3074113 | 2 | 0 | 2 | 2 | 2 | 2 | 2 | 3 | 0 | 0 | 1 | 1 | 4 | 1 | 1 | 1 | 1 | 1 | 1 | 1 | 1 | 1 | 0 | 4 | 1 |
| 1229 | 3074114 | 2 | 1 | 3 | 2 | 2 | 1 | 2 | 3 | 0 | 0 | 2 | 2 | 4 | 1 | 1 | 1 | 1 | 1 | 1 | 1 | 1 | 0 | 1 | 4 | 1 |
| 1230 | 3074115 | 2 | 1 | 3 | 3 | 2 | 1 | 2 | 3 | 0 | 0 | 4 | 2 | 4 | 1 | 1 | 1 | 1 | 1 | 1 | 1 | 1 | 0 | 1 | 4 | 1 |
| 1231 | 3074116 | 2 | 1 | 2 | 3 | 3 | 2 | 2 | 3 | 0 | 0 | 3 | 1 | 4 | 1 | 1 | 1 | 1 | 1 | 1 | 1 | 1 | 1 | 1 | 4 | 1 |
| 1232 | 3074117 | 2 | 1 | 2 | 3 | 2 | 2 | 2 | 3 | 0 | 0 | 2 | 1 | 4 | 1 | 1 | 1 | 1 | 1 | 1 | 0 | 1 | 1 | 1 | 4 | 1 |
| 1233 | 3074118 | 2 | 0 | 3 | 3 | 2 | 2 | 2 | 3 | 0 | 0 | 1 | 2 | 4 | 1 | 1 | 1 | 1 | 1 | 1 | 1 | 0 | 0 | 0 | 3 | 1 |
| 1234 | 3074119 | 2 | 0 | 5 | 4 | 2 | 1 | 2 | 3 | 0 | 0 | 5 | 3 | 4 | 1 | 1 | 1 | 1 | 1 | 1 | 1 | 1 | 0 | 1 | 4 | 1 |
| 1235 | 3074120 | 2 | 1 | 5 | 5 | 3 | 2 | 1 | 3 | 1 | 1 | 5 | 3 | 4 | 1 | 1 | 1 | 1 | 1 | 1 | 0 | 1 | 1 | 1 | 4 | 1 |
| 1236 | 3074121 | 2 | 1 | 3 | 4 | 2 | 1 | 3 | 3 | 0 | 0 | 2 | 2 | 4 | 1 | 1 | 1 | 1 | 1 | 1 | 1 | 1 | 1 | 1 | 4 | 1 |
| 1237 | 3074122 | 2 | 1 | 3 | 3 | 2 | 1 | 1 | 3 | 0 | 0 | 5 | 2 | 4 | 1 | 1 | 1 | 1 | 1 | 1 | 1 | 1 | 1 | 0 | 4 | 1 |
| 1238 | 3074123 | 2 | 0 | 3 | 4 | 2 | 1 | 1 | 3 | 0 | 0 | 1 | 2 | 4 | 1 | 1 | 1 | 1 | 0 | 1 | 1 | 0 | 0 | 0 | 3 | 1 |
| 1239 | 3074124 | 2 | 0 | 2 | 5 | 1 | 1 | 1 | 3 | 0 | 0 | 1 | 1 | 4 | 1 | 1 | 1 | 1 | 1 | 1 | 1 | 1 | 1 | 1 | 4 | 1 |
| 1240 | 3074125 | 2 | 1 | 2 | 5 | 2 | 2 | 1 | 3 | 0 | 0 | 2 | 1 | 4 | 1 | 1 | 1 | 1 | 1 | 1 | 1 | 1 | 1 | 1 | 4 | 1 |
| 1241 | 3074126 | 2 | 0 | 5 | 2 | 2 | 1 | 1 | 3 | 0 | 0 | 5 | 3 | 4 | 1 | 1 | 1 | 1 | 1 | 1 | 1 | 0 | 1 | 1 | 4 | 1 |
| 1242 | 3074127 | 2 | 0 | 2 | 5 | 2 | 2 | 1 | 3 | 0 | 0 | 1 | 1 | 4 | 1 | 1 | 1 | 1 | 1 | 1 | 0 | 1 | 1 | 1 | 4 | 1 |
| 1243 | 3074128 | 2 | 0 | 2 | 5 | 2 | 2 | 3 | 3 | 0 | 0 | 1 | 1 | 4 | 1 | 1 | 1 | 1 | 1 | 1 | 0 | 1 | 1 | 1 | 4 | 1 |
| 1244 | 3074129 | 2 | 1 | 5 | 4 | 2 | 2 | 3 | 3 | 1 | 0 | 5 | 3 | 4 | 0 | 1 | 1 | 1 | 1 | 1 | 1 | 0 | 1 | 0 | 3 | 1 |
| 1245 | 3074130 | 2 | 0 | 3 | 2 | 3 | 2 | 2 | 3 | 1 | 0 | 1 | 2 | 4 | 1 | 1 | 1 | 1 | 1 | 1 | 0 | 1 | 1 | 0 | 4 | 1 |
| 1246 | 3074131 | 2 | 1 | 2 | 5 | 2 | 2 | 2 | 3 | 0 | 0 | 2 | 1 | 4 | 0 | 1 | 1 | 1 | 1 | 1 | 1 | 0 | 1 | 1 | 4 | 1 |
| 1247 | 3074132 | 2 | 0 | 2 | 2 | 2 | 2 | 4 | 4 | 0 | 0 | 1 | 1 | 4 | 0 | 0 | 0 | 0 | 0 | 0 | 0 | 0 | 0 | 0 | 1 | 0 |
| 1248 | 3074133 | 2 | 1 | 2 | 4 | 2 | 1 | 4 | 4 | 0 | 0 | 2 | 1 | 4 | 1 | 0 | 0 | 0 | 0 | 0 | 0 | 0 | 0 | 0 | 2 | 0 |
| 1249 | 3074134 | 2 | 0 | 5 | 2 | 3 | 1 | 3 | 4 | 1 | 1 | 5 | 3 | 4 | 1 | 1 | 1 | 1 | 1 | 1 | 0 | 1 | 1 | 1 | 4 | 1 |
| 1250 | 3074135 | 2 | 1 | 2 | 5 | 2 | 1 | 3 | 4 | 0 | 0 | 2 | 1 | 4 | 0 | 1 | 1 | 1 | 0 | 1 | 0 | 1 | 1 | 1 | 3 | 1 |
| 1251 | 3074136 | 2 | 1 | 2 | 5 | 2 | 1 | 4 | 4 | 0 | 0 | 2 | 1 | 4 | 1 | 1 | 1 | 1 | 1 | 1 | 1 | 1 | 1 | 0 | 4 | 1 |
| 1252 | 3074137 | 2 | 1 | 2 | 5 | 2 | 2 | 3 | 4 | 0 | 0 | 2 | 1 | 4 | 0 | 0 | 0 | 0 | 0 | 0 | 0 | 0 | 0 | 0 | 1 | 0 |
| 1253 | 3074138 | 2 | 0 | 3 | 3 | 2 | 1 | 4 | 4 | 0 | 0 | 1 | 2 | 4 | 0 | 1 | 1 | 1 | 1 | 1 | 0 | 0 | 0 | 0 | 2 | 0 |
| 1254 | 3074139 | 2 | 1 | 2 | 3 | 2 | 1 | 3 | 4 | 0 | 0 | 4 | 1 | 4 | 0 | 0 | 0 | 0 | 0 | 0 | 0 | 0 | 0 | 0 | 1 | 0 |
| 1255 | 3074140 | 2 | 1 | 2 | 5 | 2 | 2 | 4 | 4 | 0 | 0 | 2 | 1 | 4 | 0 | 0 | 0 | 0 | 0 | 0 | 0 | 0 | 0 | 0 | 1 | 0 |
| 1256 | 308421 | 2 | 0 | 2 | 4 | 1 | 1 | 4 | 4 | 0 | 0 | 2 | 1 | 4 | 0 | 0 | 0 | 0 | 0 | 0 | 0 | 0 | 0 | 0 | 1 | 0 |
| 1257 | 308422 | 2 | 1 | 2 | 4 | 2 | 2 | 4 | 4 | 0 | 0 | 2 | 1 | 4 | 0 | 1 | 1 | 1 | 0 | 1 | 1 | 0 | 1 | 1 | 3 | 1 |
| 1258 | 308423 | 2 | 0 | 3 | 3 | 3 | 2 | 4 | 2 | 0 | 0 | 2 | 2 | 4 | 0 | 1 | 1 | 1 | 0 | 0 | 0 | 1 | 1 | 1 | 3 | 1 |
| 1259 | 308424 | 2 | 0 | 2 | 2 | 1 | 1 | 4 | 2 | 0 | 0 | 2 | 1 | 4 | 0 | 1 | 1 | 1 | 1 | 1 | 0 | 1 | 0 | 1 | 3 | 1 |
| 1260 | 308425 | 2 | 0 | 2 | 3 | 3 | 2 | 4 | 2 | 0 | 0 | 2 | 1 | 4 | 1 | 1 | 1 | 1 | 0 | 0 | 0 | 1 | 1 | 1 | 3 | 1 |
| 1261 | 308426 | 2 | 0 | 2 | 2 | 3 | 2 | 4 | 2 | 0 | 0 | 2 | 1 | 4 | 1 | 1 | 1 | 1 | 0 | 0 | 0 | 0 | 1 | 1 | 3 | 1 |
| 1262 | 308427 | 2 | 1 | 2 | 2 | 1 | 2 | 4 | 4 | 0 | 0 | 2 | 1 | 4 | 0 | 0 | 0 | 0 | 0 | 0 | 0 | 0 | 0 | 0 | 1 | 0 |
| 1263 | 308428 | 2 | 1 | 2 | 4 | 2 | 2 | 4 | 4 | 0 | 0 | 2 | 1 | 4 | 0 | 1 | 0 | 1 | 0 | 1 | 0 | 0 | 1 | 1 | 2 | 0 |
| 1264 | 308429 | 2 | 0 | 4 | 1 | 2 | 2 | 4 | 4 | 0 | 0 | 2 | 2 | 4 | 0 | 1 | 1 | 1 | 0 | 0 | 0 | 0 | 1 | 1 | 2 | 0 |
| 1265 | 3084210 | 2 | 0 | 2 | 2 | 2 | 1 | 4 | 2 | 0 | 0 | 2 | 1 | 4 | 1 | 0 | 0 | 0 | 0 | 0 | 0 | 0 | 0 | 0 | 2 | 0 |
| 1266 | 3084211 | 2 | 1 | 2 | 3 | 1 | 2 | 3 | 2 | 0 | 0 | 2 | 1 | 4 | 0 | 0 | 0 | 0 | 0 | 0 | 0 | 0 | 0 | 0 | 1 | 0 |
| 1267 | 3084212 | 2 | 0 | 1 | 3 | 2 | 1 | 1 | 2 | 0 | 0 | 2 | 1 | 4 | 0 | 0 | 0 | 0 | 0 | 0 | 0 | 0 | 0 | 0 | 1 | 0 |
| 1268 | 3084213 | 2 | 0 | 2 | 2 | 2 | 2 | 4 | 4 | 0 | 0 | 2 | 1 | 4 | 0 | 0 | 0 | 0 | 0 | 0 | 0 | 0 | 0 | 0 | 1 | 0 |
| 1269 | 3084214 | 2 | 0 | 2 | 2 | 2 | 1 | 4 | 4 | 0 | 0 | 2 | 1 | 4 | 0 | 0 | 0 | 0 | 0 | 0 | 0 | 0 | 0 | 0 | 1 | 0 |
| 1270 | 3084215 | 2 | 1 | 2 | 2 | 1 | 1 | 4 | 2 | 0 | 0 | 2 | 1 | 4 | 0 | 0 | 0 | 0 | 0 | 0 | 0 | 0 | 0 | 0 | 1 | 0 |
| 1271 | 3084216 | 2 | 1 | 2 | 3 | 2 | 2 | 4 | 2 | 0 | 0 | 2 | 1 | 4 | 0 | 0 | 0 | 0 | 0 | 0 | 0 | 0 | 0 | 0 | 1 | 0 |
| 1272 | 3084217 | 2 | 1 | 2 | 2 | 2 | 1 | 1 | 2 | 0 | 0 | 2 | 1 | 4 | 0 | 0 | 0 | 0 | 0 | 0 | 0 | 0 | 0 | 0 | 1 | 0 |
| 1273 | 3084218 | 2 | 1 | 2 | 2 | 2 | 1 | 1 | 4 | 0 | 0 | 2 | 1 | 4 | 0 | 1 | 1 | 1 | 0 | 0 | 1 | 0 | 1 | 1 | 3 | 1 |
| 1274 | 3084219 | 2 | 1 | 3 | 3 | 2 | 1 | 1 | 4 | 0 | 0 | 2 | 2 | 4 | 1 | 1 | 1 | 1 | 0 | 0 | 1 | 0 | 0 | 1 | 3 | 1 |
| 1275 | 3084220 | 2 | 1 | 4 | 4 | 3 | 2 | 1 | 4 | 1 | 1 | 5 | 2 | 4 | 1 | 0 | 0 | 0 | 0 | 0 | 0 | 0 | 0 | 0 | 2 | 0 |
| 1276 | 3084221 | 2 | 0 | 2 | 2 | 2 | 2 | 1 | 4 | 1 | 1 | 3 | 1 | 4 | 1 | 1 | 1 | 1 | 1 | 0 | 1 | 0 | 1 | 1 | 4 | 1 |
| 1277 | 3084222 | 2 | 0 | 3 | 4 | 2 | 2 | 1 | 4 | 0 | 0 | 1 | 2 | 4 | 0 | 0 | 0 | 0 | 0 | 0 | 0 | 0 | 0 | 0 | 1 | 0 |
| 1278 | 3084223 | 2 | 0 | 2 | 4 | 1 | 1 | 1 | 4 | 0 | 0 | 1 | 1 | 4 | 1 | 1 | 0 | 1 | 1 | 0 | 0 | 0 | 0 | 0 | 2 | 0 |
| 1279 | 3084224 | 2 | 0 | 3 | 2 | 2 | 2 | 1 | 4 | 0 | 0 | 1 | 2 | 4 | 0 | 0 | 0 | 0 | 0 | 0 | 0 | 0 | 0 | 0 | 1 | 0 |
| 1280 | 3084225 | 2 | 1 | 2 | 3 | 2 | 2 | 2 | 4 | 0 | 0 | 2 | 1 | 4 | 1 | 0 | 0 | 0 | 0 | 0 | 0 | 0 | 0 | 0 | 2 | 0 |
| 1281 | 3084226 | 2 | 0 | 4 | 2 | 3 | 2 | 3 | 4 | 0 | 0 | 1 | 2 | 4 | 1 | 1 | 0 | 1 | 0 | 0 | 0 | 1 | 1 | 1 | 3 | 1 |
| 1282 | 3084227 | 2 | 1 | 2 | 4 | 2 | 2 | 3 | 4 | 0 | 0 | 2 | 1 | 4 | 0 | 0 | 0 | 0 | 0 | 0 | 0 | 0 | 0 | 0 | 1 | 0 |
| 1283 | 3084228 | 2 | 1 | 1 | 5 | 2 | 1 | 2 | 4 | 0 | 0 | 2 | 1 | 4 | 1 | 0 | 0 | 0 | 0 | 0 | 0 | 0 | 0 | 0 | 2 | 0 |
| 1284 | 3084229 | 2 | 0 | 3 | 1 | 3 | 2 | 2 | 4 | 0 | 0 | 1 | 2 | 4 | 1 | 1 | 1 | 0 | 0 | 0 | 0 | 0 | 0 | 0 | 2 | 0 |
| 1285 | 3084230 | 2 | 1 | 2 | 4 | 2 | 2 | 1 | 4 | 1 | 1 | 5 | 1 | 4 | 1 | 0 | 0 | 0 | 0 | 0 | 0 | 0 | 0 | 0 | 2 | 0 |
| 1286 | 308431 | 2 | 0 | 2 | 3 | 2 | 2 | 4 | 2 | 0 | 0 | 1 | 1 | 4 | 1 | 1 | 1 | 1 | 1 | 0 | 1 | 0 | 0 | 0 | 3 | 1 |
| 1287 | 308432 | 2 | 1 | 4 | 5 | 2 | 1 | 4 | 2 | 0 | 0 | 5 | 2 | 4 | 0 | 1 | 1 | 1 | 1 | 0 | 1 | 0 | 0 | 0 | 2 | 0 |
| 1288 | 308433 | 2 | 0 | 2 | 2 | 1 | 1 | 4 | 2 | 0 | 0 | 2 | 1 | 4 | 0 | 1 | 1 | 1 | 0 | 0 | 0 | 0 | 0 | 0 | 2 | 0 |
| 1289 | 308434 | 2 | 0 | 2 | 4 | 2 | 1 | 4 | 2 | 0 | 0 | 2 | 1 | 4 | 1 | 0 | 0 | 0 | 0 | 0 | 0 | 0 | 0 | 0 | 2 | 0 |
| 1290 | 308435 | 2 | 0 | 4 | 2 | 1 | 1 | 4 | 2 | 0 | 0 | 2 | 2 | 4 | 1 | 1 | 1 | 1 | 0 | 0 | 0 | 0 | 0 | 0 | 2 | 0 |
| 1291 | 308436 | 2 | 1 | 4 | 1 | 2 | 1 | 4 | 2 | 0 | 0 | 5 | 2 | 4 | 0 | 1 | 1 | 1 | 0 | 0 | 1 | 0 | 0 | 0 | 2 | 0 |
| 1292 | 308437 | 2 | 0 | 5 | 1 | 2 | 2 | 4 | 2 | 1 | 0 | 5 | 3 | 4 | 0 | 0 | 0 | 0 | 0 | 0 | 0 | 0 | 0 | 0 | 1 | 0 |
| 1293 | 308438 | 2 | 1 | 2 | 2 | 2 | 1 | 3 | 2 | 0 | 0 | 3 | 1 | 4 | 0 | 0 | 0 | 0 | 0 | 0 | 0 | 0 | 0 | 0 | 1 | 0 |
| 1294 | 308439 | 2 | 0 | 2 | 2 | 2 | 2 | 4 | 2 | 0 | 0 | 2 | 1 | 4 | 1 | 0 | 0 | 0 | 0 | 0 | 0 | 0 | 0 | 0 | 2 | 0 |
| 1295 | 3084310 | 2 | 1 | 3 | 3 | 1 | 2 | 3 | 2 | 0 | 0 | 2 | 2 | 4 | 1 | 1 | 1 | 1 | 1 | 0 | 0 | 1 | 0 | 1 | 3 | 1 |
| 1296 | 3084311 | 2 | 0 | 2 | 2 | 2 | 2 | 4 | 2 | 0 | 0 | 2 | 1 | 4 | 0 | 1 | 1 | 1 | 0 | 0 | 0 | 0 | 0 | 0 | 2 | 0 |
| 1297 | 3084312 | 2 | 0 | 2 | 2 | 2 | 2 | 1 | 4 | 0 | 0 | 1 | 1 | 2 | 0 | 0 | 0 | 0 | 0 | 0 | 0 | 0 | 0 | 0 | 1 | 0 |
| 1298 | 3084313 | 2 | 1 | 2 | 5 | 2 | 1 | 2 | 4 | 0 | 0 | 2 | 1 | 4 | 0 | 0 | 0 | 0 | 0 | 0 | 0 | 0 | 0 | 0 | 1 | 0 |
| 1299 | 3084314 | 2 | 0 | 4 | 1 | 2 | 2 | 4 | 4 | 0 | 0 | 1 | 2 | 4 | 1 | 0 | 0 | 0 | 0 | 0 | 0 | 0 | 0 | 0 | 2 | 0 |
| 1300 | 3084315 | 2 | 1 | 4 | 4 | 3 | 2 | 2 | 4 | 0 | 0 | 3 | 2 | 4 | 1 | 0 | 0 | 0 | 0 | 0 | 0 | 0 | 0 | 0 | 2 | 0 |
| 1301 | 3084316 | 2 | 1 | 2 | 1 | 3 | 1 | 4 | 4 | 0 | 0 | 2 | 1 | 4 | 1 | 1 | 1 | 1 | 1 | 0 | 1 | 0 | 0 | 0 | 3 | 1 |
| 1302 | 3084317 | 2 | 1 | 4 | 1 | 2 | 2 | 3 | 4 | 0 | 0 | 2 | 2 | 2 | 1 | 1 | 1 | 0 | 0 | 0 | 0 | 0 | 0 | 0 | 2 | 0 |
| 1303 | 3084318 | 2 | 0 | 2 | 1 | 2 | 1 | 4 | 4 | 0 | 0 | 1 | 1 | 4 | 0 | 0 | 0 | 0 | 0 | 0 | 0 | 0 | 0 | 0 | 1 | 0 |
| 1304 | 3084319 | 2 | 0 | 2 | 2 | 2 | 2 | 4 | 4 | 0 | 0 | 1 | 1 | 4 | 1 | 0 | 0 | 0 | 0 | 0 | 0 | 0 | 0 | 0 | 2 | 0 |
| 1305 | 3084320 | 2 | 1 | 3 | 2 | 3 | 2 | 4 | 4 | 0 | 0 | 2 | 2 | 4 | 0 | 0 | 0 | 0 | 0 | 0 | 0 | 0 | 0 | 0 | 1 | 0 |
| 1306 | 3084321 | 2 | 1 | 2 | 5 | 2 | 2 | 2 | 4 | 0 | 0 | 2 | 1 | 4 | 0 | 1 | 1 | 1 | 0 | 0 | 0 | 0 | 0 | 0 | 2 | 0 |
| 1307 | 3084322 | 2 | 0 | 2 | 3 | 3 | 2 | 2 | 4 | 0 | 0 | 3 | 1 | 4 | 0 | 0 | 0 | 0 | 0 | 0 | 0 | 0 | 0 | 0 | 1 | 0 |
| 1308 | 3084323 | 2 | 0 | 2 | 3 | 2 | 2 | 3 | 4 | 0 | 0 | 2 | 1 | 4 | 1 | 0 | 0 | 0 | 0 | 0 | 0 | 0 | 0 | 0 | 2 | 0 |
| 1309 | 3084324 | 2 | 1 | 2 | 1 | 3 | 1 | 4 | 4 | 0 | 0 | 2 | 1 | 4 | 0 | 0 | 0 | 0 | 0 | 0 | 0 | 0 | 0 | 0 | 1 | 0 |
| 1310 | 3084325 | 2 | 0 | 4 | 3 | 3 | 2 | 1 | 4 | 0 | 0 | 1 | 2 | 4 | 0 | 0 | 0 | 0 | 0 | 0 | 0 | 0 | 0 | 0 | 1 | 0 |
| 1311 | 3084326 | 2 | 0 | 2 | 1 | 1 | 1 | 4 | 4 | 0 | 0 | 2 | 1 | 4 | 0 | 0 | 0 | 0 | 0 | 0 | 0 | 0 | 0 | 0 | 1 | 0 |
| 1312 | 3084327 | 2 | 0 | 2 | 1 | 1 | 1 | 4 | 4 | 0 | 0 | 2 | 1 | 4 | 1 | 0 | 0 | 0 | 0 | 0 | 0 | 0 | 0 | 0 | 2 | 0 |
| 1313 | 3084328 | 2 | 0 | 5 | 1 | 2 | 2 | 4 | 4 | 0 | 0 | 5 | 3 | 4 | 0 | 0 | 0 | 0 | 0 | 0 | 0 | 0 | 0 | 0 | 1 | 0 |
| 1314 | 3084329 | 2 | 1 | 5 | 5 | 3 | 2 | 1 | 4 | 1 | 1 | 5 | 3 | 4 | 0 | 1 | 1 | 1 | 0 | 0 | 0 | 0 | 0 | 1 | 2 | 0 |
| 1315 | 3084330 | 2 | 0 | 2 | 2 | 2 | 1 | 4 | 4 | 0 | 0 | 2 | 1 | 4 | 0 | 0 | 0 | 0 | 0 | 0 | 0 | 0 | 0 | 0 | 1 | 0 |

| | | | | | | | | | | | | | | | | | | | | | | | | | | |
|---|---|---|---|---|---|---|---|---|---|---|---|---|---|---|---|---|---|---|---|---|---|---|---|---|---|---|
| 1316 | 3084331 | 2 | 1 | 4 | 4 | 2 | 1 | 4 | 4 | 0 | 0 | 2 | 2 | 4 | 0 | 1 | 1 | 1 | 0 | 0 | 0 | 0 | 1 | 1 | 2 | 0 |
| 1317 | 3084332 | 2 | 1 | 5 | 2 | 2 | 1 | 3 | 4 | 1 | 1 | 5 | 3 | 4 | 1 | 1 | 1 | 1 | 0 | 0 | 1 | 0 | 0 | 1 | 3 | 1 |
| 1318 | 3084333 | 2 | 0 | 5 | 2 | 3 | 1 | 4 | 4 | 1 | 1 | 5 | 3 | 4 | 1 | 0 | 0 | 0 | 0 | 0 | 0 | 0 | 0 | 0 | 2 | 0 |
| 1319 | 3084334 | 2 | 1 | 5 | 4 | 3 | 2 | 4 | 4 | 1 | 1 | 5 | 3 | 4 | 1 | 0 | 0 | 0 | 0 | 0 | 0 | 0 | 0 | 0 | 2 | 0 |
| 1320 | 3084335 | 2 | 1 | 5 | 4 | 3 | 1 | 3 | 4 | 0 | 0 | 5 | 3 | 4 | 1 | 1 | 1 | 1 | 0 | 0 | 1 | 0 | 0 | 0 | 2 | 0 |
| 1321 | 3084336 | 2 | 1 | 5 | 4 | 3 | 2 | 4 | 4 | 1 | 1 | 5 | 3 | 3 | 1 | 1 | 1 | 1 | 0 | 0 | 0 | 0 | 1 | 1 | 3 | 1 |
| 1322 | 3084337 | 2 | 0 | 2 | 4 | 3 | 1 | 4 | 4 | 0 | 0 | 1 | 1 | 4 | 1 | 1 | 0 | 1 | 0 | 0 | 1 | 0 | 1 | 1 | 3 | 1 |
| 1323 | 3084338 | 2 | 1 | 4 | 3 | 3 | 2 | 3 | 4 | 1 | 0 | 3 | 2 | 4 | 1 | 0 | 0 | 0 | 0 | 0 | 0 | 0 | 0 | 0 | 2 | 0 |
| 1324 | 3084339 | 2 | 1 | 3 | 4 | 3 | 1 | 4 | 4 | 0 | 0 | 2 | 2 | 4 | 0 | 1 | 1 | 1 | 0 | 0 | 0 | 0 | 0 | 1 | 2 | 0 |
| 1325 | 308441 | 2 | 0 | 3 | 2 | 2 | 1 | 4 | 2 | 0 | 0 | 2 | 2 | 4 | 0 | 1 | 1 | 1 | 0 | 0 | 0 | 0 | 0 | 0 | 2 | 0 |
| 1326 | 308442 | 2 | 0 | 2 | 2 | 2 | 2 | 4 | 2 | 0 | 0 | 2 | 1 | 4 | 0 | 0 | 0 | 0 | 0 | 0 | 0 | 0 | 0 | 0 | 1 | 0 |
| 1327 | 308443 | 2 | 0 | 2 | 2 | 2 | 2 | 1 | 2 | 0 | 0 | 2 | 1 | 4 | 1 | 0 | 0 | 0 | 0 | 0 | 0 | 0 | 0 | 0 | 2 | 0 |
| 1328 | 308444 | 2 | 0 | 2 | 5 | 2 | 1 | 1 | 2 | 0 | 0 | 2 | 1 | 4 | 1 | 1 | 0 | 1 | 0 | 0 | 0 | 0 | 0 | 0 | 2 | 0 |
| 1329 | 308445 | 2 | 0 | 2 | 5 | 2 | 1 | 1 | 2 | 0 | 0 | 2 | 1 | 4 | 0 | 1 | 0 | 1 | 0 | 0 | 0 | 0 | 0 | 0 | 2 | 0 |
| 1330 | 308446 | 2 | 1 | 2 | 4 | 2 | 2 | 1 | 2 | 0 | 0 | 2 | 1 | 4 | 0 | 1 | 0 | 1 | 0 | 0 | 0 | 1 | 0 | 0 | 2 | 0 |
| 1331 | 308447 | 2 | 1 | 2 | 5 | 2 | 1 | 3 | 2 | 1 | 1 | 2 | 1 | 4 | 0 | 0 | 0 | 0 | 0 | 0 | 0 | 0 | 0 | 0 | 1 | 0 |
| 1332 | 308448 | 2 | 1 | 2 | 5 | 2 | 2 | 4 | 2 | 0 | 0 | 2 | 1 | 4 | 0 | 0 | 0 | 0 | 0 | 0 | 0 | 0 | 0 | 0 | 1 | 0 |
| 1333 | 308449 | 2 | 0 | 2 | 1 | 1 | 1 | 4 | 2 | 0 | 0 | 2 | 1 | 4 | 0 | 1 | 0 | 1 | 0 | 0 | 0 | 0 | 0 | 0 | 2 | 0 |
| 1334 | 3084410 | 2 | 1 | 3 | 3 | 2 | 1 | 4 | 2 | 0 | 0 | 2 | 2 | 4 | 0 | 0 | 0 | 0 | 0 | 0 | 0 | 0 | 0 | 0 | 1 | 0 |
| 1335 | 3084411 | 2 | 1 | 1 | 5 | 2 | 2 | 2 | 2 | 0 | 0 | 2 | 1 | 4 | 0 | 0 | 0 | 0 | 0 | 0 | 0 | 0 | 0 | 0 | 1 | 0 |
| 1336 | 3084412 | 2 | 1 | 4 | 3 | 3 | 1 | 1 | 2 | 1 | 0 | 3 | 2 | 4 | 0 | 1 | 0 | 1 | 0 | 0 | 0 | 0 | 0 | 0 | 2 | 0 |
| 1337 | 3084413 | 2 | 1 | 2 | 5 | 2 | 1 | 1 | 2 | 0 | 0 | 2 | 1 | 4 | 0 | 1 | 1 | 1 | 0 | 1 | 0 | 1 | 0 | 0 | 2 | 0 |
| 1338 | 3084414 | 2 | 1 | 4 | 3 | 3 | 2 | 1 | 2 | 0 | 0 | 5 | 2 | 4 | 1 | 1 | 0 | 1 | 0 | 0 | 1 | 0 | 0 | 0 | 2 | 0 |
| 1339 | 3084415 | 2 | 1 | 2 | 1 | 3 | 2 | 1 | 2 | 0 | 0 | 4 | 1 | 4 | 0 | 1 | 0 | 1 | 0 | 0 | 0 | 1 | 0 | 0 | 2 | 0 |
| 1340 | 3084416 | 2 | 0 | 2 | 3 | 2 | 1 | 1 | 2 | 0 | 0 | 2 | 1 | 4 | 0 | 0 | 0 | 0 | 0 | 0 | 0 | 0 | 0 | 0 | 1 | 0 |
| 1341 | 3084417 | 2 | 0 | 5 | 1 | 3 | 2 | 3 | 2 | 0 | 0 | 5 | 3 | 4 | 0 | 1 | 1 | 1 | 0 | 1 | 0 | 0 | 0 | 0 | 2 | 0 |
| 1342 | 3084418 | 2 | 1 | 3 | 2 | 3 | 1 | 3 | 2 | 1 | 1 | 2 | 2 | 4 | 1 | 0 | 0 | 0 | 0 | 0 | 0 | 0 | 0 | 0 | 2 | 0 |
| 1343 | 3084419 | 2 | 1 | 4 | 1 | 2 | 1 | 3 | 2 | 0 | 0 | 2 | 2 | 4 | 0 | 1 | 1 | 1 | 0 | 0 | 0 | 0 | 0 | 0 | 2 | 0 |
| 1344 | 3084420 | 2 | 1 | 2 | 5 | 1 | 2 | 1 | 2 | 0 | 0 | 2 | 1 | 4 | 0 | 0 | 0 | 0 | 0 | 0 | 0 | 0 | 0 | 0 | 1 | 0 |
| 1345 | 3084421 | 2 | 0 | 3 | 2 | 3 | 1 | 4 | 2 | 0 | 0 | 1 | 2 | 4 | 1 | 0 | 0 | 0 | 0 | 0 | 0 | 0 | 0 | 0 | 2 | 0 |
| 1346 | 3084422 | 2 | 0 | 2 | 2 | 2 | 2 | 4 | 2 | 0 | 0 | 2 | 1 | 4 | 1 | 0 | 0 | 0 | 0 | 0 | 0 | 0 | 0 | 0 | 2 | 0 |
| 1347 | 3084423 | 2 | 1 | 5 | 2 | 3 | 1 | 4 | 2 | 0 | 0 | 5 | 3 | 4 | 1 | 1 | 1 | 1 | 0 | 0 | 0 | 0 | 0 | 0 | 2 | 0 |
| 1348 | 3084424 | 2 | 1 | 5 | 1 | 3 | 1 | 4 | 2 | 0 | 0 | 5 | 3 | 4 | 1 | 1 | 1 | 1 | 0 | 1 | 0 | 0 | 0 | 0 | 2 | 0 |
| 1349 | 3084425 | 2 | 1 | 4 | 2 | 3 | 2 | 4 | 2 | 1 | 0 | 5 | 2 | 4 | 0 | 0 | 0 | 0 | 0 | 0 | 0 | 0 | 0 | 0 | 1 | 0 |
| 1350 | 3084426 | 2 | 0 | 5 | 1 | 3 | 1 | 3 | 2 | 1 | 0 | 5 | 3 | 4 | 1 | 1 | 1 | 1 | 0 | 0 | 0 | 0 | 0 | 1 | 2 | 0 |
| 1351 | 308451 | 2 | 0 | 1 | 4 | 2 | 2 | 2 | 3 | 0 | 0 | 2 | 1 | 4 | 1 | 1 | 0 | 1 | 1 | 0 | 1 | 0 | 0 | 1 | 3 | 1 |
| 1352 | 308452 | 2 | 1 | 2 | 2 | 1 | 2 | 2 | 3 | 0 | 0 | 2 | 1 | 4 | 1 | 1 | 1 | 1 | 1 | 0 | 0 | 0 | 0 | 1 | 3 | 1 |
| 1353 | 308453 | 2 | 0 | 2 | 3 | 2 | 1 | 2 | 3 | 0 | 0 | 2 | 1 | 4 | 1 | 0 | 0 | 0 | 0 | 0 | 0 | 0 | 0 | 0 | 2 | 0 |
| 1354 | 308454 | 2 | 0 | 2 | 2 | 1 | 2 | 3 | 3 | 0 | 0 | 2 | 1 | 4 | 1 | 0 | 0 | 0 | 0 | 0 | 0 | 0 | 0 | 0 | 2 | 0 |
| 1355 | 308455 | 2 | 0 | 1 | 4 | 1 | 1 | 2 | 3 | 0 | 0 | 2 | 1 | 4 | 1 | 1 | 1 | 0 | 1 | 0 | 0 | 0 | 0 | 0 | 2 | 0 |
| 1356 | 308456 | 2 | 1 | 2 | 2 | 2 | 2 | 2 | 1 | 0 | 0 | 2 | 1 | 4 | 1 | 1 | 1 | 1 | 1 | 0 | 0 | 0 | 0 | 1 | 3 | 1 |
| 1357 | 308457 | 2 | 1 | 2 | 2 | 1 | 2 | 3 | 3 | 0 | 0 | 2 | 1 | 4 | 1 | 1 | 1 | 1 | 1 | 0 | 0 | 0 | 0 | 1 | 3 | 1 |
| 1358 | 308458 | 2 | 0 | 2 | 3 | 2 | 1 | 3 | 3 | 0 | 0 | 2 | 1 | 4 | 0 | 0 | 0 | 0 | 0 | 0 | 0 | 0 | 0 | 0 | 1 | 0 |
| 1359 | 308459 | 2 | 0 | 1 | 2 | 1 | 1 | 2 | 3 | 0 | 0 | 2 | 1 | 4 | 1 | 1 | 1 | 1 | 1 | 0 | 0 | 0 | 0 | 1 | 3 | 1 |
| 1360 | 3084510 | 2 | 0 | 2 | 2 | 2 | 1 | 2 | 3 | 0 | 0 | 2 | 1 | 4 | 1 | 1 | 1 | 1 | 1 | 0 | 0 | 0 | 0 | 1 | 3 | 1 |
| 1361 | 3084511 | 2 | 1 | 1 | 4 | 1 | 2 | 2 | 3 | 0 | 0 | 2 | 1 | 4 | 1 | 1 | 1 | 1 | 0 | 0 | 1 | 0 | 0 | 1 | 3 | 1 |
| 1362 | 3084512 | 2 | 0 | 2 | 2 | 1 | 2 | 2 | 3 | 0 | 0 | 2 | 1 | 4 | 1 | 1 | 1 | 1 | 0 | 1 | 0 | 0 | 0 | 1 | 3 | 1 |
| 1363 | 3084513 | 2 | 0 | 2 | 1 | 1 | 1 | 2 | 3 | 0 | 0 | 2 | 1 | 4 | 1 | 1 | 1 | 1 | 1 | 1 | 1 | 0 | 0 | 1 | 4 | 1 |
| 1364 | 3084514 | 2 | 0 | 2 | 1 | 1 | 1 | 2 | 3 | 0 | 0 | 2 | 1 | 4 | 1 | 1 | 1 | 1 | 0 | 0 | 0 | 0 | 1 | 1 | 3 | 1 |
| 1365 | 3084515 | 2 | 1 | 1 | 4 | 1 | 1 | 2 | 3 | 0 | 0 | 2 | 1 | 4 | 1 | 1 | 1 | 1 | 1 | 0 | 0 | 0 | 0 | 1 | 3 | 1 |
| 1366 | 3084516 | 2 | 1 | 2 | 4 | 1 | 2 | 2 | 3 | 0 | 0 | 2 | 1 | 4 | 1 | 1 | 0 | 1 | 1 | 0 | 0 | 0 | 0 | 0 | 2 | 0 |
| 1367 | 3084517 | 2 | 0 | 2 | 4 | 1 | 2 | 2 | 3 | 0 | 0 | 2 | 1 | 4 | 0 | 1 | 0 | 1 | 1 | 0 | 0 | 0 | 0 | 1 | 2 | 0 |
| 1368 | 3084518 | 2 | 1 | 2 | 3 | 1 | 2 | 2 | 3 | 0 | 0 | 2 | 1 | 4 | 1 | 1 | 1 | 1 | 1 | 0 | 0 | 0 | 0 | 0 | 2 | 0 |
| 1369 | 3084519 | 2 | 0 | 1 | 4 | 1 | 1 | 2 | 3 | 0 | 0 | 2 | 1 | 4 | 1 | 1 | 1 | 1 | 0 | 0 | 0 | 1 | 0 | 0 | 2 | 0 |
| 1370 | 3084520 | 2 | 0 | 2 | 5 | 1 | 1 | 2 | 3 | 0 | 0 | 2 | 1 | 4 | 1 | 1 | 1 | 1 | 1 | 1 | 0 | 0 | 0 | 1 | 3 | 1 |
| 1371 | 3084521 | 2 | 0 | 2 | 4 | 1 | 2 | 2 | 3 | 0 | 0 | 2 | 1 | 4 | 1 | 1 | 1 | 1 | 1 | 0 | 0 | 0 | 0 | 1 | 3 | 1 |
| 1372 | 3084522 | 2 | 0 | 2 | 2 | 2 | 2 | 2 | 3 | 0 | 0 | 2 | 1 | 4 | 1 | 1 | 1 | 0 | 1 | 0 | 0 | 0 | 0 | 0 | 2 | 0 |
| 1373 | 3084523 | 2 | 0 | 2 | 1 | 1 | 1 | 2 | 3 | 0 | 0 | 2 | 1 | 4 | 1 | 1 | 1 | 1 | 1 | 1 | 0 | 0 | 0 | 1 | 3 | 1 |
| 1374 | 3084524 | 2 | 1 | 2 | 4 | 1 | 2 | 2 | 3 | 0 | 0 | 2 | 1 | 4 | 1 | 1 | 0 | 1 | 0 | 0 | 1 | 0 | 0 | 0 | 2 | 0 |
| 1375 | 3084525 | 2 | 1 | 2 | 5 | 1 | 1 | 2 | 3 | 0 | 0 | 2 | 1 | 4 | 1 | 1 | 0 | 1 | 1 | 1 | 1 | 0 | 0 | 1 | 3 | 1 |
| 1376 | 308461 | 2 | 0 | 3 | 4 | 3 | 2 | 2 | 3 | 1 | 1 | 2 | 2 | 4 | 1 | 1 | 1 | 1 | 1 | 1 | 1 | 0 | 1 | 1 | 4 | 1 |
| 1377 | 308462 | 2 | 1 | 1 | 3 | 1 | 1 | 2 | 3 | 0 | 0 | 2 | 1 | 4 | 1 | 1 | 1 | 1 | 1 | 1 | 1 | 0 | 0 | 0 | 3 | 1 |
| 1378 | 308463 | 2 | 1 | 2 | 2 | 2 | 1 | 2 | 3 | 0 | 0 | 2 | 1 | 4 | 1 | 1 | 1 | 0 | 0 | 0 | 0 | 0 | 0 | 0 | 2 | 0 |
| 1379 | 308464 | 2 | 1 | 1 | 4 | 1 | 2 | 2 | 3 | 0 | 0 | 2 | 1 | 4 | 1 | 1 | 1 | 0 | 1 | 0 | 0 | 0 | 0 | 0 | 2 | 0 |
| 1380 | 308465 | 2 | 0 | 2 | 5 | 1 | 1 | 2 | 3 | 0 | 0 | 2 | 1 | 4 | 1 | 1 | 1 | 1 | 0 | 0 | 0 | 0 | 0 | 0 | 2 | 0 |
| 1381 | 308466 | 2 | 1 | 2 | 3 | 1 | 1 | 2 | 3 | 0 | 0 | 2 | 1 | 4 | 0 | 1 | 1 | 0 | 1 | 0 | 0 | 0 | 0 | 0 | 2 | 0 |
| 1382 | 308467 | 2 | 0 | 2 | 4 | 2 | 1 | 2 | 3 | 0 | 0 | 2 | 1 | 4 | 1 | 1 | 1 | 1 | 0 | 1 | 0 | 0 | 0 | 0 | 2 | 0 |
| 1383 | 308468 | 2 | 0 | 2 | 3 | 2 | 2 | 2 | 3 | 0 | 0 | 2 | 1 | 4 | 1 | 1 | 0 | 0 | 0 | 0 | 0 | 0 | 0 | 0 | 2 | 0 |
| 1384 | 308469 | 2 | 0 | 2 | 5 | 1 | 1 | 2 | 3 | 0 | 0 | 2 | 1 | 4 | 1 | 1 | 1 | 0 | 0 | 0 | 0 | 0 | 0 | 0 | 2 | 0 |
| 1385 | 3084610 | 2 | 1 | 2 | 2 | 1 | 1 | 2 | 3 | 0 | 0 | 2 | 1 | 4 | 1 | 1 | 0 | 0 | 1 | 0 | 0 | 0 | 0 | 0 | 2 | 0 |
| 1386 | 3084611 | 2 | 1 | 2 | 2 | 1 | 2 | 2 | 3 | 0 | 0 | 2 | 1 | 4 | 1 | 1 | 1 | 0 | 1 | 0 | 0 | 0 | 0 | 0 | 2 | 0 |
| 1387 | 3084612 | 2 | 0 | 2 | 3 | 2 | 1 | 2 | 3 | 0 | 0 | 2 | 1 | 4 | 1 | 1 | 1 | 0 | 1 | 0 | 0 | 0 | 0 | 0 | 2 | 0 |
| 1388 | 3084613 | 2 | 0 | 4 | 3 | 2 | 1 | 2 | 3 | 0 | 0 | 2 | 2 | 4 | 1 | 1 | 1 | 1 | 1 | 0 | 0 | 0 | 0 | 1 | 3 | 1 |
| 1389 | 3084614 | 2 | 1 | 2 | 5 | 1 | 1 | 2 | 3 | 0 | 0 | 2 | 1 | 4 | 1 | 1 | 1 | 1 | 1 | 0 | 0 | 0 | 0 | 0 | 2 | 0 |
| 1390 | 3084615 | 2 | 1 | 2 | 5 | 2 | 1 | 2 | 3 | 0 | 0 | 2 | 1 | 4 | 1 | 1 | 1 | 0 | 1 | 1 | 0 | 0 | 0 | 0 | 2 | 0 |
| 1391 | 3084616 | 2 | 1 | 4 | 4 | 1 | 1 | 2 | 3 | 0 | 0 | 2 | 2 | 4 | 1 | 1 | 1 | 1 | 1 | 1 | 0 | 0 | 0 | 1 | 3 | 1 |
| 1392 | 3084617 | 2 | 0 | 2 | 4 | 1 | 2 | 2 | 3 | 0 | 0 | 2 | 1 | 4 | 1 | 1 | 1 | 0 | 1 | 1 | 0 | 0 | 0 | 0 | 2 | 0 |
| 1393 | 3084618 | 2 | 1 | 2 | 4 | 1 | 2 | 2 | 3 | 0 | 0 | 2 | 1 | 4 | 1 | 1 | 1 | 0 | 0 | 1 | 0 | 0 | 0 | 0 | 2 | 0 |
| 1394 | 3084619 | 2 | 1 | 2 | 4 | 1 | 2 | 2 | 3 | 0 | 0 | 2 | 1 | 4 | 1 | 1 | 1 | 0 | 0 | 1 | 0 | 0 | 0 | 0 | 2 | 0 |
| 1395 | 3084620 | 2 | 1 | 2 | 3 | 1 | 1 | 2 | 3 | 0 | 0 | 2 | 1 | 4 | 1 | 1 | 1 | 0 | 1 | 1 | 0 | 0 | 0 | 0 | 2 | 0 |
| 1396 | 3084621 | 2 | 0 | 1 | 5 | 1 | 2 | 2 | 3 | 0 | 0 | 2 | 1 | 4 | 1 | 1 | 1 | 0 | 0 | 1 | 0 | 0 | 0 | 0 | 2 | 0 |
| 1397 | 3084622 | 2 | 1 | 2 | 3 | 1 | 1 | 2 | 3 | 0 | 0 | 2 | 1 | 4 | 1 | 1 | 1 | 0 | 1 | 1 | 0 | 0 | 0 | 0 | 2 | 0 |
| 1398 | 3084623 | 2 | 1 | 2 | 4 | 1 | 1 | 2 | 3 | 0 | 0 | 2 | 1 | 4 | 1 | 1 | 1 | 0 | 1 | 1 | 0 | 0 | 0 | 0 | 2 | 0 |
| 1399 | 3084624 | 2 | 1 | 2 | 3 | 1 | 2 | 2 | 3 | 0 | 0 | 2 | 1 | 4 | 1 | 1 | 1 | 1 | 1 | 1 | 0 | 0 | 0 | 1 | 3 | 1 |
| 1400 | 3084625 | 2 | 0 | 2 | 4 | 1 | 1 | 2 | 3 | 0 | 0 | 2 | 1 | 4 | 1 | 1 | 1 | 0 | 1 | 1 | 0 | 0 | 0 | 0 | 2 | 0 |
| 1401 | 309471 | 2 | 1 | 4 | 4 | 2 | 2 | 2 | 4 | 1 | 1 | 2 | 2 | 4 | 1 | 1 | 1 | 1 | 1 | 1 | 1 | 1 | 1 | 1 | 4 | 1 |
| 1402 | 309472 | 2 | 0 | 3 | 3 | 1 | 1 | 1 | 4 | 0 | 0 | 2 | 2 | 4 | 0 | 1 | 1 | 1 | 1 | 1 | 1 | 1 | 0 | 1 | 4 | 1 |
| 1403 | 309473 | 2 | 1 | 2 | 3 | 1 | 2 | 2 | 4 | 0 | 0 | 2 | 1 | 4 | 1 | 1 | 1 | 1 | 0 | 1 | 1 | 0 | 0 | 1 | 3 | 1 |
| 1404 | 309474 | 2 | 1 | 2 | 4 | 1 | 2 | 1 | 4 | 1 | 0 | 2 | 1 | 4 | 1 | 1 | 1 | 1 | 0 | 1 | 1 | 0 | 0 | 1 | 3 | 1 |
| 1405 | 309475 | 2 | 0 | 2 | 2 | 1 | 1 | 1 | 4 | 1 | 1 | 2 | 1 | 4 | 1 | 1 | 1 | 1 | 0 | 1 | 0 | 1 | 1 | 1 | 4 | 1 |
| 1406 | 309476 | 2 | 0 | 4 | 3 | 3 | 2 | 4 | 4 | 1 | 1 | 1 | 2 | 4 | 1 | 1 | 1 | 1 | 0 | 1 | 1 | 1 | 1 | 1 | 4 | 1 |
| 1407 | 309477 | 2 | 1 | 2 | 2 | 2 | 2 | 1 | 4 | 0 | 0 | 2 | 1 | 4 | 0 | 1 | 1 | 1 | 1 | 1 | 0 | 1 | 0 | 1 | 3 | 1 |
| 1408 | 309478 | 2 | 1 | 4 | 4 | 1 | 2 | 1 | 4 | 1 | 0 | 2 | 2 | 4 | 1 | 1 | 1 | 1 | 1 | 1 | 1 | 1 | 1 | 1 | 4 | 1 |
| 1409 | 309479 | 2 | 1 | 2 | 5 | 1 | 1 | 4 | 4 | 1 | 0 | 2 | 1 | 4 | 0 | 1 | 1 | 1 | 0 | 1 | 1 | 1 | 0 | 1 | 3 | 1 |

| | | | | | | | | | | | | | | | | | | | | | | | | | | |
|---|---|---|---|---|---|---|---|---|---|---|---|---|---|---|---|---|---|---|---|---|---|---|---|---|---|---|
| 1410 | 3094710 | 2 | 1 | 2 | 2 | 3 | 1 | 4 | 4 | 0 | 0 | 2 | 1 | 4 | 0 | 1 | 1 | 1 | 0 | 0 | 1 | 1 | 0 | 0 | 2 | 0 |
| 1411 | 3094711 | 2 | 0 | 2 | 2 | 1 | 1 | 4 | 4 | 0 | 0 | 2 | 1 | 4 | 0 | 1 | 1 | 1 | 1 | 1 | 1 | 1 | 0 | 1 | 4 | 1 |
| 1412 | 3094712 | 2 | 0 | 5 | 2 | 2 | 2 | 3 | 4 | 0 | 0 | 5 | 3 | 4 | 1 | 1 | 1 | 1 | 1 | 0 | 1 | 1 | 0 | 1 | 4 | 1 |
| 1413 | 3094713 | 2 | 1 | 2 | 1 | 2 | 2 | 4 | 4 | 0 | 0 | 2 | 1 | 4 | 0 | 1 | 1 | 1 | 0 | 0 | 0 | 0 | 0 | 0 | 2 | 0 |
| 1414 | 3094714 | 2 | 1 | 2 | 1 | 1 | 1 | 4 | 4 | 0 | 0 | 2 | 1 | 4 | 0 | 1 | 1 | 1 | 0 | 0 | 0 | 0 | 0 | 0 | 2 | 0 |
| 1415 | 3094715 | 2 | 1 | 2 | 4 | 1 | 1 | 4 | 4 | 0 | 0 | 2 | 1 | 4 | 1 | 1 | 1 | 1 | 0 | 0 | 0 | 1 | 0 | 1 | 3 | 1 |
| 1416 | 3094716 | 2 | 1 | 2 | 2 | 2 | 2 | 4 | 4 | 0 | 0 | 2 | 1 | 4 | 0 | 1 | 1 | 1 | 0 | 0 | 1 | 1 | 0 | 1 | 3 | 1 |
| 1417 | 3094717 | 2 | 1 | 5 | 2 | 3 | 1 | 1 | 4 | 1 | 0 | 5 | 3 | 4 | 1 | 1 | 1 | 1 | 1 | 1 | 1 | 1 | 0 | 1 | 4 | 1 |
| 1418 | 3094718 | 2 | 1 | 2 | 2 | 1 | 2 | 2 | 4 | 0 | 0 | 2 | 1 | 4 | 0 | 1 | 1 | 1 | 0 | 1 | 1 | 1 | 0 | 1 | 3 | 1 |
| 1419 | 3094719 | 2 | 0 | 3 | 2 | 2 | 2 | 2 | 4 | 0 | 0 | 2 | 2 | 4 | 0 | 1 | 1 | 1 | 1 | 0 | 1 | 1 | 0 | 1 | 3 | 1 |
| 1420 | 3094720 | 2 | 0 | 3 | 4 | 1 | 1 | 2 | 4 | 0 | 0 | 2 | 2 | 4 | 0 | 1 | 1 | 1 | 1 | 1 | 1 | 1 | 0 | 1 | 4 | 1 |
| 1421 | 3094721 | 2 | 1 | 2 | 3 | 1 | 2 | 2 | 4 | 0 | 0 | 2 | 1 | 4 | 0 | 1 | 1 | 1 | 1 | 1 | 1 | 1 | 0 | 1 | 4 | 1 |
| 1422 | 3094722 | 2 | 0 | 2 | 3 | 1 | 2 | 2 | 4 | 0 | 0 | 2 | 1 | 4 | 0 | 1 | 1 | 1 | 1 | 1 | 1 | 1 | 0 | 1 | 4 | 1 |
| 1423 | 3094723 | 2 | 1 | 3 | 3 | 1 | 2 | 2 | 4 | 0 | 0 | 2 | 2 | 4 | 0 | 1 | 1 | 1 | 1 | 1 | 1 | 1 | 0 | 1 | 4 | 1 |
| 1424 | 3094724 | 2 | 0 | 5 | 1 | 2 | 2 | 1 | 4 | 0 | 0 | 5 | 3 | 4 | 0 | 1 | 1 | 1 | 1 | 1 | 1 | 1 | 0 | 1 | 4 | 1 |
| 1425 | 3094725 | 2 | 1 | 2 | 5 | 1 | 1 | 1 | 4 | 0 | 0 | 2 | 1 | 4 | 0 | 1 | 1 | 1 | 0 | 1 | 1 | 1 | 0 | 0 | 3 | 1 |
| 1426 | 3094726 | 2 | 0 | 2 | 4 | 1 | 1 | 1 | 4 | 0 | 0 | 2 | 1 | 4 | 0 | 1 | 1 | 1 | 0 | 0 | 1 | 1 | 0 | 1 | 3 | 1 |
| 1427 | 3094727 | 2 | 1 | 3 | 3 | 1 | 2 | 1 | 4 | 0 | 0 | 2 | 2 | 4 | 0 | 1 | 1 | 1 | 1 | 1 | 1 | 1 | 0 | 1 | 4 | 1 |
| 1428 | 3094728 | 2 | 0 | 4 | 1 | 2 | 1 | 1 | 4 | 0 | 0 | 2 | 2 | 4 | 0 | 1 | 1 | 1 | 1 | 1 | 1 | 1 | 0 | 1 | 4 | 1 |
| 1429 | 3094729 | 2 | 0 | 4 | 3 | 1 | 1 | 2 | 4 | 0 | 0 | 2 | 2 | 4 | 0 | 1 | 1 | 1 | 1 | 1 | 0 | 1 | 0 | 1 | 3 | 1 |
| 1430 | 3094730 | 2 | 1 | 3 | 3 | 1 | 2 | 2 | 4 | 0 | 0 | 2 | 2 | 4 | 0 | 1 | 1 | 1 | 1 | 1 | 1 | 1 | 0 | 1 | 4 | 1 |
| 1431 | 3094731 | 2 | 0 | 2 | 3 | 1 | 2 | 2 | 4 | 0 | 0 | 2 | 1 | 4 | 0 | 1 | 1 | 1 | 1 | 1 | 1 | 1 | 0 | 1 | 4 | 1 |
| 1432 | 3094732 | 2 | 1 | 2 | 2 | 1 | 1 | 2 | 4 | 0 | 0 | 2 | 1 | 4 | 0 | 1 | 1 | 1 | 1 | 1 | 1 | 1 | 0 | 1 | 4 | 1 |
| 1433 | 3094733 | 2 | 0 | 2 | 2 | 2 | 2 | 1 | 4 | 0 | 0 | 2 | 1 | 4 | 0 | 1 | 1 | 1 | 1 | 1 | 1 | 1 | 0 | 1 | 4 | 1 |
| 1434 | 3094734 | 2 | 0 | 4 | 2 | 3 | 2 | 1 | 4 | 0 | 0 | 2 | 2 | 4 | 0 | 1 | 1 | 1 | 1 | 1 | 1 | 1 | 0 | 1 | 4 | 1 |
| 1435 | 3094735 | 2 | 0 | 4 | 3 | 1 | 1 | 2 | 4 | 0 | 0 | 2 | 2 | 4 | 0 | 1 | 1 | 1 | 0 | 0 | 1 | 1 | 0 | 1 | 3 | 1 |
| 1436 | 3094736 | 2 | 0 | 4 | 3 | 1 | 1 | 1 | 4 | 0 | 0 | 2 | 2 | 4 | 0 | 1 | 1 | 1 | 0 | 1 | 0 | 1 | 0 | 1 | 3 | 1 |
| 1437 | 3094737 | 2 | 0 | 3 | 4 | 2 | 1 | 4 | 4 | 0 | 0 | 2 | 2 | 4 | 0 | 1 | 1 | 1 | 0 | 1 | 1 | 1 | 0 | 1 | 3 | 1 |
| 1438 | 3094738 | 2 | 0 | 3 | 3 | 1 | 1 | 2 | 4 | 0 | 0 | 2 | 2 | 4 | 0 | 1 | 1 | 1 | 0 | 1 | 1 | 1 | 0 | 1 | 3 | 1 |
| 1439 | 3094739 | 2 | 0 | 2 | 3 | 1 | 2 | 2 | 4 | 0 | 0 | 2 | 1 | 4 | 0 | 1 | 1 | 1 | 0 | 0 | 1 | 1 | 0 | 1 | 3 | 1 |
| 1440 | 3094740 | 2 | 0 | 4 | 3 | 2 | 1 | 2 | 4 | 1 | 0 | 2 | 2 | 4 | 1 | 1 | 1 | 1 | 1 | 1 | 1 | 1 | 0 | 1 | 4 | 1 |
| 1441 | 309481 | 2 | 1 | 3 | 2 | 1 | 1 | 1 | 2 | 0 | 0 | 2 | 2 | 4 | 0 | 1 | 1 | 1 | 0 | 1 | 1 | 1 | 0 | 1 | 3 | 1 |
| 1442 | 309482 | 2 | 1 | 3 | 2 | 1 | 1 | 4 | 2 | 0 | 0 | 2 | 2 | 4 | 0 | 1 | 1 | 1 | 1 | 1 | 1 | 1 | 0 | 1 | 4 | 1 |
| 1443 | 309483 | 2 | 0 | 5 | 2 | 1 | 2 | 1 | 2 | 0 | 0 | 5 | 3 | 4 | 0 | 1 | 1 | 1 | 0 | 1 | 1 | 1 | 0 | 1 | 3 | 1 |
| 1444 | 309484 | 2 | 0 | 2 | 5 | 1 | 2 | 1 | 2 | 0 | 0 | 2 | 1 | 4 | 0 | 1 | 1 | 1 | 1 | 1 | 1 | 1 | 0 | 1 | 4 | 1 |
| 1445 | 309485 | 2 | 0 | 4 | 2 | 2 | 2 | 1 | 2 | 0 | 0 | 2 | 2 | 4 | 0 | 1 | 1 | 1 | 0 | 1 | 1 | 1 | 0 | 1 | 3 | 1 |
| 1446 | 309486 | 2 | 0 | 2 | 3 | 2 | 2 | 1 | 2 | 0 | 0 | 2 | 1 | 4 | 1 | 1 | 1 | 1 | 0 | 1 | 1 | 1 | 0 | 1 | 4 | 1 |
| 1447 | 309487 | 2 | 0 | 2 | 3 | 1 | 2 | 1 | 2 | 0 | 0 | 2 | 1 | 4 | 0 | 1 | 1 | 1 | 1 | 1 | 1 | 1 | 0 | 1 | 4 | 1 |
| 1448 | 309488 | 2 | 0 | 2 | 4 | 1 | 2 | 1 | 2 | 0 | 0 | 2 | 1 | 4 | 1 | 1 | 1 | 1 | 1 | 1 | 1 | 1 | 0 | 1 | 4 | 1 |
| 1449 | 309489 | 2 | 0 | 2 | 3 | 2 | 1 | 1 | 2 | 0 | 0 | 2 | 1 | 4 | 0 | 1 | 1 | 1 | 0 | 1 | 1 | 1 | 0 | 1 | 3 | 1 |
| 1450 | 3094810 | 2 | 0 | 2 | 3 | 1 | 1 | 2 | 2 | 0 | 0 | 2 | 1 | 4 | 0 | 1 | 1 | 1 | 1 | 1 | 1 | 1 | 0 | 1 | 4 | 1 |
| 1451 | 3094811 | 2 | 0 | 2 | 4 | 1 | 2 | 2 | 2 | 0 | 0 | 2 | 1 | 4 | 0 | 1 | 1 | 1 | 1 | 1 | 0 | 1 | 0 | 1 | 3 | 1 |
| 1452 | 3094812 | 2 | 1 | 2 | 5 | 1 | 2 | 2 | 2 | 0 | 0 | 2 | 1 | 4 | 0 | 1 | 1 | 1 | 1 | 0 | 1 | 1 | 0 | 1 | 3 | 1 |
| 1453 | 3094813 | 2 | 1 | 2 | 3 | 1 | 1 | 1 | 2 | 0 | 0 | 2 | 1 | 4 | 1 | 1 | 1 | 1 | 1 | 1 | 1 | 1 | 0 | 1 | 4 | 1 |
| 1454 | 3094814 | 2 | 1 | 2 | 2 | 2 | 1 | 1 | 2 | 0 | 0 | 2 | 1 | 4 | 0 | 1 | 1 | 1 | 1 | 1 | 1 | 1 | 0 | 1 | 4 | 1 |
| 1455 | 3094815 | 2 | 1 | 4 | 2 | 1 | 2 | 1 | 2 | 0 | 0 | 2 | 2 | 4 | 0 | 1 | 1 | 1 | 1 | 1 | 1 | 1 | 0 | 1 | 4 | 1 |
| 1456 | 3094816 | 2 | 0 | 2 | 2 | 1 | 2 | 2 | 2 | 0 | 0 | 2 | 1 | 4 | 0 | 1 | 1 | 1 | 1 | 1 | 1 | 1 | 0 | 1 | 4 | 1 |
| 1457 | 3094817 | 2 | 1 | 2 | 2 | 1 | 1 | 2 | 2 | 0 | 0 | 2 | 1 | 4 | 0 | 1 | 1 | 1 | 1 | 1 | 0 | 1 | 0 | 1 | 3 | 1 |
| 1458 | 3094818 | 2 | 1 | 2 | 1 | 1 | 1 | 2 | 2 | 0 | 0 | 2 | 1 | 4 | 0 | 1 | 1 | 1 | 0 | 1 | 1 | 0 | 0 | 0 | 2 | 0 |
| 1459 | 3094819 | 2 | 1 | 2 | 3 | 1 | 2 | 2 | 2 | 0 | 0 | 2 | 1 | 4 | 0 | 1 | 1 | 1 | 1 | 1 | 1 | 1 | 0 | 1 | 4 | 1 |
| 1460 | 3094820 | 2 | 0 | 2 | 2 | 1 | 1 | 2 | 2 | 0 | 0 | 2 | 1 | 4 | 0 | 1 | 1 | 1 | 0 | 1 | 1 | 1 | 0 | 1 | 3 | 1 |
| 1461 | 3094821 | 2 | 1 | 4 | 3 | 1 | 2 | 2 | 2 | 0 | 0 | 2 | 2 | 4 | 0 | 1 | 1 | 1 | 1 | 1 | 1 | 1 | 0 | 0 | 3 | 1 |
| 1462 | 3094822 | 2 | 1 | 2 | 5 | 1 | 2 | 1 | 2 | 0 | 0 | 2 | 1 | 4 | 0 | 1 | 1 | 1 | 0 | 1 | 1 | 1 | 0 | 1 | 3 | 1 |
| 1463 | 3094823 | 2 | 0 | 2 | 4 | 1 | 1 | 1 | 2 | 0 | 0 | 2 | 1 | 4 | 0 | 1 | 1 | 1 | 1 | 1 | 1 | 1 | 0 | 0 | 3 | 1 |
| 1464 | 3094824 | 2 | 1 | 2 | 5 | 1 | 2 | 2 | 2 | 0 | 0 | 2 | 1 | 4 | 0 | 1 | 1 | 1 | 0 | 1 | 0 | 1 | 0 | 1 | 3 | 1 |
| 1465 | 3094825 | 2 | 1 | 5 | 4 | 2 | 2 | 2 | 2 | 0 | 0 | 2 | 3 | 4 | 0 | 1 | 1 | 1 | 0 | 1 | 1 | 1 | 0 | 1 | 3 | 1 |
| 1466 | 309491 | 2 | 1 | 2 | 4 | 1 | 2 | 1 | 2 | 0 | 0 | 2 | 1 | 4 | 1 | 1 | 1 | 1 | 1 | 0 | 1 | 1 | 0 | 1 | 4 | 1 |
| 1467 | 309492 | 2 | 0 | 2 | 2 | 1 | 1 | 4 | 2 | 0 | 0 | 2 | 1 | 4 | 0 | 1 | 1 | 1 | 1 | 1 | 1 | 1 | 0 | 1 | 4 | 1 |
| 1468 | 309493 | 2 | 0 | 2 | 4 | 1 | 2 | 4 | 2 | 0 | 0 | 2 | 1 | 4 | 0 | 1 | 1 | 1 | 0 | 1 | 1 | 1 | 0 | 1 | 3 | 1 |
| 1469 | 309494 | 2 | 0 | 2 | 5 | 1 | 1 | 3 | 2 | 0 | 0 | 2 | 1 | 4 | 0 | 1 | 1 | 1 | 0 | 1 | 0 | 1 | 0 | 1 | 3 | 1 |
| 1470 | 309495 | 2 | 0 | 2 | 2 | 1 | 1 | 1 | 2 | 0 | 0 | 2 | 1 | 4 | 0 | 1 | 1 | 1 | 0 | 1 | 1 | 1 | 0 | 1 | 3 | 1 |
| 1471 | 309496 | 2 | 1 | 5 | 3 | 2 | 1 | 1 | 2 | 1 | 1 | 5 | 3 | 4 | 1 | 1 | 1 | 1 | 0 | 1 | 1 | 1 | 0 | 1 | 4 | 1 |
| 1472 | 309497 | 2 | 1 | 2 | 4 | 1 | 1 | 2 | 2 | 0 | 0 | 2 | 1 | 4 | 0 | 1 | 1 | 1 | 1 | 1 | 1 | 1 | 0 | 1 | 4 | 1 |
| 1473 | 309498 | 2 | 0 | 2 | 1 | 1 | 1 | 4 | 2 | 0 | 0 | 2 | 1 | 4 | 0 | 1 | 1 | 1 | 1 | 1 | 1 | 1 | 0 | 1 | 4 | 1 |
| 1474 | 309499 | 2 | 0 | 2 | 3 | 1 | 2 | 4 | 2 | 0 | 0 | 2 | 1 | 4 | 0 | 1 | 1 | 1 | 0 | 1 | 1 | 1 | 0 | 1 | 3 | 1 |
| 1475 | 3094910 | 2 | 1 | 3 | 3 | 2 | 2 | 4 | 2 | 0 | 0 | 2 | 2 | 4 | 0 | 1 | 1 | 1 | 0 | 1 | 1 | 1 | 0 | 1 | 3 | 1 |
| 1476 | 3094911 | 2 | 0 | 2 | 3 | 1 | 2 | 4 | 2 | 0 | 0 | 2 | 1 | 4 | 0 | 1 | 1 | 1 | 0 | 1 | 1 | 1 | 0 | 1 | 3 | 1 |
| 1477 | 3094912 | 2 | 0 | 5 | 2 | 2 | 1 | 4 | 2 | 0 | 0 | 5 | 3 | 4 | 0 | 1 | 1 | 1 | 0 | 1 | 1 | 1 | 0 | 1 | 3 | 1 |
| 1478 | 3094913 | 2 | 1 | 2 | 2 | 1 | 1 | 4 | 3 | 0 | 0 | 2 | 1 | 4 | 0 | 1 | 1 | 1 | 1 | 0 | 1 | 1 | 0 | 0 | 3 | 1 |
| 1479 | 3094914 | 2 | 1 | 3 | 2 | 1 | 2 | 1 | 2 | 0 | 0 | 2 | 2 | 4 | 1 | 1 | 1 | 1 | 1 | 1 | 1 | 1 | 0 | 1 | 4 | 1 |
| 1480 | 3094915 | 2 | 1 | 2 | 3 | 1 | 1 | 1 | 2 | 0 | 0 | 2 | 1 | 4 | 0 | 1 | 1 | 1 | 0 | 1 | 1 | 1 | 0 | 1 | 3 | 1 |
| 1481 | 3094916 | 2 | 1 | 2 | 4 | 1 | 1 | 1 | 2 | 0 | 0 | 2 | 1 | 4 | 1 | 1 | 1 | 1 | 1 | 0 | 1 | 1 | 0 | 1 | 4 | 1 |
| 1482 | 3094917 | 2 | 1 | 2 | 2 | 1 | 1 | 1 | 2 | 0 | 0 | 2 | 1 | 4 | 1 | 1 | 1 | 1 | 0 | 0 | 0 | 0 | 0 | 0 | 2 | 0 |
| 1483 | 3094918 | 2 | 0 | 4 | 2 | 1 | 2 | 4 | 2 | 0 | 0 | 2 | 2 | 4 | 1 | 1 | 1 | 1 | 0 | 1 | 1 | 1 | 0 | 1 | 4 | 1 |
| 1484 | 3094919 | 2 | 0 | 2 | 2 | 1 | 2 | 4 | 2 | 0 | 0 | 2 | 1 | 4 | 0 | 1 | 1 | 1 | 0 | 1 | 1 | 1 | 0 | 1 | 3 | 1 |
| 1485 | 3094920 | 2 | 0 | 2 | 3 | 2 | 1 | 4 | 2 | 1 | 0 | 2 | 1 | 4 | 0 | 1 | 1 | 1 | 0 | 1 | 1 | 1 | 0 | 1 | 3 | 1 |
| 1486 | 3094921 | 2 | 1 | 2 | 3 | 1 | 2 | 4 | 2 | 0 | 0 | 2 | 1 | 4 | 1 | 1 | 1 | 1 | 1 | 1 | 1 | 1 | 0 | 1 | 4 | 1 |
| 1487 | 3094922 | 2 | 0 | 2 | 3 | 1 | 2 | 4 | 2 | 0 | 0 | 2 | 1 | 4 | 0 | 1 | 1 | 1 | 1 | 1 | 1 | 1 | 0 | 1 | 4 | 1 |
| 1488 | 3094923 | 2 | 0 | 2 | 2 | 1 | 1 | 1 | 2 | 0 | 0 | 2 | 1 | 4 | 1 | 1 | 1 | 1 | 1 | 1 | 1 | 1 | 0 | 1 | 4 | 1 |
| 1489 | 3094924 | 2 | 0 | 5 | 1 | 3 | 1 | 4 | 2 | 0 | 0 | 5 | 3 | 4 | 0 | 1 | 1 | 1 | 1 | 1 | 1 | 1 | 0 | 1 | 4 | 1 |
| 1490 | 3094925 | 2 | 0 | 5 | 2 | 1 | 2 | 4 | 2 | 0 | 0 | 5 | 3 | 4 | 0 | 1 | 1 | 1 | 1 | 1 | 1 | 1 | 0 | 1 | 4 | 1 |
| 1491 | 3094926 | 2 | 1 | 4 | 4 | 2 | 1 | 1 | 2 | 0 | 0 | 2 | 2 | 4 | 1 | 1 | 1 | 1 | 1 | 1 | 1 | 1 | 0 | 0 | 4 | 1 |
| 1492 | 3094927 | 2 | 1 | 2 | 4 | 2 | 1 | 1 | 2 | 0 | 0 | 2 | 1 | 4 | 1 | 1 | 1 | 1 | 0 | 0 | 0 | 0 | 0 | 0 | 2 | 0 |
| 1493 | 3094928 | 2 | 1 | 5 | 2 | 2 | 1 | 1 | 2 | 1 | 1 | 5 | 3 | 4 | 1 | 1 | 1 | 1 | 1 | 1 | 1 | 0 | 0 | 1 | 4 | 1 |
| 1494 | 3094929 | 2 | 1 | 2 | 1 | 1 | 1 | 1 | 2 | 0 | 0 | 2 | 1 | 4 | 0 | 1 | 1 | 1 | 0 | 1 | 0 | 1 | 0 | 1 | 3 | 1 |
| 1495 | 3094930 | 2 | 1 | 2 | 3 | 1 | 2 | 1 | 2 | 0 | 0 | 2 | 1 | 4 | 0 | 1 | 1 | 1 | 1 | 1 | 1 | 1 | 0 | 1 | 4 | 1 |

Variables explanation

Res_ID Respondent ID
MES Malaria endemic settings
1 High (East Sumba district)
2 Low (East Manggarai district)

| | | |
|---|---|---|
| | 3 | Moderate (Belu district) |
| G | Gender of participants | |
| | 0 | Female |
| | 1 | Male |
| Edu_1 | Education level of participants | |
| | 1 | No education |
| | 2 | Primary school |
| | 3 | Junior high school |
| | 4 | Senior high school' |
| | 5 | Diploma or above |
| AG | Age group of participants | |
| | 1 | < 30 |
| | 2 | 30 – 39 |
| | 3 | 40 – 49 |
| | 4 | 50 – 59 |
| | 5 | > 60 |
| SES | Social economic status of participants | |
| | 1 | Low |
| | 2 | Average |
| | 3 | High |
| FZ | Family size | |
| | 1 | ≤ 4 |
| | 2 | > 4 |
| DTHF | Distance to the nearest health facilities | |
| | 1 | < 1 Km |
| | 2 | 1 – 2 Km |
| | 3 | 2 – 3 Km |
| | 4 | >= 3 Km |
| HFC | The nearest health facilities | |
| | 1 | Village maternity posts |
| | 2 | Village health Post |
| | 3 | Sub Public Health centres |
| | 4 | Public Health centres |
| IMWP | Household Income in relation to Province Minimum Wage (PMW) | |
| | 0 | < PMW |
| | 1 | >= PMW |
| IIPL | Household Income in relation to Indonesia Poverty Line (IPL) | |
| | 0 | < IPL |
| | 1 | >= IPL |
| Job | Main occupation of participants | |
| | 1 | Housewife |
| | 2 | Farmer |
| | 3 | Entrepreneurs |
| | 4 | Other |
| | 5 | Govt. or non-govt. employment |
| Edu2 | Education level of participants in 3 categories | |
| | 1 | Primary or no education |
| | 2 | Secondary education |
| | 3 | Diploma or above |
| Eth | Ethnicity in East Nusa Tenggara Province | |
| | 1 | Sumba |
| | 2 | Others |
| | 3 | Atoni |
| | 4 | Manggarai |
| Q1 | Hearing malaria term | |
| | 0 | No |
| | 1 | Yes |
| Q2 | Dangerous effecf of malaria to health | |
| | 0 | No |
| | 1 | Yes |
| Q3 | Awareness that malaria can be prevented | |
| | 0 | No |
| | 1 | Yes |
| Q4 | The ability to identify fever as main sympthom of malaria | |
| | 0 | No |
| | 1 | Yes |
| Q5 | The correct knowledge on mosquito bite as transmission mode of malaria | |
| | 0 | No |
| | 1 | Yes |
| Q6 | Knowledge on sleeping under non-LLINs | |
| | 0 | No |
| | 1 | Yes |
| Q7 | Knowledge on sleeping under LLINs | |
| | 0 | No |
| | 1 | Yes |
| Q8 | Knowledge on using mosquito coil | |
| | 0 | No |
| | 1 | Yes |
| Q9 | Knowledge on keep house clean | |
| | 0 | No |

| | | |
|---|---|---|
| | 1 | Yes |
| Q10 | Seeking treatment within 24 hours at health facility | |
| | 0 | No |
| | 1 | Yes |
| MKS | Malaria knowledge score | |
| | 1 | Very poor |
| | 2 | Poor |
| | 3 | Good |
| | 4 | Excellent |
| MA | Malaria awareness status of participants | |
| | 0 | Unaware |
| | 1 | Aware |
